# An Integrated Molecular Atlas of Alzheimer’s Disease

**DOI:** 10.1101/2021.09.14.21263565

**Authors:** Maria A. Ulmer, Jan Krumsiek, Serge Nataf, Kwangsik Nho, Anna K. Greenwood, Jesse C. Wiley, Lina-Liv Willruth, Tong Wu, Orhan Bellur, Bharadwaj Marella, Kevin Huynh, Patrick Weinisch, Werner Römisch-Margl, Nick Lehner, Yacoub A. Njipouombe Nsangou, The AMP-AD Consortium, The Alzheimer’s Disease Neuroimaging Initiative, The Alzheimer’s Disease Metabolomics Consortium, Jan Baumbach, Peter J. Meikle, Andrew J. Saykin, P. Murali Doraiswamy, Cornelia van Duijn, Karsten Suhre, Rima Kaddurah-Daouk, Gabi Kastenmüller, Matthias Arnold

## Abstract

Alzheimer’s disease (AD) is a complex neurodegenerative disorder with multifactorial etiology and widespread molecular manifestations. Investigating molecular disease associations in a broader multi-level context across omics modalities remains one central challenge in AD research, despite the increasing availability of large-scale omics data. The AD Atlas, an online multi-omics resource, provides access to harmonized, disease-relevant data from over 25 large studies on 20,363 protein-coding genes, 8,396 proteins, 1,328 metabolites and 43 AD-related phenotypes interconnected by 979,190 significant associations. Results from AD-specific omics studies from AMP-AD, NIAGADS, and other initiatives are complemented with molecular associations from population-based studies in a comprehensive network resource to provide a genome-scale molecular view on AD. In a deep learning-based evaluation of the AD Atlas content, we demonstrate the utility of the network for data-driven identification of modules strongly enriched for AD-related functional domains. We provide full access to the AD Atlas at www.adatlas.org.

## INTRODUCTION

Late-onset Alzheimer’s disease (AD) is a progressive neurodegenerative disorder, for which there is currently no cure or efficacious preventive therapy and only modestly effective symptomatic treatments (*1*). AD is a multifactorial disease linked to all molecular layers from genetic and epigenetic variation through transcriptional changes to altered abundances of proteins and metabolites, which interact in complex networks (*2*). Despite significant advances in the study of AD and related dementias, there are many challenges remaining. One of the most prominent missing pieces are robust and reliable biomarkers for both diagnosis and therapeutic intervention that are embedded in the context of multi-level molecular changes observed in AD.

NIH’s Accelerating Medicines Partnership in AD (AMP-AD; https://www.nia.nih.gov/research/amp-ad; (*3*)) program is working towards this goal through generation and examination of diverse data including multi-omics profiling of different modalities across relevant tissues. Data generated through the AMP-AD initiative is shared through the AD Knowledge Portal (https://adknowledgeportal.org; (*4*)) and interactive visualizations, aimed to support the evaluation of data at the single target level, are provided through the Agora Platform (https://agora.adknowledgeportal.org/). However, an analytical tool that incorporates biological entities into their multi-omics context has so far been missing. To this end, biological networks offer an intuitive framework to integrate and store densely connected biomedical data, making them an attractive data structure for multi-omics integration efforts (*5*). Heterogeneous networks, which consist of multiple types of nodes (e.g. metabolites, genes and phenotypes) and edges (e.g. partial correlation of metabolites, gene co-expression), have been particularly useful to describe the complex interplay within and between biological layers (*6*). Yet, standardized processing of different data types, consolidation of association data across population-based and patient-related sources, and integration into a single network requires expertise across research domains. Therefore, public multi-layer network repositories are needed to enable researchers with various backgrounds to explore AD, its biomarkers, and associated (endo-)phenotypes in a multi-omics context.

In this study, we present the AD Atlas, a publicly available, network-based resource built using an extended QTL-based integration strategy combined with a composite network approach (*7*) to integrate and harmonize data from more than 25 large omics studies. Based on data from knowledge bases and healthy cohorts, we first constructed a generalized, disease-independent framework of intra- (e.g. gene-gene) and inter-omics (e.g. metabolite-gene) relationships. Using large-scale association data on different aspects of AD – including data from AMP-AD, NIAGADS, and other large studies and consortium efforts – this framework was then transformed into an integrated multi-omics knowledge base for markers of AD. The resulting comprehensive catalogue of multi-omics relationships is stored in the graph-based database management system Neo4j. Public access to the resource is provided through a web-based user interface (www.adatlas.org) featuring several data visualization and analysis tools that enable screening and meta-analysis of multi-omics data in the context of AD and independent of in-house bioinformatics capacities. (A detailed overview of the interface, including multiple showcases, is provided as supplementary material.) To evaluate the validity of our harmonization procedures and to provide global information on the content of the AD Atlas, we quantify both the agreement between integrated datasets and the complementarity of information contained by different omics data types. We further demonstrate that the AD Atlas can be used to derive biologically meaningful information by coupling deep-learning-based dimensionality reduction on the global network structure with unsupervised cluster analysis, which segregated the network into distinct functional modules significantly enriched for different AD-related biological domains.

## RESULTS

### Resource overview

The Alzheimer’s disease (AD) Atlas is a genome-scale catalogue of association results from large omics studies in network format. It builds upon and extends the concept of augmenting results of a single study with (multi-)omics association data from external sources, e.g., to strengthen confidence in and functionally annotate disease-related molecular associations (*8*, *9*) or to prioritize candidate causal genes in genome-wide association studies (GWAS) using overlapping eQTL or pQTL signals (*10*). Building upon this concept, and by projecting raw quantitative molecular data to the association results space, the heterogeneity of the underlying data is substantially reduced. As a result, the AD Atlas enables the extraction of context-specific molecular subnetworks by exploring and interlinking biological entities (e.g. metabolites, SNPs, genes) of interest in their network neighborhood.

We established the underlying association database by inferring pairwise relationships between molecular entities in large-scale studies, resulting in a complex collection of data stored in a Neo4j graph database (**Figure 1**). To this end, we first used comprehensive population-based data and genomic annotation databases, providing a generalized framework to study multi-omics relationships globally. On top of this, we integrated association data from large-scale studies on diverse aspects of AD including hundreds to hundreds of thousands of individuals in order to transform this framework into an integrated multi-omics knowledge base for markers of AD. These data include recent large genetic association (meta-)analyses of AD and AD biomarkers from NIAGADS and other large efforts, as well as differences in transcriptomics, proteomics, and metabolomics markers observed in AD or in relation to AD endophenotypes. The latter are predominantly based on data generated on thousands of human blood samples and brain tissue from multiple brain regions using state-of-the-art technologies and analyzed using standardized processing pipelines developed by the AMP-AD program and partnering initiatives.

**Figure 1.**
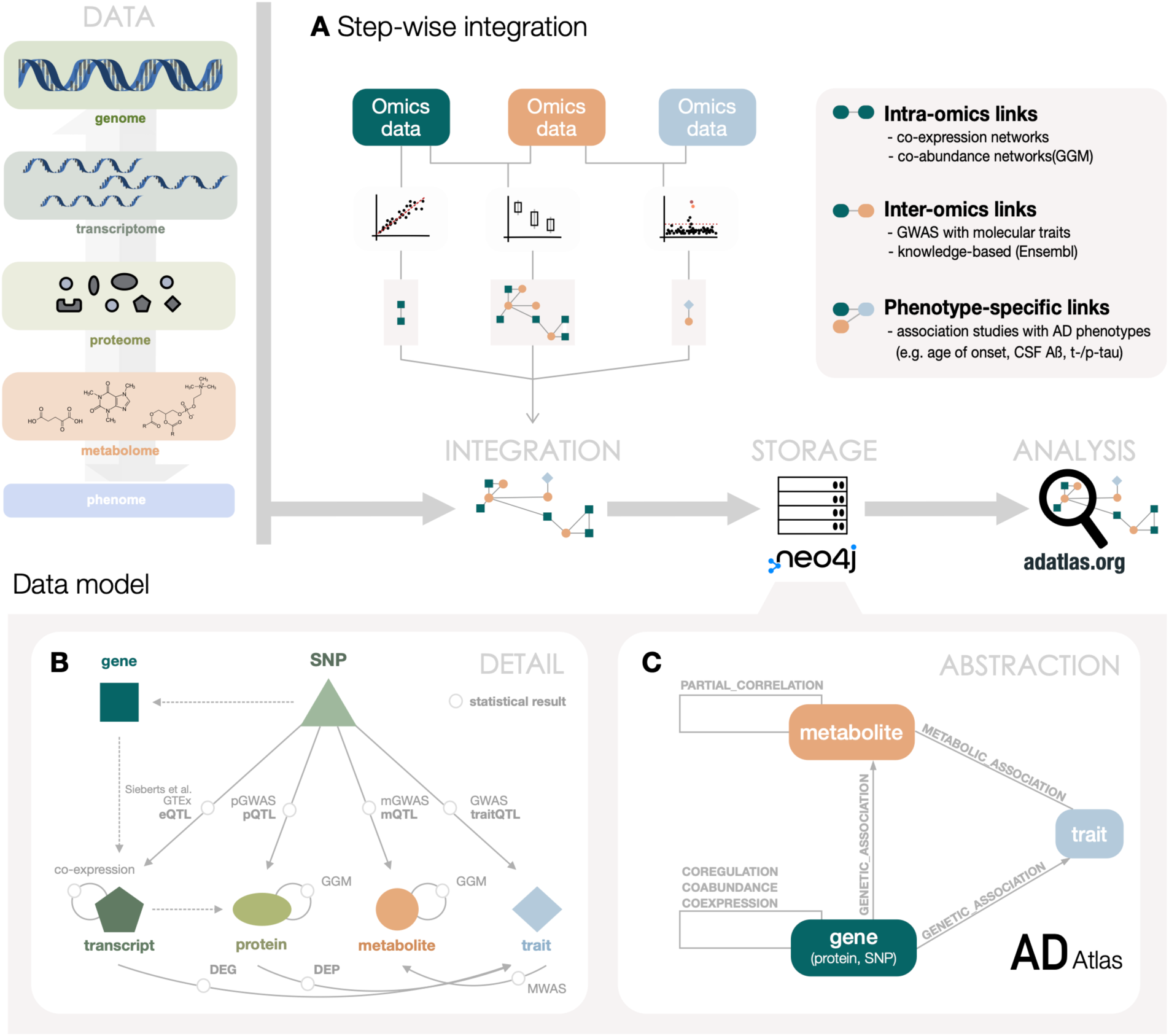
Data integration overview. The AD Atlas is a multi-omics resource that enables the integration and analysis of heterogeneous omics datasets in the context of Alzheimer’s disease (AD). **(A)** Step-wise multi-omics integration approach underlying the AD Atlas. Using statistical analysis, such as association analysis and partial correlation, omics data collected in large population-based studies is used to infer biological relationships between (inter-omics) and within (intra-omics) omics layers. Links to AD (endo-)phenotypes from large-scale case-control or biomarker studies enable multi-omics exploration in the context of AD. **(B)** The collection of omics data, i.e., information on measured entities (nodes) and statistical or knowledge-based relationships between them (edges) are stored in a comprehensive graph structure. The data model depicted is simplified for clarity. **(C)** To enable efficient data access and reduce the complexity of the resulting networks, an abstracted data view is constructed by projecting SNPs, transcripts and proteins onto genes and consolidating metabolites across analytical platforms. Multiple edges connecting the same entities are summarized, retaining as much detailed information as possible in form of edge annotations. The web-interface available at www.adatlas.org provides access to this data view.

To increase data accessibility and downstream interpretability, we next summarized and extracted the data into an abstracted data view (**Methods, Supplementary Figure 1A**). The resulting summarized network representation (**Figure 1C**) is accessible through the user interface and consists of three node types; (I) metabolites, mapped across available platforms where possible (**Supplementary Figure 1B**), (II) genes, which cover SNPs, transcripts, and proteins (**Methods**), and (III) traits, which describe AD (endo-)phenotypes and biomarkers. Various different relationship types interconnect these entities (see **Methods** and **Supplementary Table 1**). Integrating over 25 different studies and analyses, the AD Atlas includes information on 20,363 protein-coding genes, 8,396 proteins, 1,328 metabolites and 43 unique AD-related traits (**Supplementary Table 2**). Traits include cerebrospinal fluid (CSF) and imaging biomarkers, partially with different covariate settings (adjustment for *APOE* genotype), stratifications or stagings for neuropathologies, amounting to 67 traits in total (**Supplementary Table 3**), with further details provided in **Supplementary Table 4**. Biological entities and traits are linked by over 900,000 relationships, representing significant statistical associations inferred from large-scale quantitative data from population-based cohorts and AD-specific studies. A more detailed summary of the data compiled in the AD Atlas is shown in **Table 1**. Important terms and concepts used throughout this manuscript are defined in **Supplementary Box 1**.

**Table 1.** Data compiled in the AD Atlas (simplified data view).

|  |  | Nentities | References |
| --- | --- | --- | --- |
| Nodes | AD-related **meta-trait** | 10 | - |
|  | AD-related **traits** | 67<br>43 unique | - |
|  | **Genes** (protein-coding) | 20,363 |  |
|  | with differential gene expression data | 14,731 | (39) |
|  | with differential protein abundance data | 7,867 | (40, 41) |
|  | **Metabolites** | 1,328 | - |
| Relationships | **Genetic associations with AD phenotypes**<br>*traitQTLs* | 12,361 | (42–51)<br>This study* |
|  | **Genetic associations with metabolic traits**<br>*mQTLs; sample types (n=3)* | 165,719 | (52–56)<br>This study* |
|  | **Metabolic associations with AD phenotypes**<br>*mWAS; sample types (n=2)* | 1,018 | (38, 57–59) |
|  | **Gene co-expression data**<br>*brain regions (n=7)* | 232,516 | (39) |
|  | **Genetic co-regulation**<br>*eQTLs; tissues (n=49)* | 493,117 | (27, 30) |
|  | **Protein co-abundance data**<br>*partial correlation-based; sample types (n=2)* | 73,296 | (8, 40, 41) |
|  | **Metabolic pathways**<br>*partial correlation-based; sample types (n=2)* | 1,163 | (38, 60)<br>This study* |

\*Details given in the **Supplementary Material**

### Quantification of data concordance across studies and complementarity of different omics

By incorporating associations from various statistical analyses and multiple studies, the AD Atlas offers unique opportunities to explore results across studies as well as data and sample types. To quantify the agreement between networks derived from different datasets, we calculated the pairwise Jaccard Index and the Overlap Coefficient (**Methods**) and visualized the results in heat maps (**Figure 2**), which provide a global overview for identifying notable patterns with either high agreement or complementarity. Pairwise comparison of GWAS studies at the meta-trait-level (**Methods**) revealed generally high genetic overlap between different association studies of AD (**Figure 2**). Overall, this overlap was larger when the comparison was performed on the level of genes (using the SNP to gene mapping as defined by the AD Atlas (**Methods**)) rather than SNPs directly (**Supplementary Figures 3** and **4**). Particularly high concordance can be seen between recent large-scale meta-analysis of AD by proxy and diagnosed AD, as well as studies on CSF biomarkers and neuropathologic features. Varying agreement between studies are most likely due to differences in factors, such as study population, inclusion criteria, or sample size, which may affect statistical power.

**Figure 2.**
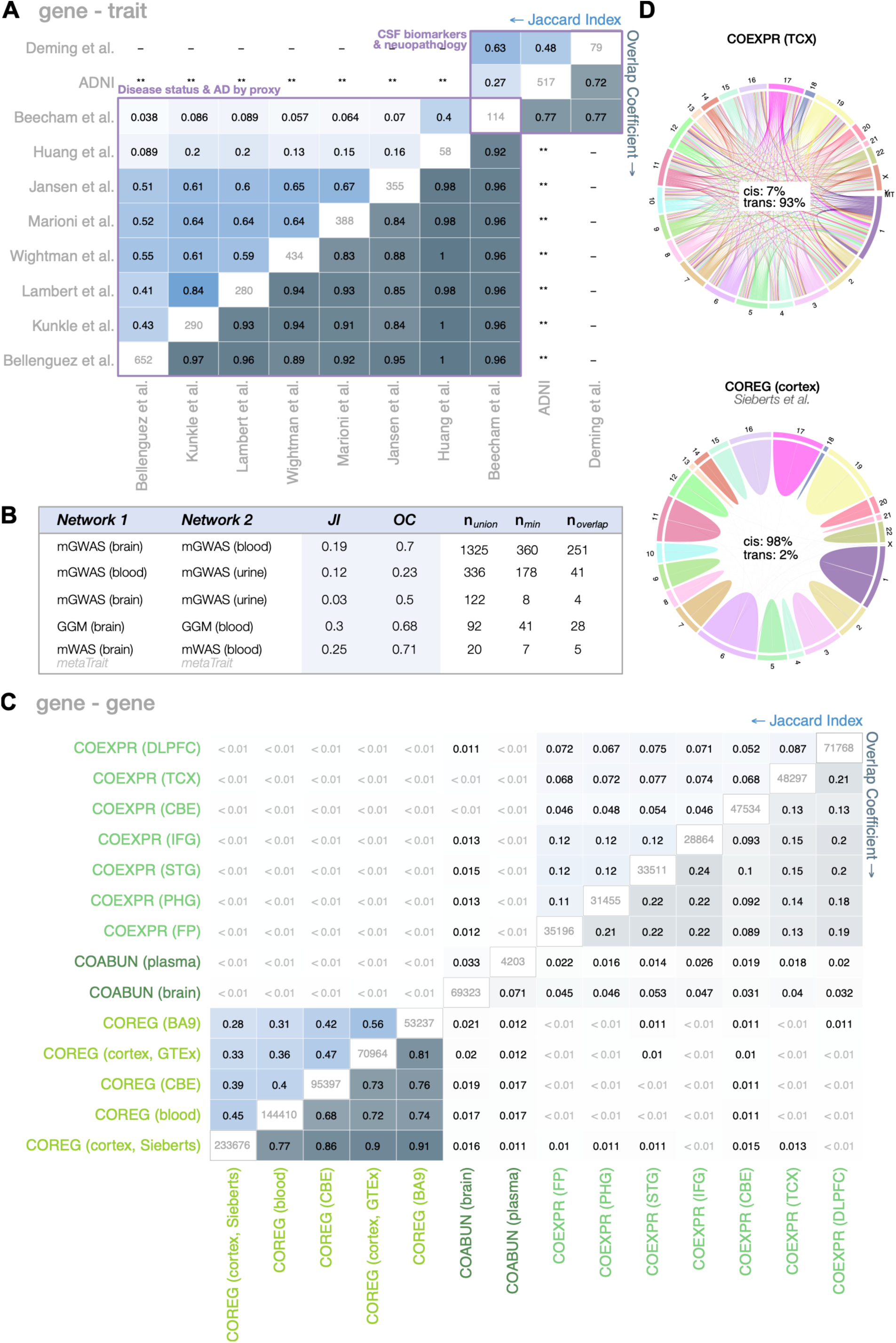
Global view on cross-study and -sample type replication. **(A)** Heatmap of genetic correlation between AD-relevant genome-wide association studies (GWAS). Pairwise comparisons were performed at the common meta-trait- and gene-level (see **METHODS**). Diagonal: total number of edges in unfiltered network. Upper triangle: Jaccard index (JI). Lower triangle: Overlap coefficient (OC). Cells are colored by value; zero (white) to one (blue). Statistics used to calculate the JI and OC are given in **Supplementary Figure 2**. **ADNI included in meta-analyses as part of IGAP. **(B)** Results of cross-tissue comparisons using metabolomics-related data. The number of edges of the union of both networks (n_union_), smaller network (n_min_) and overlap (n_overlap_) are provided for each comparison. **(C)** Heatmap of pairwise comparisons between selected networks inferred using eQTL (co-regulation; COREG), proteomics (co-abundance; COABUN) and transcriptomics data (co-expression; COEXPR). Networks were filtered for common genes prior to comparison. Details analogous to (A). Statistics used to calculate the JI and OC are given in **Supplementary Figure 5**. **(D)** Chord diagrams showing genetic cis- (within same chromosome) and trans- (across chromosomes) connections for selected co-regulation (eQTL-based) and co-expression (transcript-based) networks. Co-regulation edges in the AD Atlas mainly represent cis-effects whereas co-expression edges are predominantly in trans. Plot colored by chromosome.

Next, we examined the agreement of metabolomics studies across sample types and found that metabolite associations observed in blood can, at least to some extent, be replicated in the brain (**Figure 2B**). Interestingly, despite the blood-brain barrier being considered a robust barrier, the results from blood showed greater similarity to those from the brain than to the renal/urinary system, which is similarly isolated. Furthermore, we evaluated the complementarity of different omics studies focusing on the same entities (e.g., genes). Notably, we observed a substantial overlap within omics data types, whereas there was minimal overlap across different omics. For example, the agreement between gene co-expression and protein co-abundance networks was minimal, as illustrated in **Figure 2C**. Additionally, we found a strong similarity between co-regulation (eQTL) networks, even when considering different studies and tissues. Further analysis indicated that these networks primarily capture cis-effects, while co-expression and co-abundance edges predominantly reflect trans-effects, as depicted in **Figure 2D** and **Supplementary Figure 6**.

Lastly, by functionally characterizing the gene set implicated in AD by each omics layer (**Supplementary Figure 7**), as well as their shared number of genes (**Supplementary Figure 8**) we saw differences in contribution to AD-relevant processes which is in line with a recent meta-analysis (*11*). For example, while only the proteomic and transcriptomic layers of AD showed strong enrichment for metabolic (p=4.73e-60 and p=1.13e-09, respectively) and synaptic (p=1.52e-106 and p=1.24e-02, respectively) processes, only the genomic layer showed an enrichment for immune response-related genes (p=3.17e-03).

These findings emphasize the complementary nature of multi-omics data and underscore the importance of integrating different types of data.

### Exploration of context-specific molecular subnetworks through the AD Atlas user interface

The AD Atlas is accessible via an online interactive user interface (www.adatlas.org; **Figure 3**). Through this interface users can dynamically generate, explore, and analyze context-specific molecular subnetworks surrounding entities of their interest, i.e., specific genes, metabolites or AD-related (endo-)phenotypes, which can be chosen in the network settings panel of the network browser (**Figure 3B** and **C**). The concrete process of subnetwork generation depends on the entry point and is depicted in **Figure 4**:

i. Trait- or meta-trait-centric subnetworks The user specifies a trait or collection of traits as input. Genes and metabolites directly associated with the traits of interest are added using data from genome- and metabolome-wide association studies. Next, these entities are annotated with associated genes and metabolites (through mQTL data) and finally, all associated traits are added for this final set of genes and metabolites, again using data from genome-and metabolome-wide association studies. All relationships between these entities that meet the user-specified criteria, including intra-omics links, are included in the returned network. Here, meta-traits are pre-defined collections of traits loosely following the A-T-(N)-(C) schema (*12*) (see **Supplementary Table 3**).
ii. Gene-centric subnetworks The user provides a gene or a set of genes as input (provided as gene symbol or Ensembl gene ID). Traits and metabolites that are directly associated with the genes of interest are added using data from GWAS with AD-related traits and GWAS with metabolic traits. Lastly, all relationships between these entities that meet the user- specified criteria are added to the resulting network. **Figure 4B** provides a step-by-step example.
iii. Metabolite- or pathway-centric subnetworks The user provides a metabolite or a set of metabolites as input (provided as a biochemical name). Traits and genes that are associated with the metabolite of interest are added using data from MWAS with AD-related traits and mQTL data generated in GWAS with metabolic traits. Lastly, all relationships between these entities that meet the user-specified criteria are added to the resulting network. Here, pathways are predefined sets of metabolites, currently defined by pathway annotations (super- /sub-pathways and metabolite classification) as provided by the metabolomics lab or platform vendor that generated the data.

**Figure 3.**
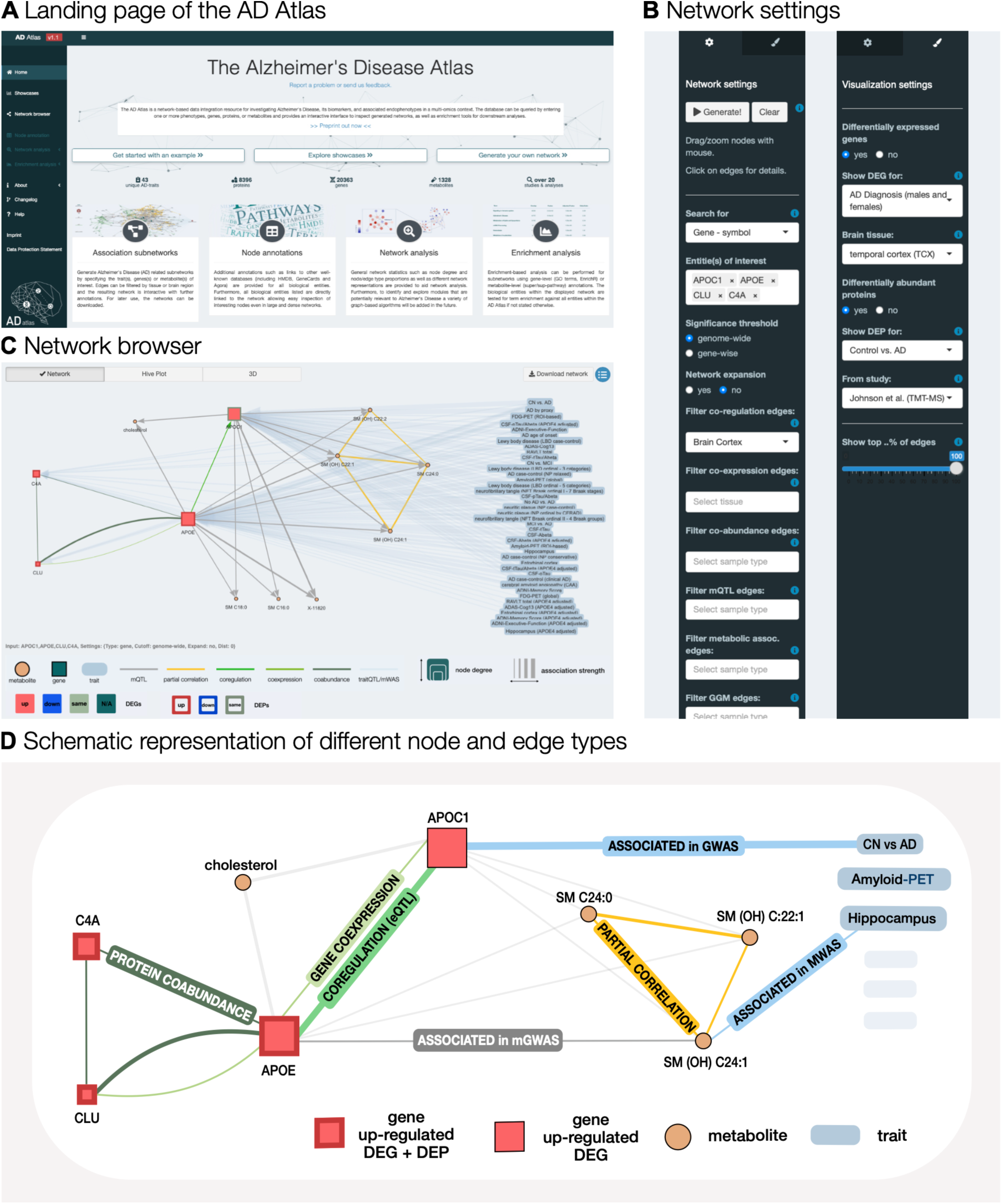
User interface. Users can generate context-specific molecular subnetworks and apply analytical tools via our interactive user interface (www.adatlas.org). **(A)** AD Atlas landing page. **(B)** In the network browser, users can use the right panel to specify parameters to generate context-specific molecular subnetworks (left). By clicking the paintbrush icon at the top of the panel, additional information, such as differential gene expression in disease, can be overlayed using the visualization options (right). **(C)** Network browser showcasing the multi-omics subnetwork surrounding the input genes *APOE, APOC1, C4A* and *CLU*. **(D)** Schematic representation of the molecular subnetwork seen in **C**, showcasing the different node and edge types. For simplicity, not all nodes and edges are included. GWAS: genome-wide association study; mGWAS: GWAS with metabolic traits; MWAS: metabolome-wide association study; protein co-abundance: partial correlation between proteins; eQTL: expression quantitative trait locus; DEG: differentially expressed gene; DEP: differentially expressed protein.

**Figure 4.**
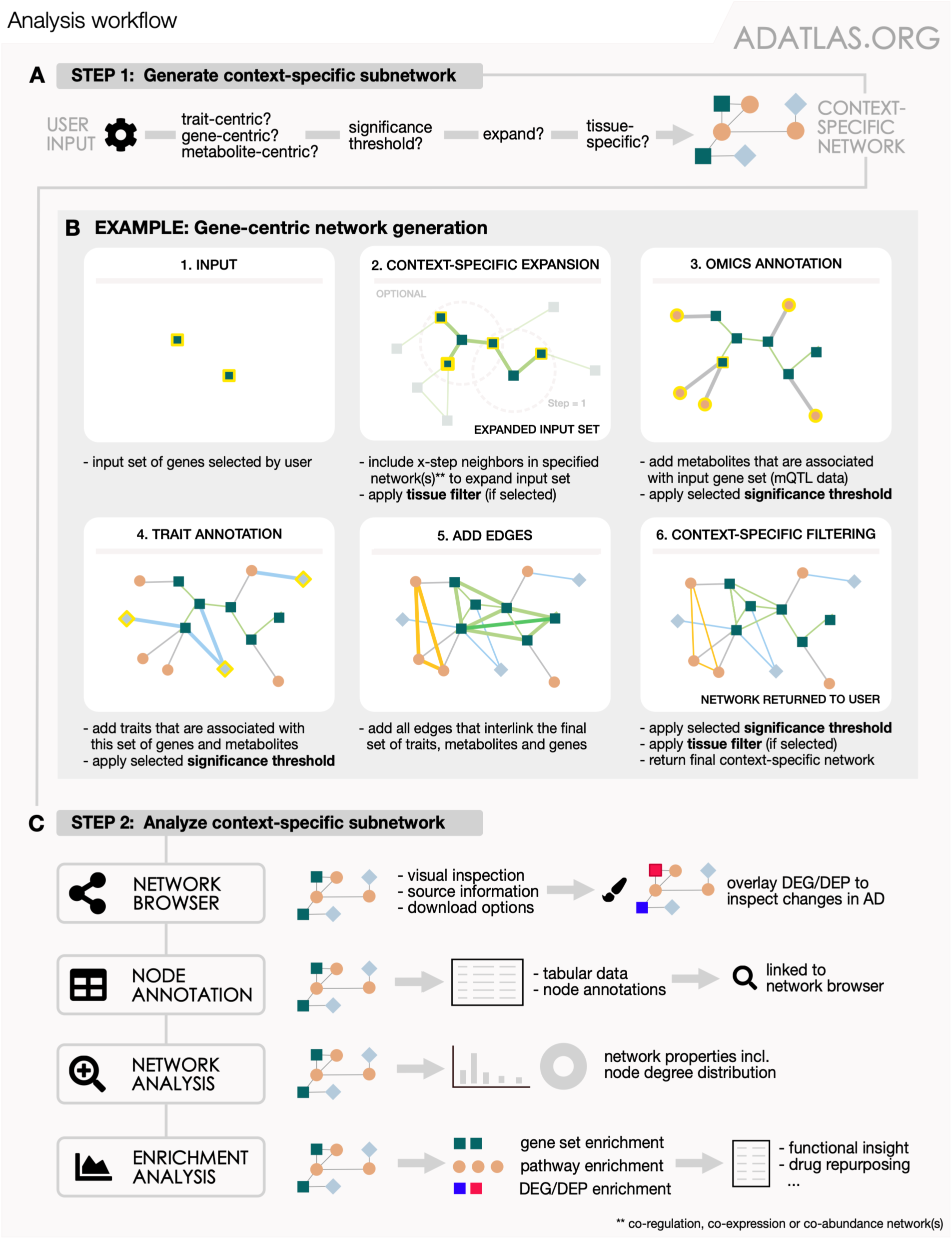
Network generation and analysis workflow. **(A)** Users can generate context-specific molecular subnetworks surrounding entities (traits, genes, metabolites) of interest. **(B)** Step-by-step description of the network generation process, here using the gene-centric approach. Statistical inter- and intra-omics links are added to the input genes provided by the user in a successive manner. Newly added nodes and edges are indicated by a yellow border and thick edges, respectively. Steps applying edge filtering (tissue-or significance-based) are indicated. **(C)** The resulting networks can be analyzed using tools, such as enrichment analysis, via our interactive user interface (www.adatlas.org; Figure 3). DEG: differentially expressed gene; DEP: differentially expressed protein.

Additionally, these molecular subnetworks can be customized by applying edge filters (e.g., by sample type, tissue or brain region) and by adjusting the significance threshold for genetic associations (**Methods**). To gain further insights into the functional neighborhood of the user-specified metabolites or genes, the interface allows expanding the set of input entities before the procedure described in (i) – (iii) to include the 1-step or 2-step neighbors of the initial input entities; neighbors are assessed using co-regulation (eQTL), transcript co-expression or protein co-abundance data for genes, or metabolite co-abundance (partial correlation) data for metabolites.

After construction, the context-specific subnetworks can be visually inspected using the network browser (**Figure 3C**) and can be subjected to a number of downstream analyses within the AD Atlas user interface. Information on differentially expressed genes (DEG) and proteins (DEP) can be overlaid onto the networks to investigate the extent and direction of dysregulation in AD using the visualization options (**Figure 3B**). Here, users can choose the underlying association model (sex-specific or pooled analysis) and brain region. Furthermore, the entities in the generated network can be functionally characterized using gene set and pathway enrichment analysis (**Methods**).

Showcases exemplifying the use of subnetwork generation and analysis in the context of specific research questions are provided in **Supplementary Material** and can be interactively explored under www.adatlas.org/?showcases. Specifically, (I) a subnetwork of lipid metabolism and transport constructed around the AD-related genes *APOE* and *CLU* highlights known candidates for drug repositioning; (II) a subnetwork constructed around the targets of statins suggests that the (off-)target *ITGAL* links to neuroinflammation through TREM2 signaling; (III) a subnetwork constructed around AD-associated sphingomyelins contextualizes the molecular link between the sphingomyelin pathway and AD pathology; (IV) a subnetwork surrounding marker genes for homeostatic microglia and disease-associated microglia suggests possible involvement of androgens. These examples demonstrate the utility of the AD Atlas resource for effective *ad hoc* exploration of the molecular landscape in AD to identify potential drug repositioning candidates or to generate new mechanistic hypotheses.

### Assessment of the utility of the global network structure to derive disease-relevant information

Next, we assessed if the structure and content of the AD Atlas network globally mirrors known biological relationships in AD despite its correlational nature. To this end, we applied a node embedding approach, where nodes in the network are reduced into a lower-dimensional vector space while retaining structural information of the network using random walks (*13*, *14*). In such an embedding space, similar/proximal nodes in the original network are represented by similar vectors (or embeddings) and can be subjected to a number of downstream analyses, including clustering, community detection and network visualization (*15*, *16*). Here, we first constructed a brain-specific network (**Methods**, node degree distribution is shown in **Supplementary Figure 9**). Next, we determined the optimal number of dimensions (n = 130) to embed the network at minimal information loss using the approach by Gu et al. (*17*) and used DeepWalk as implemented in the python package GeneWalk (*13*) for node embedding.

Visualization of the AD Atlas network content in 2-dimensional (2D) space using UMAP (Uniform Manifold Approximation and Projection) (*18*) on the 130-dimensional node embeddings revealed a clear structure (**Figure 5**) while the corresponding 2D representation of embeddings from a random network did not show any structure (**Supplementary Figure 10A**). Moreover, overlaying the gene nodes with differential expression results for AD cases vs. controls showed distinct global patterns (**Figure 5A**), indicating that genes whose expression is similarly affected in disease are located in proximal regions of the AD Atlas network. This is noteworthy, as differential expression information was not used to build the network and, thus, not provided to the embedding algorithm. Annotating gene nodes with AD-relevant biological domains (manual curation of GO terms into AD-relevant domains, see **Methods**) showed significant enrichment of specific domains, i.e., biological functions, in distinct clusters obtained by hierarchical clustering of nodes in the embedding space (**Figure 5E**, **Supplementary Table 5**). Among other examples, genes involved in immune response, a pathway previously implicated in AD pathophysiology (*19*), were highly enriched (*p*-value= 2.60e-81; one-sided *Fisher’s exact test*) in a cluster that showed up-regulation of gene expression in disease (*p*-value = 2.37e-30; one-sided *Fisher’s exact test*) and tight linkage (in terms of path length in the network; *p*-value = 3.77e-42; one-sided *t-test*) to AD phenotypes (**Figure 5**). The full set of enrichment results is provided in **Supplementary Table 6**.

**Figure 5.**
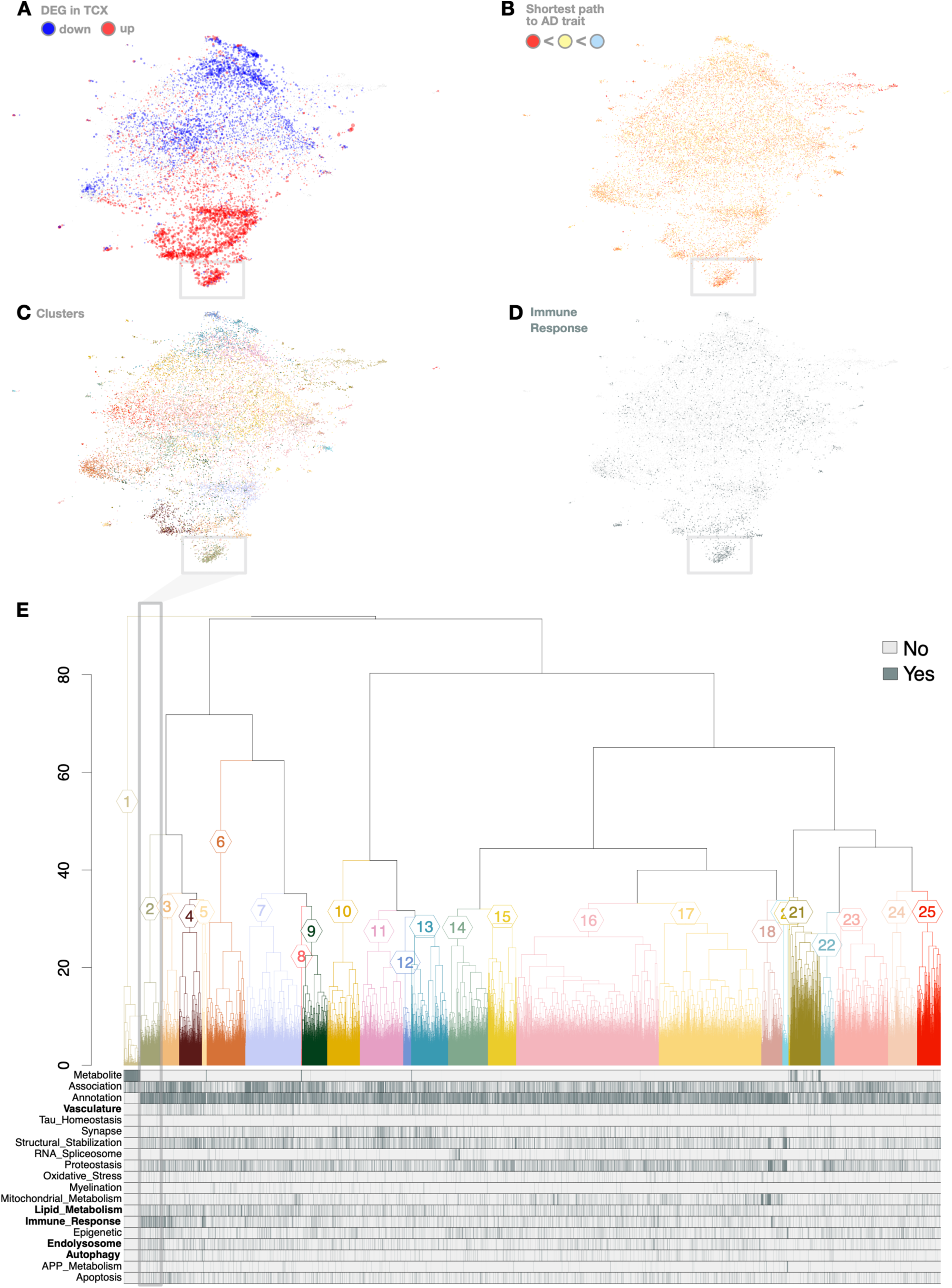
Global assessment of biological information content in the AD Atlas network structure. The nodes of the AD Atlas network were embedded into 130-dimensional vector space and projected to 2-dimensional space for visualization, as described in **Methods**. **(A)** Overlay of differential expression data from the temporal cortex (TCX). Nodes are scaled by effect size and colored by effect direction (blue: down-regulated, red: up-regulated in disease). A global pattern of up- and down-regulation can be seen. **(B)** Shortest path of each entity (gene, metabolite) to an AD trait indicated by node coloring (red: path length is 1, i.e., directly associated, orange: 2-step, yellow: 3- or 4-step, blue >4 steps). Mean length of all shortest paths is 2.25. **(C)** Coloring according to membership of clusters determined by hierarchical clustering with cut h=30. **(D)** Immune response-related genes colored grey. **(E)** Dendrogram of hierarchical clustering with bands below indicating whether entities in clusters are metabolites, have been associated with AD (through association studies or differential analysis, i.e., significant up- or down-regulation in the TCX of AD patients) or have been annotated with an AD-relevant function (dark grey: yes, light grey: no). This reveals enrichment of certain biological domains in distinct regions of the visualized embedding, indicating proximity in the AD Atlas network (see **Supplementary Figure 10B**). Biological domains that are significantly (*p*-value *<* 2 × 10^-3^, Fisher’s exact test) enriched in cluster 2, which is indicated by the grey box, are emphasized in bold. In total, 23 of 25 clusters are significantly enriched for at least one biological domain (further results given in **Supplementary Table 5** and **6**).

In conclusion, these results provide evidence that the topology of the association-based multi-omics network of the AD Atlas reflects AD-relevant biological information.

## DISCUSSION

Large-scale multi-omics data holds the potential to substantially advance our understanding of molecular changes involved in the onset and progression of complex diseases like AD. However, the complexity of analyzing multiple data modalities and bringing them together in an integrated framework for biomedical research still represents a major bottleneck that requires interdisciplinary expertise. While through open science initiatives large omics datasets are widely available to the research community, democratization of processed, harmonized, high-quality data in unified resources remains an unaddressed need in many contexts.

The AD Atlas consolidates processed experimental data from large cohort studies in a predominantly hypothesis-free and data-driven network-based integration framework. The web portal provides researchers of varying backgrounds and interests with direct access to this network, along with several tools to annotate and meta-analyze context-specific subnetworks. It thus builds upon and extends previous efforts that have provided multi-omics integration solutions for multi-disease drug repositioning (*20*, *21*), genome-guided computational analysis of AD (*22*), and multi-omics annotation of individual targets (*23*). Through the user interface, the AD Atlas enables to conduct flexible and context-specific analyses tailored to the research question at hand, without the necessity of local bioinformatics capacities. We provide multiple entry points, allowing for dynamical extraction of multi-omics subnetworks surrounding a gene, metabolite or trait of interest. Alternatively, multiple entities can be analyzed together, which can reveal non-trivial connections, and networks can be expanded to explore their functional neighborhood (for genes defined by co-expression, protein co-abundance or co-regulation networks and for metabolites by partial correlation networks). By providing a large set of filtering options, such as restricting co-expression links to specific brain regions, filtering for a specific sample type or applying different significance cutoffs, users can create and explore highly context-specific networks that integrate results from various sources. Besides dynamic generation and interactive exploration of networks, we additionally interlink entities to external databases and provide downstream analysis tools, including the exploration of experimental data on differentially expressed genes and differentially abundant proteins, as well as gene set and pathway enrichment analysis. In addition to the web-based user-interface, we also provide full download of the underlying multi-omics network to enable computationally more expensive analyses.

By integrating multiple independent datasets of the same analysis type (e.g., GWAS on the same traits conducted in different cohorts), the AD Atlas builds data confidence by independent replication. Through systematic analysis of the agreement across studies, omics levels and sample types, we highlight the power of rigorously harmonized and integrated multi-omics data to globally assess comparability and complementary of information across tissues and regulatory layers. To further strengthen confidence in the derived multi-omics network, we provided evidence that the AD Atlas network captures AD-relevant biology at a global scale by projecting the network structure into a lower-dimensional space via node embeddings and integrating this data view with established AD-related gene functions and differential expression data. By offering the complete network for download, we empower users to conduct such analyses and have future plans to incorporate global analysis tools, such as the identification of disease modules in a hypothesis-free manner, into the AD Atlas web interface.

One important but non-trivial aspect in association network analysis is the selection of p-value thresholds, as unfiltered data would result in a fully connected (i.e., meaningless) network. Hence, while the database underlying the AD Atlas contains potentially relevant evidence at fine granularity (up to a raw *p*-value ≤ 0.05 where available), this data is currently not accessible through the web interface. Instead, we provide edge filtering criteria only for genetic associations, i.e., using either a genome-wide or gene-wise Bonferroni threshold, and otherwise provide data at study-specific significance thresholds. While this leads to a mixture of significance thresholds, we believe it represents an acceptable trade-off between inflation of false positive and false negative findings, as, in contrast to the established threshold for genome-wide significance, a similar universal omics-wide threshold is not available for e.g., proteomics or metabolomics. Therefore, we choose to apply study-specific significance thresholds that limit false positives to 5% per study (corrected for multiple testing) and false negatives to the rate defined by each study’s power. We also do not compare or meta-analyze additional information, such as sample size or the set of confounders used as covariates. This aims at avoiding bias due to, for example, samples included in several integrated studies (e.g., overlap in several recent AD case-control GWAS meta-analyses), or differential weighting of over-/under-studied entities, as we include associations independent of whether they have been reported in one or multiple studies.

Just like any large-scale data integration resource, the AD Atlas in its current form has several limitations. First, the underlying molecular framework is derived primarily in a data-driven way, which leads to a largely bias-free network representation of the profiled molecular data. However, given the nature of such integrative databases, selection bias to some extent is inevitable. This arises, for example, from the (manual) selection of datasets that are included in the resource and the varying coverage (entities measured) and statistical power of individual studies. We aim to mitigate this issue by continually expanding the number of integrated data and analyses as new (and larger) studies become available, thereby increasing coverage and confidence in these data-driven representations. In addition, we currently neglect a large body of curated knowledge from experts and public databases on, for example, biochemical pathways or drug targets. As we continue to extend the AD Atlas through regular update cycles, integrating such data will continuously extend the available layers of evidence. Furthermore, based on the currently limited availability of research data obtained in diverse populations/ethnicities (*24*), the data used to generate the AD Atlas are also not representative of these populations, which reduces the generalizability and transferability of the integrated molecular observations. As more data from diverse populations are becoming available, we will put a particular emphasis on their integration in the AD Atlas resource.

In summary, through rigorous harmonization and integration of processed multi-omics data, the AD Atlas represents an extensive resource that globally mirrors known biological relationships in AD. As such it is a useful tool for the formulation and further exploration of molecular hypotheses for AD research that can be validated in follow-up analysis or experiments. In addition to providing access to a large collection of curated molecular data, the resource enables customized analyses via a multi-functional web interface, which includes tools for constructing and meta-analyzing hypothesis-guided multi-omics networks. In four showcases, we demonstrate how molecular subnetworks surrounding input genes, metabolites or traits can be dynamically retrieved based on user-specified parameters, and how overlaying and analyzing these subnetworks together with differential abundance data within the web interface enables generation of new mechanistic hypotheses and identification of potential drug repositioning candidates.

## METHODS

Core methods used to build and analyze the AD Atlas and user interface are described in the following. For full details, including a comprehensive list of integrated datasets and methods for re-analysis of data, please refer to the **Supplementary Material**. All analyses were carried out using the AD Atlas v1.1.

### Data integration strategy and storage

The AD Atlas was built using step-wise data integration in which different omics datasets, such as transcriptomics, proteomics and metabolomics, are analyzed separately or in specific combinations before integration (shown schematically in **Figure 1A**). In contrast to synchronous integration, where all data is used in one analysis step, step-wise approaches allow the integration of data across many different sources and do not require the data to come from the same set of individuals/samples. We used an extended QTL-based integration strategy paired with a composite network approach, described in detail in our recent review (*7*). Briefly, relationships between biological entities (e.g., genetic variants, genes, metabolites) are either taken from public knowledge databases or inferred through statistical analysis (e.g., genome-wide association studies (GWAS) or correlation-based analysis). These individual networks are subsequently merged into a large heterogeneous network by using overlapping entities to connect multiple layers of omics data. For example, links between metabolites and genes are established via overlapping quantitative trait loci (QTLs). The resulting heterogeneous network consist of multiple node types (biological entities) that are connected by different types of edges (inferred associations between entities). To enable efficient data storage and analysis we utilized the native graph database management system Neo4j (https://neo4j.com/).

### Network-based multi-omics harmonization and integration pipeline

The construction of the AD Atlas can be divided into two phases. In the first phase, the results of available (multi-)omics studies were collected, preprocessed, integrated and stored in a detailed network structure (**Figure 1B**). In the second phase, this complex collection of data is summarized and extracted into a simplified data view (**Figure 1C**) to reduce complexity, increase interpretability of results and enable efficient data access.

#### Phase 1: Data collection and preprocessing

Known biological relationships between omics layers were downloaded from public databases, such as gene-transcript-protein mappings from Ensembl (*25*) and mappings between single nucleotide polymorphisms (SNPs) and genes from SNiPA (*26*). SNPs, genes, transcripts and proteins were stored in the database as individual nodes and relationships were added as edges between them. Furthermore, metabolites (and their corresponding meta-information, i.e., biochemical name, pathway annotations, and additional identifiers) measured across different metabolomics platforms were also collected and stored as nodes in the database. Large-scale quantitative data from population-based studies were then used to establish data-driven relationships within (e.g., tissue-specific gene co-expression) and across omics (e.g., eQTLs or mQTLs) layers. To identify entities within this network that are relevant to AD, we used large-scale association data for AD from case-control and AD biomarker GWAS, MWAS, data on differentially expressed genes and differentially abundant proteins, and brain region-specific gene co-expression and protein co-abundance. Summary statistics of these analyses were either downloaded from publicly available data repositories, supplementary data or calculated in additional analysis steps (see the **Supplementary Material** for more details). A summary of data types and their respective sources is listed in **Table 1**.

##### Data format and integration

All data were transformed into a standard data format, where one (or more) comma-separated values (CSV) files were constructed for each study, containing one relationship per row (e.g., gene 1, gene 2, p-value, etc.) and an additional CSV file containing information on the data source (e.g., publication, author, year, etc.). Data integration across studies/sources is performed by overlaying the inferred knowledge-based and data-driven relationships. Data-driven associations between two entities are modeled through an intermediate node containing the respective summary statistic (**Figure 1B**). Additionally, every statistical results node is connected to a node of type *source*, enabling a highly granular view of the data and preserving information, for example, on the cohort, publication, and sample type.

##### ID harmonization

To facilitate integration through overlap, each biological entity (e.g., gene, protein, metabolite) was mapped to a unique identifier, either using the mapping provided in the data source or through manual curation. The unique identifier for SNPs is defined as their rsID, while genes, transcripts and proteins are identified by their respective Ensembl IDs and measured metabolites are identified by their platform-specific ID. Of note, these vendor-specific metabolite labels, i.e., biochemical names assigned by the respective metabolomics platform provider, will be updated in future releases to better reflect the currently accepted notation, especially concerning lipid classes such as sphingomyelins. Additional identifiers, including biochemical metabolite names, gene symbols and UniProt identifiers, have also been annotated but are not required to be unique. AD-specific (endo-)phenotypes from different studies (GWAS, MWAS) were harmonized through manual curation and are listed in **Supplementary Table 3.** Further information regarding each phenotype is given in **Supplementary Table 4**.

#### Phase 2: Summary and abstraction

The resulting network is highly complex. To make the resource more efficient and allow a more intuitive (visual) interpretation of the results, the data model was projected into a simpler network view which is accessible via the AD Atlas user interface (www.adatlas.org, seen in **Figure 1C**). **Supplementary Figure 1** schematically shows an example of the abstraction pipeline described in the following.

##### Projection onto genes

SNPs, transcripts and encoded proteins are projected onto genes using information from Ensembl (*25*), GTEx (*27*) and SNiPA (*26*). Proteins are mapped to genes via transcripts using the mapping provided by Ensembl. For projecting SNPs to genes, we use (i) the genomic location of the SNP: if a variant is either directly located within the gene body or within 2.5kb up- or downstream, it is assigned to the respective gene(s); if a variant is located in a gene-associated regulatory element (promoters or enhancers from ENCODE (*29*) and FANTOM5 (*29*)), it is assigned to the respective associated genes; and (ii) association data: a SNP is further assigned to a gene for which significant eQTL and pQTL associations exist. Consequently, one-to-many SNP-to-gene mappings are possible. GWAS signals are therefore not limited to a single gene for each locus, but instead mirror shared genetic and epigenetic regulation to provide a comprehensive representation of each locus. The information used for this projection is partly extracted from SNiPA (*26*), and partly comes from the GTEx (*27*) or Sieberts et al. (*30*) studies. Additionally, genes displayed in the current version of the AD Atlas have been limited to protein-coding genes. The resulting meta-gene nodes further contain all relationships from the individual omics layers (e.g., from transcriptomics and proteomics analyses).

##### Metabolite consolidation

Metabolites that were measured on more than one platform were consolidated using manual mappings between platform-specific IDs and information on identified unknown metabolites to construct so-called “meta-metabolite” nodes (statistics given in **Supplementary Tables 7 and 8**). Furthermore, metabolites measured by the Metabolon platform, that were annotated with the same biochemical name and chemical ID (but differing compound ID), were also merged. A detailed example of metabolite consolidation can be seen in (**Supplementary Figure 1B**)

##### Edge summary and significance thresholds

Statistical associations between biological entities (edges) were summarized while retaining all information on the underlying SNPs and proteins. This information is stored in comprehensive edge labels. Edges are further pre-filtered using study-specific, gene-wise, or genome-wide significance thresholds. Genetic associations with AD traits (trait QTLs) and metabolites (mQTLs) can be filtered using either a genome-wide (*p*-value ≤ 5×10^-8^) or a Bonferroni-like gene-wise cutoff. The gene-wise cutoff is defined as *p*-value ≤ 0.05/#SNPs_geneA_, where #SNPs_geneA_ is the number of SNPs that have been annotated to gene_A_. A comprehensive list of study-specific significance thresholds is given in **Supplementary Table 2**.

### Definition of inferred edge types

Different edge types categorize statistically significant associations/correlations between two entities, represented as an edge in the multi-omics network. In the context of the AD Atlas, they are defined as following:

***Co-regulation –*** inferred from expression quantitative trait loci (eQTL studies) to inform about shared genetic and epigenetic regulation of gene or protein expression. Edges link two or more genes mostly within one locus, but can also be in trans. Interpretation examples of an edge (A)-[:COREGULATION]-(B) include:

*A SNP is an eQTL for both gene A and gene B;*
*A SNP is located in gene A and is an eQTL for gene B;*
*A SNP is located in a promoter/enhancer linked to gene A and is an eQTL for gene B;*

***Genetic association –*** inferred from GWAS and MWAS to inform about genetic risk for AD and metabolite quantitative trait loci (mQTL). Edges link genes with AD-related traits or metabolic traits. Interpretation examples of an edge (A)-[:GENETIC ASSOCIATION]-(B) include:

*A SNP is located in gene A and is a QTL for (AD or metabolic) trait B;*
*A SNP is an eQTL for gene A and is a QTL for (AD or metabolic) trait B;*
*A SNP is located in a promoter/enhancer linked to gene A and QTL for (AD or metabolic) trait B;*

***Metabolic association –*** inferred from metabolome-wide association studies (MWAS). Edges link metabolites with AD-related traits and indicate a significant association.

***Co-expression –*** inferred from gene co-expression analysis. An edge indicates a statistically significant positive or negative correlation between the levels of two transcripts.

***Co-abundance –*** inferred from gaussian graphical models (GGMs) using proteomics data. Edges link two or more genes and indicate a significant positive or negative partial correlation between the abundance of proteins that are encoded by the genes.

***Partial correlation –*** inferred from GGMs using metabolomics data. An edge indicates a significant positive or negative partial correlation between the levels of two metabolites.

### Calculation of pairwise network similarity through edge overlap

To quantify the concordance of two omics- and study-specific networks A and B we used two different measures; the Jaccard index and the Overlap similarity. The measures are defined as following:

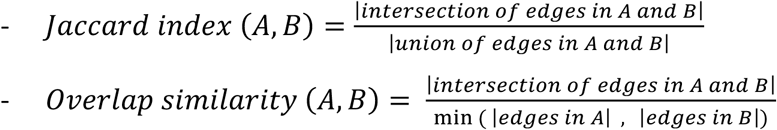

Values of both metrics range between 0-1, with 0 indicating no overlap and 1 indicating complete overlap (for the Jaccard index) or the complete inclusion of edges of the smaller set in the larger set (for Overlap similarity). It is worth noting that these measures only consider the presence or absence of associations, without considering their strength or direction. Meta-trait-gene (GWAS) networks were constructed by mapping all traits to meta-traits (as defined in **Supplementary Table 3**). This allowed the comparison of broader trait categories. Before each pairwise comparison, the networks were filtered for overlapping meta-traits, that is, we only included genome-wide significant edges between genes and meta-traits that were analyzed in both studies to ensure valid comparisons. In a similar fashion, gene-gene networks were filtered for common genes before comparison, i.e., only significant edges between genes that have been investigated in both studies were included in each pairwise comparison. This was done to account for discrepancies introduced through different sets of entities measured by different omics technologies. Comparison of metabolomics-related data was conducted analogously. Metabolite-gene (mGWAS) networks were filtered for (pairwise) common metabolites prior to comparison and only genome-wide significant edges were included. Metabolite-metabolite networks (GGM) were filtered for common metabolites before comparison. Lastly, trait-metabolite (MWAS) networks were compared at the meta-trait-level, that is, all traits were mapped to meta-traits as described above. Before each pairwise comparison, the networks were filtered for common meta-traits and metabolites.

### Network representation learning and visualization of global network structure

Network representation learning on the global AD Atlas network was performed using a slightly modified version of the node embedding framework implemented in the GeneWalk Python package. Details of this approach can be found elsewhere (*13*, *31*). Briefly, vector representations of nodes are learnt by sampling unbiased random walks over the network and using neighboring node pairs from these walk sequences as input-output training sets for a fully connected neural network. The package also enables the generation and embedding of randomized networks that retain the same degree distribution which was used to compare structures seen in the AD Atlas (**Supplementary Figure 10A**). A simplified graph was abstracted from the AD Atlas, i.e., an undirected graph that includes no self-loops and multiple edges between two nodes are only represented by one edge. Furthermore, no distinction was made between the different node and edge types as this information cannot be taken into account by the algorithm. As we were interested to see if the global structure of the molecular network provides relevant biological information related to AD, we excluded AD phenotype nodes from the network. A brain-specific network was constructed by restricting the sample type of all edges to ‘brain’ (protein co-abundance, metabolite partial correlation, metabolite-trait associations and mQTL data). A genome-wide significance filter (*p*-value ≤ 5×10^-8^) was applied to genetic associations. As the SNP-to-gene annotation underlying QTL-based edges incorporates eQTL information, we did not include co-regulation edges in the network to avoid overweighting this data modality (**Supplementary Figure 10C)**. GeneWalk was run setting the window size (context surrounding each node that is regarded for embedding) to 2 and the dimension of the vector representations (embeddings) to 130 (–dim rep=130). The appropriate embedding dimension was determined by using the approach described in (*32*). As this method uses the embedding algorithm Node2Vec (*32*) we set the parameters *p* and *q* both to 1 and the *window_size* to 2 to make the embedding algorithm similar to the embedding algorithm used by GeneWalk (DeepWalk (*31*)). The resulting node vectors were projected to two dimensions for visualization using Uniform Manifold Approximation and Projection (UMAP) (*35*) as implemented in the Python library *umap* (settings: n_neighbors=15, min_dist=0.1, metric: ‘cosine’). Hierarchical clustering and visualization as a dendrogram were performed using the R packages *stats* (v4.2.0) and *dendextend* (v1.16.0). The 130-dimensional node vectors were used as input and the method “ward.D2” with Euclidean distance was used for clustering.

### Alzheimer’s disease biological domain information

For AD-relevant gene function annotation, over 8200 GO term definitions were categorized into 16 AD-related biological domains: Synaptic Function, Immune Response, Endolysosomal Trafficking, Structural Stabilization, RNA Spliceosome, Myelination, Vascular Function, Mitochondria and Metabolism, Autophagy, Apoptosis, Epigenetics, Oxidative Stress, Lipid Metabolism, APP Processing, Tau homeostasis, Proteostasis. This manual curation effort currently covers 54% of all genes but will be continuously expanded in the future. Further information on the approach and the complete mapping of genes and GO terms to biological domains is available at https://www.synapse.org/#!Synapse:syn26529354.

### Enrichment analysis

The AD Atlas offers access to a variety of different tools that enable enrichment analysis of the generated subnetworks. Gene set enrichment can be conducted using the R package *enrichR* (v3.0) (*33*), which enables the analysis of a variety of different gene sets, including drug perturbation signatures and biochemical pathways, via the Enrichr webservice (https://maayanlab.cloud/Enrichr/). Here, the Benjamini-Hochberg (BH) method is used to account for multiple testing and the corrected *p*-values are reported as *q*-value. Enrichr does not allow the definition of a custom background list. Additionally, enrichment can be performed using the R package *gprofiler2* (v0.2.0) (*34*), which provides an interface to the gene list functional profiling toolset, g:Profiler (https://biit.cs.ut.ee/gprofiler/gost). The user can specify the correction method applied for multiple testing; the background set consists of all protein coding genes included in the AD Atlas. Furthermore, we provide access to Rummagene (https://rummagene.com/) (*35*) and RummaGEO (https://rummageo.com/) (36) utilizing their GraphQL API. Classical GO term enrichment analysis is provided via the R package *topGO* (v2.38.1) (*37*). As background list, again, all protein coding genes included in the AD Atlas are used. Metabolic pathway enrichment analysis for metabolites included in the constructed molecular subnetworks is performed using platform-specific annotations of metabolites into classes and super-/sub-pathways. Contingency tables are calculated and then tested using Fisher’s exact test (function *fisher.test* with alternative = ‘greater’ from the R package *stats*). The same method is used to test for enrichment of AD biological domains and differentially expressed genes and proteins. For the latter, up-regulated, unchanged and down-regulated genes are treated as individual gene sets. Multiple testing correction is performed using *p.adjust* (method = ‘fdr’). As background, we use all metabolites for which pathway information is available and all genes/proteins that have been annotated with biological domains or tested for differential expression.

### Technical framework

The integrated multi-omics data is stored as a heterogeneous graph using the graph database management system Neo4j (community v4.4.11). To enable easy access to the data, we implemented a network- and web-based user interface to the AD Atlas, reachable via www.adatlas.org. This frontend was built as a ShinyApp using R v3.6.1 and is deployed using ShinyProxy (v2.3.1_amd64.deb). Communication between Neo4j and R is established through the official Neo4j python driver *neo4j* (https://github.com/neo4j/neo4j-python-driver) and the R-to-python interface package *reticulate* (v1.17). Molecular subnetworks are constructed by translating the user-specified options into a cypher query that is subsequently used to communicate with the underlying Neo4j graph database. The resulting data matrices, one with node information and one containing the edges, are processed using R and finally visualized using *VisNetwork* (v2.0.9). A detailed example query is shown in **Supplementary Figure 11**.

All tools used are publicly available and free for academic use. In detail, we used the following:

#### Data storage and app deployment

- **Neo4j**: Version: v4.4.11 (community). Available from: https://neo4j.com
- **Docker**: Version 1.13.1, Build cccb291/1.13.1. Available from: https://www.docker.com
- **ShinyProxy**: Version 2.3.1_amd64.deb. Available from: https://www.shinyproxy.io/

#### User interface - R packages

- **shiny**: Version: 1.6.0. Web application framework for R.
- **shinyjs**: Version: 2.0.0. Easily improve the user experience of shiny apps in seconds.
- **scroller**: Version: 0.1.1. Scroll to any element in Shiny.
- **shinyEffects**: Version: 0.2.0. Customize your web apps with fancy effects.
- **shinybusy**: Version: 0.3.1. Busy indicator for ‘Shiny’ applications.
- **shinydashboard**: Version: 0.7.1. Create dashboards with ‘Shiny’.
- **shinydashboardPlus**: Version: 0.7.0. Add more ‘AdminLTE2’components to ‘shinydashboard’.
- **shinyWidgets**: Version: 0.6.0. Custom inputs widgets for shiny.
- **shinycustomloader**: Version: 0.9.0. Custom loader for shiny outputs.
- **particlesjs:** Version: 0.1.1 Beautiful background for ‘Shiny’ Applications and ‘Rmarkdown’ Documents

#### Data visualization - R packages

After preprocessing of the edges and nodes data.frame returned from the user defined cypher query (**Supplementary Figure 11**), the *igraph* function *degree* is used to extract the node degrees (used for scaling of node size) and the functions *layout_with_drl*, *layout_with_graphopt* from the *igraph* package or *qgraph.layout.fruchtermanreingold* from the *qgraph* package is used to precalculate the network layout. Depending on user input, this is either done for the whole network (**Include traits in layout=yes**) or excluding trait nodes and arranging these separately, to the right of the layout (**Include traits in layout=no**). Finally, the visualization of interactive networks is performed using the R package *VisNetwork*. Network representation as hive plots is rendered with *HiveR,* 3D networks are rendered with *threejs* and interactive bar- and donut-plots of the network statistics are realized using *plotly*. The package *DT* is used to create HTML table widgets that allow users to sort, search and download tables.

- **VisNetwork**: Version: 2.0.9. Network visualization using ‘vis.js’ library.
- **qgraph**: Version 1.6.5. Graph plotting methods and psychometric data visualization.
- **igraph**: Version 1.2.4.1. Network analysis and visualization.
- **threejs**: Version 0.3.3. Interactive 3D scatter plots, networks and globes.
- **plotly**: Version 4.9.4.1. Creative interactive web graphics via ‘plotly.js’.
- **DT**: Version: 0.16. A wrapper of the JavaScript library ‘DataTables’.
- **HiveR**: Version: 0.3.63. 2D and 3D Hive plots for R.

#### Enrichment analysis – R packages

- **topGO**: Version: 2.38.1. Enrichment analysis for Gene Ontology.
- **enrichR:** Version: 3.0. Provides an R interface to ‘Enrichr’.
- **gprofiler2:** Version: 0.2.1 Provides an R interface to ‘g:Profiler’ toolset.

### Integrated datasets

#### Public databases and population-based studies

- **Ensembl** (PMID: 30407521)
- **SNiPA** (PMID: 2543133)
- **Tissue-specific gene regulation**

- <u>GTEx v8</u> - The Genotype-Tissue Expression (GTEx) Project (doi: 10.1101/787903)
- <u>Sieberts et al.</u> – Large brain eQTL meta-analysis (AD Knowledge Portal: https://adknowledgeportal.synapse.org/Explore/Studies/DetailsPage?Study=syn25398075)
- **Genetic associations with metabolic traits** Summary statistics from genome-wide association studies (GWAS) with metabolic traits were downloaded from *[SNiPA](https://snipa.helmholtz-muenchen.de/snipa3/)* (https://snipa.org) and the *[mGWAS server](http://metabolomics.helmholtz-muenchen.de/gwas/index.php)* (https://metabolomics.helmholtz-muenchen.de/gwas). This includes the following publications:

- <u>Raffler et al.</u> - Loci of urinary human metabolic individuality (PMID: 26352407)
- <u>Shin et al.</u> - Atlas of genetic influences on blood metabolites (PMID: 24816252)
- <u>Long et al.</u> **-** Genetic factors modifying the blood metabolome (PMID: 28263315)
- <u>Draisma et al.</u> - Genetic variants contributing to variation in blood metabolite levels (PMID: 26068415)
- <u>Suhre et al.</u> - Human metabolic individuality (PMID: 21886157)
- **Partial correlation networks**

- <u>Krumsiek et al.</u> - A systems approach to metabolite identification (PMID: 23093944)
- <u>Suhre et al.</u> - GWAS with the human blood plasma proteome (PMID: 28240269)

#### Alzheimer’s disease-related associations

- **Genetic associations with Alzheimer’s (endo-)phenotypes**

- <u>Beecham et al.</u> - GWAS Meta-analysis of Neuropathologic Features of AD (PMID: 25188341) NIAGADS id: https://www.niagads.org/datasets/ng00041
- <u>Deming et al.</u> - GWAS with cerebrospinal fluid clusterin (PMID: 26545630) NIAGADS id: https://www.niagads.org/datasets/ng00052
- <u>Deming et al.</u> - GWAS with Alzheimer’s endophenotypes (PMID: 28247064) NIAGADS id: https://www.niagads.org/datasets/ng00055
- <u>Lambert et al.</u> - Two-stage meta-analysis of GWAS with late-onset AD (PMID: 24162737) NIAGADS id: https://www.niagads.org/datasets/ng00036
- <u>Huang et al.</u> - GWAS with AD age of onset (PMID: 28628103) NIAGADS id: https://www.niagads.org/datasets/ng00058
- <u>Marioni et al.</u> - GWAS on family history of Alzheimer’s disease (PMID: 29777097) Data link: https://www.ccace.ed.ac.uk/node/335
- <u>Kunkle et al.</u> - Genetic meta-analysis of diagnosed Alzheimer’s disease (PMID: 30820047) NIAGADS id: https://www.niagads.org/datasets/ng00075
- <u>Jansen et al.</u> - Genetic meta-analysis using clinically diagnosed and proxy AD (PMID: 30617256) Data: https://ctg.cncr.nl/software/summary_statistics
- <u>Wightman et al.</u> - Genetic meta-analysis using clinically diagnosed and proxy AD (PMID: 34493870) Data: https://ctg.cncr.nl/software/summary_statistics
- <u>Bellenguez et al.</u> - Genetic meta-analysis using clinically diagnosed and proxy AD (PMID: 30820047) Data: *GWAS catalog (GCST90027158)*
- **Brain co-expression networks**

- <u>Wan et al</u>. - Meta-analysis of the human brain transcriptome (doi: 10.1016/j.celrep.2020.107908) Synapse id: https://www.synapse.org/#!Synapse:syn12523173
- **Brain partial correlation networks**

- <u>Batra et al.</u> **-** The landscape of metabolic brain alterations in Alzheimer’s disease (PMID: 35829654)
- **Differential analysis in AD cohorts**

- <u>Wan et al.</u> - Meta-analysis of the human brain transcriptome (doi: 10.1016/j.celrep.2020.107908) Synapse id (brain-region specific): https://www.synapse.org/#!Synapse:syn12177499 Synapse id (meta-analysis): https://doi.org/10.7303/syn11914606
- <u>Johnson et al.</u> - Consensus Proteomic Analysis of Alzheimer’s Disease Brain (doi: 10.1101/802959) Synapse id (study): https://www.synapse.org/#!Synapse:syn20933797
- <u>Johnson et al.</u> – Large-scale deep multi-layer analysis of Alzheimer’s disease brain (doi:10.1038/s41593-021-00999-y) Synapse id (study): https://doi.org/10.7303/syn25006611
- **Metabolic associations with Alzheimer’s (endo-)phenotypes**

- <u>MahmoudianDehkordi et al.</u> and <u>Nho et al.</u> - Bile acid MWAS with AD biomarkers (PMID: 30337151)
- <u>Arnold et al.</u> - MWAS with AD biomarkers (PMID: 32123170)
- <u>Batra et al.</u> **-** The landscape of metabolic brain alterations in Alzheimer’s disease (PMID: 35829654)

#### Additional (data re-) analysis

- **Alzheimer’s disease biological domain information**

- Complete mapping of genes and GO terms to biological domains is available at https://www.synapse.org/#!Synapse:syn26529354.
- **Partial correlation networks**

- <u>Johnson et al.</u> - Consensus Proteomic Analysis of Alzheimer’s Disease Brain (doi: 10.1101/802959) Synapse id (study): https://www.synapse.org/#!Synapse:syn20933797 Synapse id (data): https://www.synapse.org/#!Synapse:syn21449534
- <u>Johnson et al.</u> - Large-scale deep multi-layer analysis of Alzheimer’s disease brain (doi:10.1038/s41593-021-00999-y) Synapse id (study): https://doi.org/10.7303/syn25006611 Synapse id (data): https://www.synapse.org/#!Synapse:syn25006802
- **Genetic associations with brain metabolites**

- Subset of data used in Batra et al. (*38*) from participants of the ROS/MAP cohorts and the Mayo brain bank (unpublished data).
- **Genetic associations with blood metabolites and Alzheimer’s (endo)phenotypes**

- <u>Alzheimer’s Disease Neuroimaging Initiative (ADNI)</u> Data used in the preparation of this article were obtained from the Alzheimer’s Disease Neuroimaging Initiative (ADNI) database (*adni.loni.usc.edu*). The ADNI was launched in 2003 as a public-private partnership, led by Principal Investigator Michael W. Weiner, MD. The primary goal of ADNI has been to test whether serial magnetic resonance imaging (MRI), positron emission tomography (PET), other biological markers, and clinical and neuropsychological assessment can be combined to measure the progression of mild cognitive impairment (MCI) and early Alzheimer’s disease (AD). For up-to-date information, see www.adni-info.org. Metabolomics data are available via the Alzheimer’s Disease (AD) Knowledge Portal (https://adknowledgeportal.org):

▪ AbsoluteIDQ® p180 kit (Biocrates Life Sciences AG, Innsbruck, Austria): https://doi.org/10.7303/syn5592519 (ADNI-1) https://doi.org/10.7303/syn9705278 (ADNI-GO/-2)
▪ Bile Acids Kit (Biocrates Life Sciences AG, Innsbruck, Austria): https://doi.org/10.7303/syn12036817.1 (ADNI-1) https://doi.org/10.7303/syn9779093.1 (ADNI-GO/2)

## Author contributions

*All authors read and approved the final manuscript*.

Conceptualization: MAW, JK, GK, MA; data analysis and resource development: MAW, JK, SN, KN, AKG, JCW, LLW, TW, YANN, KH, OB, BM, PW, WRM, NL, GK, MA; funding acquisition: RKD, CVD, GK, MA; visualization: MAW; supervision: RKD, GK, MA; writing - original draft: MAW, SN, PMD, GK, MA; writing - review & editing: JK, AKG, JB, PJM, AJS, CVD, KS; all authors read and approved the final manuscript.

## Data and materials availability

All data needed to evaluate the conclusions in the paper are present in the paper and/or the **Supplementary Materia**l. The AD Atlas is accessible via the user interface at adatlas.org. A comprehensive listing of the exact data sources that were integrated in the AD Atlas is given in the **Supplementary Material**. The results published here are in whole or in part based on data obtained from the AD Knowledge Portal (https://adknowledgeportal.org). Data used in the preparation of this article were further obtained from the Alzheimer’s Disease Neuroimaging Initiative (ADNI) database (https://adni.loni.usc.edu). A standalone version of each release of the AD Atlas shiny app with a data freeze of the Neo4j database (abstracted data model) will be made available upon publication on adatlas.org and in the *AD Knowledge Portal*. The results presented in this paper are based on version 1.1 of the AD Atlas.

## REFERENCES

1. J. M. Long, D. M. Holtzman, Alzheimer Disease: An Update on Pathobiology and Treatment Strategies. Cell 179, 312–339 (2019).

2. G. M. Sancesario, S. Bernardini, Alzheimer’s disease in the omics era. Clin. Biochem. 59, 9–16 (2018).

3. R. J. Hodes, N. Buckholtz, Accelerating Medicines Partnership: Alzheimer’s Disease (AMP-AD) Knowledge Portal Aids Alzheimer’s Drug Discovery through Open Data Sharing. Expert Opin. Ther. Targets 20, 389–391 (2016).

4. A. K. Greenwood, K. S. Montgomery, N. Kauer, K. H. Woo, Z. J. Leanza, W. L. Poehlman, J. Gockley, S. K. Sieberts, L. Bradic, B. A. Logsdon, M. A. Peters, L. Omberg, L. M. Mangravite, The AD Knowledge Portal: A Repository for Multi-Omic Data on Alzheimer’s Disease and Aging. Curr. Protoc. Hum. Genet. 108, e105 (2020).

5. A. Lysenko, I. A. Roznovăţ, M. Saqi, A. Mazein, C. J. Rawlings, C. Auffray, Representing and querying disease networks using graph databases. BioMed Central [Preprint] (2016). 10.1186/s13040-016-0102-8.

6. B. Lee, S. Zhang, A. Poleksic, L. Xie, Heterogeneous Multi-Layered Network Model for Omics Data Integration and Analysis. Front. Genet. 10, 1381 (2019).

7. M. A. Wörheide, J. Krumsiek, G. Kastenmüller, M. Arnold, Multi-omics integration in biomedical research - A metabolomics-centric review. Anal. Chim. Acta 1141, 144–162 (2021).

8. K. Suhre, M. Arnold, A. M. Bhagwat, R. J. Cotton, R. Engelke, J. Raffler, H. Sarwath, G. Thareja, A. Wahl, R. K. Delisle, L. Gold, M. Pezer, G. Lauc, M. A. E. D. Selim, D. O. Mook-Kanamori, E. K. Al-Dous, Y. A. Mohamoud, J. Malek, K. Strauch, H. Grallert, A. Peters, G. Kastenmüller, C. Gieger, J. Graumann, Connecting genetic risk to disease end points through the human blood plasma proteome. Nat. Commun. 8 (2017).

9. P. Baloni, M. Arnold, L. Buitrago, K. Nho, H. Moreno, K. Huynh, B. Brauner, G. Louie, A. Kueider-Paisley, K. Suhre, A. J. Saykin, K. Ekroos, P. J. Meikle, L. Hood, N. D. Price, Alzheimer’s Disease Metabolomics Consortium, P. M. Doraiswamy, C. C. Funk, A. I. Hernández, G. Kastenmüller, R. Baillie, X. Han, R. Kaddurah-Daouk, Multi-Omic analyses characterize the ceramide/sphingomyelin pathway as a therapeutic target in Alzheimer’s disease. Commun Biol 5, 1074 (2022).

10. M. Pietzner, E. Wheeler, J. Carrasco-Zanini, A. Cortes, M. Koprulu, M. A. Wörheide, E. Oerton, J. Cook, I. D. Stewart, N. D. Kerrison, J. Luan, J. Raffler, M. Arnold, W. Arlt, S. O’Rahilly, G. Kastenmüller, E. R. Gamazon, A. D. Hingorani, R. A. Scott, N. J. Wareham, C. Langenberg, Mapping the proteo-genomic convergence of human diseases. Science 374, eabj1541 (2021).

11. N. Ruffini, S. Klingenberg, R. Heese, S. Schweiger, S. Gerber, The big picture of neurodegeneration: A meta study to extract the essential evidence on neurodegenerative diseases in a network-based approach. Front. Aging Neurosci. 14, 866886 (2022).

12. C. R. Jack Jr, D. A. Bennett, K. Blennow, M. C. Carrillo, B. Dunn, S. B. Haeberlein, D. M. Holtzman, W. Jagust, F. Jessen, J. Karlawish, E. Liu, J. L. Molinuevo, T. Montine, C. Phelps, K. P. Rankin, C. C. Rowe, P. Scheltens, E. Siemers, H. M. Snyder, R. Sperling, Contributors, NIA-AA Research Framework: Toward a biological definition of Alzheimer’s disease. Alzheimers. Dement. 14, 535–562 (2018).

13. R. Ietswaart, B. M. Gyori, J. A. Bachman, P. K. Sorger, L. S. Churchman, GeneWalk identifies relevant gene functions for a biological context using network representation learning. Genome Biol. 22, 55 (2021).

14. W. L. Hamilton, R. Ying, J. Leskovec, Representation Learning on Graphs: Methods and Applications, arXiv [cs.SI] (2017)pp. 1–24.

15. W. Nelson, M. Zitnik, B. Wang, J. Leskovec, A. Goldenberg, R. Sharan, To Embed or Not: Network Embedding as a Paradigm in Computational Biology. Front. Genet. 10, 381 (2019).

16. H. Cai, V. W. Zheng, K. C.-C. Chang, A Comprehensive Survey of Graph Embedding: Problems, Techniques, and Applications. IEEE Trans. Knowl. Data Eng. 30, 1616–1637 (2018).

17. W. Gu, A. Tandon, Y.-Y. Ahn, F. Radicchi, Principled approach to the selection of the embedding dimension of networks. Nat. Commun. 12, 3772 (2021).

18. L. McInnes, J. Healy, J. Melville, UMAP: Uniform Manifold Approximation and Projection for Dimension Reduction. arXiv [stat.ML] (2018).

19. Y. Shi, D. M. Holtzman, Interplay between innate immunity and Alzheimer disease: APOE and TREM2 in the spotlight. Nat. Rev. Immunol. 18, 759–772 (2018).

20. D. S. Himmelstein, S. E. Baranzini, Heterogeneous Network Edge Prediction: A Data Integration Approach to Prioritize Disease-Associated Genes. PLoS Comput. Biol. 11, e1004259 (2015).

21. D. S. Himmelstein, A. Lizee, C. Hessler, L. Brueggeman, S. L. Chen, D. Hadley, A. Green, P. Khankhanian, S. E. Baranzini, Systematic integration of biomedical knowledge prioritizes drugs for repurposing. Elife 6, 1–35 (2017).

22. E. Sügis, J. Dauvillier, A. Leontjeva, P. Adler, V. Hindie, T. Moncion, V. Collura, R. Daudin, Y. Loe-Mie, Y. Herault, J.-C. Lambert, H. Hermjakob, T. Pupko, J.-C. Rain, I. Xenarios, J. Vilo, M. Simonneau, H. Peterson, HENA, heterogeneous network-based data set for Alzheimer’s disease. Sci Data 6, 151 (2019).

23. Y. Zhou, J. Fang, L. M. Bekris, Y. H. Kim, A. A. Pieper, J. B. Leverenz, J. Cummings, F. Cheng, AlzGPS: a genome-wide positioning systems platform to catalyze multi-omics for Alzheimer’s drug discovery. Alzheimers. Res. Ther. 13, 24 (2021).

24. C. Reitz, M. A. Pericak-Vance, T. Foroud, R. Mayeux, A global view of the genetic basis of Alzheimer disease. Nat. Rev. Neurol. 19, 261–277 (2023).

25. F. Cunningham, P. Achuthan, W. Akanni, J. Allen, M. R. Amode, I. M. Armean, R. Bennett, J. Bhai, K. Billis, S. Boddu, C. Cummins, C. Davidson, K. J. Dodiya, A. Gall, C. G. Girón, L. Gil, T. Grego, L. Haggerty, E. Haskell, T. Hourlier, O. G. Izuogu, S. H. Janacek, T. Juettemann, M. Kay, M. R. Laird, I. Lavidas, Z. Liu, J. E. Loveland, J. C. Marugán, T. Maurel, A. C. McMahon, B. Moore, J. Morales, J. M. Mudge, M. Nuhn, D. Ogeh, A. Parker, A. Parton, M. Patricio, A. I. Abdul Salam, B. M. Schmitt, H. Schuilenburg, D. Sheppard, H. Sparrow, E. Stapleton, M. Szuba, K. Taylor, G. Threadgold, A. Thormann, A. Vullo, B. Walts, A. Winterbottom, A. Zadissa, M. Chakiachvili, A. Frankish, S. E. Hunt, M. Kostadima, N. Langridge, F. J. Martin, M. Muffato, E. Perry, M. Ruffier, D. M. Staines, S. J. Trevanion, B. L. Aken, A. D. Yates, D. R. Zerbino, P. Flicek, Ensembl 2019. Nucleic Acids Res. 47, D745–D751 (2019).

26. M. Arnold, J. Raffler, A. Pfeufer, K. Suhre, G. Kastenmüller, SNiPA: an interactive, genetic variant-centered annotation browser. Bioinformatics 31, 1334–1336 (2015).

27. GTEx Consortium, The GTEx Consortium atlas of genetic regulatory effects across human tissues. Science 369, 1318–1330 (2020).

28. R. E. Thurman, E. Rynes, R. Humbert, J. Vierstra, M. T. Maurano, E. Haugen, N. C. Sheffield, A. B. Stergachis, H. Wang, B. Vernot, K. Garg, S. John, R. Sandstrom, D. Bates, L. Boatman, T. K. Canfield, M. Diegel, D. Dunn, A. K. Ebersol, T. Frum, E. Giste, A. K. Johnson, E. M. Johnson, T. Kutyavin, B. Lajoie, B.-K. Lee, K. Lee, D. London, D. Lotakis, S. Neph, F. Neri, E. D. Nguyen, H. Qu, A. P. Reynolds, V. Roach, A. Safi, M. E. Sanchez, A. Sanyal, A. Shafer, J. M. Simon, L. Song, S. Vong, M. Weaver, Y. Yan, Z. Zhang, Z. Zhang, B. Lenhard, M. Tewari, M. O. Dorschner, R. S. Hansen, P. A. Navas, G. Stamatoyannopoulos, V. R. Iyer, J. D. Lieb, S. R. Sunyaev, J. M. Akey, P. J. Sabo, R. Kaul, T. S. Furey, J. Dekker, G. E. Crawford, J. A. Stamatoyannopoulos, The accessible chromatin landscape of the human genome. Nature 489, 75–82 (2012).

29. FANTOM Consortium and the RIKEN PMI and CLST (DGT), A. R. R. Forrest, H. Kawaji, M. Rehli, J. K. Baillie, M. J. L. de Hoon, V. Haberle, T. Lassmann, I. V. Kulakovskiy, M. Lizio, M. Itoh, R. Andersson, C. J. Mungall, T. F. Meehan, S. Schmeier, N. Bertin, M. Jørgensen, E. Dimont, E. Arner, C. Schmidl, U. Schaefer, Y. A. Medvedeva, C. Plessy, M. Vitezic, J. Severin, C. A. Semple, Y. Ishizu, R. S. Young, M. Francescatto, I. Alam, D. Albanese, G. M. Altschuler, T. Arakawa, J. A. C. Archer, P. Arner, M. Babina, S. Rennie, P. J. Balwierz, A. G. Beckhouse, S. Pradhan-Bhatt, J. A. Blake, A. Blumenthal, B. Bodega, A. Bonetti, J. Briggs, F. Brombacher, A. M. Burroughs, A. Califano, C. V. Cannistraci, D. Carbajo, Y. Chen, M. Chierici, Y. Ciani, H. C. Clevers, E. Dalla, C. A. Davis, M. Detmar, A. D. Diehl, T. Dohi, F. Drabløs, A. S. B. Edge, M. Edinger, K. Ekwall, M. Endoh, H. Enomoto, M. Fagiolini, L. Fairbairn, H. Fang, M. C. Farach-Carson, G. J. Faulkner, A. V. Favorov, M. E. Fisher, M. C. Frith, R. Fujita, S. Fukuda, C. Furlanello, M. Furino, J.-I. Furusawa, T. B. Geijtenbeek, A. P. Gibson, T. Gingeras, D. Goldowitz, J. Gough, S. Guhl, R. Guler, S. Gustincich, T. J. Ha, M. Hamaguchi, M. Hara, M. Harbers, J. Harshbarger, A. Hasegawa, Y. Hasegawa, T. Hashimoto, M. Herlyn, K. J. Hitchens, S. J. Ho Sui, O. M. Hofmann, I. Hoof, F. Hori, L. Huminiecki, K. Iida, T. Ikawa, B. R. Jankovic, H. Jia, A. Joshi, G. Jurman, B. Kaczkowski, C. Kai, K. Kaida, A. Kaiho, K. Kajiyama, M. Kanamori-Katayama, A. S. Kasianov, T. Kasukawa, S. Katayama, S. Kato, S. Kawaguchi, H. Kawamoto, Y. I. Kawamura, T. Kawashima, J. S. Kempfle, T. J. Kenna, J. Kere, L. M. Khachigian, T. Kitamura, S. P. Klinken, A. J. Knox, M. Kojima, S. Kojima, N. Kondo, H. Koseki, S. Koyasu, S. Krampitz, A. Kubosaki, A. T. Kwon, J. F. J. Laros, W. Lee, A. Lennartsson, K. Li, B. Lilje, L. Lipovich, A. Mackay-Sim, R.-I. Manabe, J. C. Mar, B. Marchand, A. Mathelier, N. Mejhert, A. Meynert, Y. Mizuno, D. A. de Lima Morais, H. Morikawa, M. Morimoto, K. Moro, E. Motakis, H. Motohashi, C. L. Mummery, M. Murata, S. Nagao-Sato, Y. Nakachi, F. Nakahara, T. Nakamura, Y. Nakamura, K. Nakazato, E. van Nimwegen, N. Ninomiya, H. Nishiyori, S. Noma, S. Noma, T. Noazaki, S. Ogishima, N. Ohkura, H. Ohimiya, H. Ohno, M. Ohshima, M. Okada-Hatakeyama, Y. Okazaki, V. Orlando, D. A. Ovchinnikov, A. Pain, R. Passier, M. Patrikakis, H. Persson, S. Piazza, J. G. D. Prendergast, O. J. L. Rackham, J. A. Ramilowski, M. Rashid, T. Ravasi, P. Rizzu, M. Roncador, S. Roy, M. B. Rye, E. Saijyo, A. Sajantila, A. Saka, S. Sakaguchi, M. Sakai, H. Sato, S. Savvi, A. Saxena, C. Schneider, E. A. Schultes, G. G. Schulze-Tanzil, A. Schwegmann, T. Sengstag, G. Sheng, H. Shimoji, Y. Shimoni, J. W. Shin, C. Simon, D. Sugiyama, T. Sugiyama, M. Suzuki, N. Suzuki, R. K. Swoboda, P. A. C. ’t Hoen, M. Tagami, N. Takahashi, J. Takai, H. Tanaka, H. Tatsukawa, Z. Tatum, M. Thompson, H. Toyodo, T. Toyoda, E. Valen, M. van de Wetering, L. M. van den Berg, R. Verado, D. Vijayan, I. E. Vorontsov, W. W. Wasserman, S. Watanabe, C. A. Wells, L. N. Winteringham, E. Wolvetang, E. J. Wood, Y. Yamaguchi, M. Yamamoto, M. Yoneda, Y. Yonekura, S. Yoshida, S. E. Zabierowski, P. G. Zhang, X. Zhao, S. Zucchelli, K. M. Summers, H. Suzuki, C. O. Daub, J. Kawai, P. Heutink, W. Hide, T. C. Freeman, B. Lenhard, V. B. Bajic, M. S. Taylor, V. J. Makeev, A. Sandelin, D. A. Hume, P. Carninci, Y. Hayashizaki, A promoter-level mammalian expression atlas. Nature 507, 462–470 (2014).

30. S. K. Sieberts, T. M. Perumal, M. M. Carrasquillo, M. Allen, J. S. Reddy, G. E. Hoffman, K. K. Dang, J. Calley, P. J. Ebert, J. Eddy, X. Wang, A. K. Greenwood, S. Mostafavi, CommonMind Consortium (CMC), The AMP-AD Consortium, L. Omberg, M. A. Peters, B. A. Logsdon, P. L. De Jager, N. Ertekin-Taner, L. M. Mangravite, Large eQTL meta-analysis reveals differing patterns between cerebral cortical and cerebellar brain regions. Sci Data 7, 340 (2020).

31. B. Perozzi, R. Al-Rfou, S. Skiena, “DeepWalk: online learning of social representations” in Proceedings of the 20th ACM SIGKDD International Conference on Knowledge Discovery and Data Mining (Association for Computing Machinery, New York, NY, USA, 2014)KDD ‘14, pp. 701–710.

32. A. Grover, J. Leskovec, node2vec: Scalable Feature Learning for Networks. KDD 2016, 855–864 (2016).

33. Z. Xie, A. Bailey, M. V. Kuleshov, D. J. B. Clarke, J. E. Evangelista, S. L. Jenkins, A. Lachmann, M. L. Wojciechowicz, E. Kropiwnicki, K. M. Jagodnik, M. Jeon, A. Ma’ayan, Gene Set Knowledge Discovery with Enrichr. Curr Protoc 1, e90 (2021).

34. L. Kolberg, U. Raudvere, I. Kuzmin, J. Vilo, H. Peterson, gprofiler2 -- an R package for gene list functional enrichment analysis and namespace conversion toolset g:Profiler. F1000Res. 9, 709 (2020).

35. D. J. B. Clarke, G. B. Marino, E. Z. Deng, Z. Xie, J. E. Evangelista, A. Ma’ayan, Rummagene: massive mining of gene sets from supporting materials of biomedical research publications. Commun. Biol. 7, 482 (2024).

36. G. B. Marino, D. J. B. Clarke, A. Lachmann, E. Z. Deng, A. Ma’ayan, RummaGEO: Automatic mining of human and mouse gene sets from GEO. Patterns (N. Y.) 5, 101072 (2024).

37. A. Alexa, J. Rahnenfuhrer, TopGO: Enrichment Analysis for Gene Ontology (2019).

38. R. Batra, M. Arnold, M. A. Wörheide, M. Allen, X. Wang, C. Blach, A. I. Levey, N. T. Seyfried, N. Ertekin-Taner, D. A. Bennett, G. Kastenmüller, R. F. Kaddurah-Daouk, J. Krumsiek, Alzheimer’s Disease Metabolomics Consortium (ADMC), The landscape of metabolic brain alterations in Alzheimer’s disease. Alzheimers. Dement., 1–19 (2022).

39. Y.-W. Wan, R. Al-Ouran, C. G. Mangleburg, T. M. Perumal, T. V. Lee, K. Allison, V. Swarup, C. C. Funk, C. Gaiteri, M. Allen, M. Wang, S. M. Neuner, C. C. Kaczorowski, V. M. Philip, G. R. Howell, H. Martini-Stoica, H. Zheng, H. Mei, X. Zhong, J. W. Kim, V. L. Dawson, T. M. Dawson, P.-C. Pao, L.-H. Tsai, J.-V. Haure-Mirande, M. E. Ehrlich, P. Chakrabarty, Y. Levites, X. Wang, E. B. Dammer, G. Srivastava, S. Mukherjee, S. K. Sieberts, L. Omberg, K. D. Dang, J. A. Eddy, P. Snyder, Y. Chae, S. Amberkar, W. Wei, W. Hide, C. Preuss, A. Ergun, P. J. Ebert, D. C. Airey, S. Mostafavi, L. Yu, H.-U. Klein, Accelerating Medicines Partnership-Alzheimer’s Disease Consortium, G. W. Carter, D. A. Collier, T. E. Golde, A. I. Levey, D. A. Bennett, K. Estrada, T. M. Townsend, B. Zhang, E. Schadt, P. L. De Jager, N. D. Price, N. Ertekin-Taner, Z. Liu, J. M. Shulman, L. M. Mangravite, B. A. Logsdon, Meta-Analysis of the Alzheimer’s Disease Human Brain Transcriptome and Functional Dissection in Mouse Models. Cell Rep. 32, 107908 (2020).

40. E. C. B. Johnson, E. B. Dammer, D. M. Duong, L. Ping, M. Zhou, L. Yin, L. A. Higginbotham, A. Guajardo, B. White, J. C. Troncoso, M. Thambisetty, T. J. Montine, E. B. Lee, J. Q. Trojanowski, T. G. Beach, E. M. Reiman, V. Haroutunian, M. Wang, E. Schadt, B. Zhang, D. W. Dickson, N. Ertekin-Taner, T. E. Golde, V. A. Petyuk, P. L. De Jager, D. A. Bennett, T. S. Wingo, S. Rangaraju, I. Hajjar, J. M. Shulman, J. J. Lah, A. I. Levey, N. T. Seyfried, Large-scale proteomic analysis of Alzheimer’s disease brain and cerebrospinal fluid reveals early changes in energy metabolism associated with microglia and astrocyte activation. Nat. Med. 26, 769–780 (2020).

41. E. C. B. Johnson, E. K. Carter, E. B. Dammer, D. M. Duong, E. S. Gerasimov, Y. Liu, J. Liu, R. Betarbet, L. Ping, L. Yin, G. E. Serrano, T. G. Beach, J. Peng, P. L. De Jager, V. Haroutunian, B. Zhang, C. Gaiteri, D. A. Bennett, M. Gearing, T. S. Wingo, A. P. Wingo, J. J. Lah, A. I. Levey, N. T. Seyfried, Large-scale deep multi-layer analysis of Alzheimer’s disease brain reveals strong proteomic disease-related changes not observed at the RNA level. Nat. Neurosci. 25, 213–225 (2022).

42. J. C. Lambert, C. A. Ibrahim-Verbaas, D. Harold, A. C. Naj, R. Sims, C. Bellenguez, G. Jun, A. L. DeStefano, J. C. Bis, G. W. Beecham, B. Grenier-Boley, G. Russo, T. A. Thornton-Wells, N. Jones, A. V. Smith, V. Chouraki, C. Thomas, M. A. Ikram, D. Zelenika, B. N. Vardarajan, Y. Kamatani, C. F. Lin, A. Gerrish, H. Schmidt, B. Kunkle, N. Fiévet, P. Amouyel, F. Pasquier, V. Deramecourt, R. F. A. G. De Bruijn, N. Amin, A. Hofman, C. M. Van Duijn, M. L. Dunstan, P. Hollingworth, M. J. Owen, M. C. O’Donovan, L. Jones, P. A. Holmans, V. Moskvina, J. Williams, C. Baldwin, L. A. Farrer, S. H. Choi, K. L. Lunetta, A. L. Fitzpatrick, T. B. Harris, B. M. Psaty, J. R. Gilbert, K. L. Hamilton-Nelson, E. R. Martin, M. A. Pericak-Vance, J. L. Haines, V. Gudnason, P. V. Jonsson, G. Eiriksdottir, M. T. Bihoreau, M. Lathrop, O. Valladares, L. B. Cantwell, L. S. Wang, G. D. Schellenberg, A. Ruiz, M. Boada, C. Reitz, R. Mayeux, A. Ramirez, W. Maier, O. Hanon, W. A. Kukull, J. D. Buxbaum, D. Campion, D. Wallon, D. Hannequin, P. K. Crane, E. B. Larson, T. Becker, C. Cruchaga, A. M. Goate, D. Craig, J. A. Johnston, B. Mc-Guinness, S. Todd, P. Passmore, C. Berr, K. Ritchie, O. L. Lopez, P. L. De Jager, D. Evans, S. Lovestone, P. Proitsi, J. F. Powell, L. Letenneur, P. Barberger-Gateau, C. Dufouil, J. F. Dartigues, F. J. Morón, D. C. Rubinsztein, P. St. George-Hyslop, K. Sleegers, K. Bettens, C. Van Broeckhoven, M. J. Huentelman, M. Gill, K. Brown, K. Morgan, M. I. Kamboh, L. Keller, L. Fratiglioni, R. Green, A. J. Myers, S. Love, E. Rogaeva, J. Gallacher, A. Bayer, J. Clarimon, A. Lleo, D. W. Tsuang, L. Yu, D. A. Bennett, M. Tsolaki, P. Bossù, G. Spalletta, J. Collinge, S. Mead, S. Sorbi, B. Nacmias, F. Sanchez-Garcia, M. C. Deniz Naranjo, N. C. Fox, J. Hardy, P. Bosco, R. Clarke, C. Brayne, D. Galimberti, M. Mancuso, F. Matthews, S. Moebus, P. Mecocci, M. Del Zompo, H. Hampel, A. Pilotto, M. Bullido, F. Panza, P. Caffarra, M. Mayhaus, S. Pichler, W. Gu, M. Riemenschneider, L. Lannfelt, M. Ingelsson, H. Hakonarson, M. M. Carrasquillo, F. Zou, S. G. Younkin, D. Beekly, V. Alvarez, E. Coto, C. Razquin, P. Pastor, I. Mateo, O. Combarros, K. M. Faber, T. M. Foroud, H. Soininen, M. Hiltunen, D. Blacker, T. H. Mosley, C. Graff, C. Holmes, T. J. Montine, J. I. Rotter, A. Brice, M. A. Nalls, J. S. K. Kauwe, E. Boerwinkle, R. Schmidt, D. Rujescu, C. Tzourio, M. M. Nöthen, L. J. Launer, S. Seshadri, Meta-analysis of 74,046 individuals identifies 11 new susceptibility loci for Alzheimer’s disease. Nat. Genet. 45, 1452–1458 (2013).

43. G. W. Beecham, K. Hamilton, A. C. Naj, E. R. Martin, M. Huentelman, A. J. Myers, J. J. Corneveaux, J. Hardy, J.-P. Vonsattel, S. G. Younkin, D. A. Bennett, P. L. De Jager, E. B. Larson, P. K. Crane, M. I. Kamboh, J. K. Kofler, D. C. Mash, L. Duque, J. R. Gilbert, H. Gwirtsman, J. D. Buxbaum, P. Kramer, D. W. Dickson, L. A. Farrer, M. P. Frosch, B. Ghetti, J. L. Haines, B. T. Hyman, W. A. Kukull, R. P. Mayeux, M. A. Pericak-Vance, J. A. Schneider, J. Q. Trojanowski, E. M. Reiman, Alzheimer’s Disease Genetics Consortium (ADGC), G. D. Schellenberg, T. J. Montine, Genome-wide association meta-analysis of neuropathologic features of Alzheimer’s disease and related dementias. PLoS Genet. 10, e1004606 (2014).

44. Y. Deming, J. Xia, Y. Cai, J. Lord, P. Holmans, S. Bertelsen, D. Holtzman, J. C. Morris, K. Bales, E. H. Pickering, J. Kauwe, A. Goate, C. Cruchaga, A potential endophenotype for Alzheimer’s disease: cerebrospinal fluid clusterin. Neurobiol. Aging 37, 208.e1–208.e9 (2016).

45. Y. Deming, Z. Li, M. Kapoor, O. Harari, J. L. Del-Aguila, K. Black, D. Carrell, Y. Cai, M. V. Fernandez, J. Budde, S. Ma, B. Saef, B. Howells, K.-L. Huang, S. Bertelsen, A. M. Fagan, D. M. Holtzman, J. C. Morris, S. Kim, A. J. Saykin, P. L. De Jager, M. Albert, A. Moghekar, R. O’Brien, M. Riemenschneider, R. C. Petersen, K. Blennow, H. Zetterberg, L. Minthon, V. M. Van Deerlin, V. M.-Y. Lee, L. M. Shaw, J. Q. Trojanowski, G. Schellenberg, J. L. Haines, R. Mayeux, M. A. Pericak-Vance, L. A. Farrer, E. R. Peskind, G. Li, A. F. Di Narzo, Alzheimer’s Disease Neuroimaging Initiative (ADNI), Alzheimer Disease Genetic Consortium (ADGC), J. S. K. Kauwe, A. M. Goate, C. Cruchaga, Genome-wide association study identifies four novel loci associated with Alzheimer’s endophenotypes and disease modifiers. Acta Neuropathol. 133, 839–856 (2017).

46. K.-L. Huang, E. Marcora, A. A. Pimenova, A. F. Di Narzo, M. Kapoor, S. C. Jin, O. Harari, S. Bertelsen, B. P. Fairfax, J. Czajkowski, V. Chouraki, B. Grenier-Boley, C. Bellenguez, Y. Deming, A. McKenzie, T. Raj, A. E. Renton, J. Budde, A. Smith, A. Fitzpatrick, J. C. Bis, A. DeStefano, H. H. H. Adams, M. A. Ikram, S. van der Lee, J. L. Del-Aguila, M. V. Fernandez, L. Ibañez, International Genomics of Alzheimer’s Project, Alzheimer’s Disease Neuroimaging Initiative, R. Sims, V. Escott-Price, R. Mayeux, J. L. Haines, L. A. Farrer, M. A. Pericak-Vance, J. C. Lambert, C. van Duijn, L. Launer, S. Seshadri, J. Williams, P. Amouyel, G. D. Schellenberg, B. Zhang, I. Borecki, J. S. K. Kauwe, C. Cruchaga, K. Hao, A. M. Goate, A common haplotype lowers PU.1 expression in myeloid cells and delays onset of Alzheimer’s disease. Nat. Neurosci. 20, 1052–1061 (2017).

47. R. E. Marioni, S. E. Harris, Q. Zhang, A. F. McRae, S. P. Hagenaars, W. D. Hill, G. Davies, C. W. Ritchie, C. R. Gale, J. M. Starr, A. M. Goate, D. J. Porteous, J. Yang, K. L. Evans, I. J. Deary, N. R. Wray, P. M. Visscher, GWAS on family history of Alzheimer’s disease. Transl. Psychiatry 8, 0–6 (2018).

48. I. E. Jansen, J. E. Savage, K. Watanabe, J. Bryois, D. M. Williams, S. Steinberg, J. Sealock, I. K. Karlsson, S. Hägg, L. Athanasiu, N. Voyle, P. Proitsi, A. Witoelar, S. Stringer, D. Aarsland, I. S. Almdahl, F. Andersen, S. Bergh, F. Bettella, S. Bjornsson, A. Brækhus, G. Bråthen, C. de Leeuw, R. S. Desikan, S. Djurovic, L. Dumitrescu, T. Fladby, T. J. Hohman, P. V. Jonsson, S. J. Kiddle, A. Rongve, I. Saltvedt, S. B. Sando, G. Selbæk, M. Shoai, N. G. Skene, J. Snaedal, E. Stordal, I. D. Ulstein, Y. Wang, L. R. White, J. Hardy, J. Hjerling-Leffler, P. F. Sullivan, W. M. van der Flier, R. Dobson, L. K. Davis, H. Stefansson, K. Stefansson, N. L. Pedersen, S. Ripke, O. A. Andreassen, D. Posthuma, Genome-wide meta-analysis identifies new loci and functional pathways influencing Alzheimer’s disease risk. Nat. Genet. 51, 404–413 (2019).

49. B. W. Kunkle, B. Grenier-Boley, R. Sims, J. C. Bis, V. Damotte, A. C. Naj, A. Boland, M. Vronskaya, S. J. van der Lee, A. Amlie-Wolf, C. Bellenguez, A. Frizatti, V. Chouraki, E. R. Martin, K. Sleegers, N. Badarinarayan, J. Jakobsdottir, K. L. Hamilton-Nelson, S. Moreno-Grau, R. Olaso, R. Raybould, Y. Chen, A. B. Kuzma, M. Hiltunen, T. Morgan, S. Ahmad, B. N. Vardarajan, J. Epelbaum, P. Hoffmann, M. Boada, G. W. Beecham, J. G. Garnier, D. Harold, A. L. Fitzpatrick, O. Valladares, M. L. Moutet, A. Gerrish, A. V. Smith, L. Qu, D. Bacq, N. Denning, X. Jian, Y. Zhao, M. Del Zompo, N. C. Fox, S. H. Choi, I. Mateo, J. T. Hughes, H. H. Adams, J. Malamon, F. Sanchez-Garcia, Y. Patel, J. A. Brody, B. A. Dombroski, M. C. D. Naranjo, M. Daniilidou, G. Eiriksdottir, S. Mukherjee, D. Wallon, J. Uphill, T. Aspelund, L. B. Cantwell, F. Garzia, D. Galimberti, E. Hofer, M. Butkiewicz, B. Fin, E. Scarpini, C. Sarnowski, W. S. Bush, S. Meslage, J. Kornhuber, C. C. White, Y. Song, R. C. Barber, S. Engelborghs, S. Sordon, D. Voijnovic, P. M. Adams, R. Vandenberghe, M. Mayhaus, L. A. Cupples, M. S. Albert, P. P. De Deyn, W. Gu, J. J. Himali, D. Beekly, A. Squassina, A. M. Hartmann, A. Orellana, D. Blacker, E. Rodriguez-Rodriguez, S. Lovestone, M. E. Garcia, R. S. Doody, C. Munoz-Fernadez, R. Sussams, H. Lin, T. J. Fairchild, Y. A. Benito, C. Holmes, H. Karamujić-Čomić, M. P. Frosch, H. Thonberg, W. Maier, G. Roschupkin, B. Ghetti, V. Giedraitis, A. Kawalia, S. Li, R. M. Huebinger, L. Kilander, S. Moebus, I. Hernández, M. I. Kamboh, R. M. Brundin, J. Turton, Q. Yang, M. J. Katz, L. Concari, J. Lord, A. S. Beiser, C. D. Keene, S. Helisalmi, I. Kloszewska, W. A. Kukull, A. M. Koivisto, A. Lynch, L. Tarraga, E. B. Larson, A. Haapasalo, B. Lawlor, T. H. Mosley, R. B. Lipton, V. Solfrizzi, M. Gill, W. T. Longstreth, T. J. Montine, V. Frisardi, M. Diez-Fairen, F. Rivadeneira, R. C. Petersen, V. Deramecourt, I. Alvarez, F. Salani, A. Ciaramella, E. Boerwinkle, E. M. Reiman, N. Fievet, J. I. Rotter, J. S. Reisch, O. Hanon, C. Cupidi, A. G. Andre Uitterlinden, D. R. Royall, C. Dufouil, R. G. Maletta, I. de Rojas, M. Sano, A. Brice, R. Cecchetti, P. S. George-Hyslop, K. Ritchie, M. Tsolaki, D. W. Tsuang, B. Dubois, D. Craig, C. K. Wu, H. Soininen, D. Avramidou, R. L. Albin, L. Fratiglioni, A. Germanou, L. G. Apostolova, L. Keller, M. Koutroumani, S. E. Arnold, F. Panza, O. Gkatzima, S. Asthana, D. Hannequin, P. Whitehead, C. S. Atwood, P. Caffarra, H. Hampel, I. Quintela, Á. Carracedo, L. Lannfelt, D. C. Rubinsztein, L. L. Barnes, F. Pasquier, L. Frölich, S. Barral, B. McGuinness, T. G. Beach, J. A. Johnston, J. T. Becker, P. Passmore, E. H. Bigio, J. M. Schott, T. D. Bird, J. D. Warren, B. F. Boeve, M. K. Lupton, J. D. Bowen, P. Proitsi, A. Boxer, J. F. Powell, J. R. Burke, J. S. K. Kauwe, J. M. Burns, M. Mancuso, J. D. Buxbaum, U. Bonuccelli, N. J. Cairns, A. McQuillin, C. Cao, G. Livingston, C. S. Carlson, N. J. Bass, C. M. Carlsson, J. Hardy, R. M. Carney, J. Bras, M. M. Carrasquillo, R. Guerreiro, M. Allen, H. C. Chui, E. Fisher, C. Masullo, E. A. Crocco, C. DeCarli, G. Bisceglio, M. Dick, L. Ma, R. Duara, N. R. Graff-Radford, D. A. Evans, A. Hodges, K. M. Faber, M. Scherer, K. B. Fallon, M. Riemenschneider, D. W. Fardo, R. Heun, M. R. Farlow, H. Kölsch, S. Ferris, M. Leber, T. M. Foroud, I. Heuser, D. R. Galasko, I. Giegling, M. Gearing, M. Hüll, D. H. Geschwind, J. R. Gilbert, J. Morris, R. C. Green, K. Mayo, J. H. Growdon, T. Feulner, R. L. Hamilton, L. E. Harrell, D. Drichel, L. S. Honig, T. D. Cushion, M. J. Huentelman, P. Hollingworth, C. M. Hulette, B. T. Hyman, R. Marshall, G. P. Jarvik, A. Meggy, E. Abner, G. E. Menzies, L. W. Jin, G. Leonenko, L. M. Real, G. R. Jun, C. T. Baldwin, D. Grozeva, A. Karydas, G. Russo, J. A. Kaye, R. Kim, F. Jessen, N. W. Kowall, B. Vellas, J. H. Kramer, E. Vardy, F. M. LaFerla, K. H. Jöckel, J. J. Lah, M. Dichgans, J. B. Leverenz, D. Mann, A. I. Levey, S. Pickering-Brown, A. P. Lieberman, N. Klopp, K. L. Lunetta, H. E. Wichmann, C. G. Lyketsos, K. Morgan, D. C. Marson, K. Brown, F. Martiniuk, C. Medway, D. C. Mash, M. M. Nöthen, E. Masliah, N. M. Hooper, W. C. McCormick, A. Daniele, S. M. McCurry, A. Bayer, A. N. McDavid, J. Gallacher, A. C. McKee, H. van den Bussche, M. Mesulam, C. Brayne, B. L. Miller, S. Riedel-Heller, C. A. Miller, J. W. Miller, A. Al-Chalabi, J. C. Morris, C. E. Shaw, A. J. Myers, J. Wiltfang, S. O’Bryant, J. M. Olichney, V. Alvarez, J. E. Parisi, A. B. Singleton, H. L. Paulson, J. Collinge, W. R. Perry, S. Mead, E. Peskind, D. H. Cribbs, M. Rossor, A. Pierce, N. S. Ryan, W. W. Poon, B. Nacmias, H. Potter, S. Sorbi, J. F. Quinn, E. Sacchinelli, A. Raj, G. Spalletta, M. Raskind, C. Caltagirone, P. Bossù, M. D. Orfei, B. Reisberg, R. Clarke, C. Reitz, A. D. Smith, J. M. Ringman, D. Warden, E. D. Roberson, G. Wilcock, E. Rogaeva, A. C. Bruni, H. J. Rosen, M. Gallo, R. N. Rosenberg, Y. Ben-Shlomo, M. A. Sager, P. Mecocci, A. J. Saykin, P. Pastor, M. L. Cuccaro, J. M. Vance, J. A. Schneider, L. S. Schneider, S. Slifer, W. W. Seeley, A. G. Smith, J. A. Sonnen, S. Spina, R. A. Stern, R. H. Swerdlow, M. Tang, R. E. Tanzi, J. Q. Trojanowski, J. C. Troncoso, V. M. Van Deerlin, L. J. Van Eldik, H. V. Vinters, J. P. Vonsattel, S. Weintraub, K. A. Welsh-Bohmer, K. C. Wilhelmsen, J. Williamson, T. S. Wingo, R. L. Woltjer, C. B. Wright, C. E. Yu, L. Yu, Y. Saba, A. Pilotto, M. J. Bullido, O. Peters, P. K. Crane, D. Bennett, P. Bosco, E. Coto, V. Boccardi, P. L. De Jager, A. Lleo, N. Warner, O. L. Lopez, M. Ingelsson, P. Deloukas, C. Cruchaga, C. Graff, R. Gwilliam, M. Fornage, A. M. Goate, P. Sanchez-Juan, P. G. Kehoe, N. Amin, N. Ertekin-Taner, C. Berr, S. Debette, S. Love, L. J. Launer, S. G. Younkin, J. F. Dartigues, C. Corcoran, M. A. Ikram, D. W. Dickson, G. Nicolas, D. Campion, J. A. Tschanz, H. Schmidt, H. Hakonarson, J. Clarimon, R. Munger, R. Schmidt, L. A. Farrer, C. Van Broeckhoven, M. C. O’Donovan, A. L. DeStefano, L. Jones, J. L. Haines, J. F. Deleuze, M. J. Owen, V. Gudnason, R. Mayeux, V. Escott-Price, B. M. Psaty, A. Ramirez, L. S. Wang, A. Ruiz, C. M. van Duijn, P. A. Holmans, S. Seshadri, J. Williams, P. Amouyel, G. D. Schellenberg, J. C. Lambert, M. A. Pericak-Vance, Genetic meta-analysis of diagnosed Alzheimer’s disease identifies new risk loci and implicates Aβ, tau, immunity and lipid processing. Nat. Genet. 51, 414–430 (2019).

50. D. P. Wightman, I. E. Jansen, J. E. Savage, A. A. Shadrin, S. Bahrami, D. Holland, A. Rongve, S. Børte, B. S. Winsvold, O. K. Drange, A. E. Martinsen, A. H. Skogholt, C. Willer, G. Bråthen, I. Bosnes, J. B. Nielsen, L. G. Fritsche, L. F. Thomas, L. M. Pedersen, M. E. Gabrielsen, M. B. Johnsen, T. W. Meisingset, W. Zhou, P. Proitsi, A. Hodges, R. Dobson, L. Velayudhan, K. Heilbron, A. Auton, 23andMe Research Team, J. M. Sealock, L. K. Davis, N. L. Pedersen, C. A. Reynolds, I. K. Karlsson, S. Magnusson, H. Stefansson, S. Thordardottir, P. V. Jonsson, J. Snaedal, A. Zettergren, I. Skoog, S. Kern, M. Waern, H. Zetterberg, K. Blennow, E. Stordal, K. Hveem, J.-A. Zwart, L. Athanasiu, P. Selnes, I. Saltvedt, S. B. Sando, I. Ulstein, S. Djurovic, T. Fladby, D. Aarsland, G. Selbæk, S. Ripke, K. Stefansson, O. A. Andreassen, D. Posthuma, A genome-wide association study with 1,126,563 individuals identifies new risk loci for Alzheimer’s disease. Nat. Genet. 53, 1276–1282 (2021).

51. C. Bellenguez, F. Küçükali, I. E. Jansen, L. Kleineidam, S. Moreno-Grau, N. Amin, A. C. Naj, R. Campos-Martin, B. Grenier-Boley, V. Andrade, P. A. Holmans, A. Boland, V. Damotte, S. J. van der Lee, M. R. Costa, T. Kuulasmaa, Q. Yang, I. de Rojas, J. C. Bis, A. Yaqub, I. Prokic, J. Chapuis, S. Ahmad, V. Giedraitis, D. Aarsland, P. Garcia-Gonzalez, C. Abdelnour, E. Alarcón-Martín, D. Alcolea, M. Alegret, I. Alvarez, V. Álvarez, N. J. Armstrong, A. Tsolaki, C. Antúnez, I. Appollonio, M. Arcaro, S. Archetti, A. A. Pastor, B. Arosio, L. Athanasiu, H. Bailly, N. Banaj, M. Baquero, S. Barral, A. Beiser, A. B. Pastor, J. E. Below, P. Benchek, L. Benussi, C. Berr, C. Besse, V. Bessi, G. Binetti, A. Bizarro, R. Blesa, M. Boada, E. Boerwinkle, B. Borroni, S. Boschi, P. Bossù, G. Bråthen, J. Bressler, C. Bresner, H. Brodaty, K. J. Brookes, L. I. Brusco, D. Buiza-Rueda, K. Bûrger, V. Burholt, W. S. Bush, M. Calero, L. B. Cantwell, G. Chene, J. Chung, M. L. Cuccaro, Á. Carracedo, R. Cecchetti, L. Cervera-Carles, C. Charbonnier, H.-H. Chen, C. Chillotti, S. Ciccone, J. A. H. R. Claassen, C. Clark, E. Conti, A. Corma-Gómez, E. Costantini, C. Custodero, D. Daian, M. C. Dalmasso, A. Daniele, E. Dardiotis, J.-F. Dartigues, P. P. de Deyn, K. de Paiva Lopes, L. D. de Witte, S. Debette, J. Deckert, T. Del Ser, N. Denning, A. DeStefano, M. Dichgans, J. Diehl-Schmid, M. Diez-Fairen, P. D. Rossi, S. Djurovic, E. Duron, E. Düzel, C. Dufouil, G. Eiriksdottir, S. Engelborghs, V. Escott-Price, A. Espinosa, M. Ewers, K. M. Faber, T. Fabrizio, S. F. Nielsen, D. W. Fardo, L. Farotti, C. Fenoglio, M. Fernández-Fuertes, R. Ferrari, C. B. Ferreira, E. Ferri, B. Fin, P. Fischer, T. Fladby, K. Fließbach, B. Fongang, M. Fornage, J. Fortea, T. M. Foroud, S. Fostinelli, N. C. Fox, E. Franco-Macías, M. J. Bullido, A. Frank-García, L. Froelich, B. Fulton-Howard, D. Galimberti, J. M. García-Alberca, P. García-González, S. Garcia-Madrona, G. Garcia-Ribas, R. Ghidoni, I. Giegling, G. Giorgio, A. M. Goate, O. Goldhardt, D. Gomez-Fonseca, A. González-Pérez, C. Graff, G. Grande, E. Green, T. Grimmer, E. Grünblatt, M. Grunin, V. Gudnason, T. Guetta-Baranes, A. Haapasalo, G. Hadjigeorgiou, J. L. Haines, K. L. Hamilton-Nelson, H. Hampel, O. Hanon, J. Hardy, A. M. Hartmann, L. Hausner, J. Harwood, S. Heilmann-Heimbach, S. Helisalmi, M. T. Heneka, I. Hernández, M. J. Herrmann, P. Hoffmann, C. Holmes, H. Holstege, R. H. Vilas, M. Hulsman, J. Humphrey, G. J. Biessels, X. Jian, C. Johansson, G. R. Jun, Y. Kastumata, J. Kauwe, P. G. Kehoe, L. Kilander, A. K. Ståhlbom, M. Kivipelto, A. Koivisto, J. Kornhuber, M. H. Kosmidis, W. A. Kukull, P. P. Kuksa, B. W. Kunkle, A. B. Kuzma, C. Lage, E. J. Laukka, L. Launer, A. Lauria, C.-Y. Lee, J. Lehtisalo, O. Lerch, A. Lleó, W. Longstreth Jr, O. Lopez, A. L. de Munain, S. Love, M. Löwemark, L. Luckcuck, K. L. Lunetta, Y. Ma, J. Macías, C. A. MacLeod, W. Maier, F. Mangialasche, M. Spallazzi, M. Marquié, R. Marshall, E. R. Martin, A. M. Montes, C. M. Rodríguez, C. Masullo, R. Mayeux, S. Mead, P. Mecocci, M. Medina, A. Meggy, S. Mehrabian, S. Mendoza, M. Menéndez-González, P. Mir, S. Moebus, M. Mol, L. Molina-Porcel, L. Montrreal, L. Morelli, F. Moreno, K. Morgan, T. Mosley, M. M. Nöthen, C. Muchnik, S. Mukherjee, B. Nacmias, T. Ngandu, G. Nicolas, B. G. Nordestgaard, R. Olaso, A. Orellana, M. Orsini, G. Ortega, A. Padovani, C. Paolo, G. Papenberg, L. Parnetti, F. Pasquier, P. Pastor, G. Peloso, A. Pérez-Cordón, J. Pérez-Tur, P. Pericard, O. Peters, Y. A. L. Pijnenburg, J. A. Pineda, G. Piñol-Ripoll, C. Pisanu, T. Polak, J. Popp, D. Posthuma, J. Priller, R. Puerta, O. Quenez, I. Quintela, J. Q. Thomassen, A. Rábano, I. Rainero, F. Rajabli, I. Ramakers, L. M. Real, M. J. T. Reinders, C. Reitz, D. Reyes-Dumeyer, P. Ridge, S. Riedel-Heller, P. Riederer, N. Roberto, E. Rodriguez-Rodriguez, A. Rongve, I. R. Allende, M. Rosende-Roca, J. L. Royo, E. Rubino, D. Rujescu, M. E. Sáez, P. Sakka, I. Saltvedt, Á. Sanabria, M. B. Sánchez-Arjona, F. Sanchez-Garcia, P. S. Juan, R. Sánchez-Valle, S. B. Sando, C. Sarnowski, C. L. Satizabal, M. Scamosci, N. Scarmeas, E. Scarpini, P. Scheltens, N. Scherbaum, M. Scherer, M. Schmid, A. Schneider, J. M. Schott, G. Selbæk, D. Seripa, M. Serrano, J. Sha, A. A. Shadrin, O. Skrobot, S. Slifer, G. J. L. Snijders, H. Soininen, V. Solfrizzi, A. Solomon, Y. Song, S. Sorbi, O. Sotolongo-Grau, G. Spalletta, A. Spottke, A. Squassina, E. Stordal, J. P. Tartan, L. Tárraga, N. Tesí, A. Thalamuthu, T. Thomas, G. Tosto, L. Traykov, L. Tremolizzo, A. Tybjærg-Hansen, A. Uitterlinden, A. Ullgren, I. Ulstein, S. Valero, O. Valladares, C. Van Broeckhoven, J. Vance, B. N. Vardarajan, A. van der Lugt, J. Van Dongen, J. van Rooij, J. van Swieten, R. Vandenberghe, F. Verhey, J.-S. Vidal, J. Vogelgsang, M. Vyhnalek, M. Wagner, D. Wallon, L.-S. Wang, R. Wang, L. Weinhold, J. Wiltfang, G. Windle, B. Woods, M. Yannakoulia, H. Zare, Y. Zhao, X. Zhang, C. Zhu, M. Zulaica, EADB, GR@ACE, DEGESCO, EADI, GERAD, Demgene, FinnGen, ADGC, CHARGE, L. A. Farrer, B. M. Psaty, M. Ghanbari, T. Raj, P. Sachdev, K. Mather, F. Jessen, M. A. Ikram, A. de Mendonça, J. Hort, M. Tsolaki, M. A. Pericak-Vance, P. Amouyel, J. Williams, R. Frikke-Schmidt, J. Clarimon, J.-F. Deleuze, G. Rossi, S. Seshadri, O. A. Andreassen, M. Ingelsson, M. Hiltunen, K. Sleegers, G. D. Schellenberg, C. M. van Duijn, R. Sims, W. M. van der Flier, A. Ruiz, A. Ramirez, J.-C. Lambert, New insights into the genetic etiology of Alzheimer’s disease and related dementias. Nat. Genet. 54, 412–436 (2022).

52. K. Suhre, S.-Y. Shin, A.-K. Petersen, R. P. Mohney, D. Meredith, B. Wägele, E. Altmaier, CARDIoGRAM, P. Deloukas, J. Erdmann, E. Grundberg, C. J. Hammond, M. H. de Angelis, G. Kastenmüller, A. Köttgen, F. Kronenberg, M. Mangino, C. Meisinger, T. Meitinger, H.-W. Mewes, M. V. Milburn, C. Prehn, J. Raffler, J. S. Ried, W. Römisch-Margl, N. J. Samani, K. S. Small, H.-E. Wichmann, G. Zhai, T. Illig, T. D. Spector, J. Adamski, N. Soranzo, C. Gieger, Human metabolic individuality in biomedical and pharmaceutical research. Nature 477, 54–60 (2011).

53. S.-Y. Shin, E. B. Fauman, A.-K. Petersen, J. Krumsiek, R. Santos, J. Huang, M. Arnold, I. Erte, V. Forgetta, T.-P. Yang, K. Walter, C. Menni, L. Chen, L. Vasquez, A. M. Valdes, C. L. Hyde, V. Wang, D. Ziemek, P. Roberts, L. Xi, E. Grundberg, Multiple Tissue Human Expression Resource (MuTHER) Consortium, M. Waldenberger, J. B. Richards, R. P. Mohney, M. V. Milburn, S. L. John, J. Trimmer, F. J. Theis, J. P. Overington, K. Suhre, M. J. Brosnan, C. Gieger, G. Kastenmüller, T. D. Spector, N. Soranzo, An atlas of genetic influences on human blood metabolites. Nat. Genet. 46, 543–550 (2014).

54. J. Raffler, N. Friedrich, M. Arnold, T. Kacprowski, R. Rueedi, E. Altmaier, S. Bergmann, K. Budde, C. Gieger, G. Homuth, M. Pietzner, W. Römisch-Margl, K. Strauch, H. Völzke, M. Waldenberger, H. Wallaschofski, M. Nauck, U. Völker, G. Kastenmüller, K. Suhre, Genome-Wide Association Study with Targeted and Non-targeted NMR Metabolomics Identifies 15 Novel Loci of Urinary Human Metabolic Individuality. PLoS Genet. 11 (2015).

55. H. H. M. Draisma, R. Pool, M. Kobl, R. Jansen, A.-K. Petersen, A. A. M. Vaarhorst, I. Yet, T. Haller, A. Demirkan, T. Esko, G. Zhu, S. Böhringer, M. Beekman, J. B. van Klinken, W. Römisch-Margl, C. Prehn, J. Adamski, A. J. M. de Craen, E. M. van Leeuwen, N. Amin, H. Dharuri, H.-J. Westra, L. Franke, E. J. C. de Geus, J. J. Hottenga, G. Willemsen, A. K. Henders, G. W. Montgomery, D. R. Nyholt, J. B. Whitfield, B. W. Penninx, T. D. Spector, A. Metspalu, P. E. Slagboom, K. W. van Dijk, P. A. C. ’t Hoen, K. Strauch, N. G. Martin, G.-J. B. van Ommen, T. Illig, J. T. Bell, M. Mangino, K. Suhre, M. I. McCarthy, C. Gieger, A. Isaacs, C. M. van Duijn, D. I. Boomsma, Genome-wide association study identifies novel genetic variants contributing to variation in blood metabolite levels. Nat. Commun. 6, 7208 (2015).

56. T. Long, M. Hicks, H.-C. Yu, W. H. Biggs, E. F. Kirkness, C. Menni, J. Zierer, K. S. Small, M. Mangino, H. Messier, S. Brewerton, Y. Turpaz, B. A. Perkins, A. M. Evans, L. A. D. Miller, L. Guo, C. T. Caskey, N. J. Schork, C. Garner, T. D. Spector, J. C. Venter, A. Telenti, Whole-genome sequencing identifies common-to-rare variants associated with human blood metabolites. Nat. Genet. 49, 568–578 (2017).

57. S. MahmoudianDehkordi, M. Arnold, K. Nho, S. Ahmad, W. Jia, G. Xie, G. Louie, A. Kueider-Paisley, M. A. Moseley, J. W. Thompson, L. St John Williams, J. D. Tenenbaum, C. Blach, R. Baillie, X. Han, S. Bhattacharyya, J. B. Toledo, S. Schafferer, S. Klein, T. Koal, S. L. Risacher, M. A. Kling, A. Motsinger-Reif, D. M. Rotroff, J. Jack, T. Hankemeier, D. A. Bennett, P. L. De Jager, J. Q. Trojanowski, L. M. Shaw, M. W. Weiner, P. M. Doraiswamy, C. M. van Duijn, A. J. Saykin, G. Kastenmüller, R. Kaddurah-Daouk, Altered bile acid profile associates with cognitive impairment in Alzheimer’s disease—An emerging role for gut microbiome. Alzheimer’s and Dementia 15, 76–92 (2019).

58. K. Nho, A. Kueider-Paisley, S. MahmoudianDehkordi, M. Arnold, S. L. Risacher, G. Louie, C. Blach, R. Baillie, X. Han, G. Kastenmüller, W. Jia, G. Xie, S. Ahmad, T. Hankemeier, C. M. van Duijn, J. Q. Trojanowski, L. M. Shaw, M. W. Weiner, P. M. Doraiswamy, A. J. Saykin, R. Kaddurah-Daouk, Altered bile acid profile in mild cognitive impairment and Alzheimer’s disease: Relationship to neuroimaging and CSF biomarkers. Alzheimers. Dement. 15, 232–244 (2019).

59. M. Arnold, K. Nho, A. Kueider-Paisley, T. Massaro, K. Huynh, B. Brauner, S. MahmoudianDehkordi, G. Louie, M. A. Moseley, J. W. Thompson, L. S. John-Williams, J. D. Tenenbaum, C. Blach, R. Chang, R. D. Brinton, R. Baillie, X. Han, J. Q. Trojanowski, L. M. Shaw, R. Martins, M. W. Weiner, E. Trushina, J. B. Toledo, P. J. Meikle, D. A. Bennett, J. Krumsiek, P. M. Doraiswamy, A. J. Saykin, R. Kaddurah-Daouk, G. Kastenmüller, Sex and APOE ε4 genotype modify the Alzheimer’s disease serum metabolome. Nat. Commun. 11, 1148 (2020).

60. J. Krumsiek, K. Suhre, A. M. Evans, M. W. Mitchell, R. P. Mohney, M. V. Milburn, B. Wägele, W. Römisch-Margl, T. Illig, J. Adamski, C. Gieger, F. J. Theis, G. Kastenmüller, Mining the Unknown: A Systems Approach to Metabolite Identification Combining Genetic and Metabolic Information. PLoS Genet. 8 (2012).

