## Supplementary Material for "An Integrated Molecular Atlas of Alzheimer’s Disease"

[†] The AMP-AD Consortium:
The full list of contributing scientists is available at *https://adknowledgeportal.org/AMPADConsortiumMembers*.

[‡] The Alzheimer's Disease Neuroimaging Initiative (ADNI):
Data used in preparation of this article were obtained from the ADNI database (adni.loni.usc.edu). As such, the investigators within the ADNI contributed to the design and implementation of ADNI and/or provided data but did not participate in analysis or writing of this report. A complete listing of ADNI investigators can be found at: *http://adni.loni.usc.edu/wp-content/uploads/how_to_apply/ADNI_Acknowledgement_List.pdf*.

[§] The Alzheimer's Disease Metabolomics Consortium (ADMC):
The full list of contributing scientists is available at *https://sites.duke.edu/adnimetab/team/*.

[*] Corresponding author information:

#### MATERIALS and METHODS

#### 1 NETWORK-BASED MULTI-OMICS INTEGRATION

##### 1.1 Integration strategy

The large-scale integration of heterogenous data is not straightforward and often complicated by data-use restrictions, laborious pre-processing steps, analytical differences and varying data formats and identifiers. Besides synchronous integration strategies, that enable the integration of data in a single analysis step, step-wise strategies allow the integration of heterogenous datasets across different cohorts, studies and analytical techniques (*1*). These types of integration strategies do not require the same set of samples for each omics layer and datasets are often analyzed either separately or in specific combinations prior to integration. For example, in genome-wide association studies with intermediate phenotypes, genotyping data is analyzed in combination with another omics layer, such as proteomics or metabolomics. The resulting summary statistics from different analyses can then be integrated in an additional step. To utilize a maximum of available data, the Alzheimer's Disease (AD) Atlas was built using such a step-wise strategy, in which networks are built by establishing pairwise inter-omics, intra-omics and phenotype-specific links using a) knowledge-based information and b) data-driven information (schematically seen in the overview in **Figure 1A**). The thereby established links between biological entities (e.g., genes and metabolites) are merged in a final step by using overlapping entities to construct a composite network. This comprehensive catalogue of multi-omics relationships and influences on AD is represented by a large, heterogenous network in which nodes represent the individual entities (traits, genes and metabolites) and different types of edges represent different types of relationships that link these. A summary of the network can be seen in **Supplementary Table 1**.

##### 1.2 Inferred relationship types

Step-wise data integration for the AD Atlas was achieved by inferring study- and/or analysis-specific significant edges to build multiple networks, and finally overlaying these networks using overlapping entities to connect multiple layers of omics data. In general, three different types of inferred relationships can be distinguished: inter-omics, intra-omics and phenotype-specific links (**Figure 1A**).

***Intra-omics links*** establish relationships within one omics layer using the intrinsic correlation structure of omics datasets. In the AD Atlas, we have included brain region-specific gene co-expression networks (gene-gene links), and partial correlation-based protein and metabolite co-abundance networks (protein-protein and metabolite-metabolite links, respectively).

***Inter-omics links*** establish links between omics layers by using knowledge-based gene-transcript-protein links from Ensembl as well as overlapping genetic associations from genome-wide association studies with molecular traits. The AD Atlas has further incorporated tissue-specific eQTL information (taken from GTEx (*2*), SNiPA (*3*) and Sieberts et al. (*4*)) as well as metabolite quantitative trait loci (mQTL) data from large population-based and brain-based studies (listed in **Supplementary Table 2**).

***Phenotype specific links*** enable the identification of entities that are associated with Alzheimer's disease specific phenotypes (referred to as traits), including CSF biomarkers and neuroimaging. They are inferred from large-scale case-control genome-wide association studies (GWAS) and metabolome-wide association studies (MWAS), as listed in

**Supplementary Table 2**. This layer of information enables the identification of entities within the network that are associated with AD pathomechanisms.

### 2 DATA

The Alzheimer's Disease (AD) Atlas is an integration framework that interconnects and stores diverse biological domains in accessible network structures. The backbone of this multi-layered network comes from known biological relationships such as gene-transcript-protein relations and functional/pathway annotations (e.g., for proteins or metabolites) available in public databases. Large-scale quantitative data from population-based studies are included to establish data-driven relationships within (e.g., tissue-specific gene expression) and across omics (e.g. eQTLs, pQTLs, or mQTLs) as a reference. To identify entities within this network that are relevant to AD, we extend the framework using large-scale association data for AD yielded in case-control and AD biomarker GWAS, MWAS, data on differentially expressed genes and proteins, and brain region-specific gene and protein co-expression. In the following, the currently integrated datasets are listed and additional information regarding their generation and preprocessing is given.

#### 2.1 Public databases and population-based studies

##### 2.1.1 Ensembl

Ensembl (*5*) is a bioinformatics resource that was built to store, annotate and display genome information. The AD Atlas uses the Ensembl database to establish knowledge-based relationships that link genes, transcripts and proteins. Furthermore, Ensembl identifiers (IDs) are used as the primary and unique ID for genes, transcripts and proteins. Ensembl version 97 was accessed in April 2019 using the R package biomaRt (*6*). Genes are further annotated with approved human gene nomenclature provided by the HUGO Gene Nomenclature Committee (HGNC) (*7*). The complete set of annotations (gene id and gene symbol) was downloaded from ftp://ftp.ebi.ac.uk/pub/databases/genenames/new/tsv/hgnc_complete_set.txt.

##### 2.1.2 SNiPA

The single nucleotide polymorphism (SNP) annotation database SNiPA (*3*) v3.3. was used to project SNPs to genes using multiple layers of information, including the genomic location, quantitative trait loci (QTL), such as eQTL associations (cis- and trans-) and protein quantitative trait loci (pQTL) associations (cis- and trans-), and gene-associated regulated elements (ENCODE (*8*), FANTOM5 (*9*)) (*10*). SNP-to-gene mappings determined by SNiPA were downloaded and included in the AD Atlas. Furthermore, this mapping was used to determine the gene-specific significance threshold described in the main manuscript.

##### 2.1.3 Tissue-specific gene regulation

Genotype-Tissue Expression (GTEx) Project (2020)

Information on genetic loci that effect the expression of protein coding genes was taken from the Genotype-Tissue Expression (GTEx) project (*2*). The version 8 (v8) data release examines n=15,201 RNA-sequencing samples from 49 tissues of 838 postmortem donors with genotype data from whole-genome sequencing (WGS). Donors were primarily (85.3%) European Americans. Significant variant-gene associations (cis- and trans-) based on permutations were downloaded from the GTEx Portal for each tissue (*GTEx_Analysis_v8_eQTL.tar* and *GTEx_Analysis_v8_trans_eGenes_fdr05.txt*). GTEx variant IDs were mapped to rsIDs prior to integration using the provided lookup table for genotyped variants (*GTEx_Analysis_2017-06-*

*05_v8_WholeGenomeSeq_838Indiv_Analysis_Freeze.lookup_table.txt.gz*). Sample sizes for brain tissues with RNA-Seq and genotype data available:

| Tissue Type | Samples with RNA-seq and genotyped |
|---|---|
| Brain - Amygdala | 129 |
| Brain - Anterior cingulate cortex (BA24) | 147 |
| Brain - Caudate (basal ganglia) | 194 |
| Brain - Cerebellar Hemisphere | 175 |
| Brain - Cerebellum | 209 |
| Brain - Cortex | 205 |
| Brain - Frontal Cortex (BA9) | 175 |
| Brain - Hippocampus | 165 |
| Brain - Hypothalamus | 170 |
| Brain - Nucleus accumbens (basal ganglia) | 202 |
| Brain - Putamen (basal ganglia) | 170 |
| Brain - Spinal cord (cervical c-1) | 126 |
| Brain - Substantia nigra | 114 |

Sieberts et al. (2020) – Brain cis-eQTL meta-analysis

Sieberts et al. (*4*) conducted a large-scale analysis of cortical cis-eQTL using n=1433 samples from four cohorts from the AMP-AD Consortium and the CommonMind Consortium. eQTLs were generated separately for each cohort/tissue, adjusting for diagnosis and principal components of ancestry. Then meta-analysis was performed via a fixed-effect model. This included following brain regions: DLPFC from ROS/MAP, MSSM-Penn-Pitt, and HBCC, and TCX from Mayo. Results of the cortical meta-analysis were downloaded from Synapse. A significance cutoff of FDR($P_{\text{meta}}$) ≤ 0.05 was applied and SNP rsIDs and Ensembl IDs (for genes) were used for integration.

##### 2.1.4 Genetic associations with metabolic traits (mGWAS)

Summary statistics from genome-wide association studies (GWAS) with metabolic traits were downloaded via SNiPA (*3*) and the metabolomics GWAS server (*11*, *12*). Results filtered to a default p-value cutoff of $P \leq 1 \times 10^{-4}$ were available, if not stated otherwise. Where possible, IDs for genes and metabolites were mapped to Ensembl or metabolomics platform-specific IDs, respectively. The included studies cover metabolic traits measured in serum, plasma, and urine samples using both targeted and non-targeted metabolomics approaches.

Suhre et al. (2011) - Human metabolic individuality

Suhre et al. (*12*) profiled fasting serum samples from participants of the KORA F4 study (n=1,768) and the TwinsUK study (n=1,052) using the non-targeted metabolomics platform Metabolon. After quality control 276 (KORA) and 258 (TwinsUK) metabolites were used for further analysis. Linear models were fitted for each cohort separately to log-transformed metabolic traits and adjusted for age, gender and family structure. Associations with single metabolites ($P < 0.001$) for both cohorts were downloaded and included in the AD Atlas. SNP rsIDs and Metabolon specific COMP_IDs for metabolites were used for integration.

Shin et al. (2014) - Atlas of genetic influences on blood metabolites

Shin et al. (*11*) investigated how genetic variation influences metabolism using data from n=7,824 individuals from two population-based studies (Kora and TwinsUK). In total, 486 metabolite concentrations profiled in either plasma or serum were present in both datasets after quality control. The authors tested for associations between each SNP and metabolite concentration using linear regression models adjusted for age and sex. Batch effect was only added to the model for the TwinsUK analysis. Both cohorts were analyzed separately, using the software QUICKTEST in KORA and Merlin in TwinsUK. The resulting cohort-level summary statistics were combined using inverse variance meta-analysis based on effect size estimates and standard errors, adjusting for genomic control. Association data for all 486 metabolites were downloaded and SNP rsIDs and Metabolon-specific COMP IDs for metabolites were used for integration.

Raffler et al. (2015) - Loci of urinary human metabolic individuality

Raffler et al. (*10*) performed a GWAS using metabolically characterized urine samples (targeted metabolomics) and genotype data available for n=3,861 study participants from the longitudinal population study SHIP-0. The software PLINK (v1.07) was used to fit age- and sex-corrected linear regression models and a Bonferroni-adjusted significance threshold was applied to correct for multiple testing. The association results for 55 targeted metabolic traits in urine were included in the AD Atlas. SNP rsIDs and biochemical names for metabolites were used for integration.

Draisma et al. (2015) - Genetic variants contributing to variation in blood metabolite levels

Draisma et al. (*13*) performed a meta-analysis of genome-wide association analysis using blood serum samples from n=7,478 individuals across seven cohorts and five countries (the Netherlands, Germany, Australia, Estonia, and the United Kingdom). Genotype data and targeted metabolomic measurements (129 metabolites), performed using the Biocrates platform, were first analyzed assuming a linear model of association, adjusting for age, sex, relatedness, and study-specific covariates as necessary. In the next step, these cohort-level summary statistics were pooled in an inverse variance-weighted, fixed-effects meta-analysis using the software METAL. The association results were downloaded and SNP rsIDs and biochemical names metabolites were used for integration.

Long et al. (2017) - Common-to-rare variants associated with human blood metabolites

Long et al. (*14*) conducted a whole-genome sequencing study of common, low-frequency, and rare variants in n=1,960 participants of the TwinsUK study. Furthermore, the authors profiled serum samples from these participants collected at three clinical visits using the non-targeted metabolomics platform Metabolon. Focusing on 644 metabolites that showed consistent levels across these data collections and measurable heritability, a linear mixed model was applied to test for associations between genetic variants and metabolite levels while accounting for family structure. As a quantitative trait the log10-transformed mean of the median-normalized values from three visits was used. Sex and mean age at serum collection were included as covariates. Summary statistics for each of the 644 metabolites were downloaded ($P \leq 1 \times 10^{-5}$) and included in the AD Atlas. rsIDs and biochemical names for SNPs and metabolites were used for integration, respectively.

##### 2.1.5 Partial correlation networks

Krumsiek et al. (2012) - A systems approach to metabolite identification

Krumsiek et al. (*15*) combined GWAS and Gaussian Graphical Models (GGM) to identify measured metabolites with yet unknown chemical structure. GGMs, which are based on partial correlation coefficients, were applied to a dataset of 517 metabolic traits and genotype information on 655,658 genetic variants measured in n=1,768 fasting serum samples from the German population cohort KORA. Confounding through age, gender, and SNP effects were removed by including these variables in the constructed linear models. Partial correlations between metabolite pairs that are significantly different from zero at α = 0.05 after Bonferroni correction ($P \leq 7.96\times10^{-7}$) and abs(cor) ≥ 0.1603, were downloaded from the supporting information of the paper and included in the AD Atlas using metabolite biochemical names for the integration.

Suhre et al. (2017) - GWAS with the human blood plasma proteome

Suhre et al. (*16*) performed a large-scale proteomics-based genetic association study in the German KORA cohort, identifying associations between genetic variants and protein levels, measured by the aptamer-based proteomics platform SOMAscan. In the scope of the study, a GGM was computed for n=997 blood plasma samples using unscaled data for 1,124 proteins and correcting for age, gender, and body mass index (BMI). Partial correlation edges between proteins were included in the network after applying a significance threshold of α = 0.05 after Bonferroni correction for all possible edges in the model ($P \leq 7.9 \times 10-8$). Supplementary dataset 2 (annotation of the SOMAmer probes) and dataset 3 (GGM edges) were downloaded. Using the annotations supplied, SOMAmer probe identifiers were mapped to Uniprot IDs (in some cases multiple) for subsequent integration into the AD Atlas.

#### 2.2 Alzheimer's disease-related associations

##### 2.2.1 Genetic associations with Alzheimer's (endo-)phenotypes

Summary statistics for the following large-scale genome-wide association studies on CSF biomarkers, including amyloid-β, tau and clusterin, and neuropathological burden, as well as case-control meta-analyses, were integrated into the AD Atlas. Datasets were downloaded from the National Institute on Aging Genetics of Alzheimer's Disease Data Storage Site (NIAGADS; *https://www.niagads.org/*) if not stated otherwise. SNP rsIDs were used as unique identifiers and AD (endo-)phenotypes were manually harmonized.

Lambert et al. (2013) - Two-stage meta-analysis of GWAS with late-onset AD

Lambert et al. (*17*) performed a meta-analysis across four published AD case-control studies from the IGAP consortium, including the Alzheimer's Disease Genetics Consortium (ADGC), Genetic and Environmental Risk in Alzheimer's Disease (GERAD), European Alzheimer's Disease Initiative (EADI), and Cohorts for Heart and Aging Research in Genomic Epidemiology (CHARGE). In total, n=54,162 samples were included in the meta-analysis, which was performed using a fixed-effects inverse variance-weighted method. Before the combination of the summary statistics, the study-specific genomic inflation factors were estimated and their square roots were used to scale the standard errors of the beta coefficient. Stage 1 summary statistics were integrated into the AD Atlas.

Beecham et al. (2014) - GWAS Meta-analysis of neuropathologic features of AD

Beecham et al. (*18*) performed a GWAS using harmonized neuropathological data and genotyping data from n=4,914 brain autopsies. Samples were contributed by the NIA Alzheimer's Disease Centers (ADCs) and ADGC-collaborating studies. Genome-wide association analysis was performed for 14 traits in total, including neuropathological AD features and other brain pathologies. The analysis was performed for each cohort separately. For binary traits, logistic regression was performed and for ordinal traits, polytomous logistic regression. The first three principal components were included as covariates to account for population structure. Inverse-weighted meta-analysis was performed afterward using METAL, accounting for small sample sizes and incomplete phenotyping data by only regarding specific sets for analysis (further described in (*18*)).

Deming et al. (2016) - GWAS with cerebrospinal fluid clusterin

Deming et al. (*19*) used cerebrospinal fluid clusterin (*CLU*) levels as an endophenotype for a GWAS using data from n=673 individuals from the Charles F. and Joanne Knight Alzheimer's Disease Research Center (Knight ADRC) (n=400) and the ADNI (n=273). ADNI and Knight ADRC datasets were combined and the log-transformed and standardized values tested for normality. To test for the association of CSF clusterin levels, statistical analysis was performed using an additive model in PLINK, and study, age, gender, and the first two principal components were used as covariates.

Deming et al. (2017) - GWAS with AD endophenotypes

Deming et al. (*20*) performed a genome-wide association analysis of three endophenotypes (CSF levels of amyloid beta - $A\beta_{42}$, tau and phosphorylated tau - $ptau_{181}$) using data collected from n=3,146 participants across nine studies, including the Knight ADRC, ADNI, Predictors of Cognitive Decline Among Normal Individuals (BIOCARD) and the Mayo clinic. Raw protein levels were log-transformed and normalized within each study before combination for association testing using an additive linear regression model. Study, age, sex, and the first two principal components were tested for confounding using a step-wise regression analysis and included as covariates for each protein where applicable.

Huang et al.  (2017) - GWAS with AD age of onset

Huang et al. (*21*) conducted a genome-wide survival association study to identify genetic loci associated with AD age at onset. Genotyped samples (n=40,255) from IGAP, including the ADGC, GERAD, EADI, and CHARGE, were used to perform genome-wide Cox proportional hazards regression using an additive model. Sex, site, and the first three (four for EADI) principal components from EIGENSTRAT were included as covariates in all models. This analysis was performed for each dataset separately and then combined using inverse-variance meta-analysis using METAL.

Marioni et al. (2018) - GWAS on family history of Alzheimer's disease

Marioni et al. (*22*) used samples from the UK Biobank (UKBB) cohort to conduct a GWAS using an AD-proxy phenotype. Analysis was conducted separately for maternal and paternal AD. GWAS was conducted using an additive model and as outcome, the residuals of a linear regression model of maternal/paternal AD status on age of parent (at death or self-report), assessment center, genotype batch, array, and genetic principal components were used. The results were subsequently combined in two meta-analyses, performed using a standard errorweighted meta-analysis in METAL; 1) meta-analysis of UKBB maternal and paternal analysis (n= 314,278) and 2) meta-analysis of UKBB and published stage II summary statistics from IGAP (*17*) (total n = 388,324). Summary statistics of both meta-analyses are included in the AD Atlas.

Kunkle et al. (2019) - Genetic meta-analysis of diagnosed Alzheimer's disease

Kunkle et al. (*23*) conducted a genetic meta-analysis across consortia of IGAP, which includes the ADGC, GERAD, EADI, and CHARGE. In total, the analysis included 35,274 AD cases and 59,163 controls (total n=94,437). In discovery stage 1, an additive genotype model was used to test for associations between case-control status and genotype for each dataset. Models were adjusted for age (age at onset or age at last exam), sex and principal components. Results across cohorts were combined using inverse-variance meta-analysis as implemented in METAL. In stage 2, replication analysis was carried out using a custom genotyping chip (*17*) and stages 3A and 3B provided further replication for variants in regions not well captured by the chip. Summary statistics for stages 1, 2 (combined stage 1 and stage 2 p-values), 3A, and 3B were downloaded and merged, such that p-values measured in later stages (2, 3A, and 3B) were reported (if available) for each SNP.

Jansen et al. (2019) - Genetic meta-analysis using clinically diagnosed and proxy AD cases

Jansen et al. (*24*) performed a large-scale genome-wide association analysis using both clinically diagnosed AD and AD-by-proxy cases (total n=455,258). This included n=79,145 samples with clinically diagnosed AD case-control status across three consortia; Alzheimer's disease working group of the Psychiatric Genomics Consortium (PGC-ALZ), IGAP, and the Alzheimer's Disease Sequencing Project (ADSP), as well as n=376,113 samples with a weighted AD-by-proxy phenotype from the UKBB. Here, parental AD status from self-report questionnaires was used to generate a score where the number of affected parents was included and unaffected parents were weighed by their age or age at death. Association analyses were performed for each cohort using linear (UKBB - adjusted for ancestry principal components, age, sex, genotyping array, assessment center) and logistic regression (ADSP - adjusted for sex, batch, ancestry principal components; PGC-ALZ - adjusted for sex, batch, ancestry principal components, and age). For IGAP, summary data was used. All cohorts were meta-analyzed using a multivariate genome-wide meta-analysis that takes into account the partial overlap between cohorts by defining a custom per SNP test statistic. Summary statistics from this analysis (phase 3) were downloaded and integrated.

Wightman et al. (2021) - Genetic meta-analysis using clinically diagnosed and proxy AD cases

Wightman et al. (*25*) extended the analysis of Jansen et al. [273] to include samples from additional cohorts, including Finngen and GR@CE. A comprehensive list of cohorts can be found in (*25*). Analysis and meta-analysis were conducted as described above for Jansen et al. using a total of n=762,917 samples. Summary statistics, excluding the 23andMe data, were downloaded and integrated.

Bellenguez et al. (2022) - Genetic meta-analysis using clinically diagnosed and proxy AD cases

Bellenguez et al. (*26*) meta-analyzed a large dataset of clinically diagnosed case-control samples from cohorts across the European Alzheimer & Dementia Biobank (EADB) consortium and proxy-AD/Dementia cases from the UKBB. In total, n=487,511 samples were included; 39,106 clinically diagnosed AD cases, 46,828 proxy-AD/Dementia cases, and 401,577 controls. Proxy-AD/Dementia status was determined using self-report questionnaires and participants were included if at least one biological relative (parent or sibling) was reported to have dementia. Association analysis was performed in each dataset separately using logistic regression and an additive genetic mode. For the UKBB dataset, a logistic mixed model was used. All analyses were adjusted for principal components and genotyping center where necessary. To combine results across studies, METAL was used to perform an inverse-variance meta-analysis. Summary statistics of this analysis (stage I) were downloaded and integrated.

###### 2.2.2 Brain co-expression networks

Wan et al. (2020) - Meta-analysis of the human brain transcriptome

Wan et al. (*27*) conducted a consensus gene co-expression analysis using RNA-seq data to evaluate and robustly identify AD-related molecular signatures. The authors used data from three postmortem brain studies (*28–30*) collected within the AMP-AD consortium. Covariate adjustment was performed for each cohort separately, including diagnosis, sex, identified biological and technical covariates, and donor information (as random effect). Multiple gene co-expression network inference methods to n=2,114 samples. Subsequently, the resulting networks were merged into a single meta-network using an ensemble network inference algorithm. The samples were collected from seven distinct brain regions, resulting in seven tissue-specific meta-co-expression networks - DLPFC, TCX, CBE, inferior frontal gyrus (IFG), superior temporal gyrus (STG), frontal pole (FP), and parahippocampal gyrus (PHG). This data is publicly available via Agora, the results explorer of the AMP-AD Knowledge Portal, and was downloaded from Synapse.

###### 2.2.3 Brain partial correlation networks

Batra et al. (2022) - The landscape of metabolic brain alterations in Alzheimer's disease

Batra et al. (*31*) investigated metabolic changes in the brain using n=500 postmortem DLPFC brain tissue samples from the ROS/MAP cohorts. The samples included n=352 females and n=148 males. In total, out of 500 individuals, n=220 were diagnosed with AD, n=119 MCI and n=153 without cognitive impairment, and eight with other forms of dementia. Non-targeted metabolomics profiling was performed using the Metabolon platform and after quality control 667 metabolites were used to compute a partial correlation-based GGM. P-values of the resulting partial correlations were corrected using the Bonferroni method and partial correlations with $P_{adj} \leq 0.05$ were included in the network. Metabolon- specific COMP IDs were used as metabolite identifiers for integration into the AD Atlas.

###### 2.2.4 Differential analysis in AD cohorts

Wan et al. (2020) - Meta-analysis of the human brain transcriptome

As described above, Wan et al. (*27*) used RNA-seq data collected across three postmortem brain tissue studies covering seven different brain regions. Differential expression analysis and meta-differential expression analysis across brain regions were performed for n=778 samples using weighted fixed- and mixed-effect linear models. Covariate adjustment was performed for each cohort separately and included diagnosis, sex, identified biological and technical covariates, and donor information (as random effect). Differentially expressed genes were determined as those with an adjusted PF DR ≤ 0.05. Both brain region-specific and meta-analysis differential expression results were downloaded from Synapse. Four different models were included in the AD Atlas; AD Diagnosis (males and females): changes in gene expression between AD case and controls; AD Diagnosis x age of death (AOD) (males and females): changes in gene expression between AD cases and controls and whether AOD has an impact;

AD Diagnosis x Sex (females only): changes in gene expression between female AD cases and controls; AD Diagnosis x Sex (males only): changes in gene expression between male AD cases and controls.

Johnson et al. (2020) - Consensus proteomic analysis of Alzheimer's disease brain

Johnson et al. (*32*) performed a large-scale consensus proteomics analysis of AD brain across the AMP-AD consortium, including BLSA, Banner, MSBB and ACT. DLPFC tissue samples from a total of 453 control, asymptomatic AD and AD brains were analyzed using LFQ-MS proteomics. After quality control, n=419 samples and 3334 proteins were used for further analysis. Differentially abundant proteins were identified using one-way ANOVA followed by Tukey's comparison post hoc test across control, asymptomatic AD and AD brain tissue samples. The results of this analysis were downloaded. Proteins were categorized as unchanged (Tukey's $P > 0.05$), up-regulated and down-regulated in AD (Tukey's $P \leq 0.05$ and $log_2$(FC) > or < 0, respectively) for the AD Atlas. Uniprot IDs were used for integration.

Johnson et al. (2022) - Large-scale deep multi-layer analysis of Alzheimer's disease brain

Johnson et al. (*33*) performed a large-scale consensus proteomics analysis of AD brain obtained from the autopsy collections of Banner and ROS/MAP. DLPFC and BA9 tissue samples from a total of 516 control, asymptomatic AD and AD brains were analyzed using a TMT-MS proteomics approach. After quality control, n=488 samples and 8619 proteins were used for further analysis. Differentially expressed proteins were identified using one-way ANOVA followed by Holm post hoc correction of all pairwise comparisons. The results of this analysis were downloaded. Proteins were categorized as unchanged (Holm's $P > 0.05$), up-regulated and down-regulated in AD (Holm's $P \leq 0.05$ and $log_2$(FC) > or < 0, respectively) for the AD Atlas. Uniprot IDs were used for integration.

##### 2.2.5 Metabolic associations with AD (endo-)phenotypes

Metabolite associations with AD (endo-)phenotypes that were integrated into the AD Atlas were identified through metabolome-wide association studies (MWAS) using samples from ADNI phases 1, GO, and 2.

MahmoudianDehkordi et al. (2019) and Nho et al. (2019) - Bile acid MWAS with AD biomarkers

Using n=1555 baseline serum samples of fasting participants of ADNI, including cognitively normal individuals, individuals with early or late MCI, as well as AD cases, targeted metabolomics profiling of bile acids was performed using the Biocrates Life Sciences Bile Acids Kit (BIOCRATES Life Science AG, Insbruck, Austria). Bile acid levels were adjusted for medication effects. Linear regression models were used to analyze the association of the levels of 15 bile acids with 19 AD-related traits, including CSF and imaging biomarkers (*34*, *35*). Age, sex, study phase, BMI and APOE*ε4 status were included as covariates. Significant associations that were integrated into the AD Atlas were determined by applying a Bonferroni significance threshold of $P \leq 3.33 \times 10^{-3}$ (0.05/15).

Arnold et al. (2020) - MWAS with AD biomarkers

Using n=1517 baseline serum samples of fasting participants pooled from ADNI phases 1, GO, and 2, Arnold et al. (*36*) analyzed the association of 19 AD-related traits, including CSF and imaging biomarkers, with the levels of 140 metabolites using standard linear and logistic regression. Metabolite levels were adjusted for significant medication effects using step-wise backward selection. Regression models were adjusted for age, sex, ADNI study phase, and the number of copies of APOE*ε4 and could also include BMI and education (selected by backward selection). The significant associations that were integrated into the AD Atlas were determined by applying a Bonferroni significance threshold of $P \leq 9.09 \times 10^{-4}$ (0.05/55), as the number of independent metabolic features was determined to be 55.

Batra et al. (2022) **-** The landscape of metabolic brain alterations in Alzheimer's disease

As described above, Batra et al. (*31*) investigated metabolic changes in the brain using n=500 *post mortem* DLPFC brain tissue samples from the ROS/MAP cohorts. Non-targeted metabolomics profiling was performed using the Metabolon platform and after quality control 667 metabolites were tested for association with eight AD-related traits: clinical diagnosis at the time of death, level of cognition proximate to death, cognitive decline during lifetime, amyloid-β load, tau tangle load, global burden of AD pathology, NIA-Reagan score and neuropathology diagnosis. Generalized linear models with traits as response variable and appropriate link functions were used to test for association between metabolite concentrations and traits. Age, sex, BMI, *post mortem* interval (PMI), number of years of education and the number of APOE*ε4 alleles were included as co-variates in the models. P-values were corrected using the Benjamini-Hochberg method to account for multiple testing. Summary statistics for significant associations ($P_{adj} \leq 0.05$) were downloaded and integrated into the AD Atlas using Metabolon specific COMP IDs as metabolite identifiers.

#### 2.3 Additional analysis

##### 2.3.1 Partial correlation networks

Johnson et al. (2020) - Consensus proteomic analysis of Alzheimer's disease brain

As described above, Johnson et al. (*32*) performed a meta-analysis of brain tissue across four studies of the AMP-AD consortium. Minimally regressed (batch- and site-corrected) data were downloaded from Synapse and a GGM was estimated to construct a partial correlation network as described in (*37*), correcting for age, sex, and PMI. Uniprot IDs were used to integrate the data.

Johnson et al. (2022) - Large-scale deep multi-layer analysis of Alzheimer's disease brain

As described above, Johnson et al. (*33*) performed a meta-analysis of brain tissue across two studies of the AMP-AD consortium. Minimally regressed (batch- and site-corrected) data were downloaded from Synapse and a GGM was estimated to construct a partial correlation network as described in (*37*), correcting for age, sex, and PMI. Uniprot IDs were used to integrate the data.

Alzheimer's Disease Neuroimaging Initiative (ADNI)

Two metabolite partial correlation networks were estimated for metabolites measured by the targeted AbsoluteIDQ-p180 and Bile Acids Kit (BIOCRATES Life Science AG, Innsbruck, Austria) using samples from ADNI (*38*) phases 1, GO, and 2. Detailed data processing is described in (*36*) (p180) and (*34*) (bile acids). In summary, the AbsoluteIDQ-p180 dataset included n=1517 baseline fasting serum samples and 139 metabolites after quality control and the bile acid dataset included a total of 15 bile acids for n=1464 baseline fasting serum samples after quality control. Metabolites were adjusted for medications and dietary supplements. GGMs were estimated for each dataset separately as described in (*37*), applying a significance threshold of 0.05 after Bonferroni correction for all possible edges in the respective models. Age, sex, number of *APOE**ε4 alleles, and education were included as covariates. Biocrates-given metabolite names were used for integration.

Metabolomics data are available via the Alzheimer's Disease (AD) Knowledge Portal (*https://adknowledgeportal.org*).

AbsoluteIDQ® p180 kit (Biocrates Life Sciences AG, Innsbruck, Austria):

- *https://doi.org/10.7303/syn5592519* (ADNI-1)
- *https://doi.org/10.7303/syn9705278* (ADNI-GO/-2)

Bile Acids Kit (Biocrates Life Sciences AG, Innsbruck, Austria):

- *https://doi.org/10.7303/syn12036817.1* (ADNI-1)
- *https://doi.org/10.7303/syn9779093.1* (ADNI-GO/2)

##### 2.3.2 Genetic associations with blood metabolites and Alzheimer's (endo-)phenotypes

Alzheimer's Disease Neuroimaging Initiative (ADNI)

Genome-wide genotyping data of ADNI-1/GO/2 participants were collected using the Illumina Human 610-Quad, HumanOmni Express, and HumanOmni 2.5M BeadChips. Before imputation, standard quality control (QC) procedures of GWAS data for genetic markers and subjects were performed (variant call rate < 95%, Hardy-Weinberg-Equilibrium test $P < 1 \times 10^{-6}$, and minor allele frequency (MAF) < 1%, participant call rate < 95%, sex check and identity check for related relatives). Then, non-Hispanic Caucasian participants were selected using HapMap 3 genotype data and MDS analysis. Genotype imputation was performed for each genotyping platform separately using the Haplotype Reference Consortium (HRC) reference Panel r1.1 and merged afterward, resulting in data on n=1,576 individuals and 20,779,509 variants. Using this dataset, we ran GWAS analyses for each outcome (A-T-N-C measures, clinical diagnosis, and metabolite levels) that included outcome-specific sets of covariates, including age, sex, study phase, education, and *APOE**ε4 status. All models testing for association with metabolite levels were corrected for clinical diagnosis.

##### 2.3.3 Genetic associations with brain metabolites

We analyzed brain tissue samples of participants of the ROS/MAP cohorts and the Mayo using the non-targeted Metabolon Discovery HD4 platform. ROS/MAP samples were taken from the DLPFC (n = 459), in Mayo samples were taken partly from the TCX (n = 159) and the CBE (n = 177). Metabolic profiles after thorough QC as described in Batra et al. (*31*) were available for 667 metabolites in ROS/MAP and 658 in Mayo, with an overlap of 576 metabolites available in both datasets. Imputed genotypes were available for all samples. Genotype QC included filters for MAF ≥ 0.05, individual genotyping rate ≥ 95%, and genotype call rate ≥ 95%, as well as a test $P \geq 1 \times 10^{-5}$ for Hardy-Weinberg-Equilibrium. These filters yielded a total of 4.386 million genotypes, 5.434 million genotypes, and 5.415 million genotypes in ROS/MAP (DLPFC), Mayo (TCX), and Mayo (CBE), respectively. The overlap in genotypes was 4.062 million across datasets. Using z-scored metabolite levels (centered to zero mean and unit variance), we first ran independent genome-wide association studies with metabolite traits (mGWAS) analyses in each brain region for all available metabolites, where the ROS/MAP dataset, being the largest dataset available, was considered to serve as the phase 1 discovery sample. Linear regression was performed for 667 metabolites adjusting for age at death, sex, PMI, and neuropathology-based diagnosis using PLINK2. Replication analyses in the two brain regions available in Mayo TCX, n = 159; CBE, n = 177) were performed analogously for 658 metabolites. Mayo datasets were used as phase 2 replication sets. We afterward conducted an inverse-weighted meta-analysis across all three studies using random-effect models to account for between-study variance caused by differences in cohort recruitment, sample collection, and brain region specificity of metabolic readouts. We included the summary statistics of the meta-analysis for the 576 overlapping metabolites. For 91 metabolites only measured in the ROS/MAP cohort, we included discovery phase p-values. Metabolon-given COMP IDs were used as metabolite identifiers for integration.

### 3 AD ATLAS USER INTERFACE/FEATURES

The AD Atlas is a network-based data integration resource for investigating Alzheimer's disease, its biomarkers, and associated endophenotypes in a multi-omics context. To facilitate easy access and exploration of the resource, we implemented a network-based web interface through which users can query the database by entering one or more phenotypes, genes or metabolites. The network browser provides an interactive interface to inspect generated networks, as well as enrichment tools for downstream analyses.

#### 3.1 Network browser

The network browser allows users to generate and visually explore Alzheimer’s disease related (i) trait-centric, (ii) gene-centric or (iii) metabolite-centric molecular subnetworks. Users provide trait(s), gene(s) or metabolite(s) of interest, that are then annotated with associations from the AD Atlas. The initial set of query entities can be expanded to the functional 1- or 2-step neighborhood. Furthermore, the resulting network can be filtered by sample type, tissue or brain region to provide comprehensive, context-specific results. The association structure between traits, genes and metabolites is visually represented as an interactive graph in which nodes (representing biological entities, such as metabolites or genes) can be clicked to highlight immediate neighbors and edges (indicating associations) and reveal additional information, such as the underlying SNPs and their corresponding *p*-values. Generated networks can be downloaded as an interactive graph (.html file) or as an edge list (.csv file) for further analysis, i.e., using network-based software, such as Cytoscape. In the following, we provide in depth explanations of the search query parameters.

##### Molecular subnetwork generation

Users can provide one or more entities (traits, meta-traits, genes or metabolites) to query the underlying database. We provide 3 distinct entry points (further discussed in the manuscript);

i. Trait- or meta-trait-centric subnetworks
ii. Gene-centric subnetworks
iii. Metabolite- or pathway-centric subnetworks

##### Network expansion

Prior to building the network, the initial set of input genes or metabolites can be expanded to include the 1-step or 2-step functional neighbors. For gene-centric networks (input: gene(s)) these neighbors can be defined via gene co-expression, protein co-abundance, and/or eQTL co-regulation networks. For metabolite-centric networks (input: metabolite(s)), expansion is performed using partial correlation networks (GGMs). Context filtering is applied for this step if the user has specified a sample type, tissue, or brain region. Once the selection of input nodes has been expanded, they are annotated with associated entities.

**Significance threshold**

Inferred links between and within omics layers are filtered by applying basic significance thresholds. These are predominantly study-specific and can be found in **Supplementary Table 2**. For genetic associations, that is links between genes and metabolites (inferred from mGWAS) and genes and traits (inferred from GWAS), users can select either a gene-wise or genome-wide threshold. Thereby, the gene-wise cutoff is an adapted Bonferroni cutoff that is defined by $p$-value $\leq 0.05/\#SNPs_{geneA}$ , where $\#SNPs_{geneA}$ is the number of SNPs that have been annotated to gene A. The annotation of SNPs to gene was performed using the variant annotation tool SNiPA. This gene-wise significant threshold is less stringent than the genome-wide cutoff of at $p$-value $\leq 5x10^{-8}$ and will yield more results.

**Context/Tissue-specific filtering**

Edges can be filtered by sample type, tissue, or brain region. Currently, links established through tissue-specific eQTL analysis (COREGULATION) taken from GTEx (*2*) and large-scale analysis of cortical cis-eQTLs by Sieberts et al. (*4*) can be filtered by 49 tissues, of which 13 are brain-specific. Furthermore, gene co-expression networks (COEXPRESSION) were constructed for seven different brain regions by which these links can be filtered. GENETIC_ASSOCIATION (gene-metabolite), METABOLIC_ASSOCIATION, COABUNDACE, and PARTIAL_CORRELATION edges can be filtered by sample type, which currently includes brain, plasma, serum, and urine. If nothing is specified, this setting defaults to no filtering. The filtering, when specified, is also applied to the network expansion step.

**Additional visualization options**

To assess the extent and direction of dysregulation in AD, data from differential analysis in AD cohorts can be visually projected onto the built multi-omics networks. This includes information about genes that are differentially expressed in the brain (*27*), called differentially expressed genes (DEGs), and proteins that are differentially abundant in the brain (*32*, *33*), called differentially abundant proteins (DEPs). Up-regulation, down-regulation, or non-regulation at the transcriptional level is indicated by coloring the gene nodes in the network in red, blue, and light green, respectively. Differential protein abundance is indicated by coloring the edge of the gene nodes, using the same color coding as for genes.

#### 3.2 Subnetwork annotation and analysis

**Node annotations**

Entities of the generated molecular network are annotated with additional information such as biochemical pathway and links to other well-known databases (including HMDB, GeneCards and Agora) or descriptions (for AD traits). Furthermore, all biological entities are directly linked to the network allowing the search for and inspection of interesting nodes even in large and dense networks. Metabolite, gene and trait annotations can be downloaded as a CSV file.

**Network analysis**

General network statistics such as node degree and node/edge type proportions as well as different network representations are provided to aid network analysis and exploration.

#### Enrichment analysis

Enrichment-based analysis can be performed for the generated molecular subnetworks using gene-level (GO terms, EnrichR, g:Profiler, Rummagene, RummaGEO, biodomains, differential expression) or metabolite-level (using Metabolon's and Biocrates' annotation into classes and super-/sup-pathways) annotations. The biological entities within the displayed network are tested for term enrichment against all entities within the AD Atlas if not stated otherwise. The results of this analysis can be downloaded as CSV files.

### 4 SHOWCASES

In the following, we used the AD Atlas online resource (www.adatlas.org) to explore AD-related molecular hypotheses by building context-specific networks surrounding either genes or metabolites. In the **Showcases 1** and **2**, we exemplify how molecular networks constructed using the AD Atlas may assist in drug repositioning efforts. Drug repositioning has gained increasing attention as a promising alternative to *de novo* drug development (*39*). This method involves repurposing approved compounds for novel disease contexts. However, a significant challenge to enabling such endeavors is the evaluation of suggested candidates or drug targets within a multi-omics framework, which often entails additional time-consuming and costly analysis. In **Showcases 3** and **4**, we show two explorative examples, in which we used the AD Atlas to investigate molecular disease mechanisms; we thereby demonstrate the utility of our resource to formulate and prioritize metabolic and immune-related hypotheses.

To test the validity and relevance of the resulting multi-omics networks, we evaluated them with regard to their network structure, relevance to AD (association with AD-related phenotypes or evidence of dysregulation at the metabolite, gene or protein level in AD) and gene set enrichment analysis. To strengthen confidence in both the network generation procedure as well as the quality of the harmonization of underlying data, we compared obtained findings with established knowledge in the field.

We performed additional statistical evaluation across several network statistics by calculating empirical *p*-values derived from a background distribution of 1000 randomly generated networks using permutations of the same type of query input. To this end, we calculated empirical *p*-values for the following network properties: network size – the number of nodes (genes, metabolites and traits in the network); proportion of entities (genes or metabolites) significantly associated with AD-related traits (number of trait-associated entities in network / total number of entities in network); proportion of genes significantly associated with AD-related traits (number of trait-associated genes in network / total number of genes in network); proportion of metabolites significantly associated with AD-related traits – analogous to the previous; proportion of differentially expressed genes (DEGs) (DEGs in network / all entities tested for DE in network); proportion of differentially expressed genes (DEPs) – analogous to DEGs. The empirical *p*-values were estimated from the background distribution of the proposed statistics. For each showcase we generated 1000 random networks using the same input parameters but replacing the hypothesis-guided input genes/metabolites with randomly selected genes/metabolites. For the trait-centric network, the genes and metabolites associated with the input trait were replaced by random entities of the respective type and then subjected to network generation by annotation of direct associations (GENETIC_ASSOCIATION and METABOLIC_ASSOCIATION edges). We then approximate an empirical *p*-value using the empirical cumulative distribution function (function *ecdf* from R package *stats*) of the random network statistics and report the fraction of observations larger than the statistic observed in the respective showcase.

All showcases, including networks and enrichment results, can be interactively explored online at [www.adatlas.org/?showcases](www.adatlas.org/?showcases).

**A - Molecular subnetwork surrounding *APOE* and *CLU***

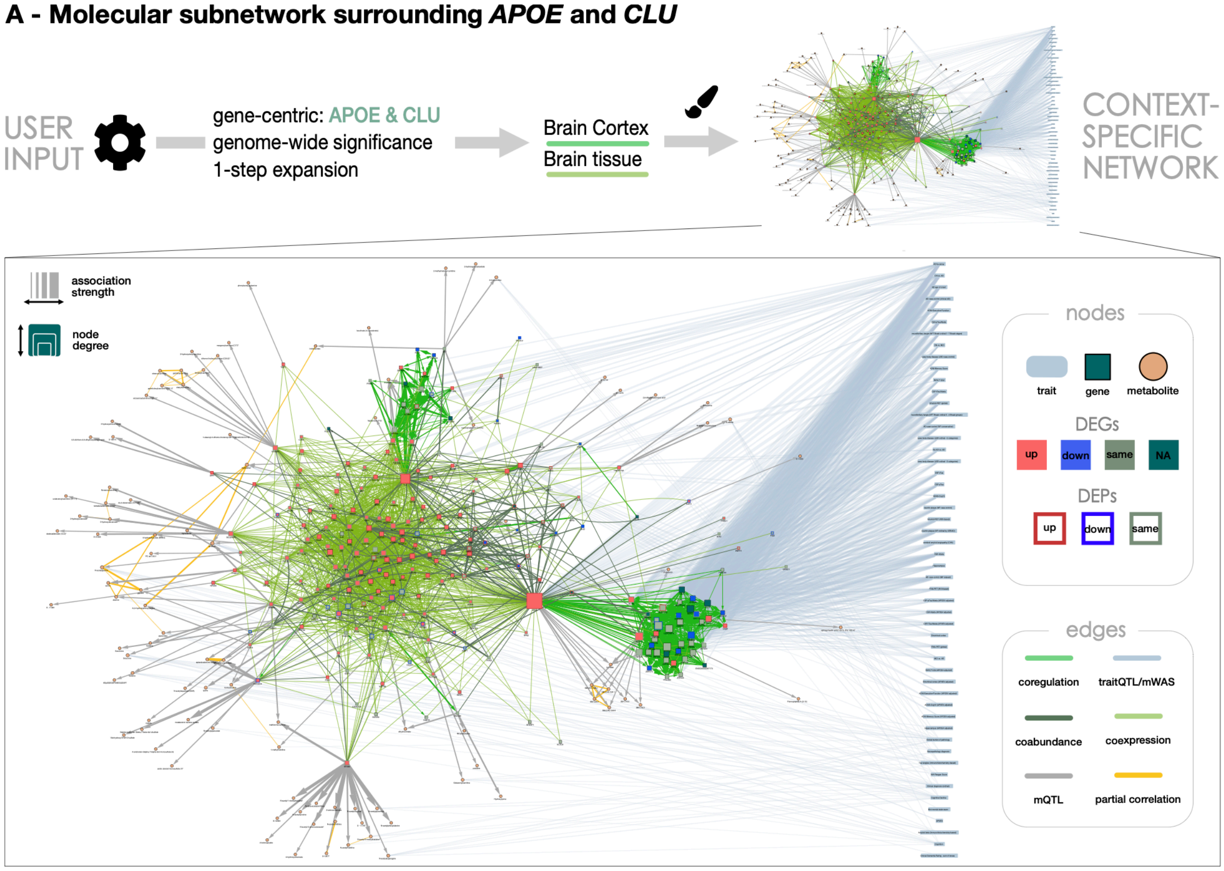

**B - Selected gene set enrichment results**

| *Drug Perturbations from GEO down (EnrichR)* | *rank* | *overlap** | *q-value*** | *OR**** | *genes* |
|---|---|---|---|---|---|
| **Candesartan** DB00796 rat GSE2739 sample 2687 | 2 | 26/369 | 3.20e-10 | 6.72 | APP, SERPINE2, SDC4, SLC1A3, STMN4, AQP4, NDRG2, CLU, CST3, VTN, GJA1, SPOCK2, APOD, PHGDH, APOE, PTGDS, NTSR2, SREBF1, ATP1B2, S100B, AGT, BCAN, SYT11, DNAJA4, TMBIM6, ITM2B |
| **Levetiracetam** DB01202 rat GSE2880 sample 2777 | 17 | 13/181 | 1.99e-05 | 6.53 | WFS1, STMN4, ENO1, PTN, NDRG2, S100B, CST3, DPYSL2, TRIM35, CPE, APOE, ITM2B, ITM2C |
| **Levetiracetam** 5284583 rat GSE2880 sample 2669 | 22 | 19/399 | 2.21e-05 | 4.29 | APP, ENO1, ATP1B2, CLU, AGT, CNN3, GCSH, GLUD1, EDNRB, TRIM35, PHGDH, ACSBG1, TMBIM6, APOE, VIM, PTGDS, ITM2B, NTSR2, ITM2C |

*overlap network (nominator) with drug signature gene set (denominator) **adjusted *p*-value using Benjamini-Hochberg ***odds-ratio

**Figure S12. *APOE*/*CLU* network identifies repositioning candidates. (A)** Multi-omics network surrounding *APOE* and *CLU* as contained in the AD Atlas. **(B)** Gene set enrichment analysis for drug-associated gene expression changes using EnrichR. By focusing on signatures which are opposed to the overall change in AD, we identify Levetiracetam and Candesartan as most promising. Genes that overlap between drug signature sets (column 'genes') and the network are color coded to indicate direction of expression change or protein abundance in disease (red: up-regulation, blue: down-regulation). Genes with opposing direction of change at the transcript (color code) and protein level are underlined.

#### 4.1 Showcase 1: Molecular network of lipid metabolism and transport identifies known repositioning candidates

Multiple genetic risk factors for late-onset Alzheimer's disease (LOAD) have been identified through GWAS (*39*). The earliest was the discovery of the genetic risk exerted by the ε4 allele of apolipoprotein E (*APOE*) (*40*), which was followed by the identification of several risk variants in clusterin (*CLU,* also referred to as *APOJ*) (*41, 42*). Both proteins are involved in lipid metabolism and transport, and we hypothesized that these mechanisms could potentially be targeted by available repositioning candidates. We therefore used the AD Atlas to build a molecular context network (**Supplementary Figure 12A**) surrounding these two genes including their 1-step functional neighborhood defined by gene co-expression, co-regulation data and protein co-abundance data (for definitions, see **Supplementary Box 1**). Nearly a quarter of the genes in the resulting network (57 of 245 genes; $P_{emp}$=0.057; **Supplementary Figure 13**) are genetically linked to AD (endo-)phenotypes through genome-wide significant associations. Furthermore, overlay of differential gene expression and protein abundance data from large-scale case-control studies (*33, 43*) reveals a significant up-regulation in AD both at the level of transcripts and proteins ($P_{emp}$=0.035 and $P_{emp}$=0.029, respectively; **Supplementary Figure 13**).

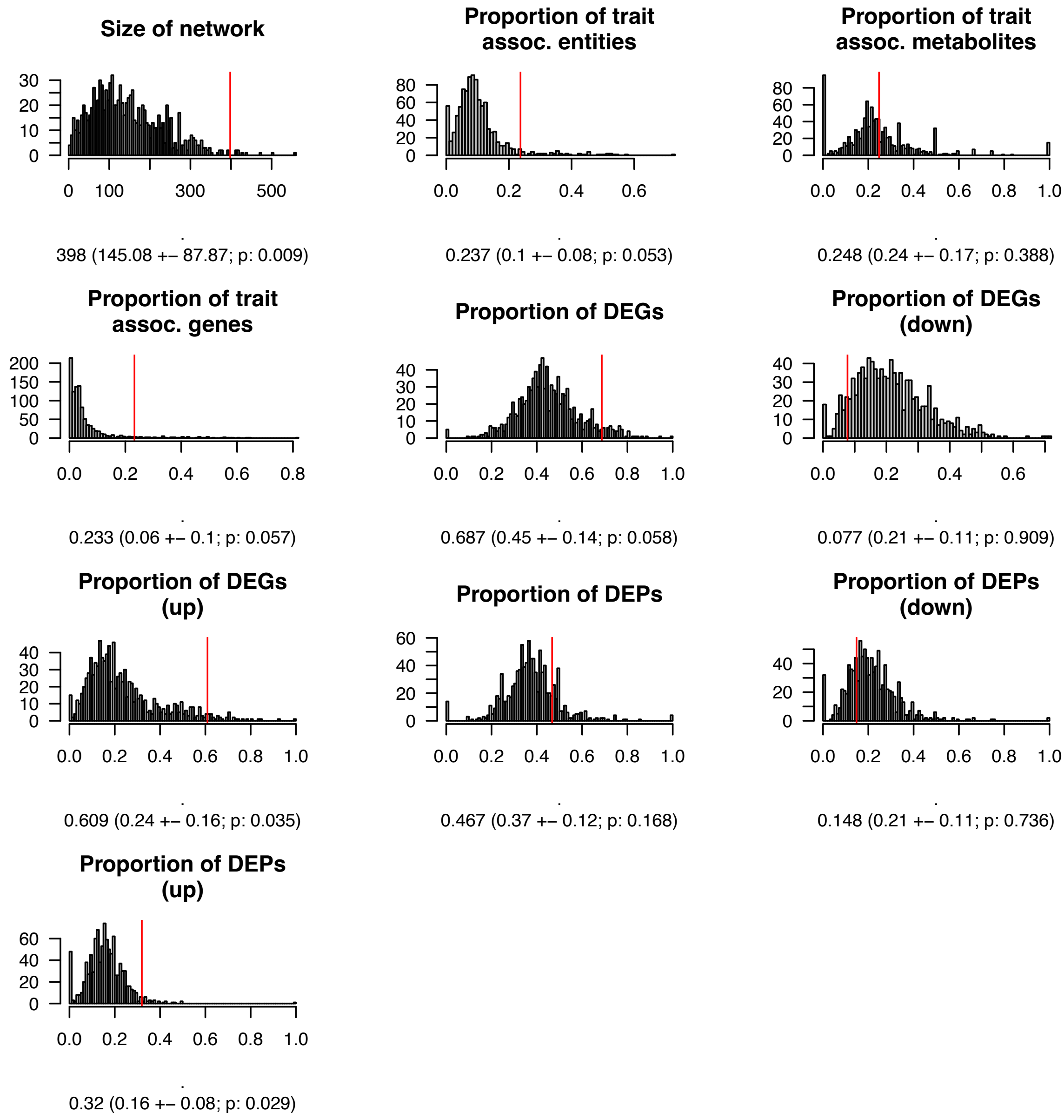

**Figure S13. Background distributions for the APOE/CLU example.** Network properties of 1000 random networks built by selecting genes (n=2) and expanding to their 1-step neighborhood, filtering co-regulation edges to the brain cortex. Red line indicates observed property for the APOE and CLU molecular network (**Supplementary Figure 12**). Observed value with mean, standard deviation and empirical *p*-value are provided below each histogram.

As a network around two central AD risk genes is expected to show significant enrichment in terms of disease relevance, we next explored the potential of this network to identify drug repositioning candidates. As described above, the network surrounding *APOE* and *CLU* displays an up-regulation signature in disease. Therefore, we hypothesize that drugs that perturb these genes in an opposing manner may be the most promising candidates to exert AD-relevant therapeutic effects (*39*). To test this hypothesis, we performed gene set enrichment analysis on all the genes in this network using molecular drug signatures from the EnrichR (*44-46*) database (EnrichR, *Drug Perturbations from GEO down*). This resulted in a list of 100 unique compounds, which we cross-referenced with drugs that have been previously proposed for and tested in clinical trials (*45*, *46*). Selected results of this analysis can be seen in **Supplementary Figure 12B.**

Among others, the list of FDR-significant compounds includes Candesartan ($P$=7.19e-13), an angiotensin receptor blocker typically used for the treatment of hypertension. Candesartan predominantly affects genes in the sub-network that are up-regulated in AD (mRNA: $P_{emp}$=0.004; protein: $P_{emp}$=0.01; **Supplementary Figure 14B**) and has recently been studied in a phase II trial (NCT02646982) to investigate its effect on individuals with mild cognitive impairment that are positive for AD biomarkers. Although no results have been published for this trial, previous clinical trials suggest beneficial neurocognitive effects following Candesartan treatment in older individuals with hypertension and mild cognitive impairment (*47*, *48*). Another interesting candidate that was identified by this analysis is Levetiracetam ($P$=3.81e-07), a medication that is used to treat epilepsy and is currently being studied to determine whether or not it is able to improve synaptic function and reduce amyloid-induced neuronal hyperactivity as a disease-modifying therapy in AD (*49*). With Levetiracetam being investigated in multiple phase II trials (NCT02002819, NCT03489044, NCT03875638) and a low-dose formulation (AGB101) currently being tested in phase II (NCT03461861) and phase III trials (NCT03486938), Levetiracetam is one of the most represented agents among ongoing clinical trials (as of February, 2020) (*49–51*). Furthermore, genes in this network that are affected by Levetiracetam display an up-regulation in AD (mRNA: $P_{emp}$<0.001; protein: $P_{emp}$=0.006; **Supplementary Figure 14A**). Of note, both drugs affect the AD-risk genes *APOE* and *CLU*, which were provided as input, as well as *APP.*

This example showcases how context-specific molecular networks can provide additional AD-relevant insights for known AD risk genes. By applying a simple enrichment approach requiring minimal analysis steps, we identify multiple drug repositioning candidates that are either being tested or have been tested in clinical trials, providing evidence for the relevance of the generated results. Through additional analysis using the AD Atlas we propose Levetiracetam and Candesartan as most promising candidates as their associated list of genes affected in drug screens overlaps significantly with the molecular context network surrounding *APOE* and *CLU* and they affect disease-perturbed genes in an opposing manner. Besides the presented examples, we find additional FDR significant candidates that may be equally interesting to follow up on, including Fluticasone, a glucocorticoid used to treat nasal symptoms (*50*), Dexamethasone, a systemic corticosteroid and Pioglitazone, an FDA-approved drug for type 2 diabetes. Interestingly, these drugs have recently been prioritized and validated using real-world data by studies using more sophisticated drug-repositioning approaches (*51–54*).

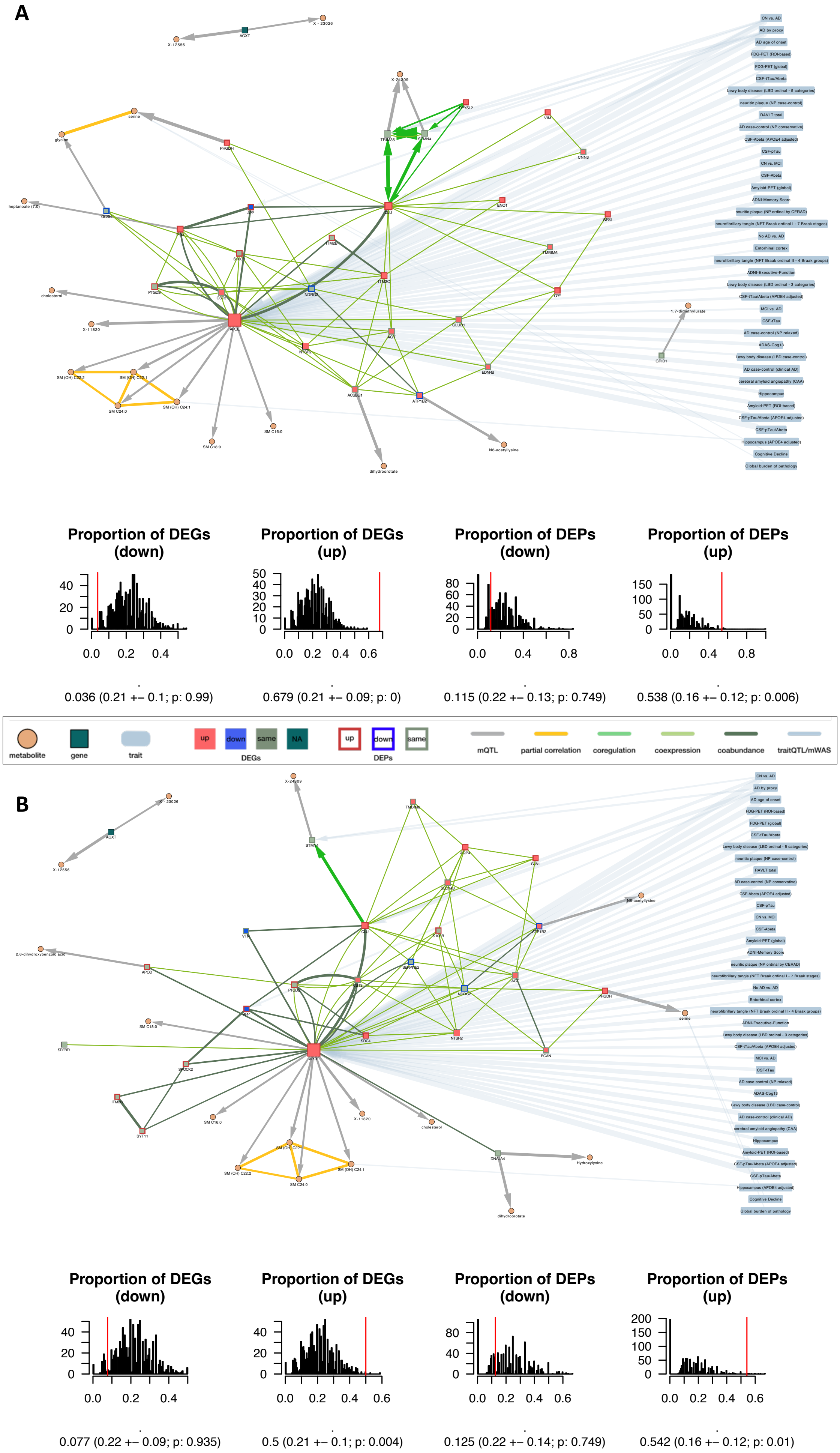
A
Proportion of DEGs (down)
0.036 (0.21 +– 0.1; p: 0.99)
Proportion of DEGs (up)
0.679 (0.21 +– 0.09; p: 0)
Proportion of DEPs (down)
0.115 (0.22 +– 0.13; p: 0.749)
Proportion of DEPs (up)
0.538 (0.16 +– 0.12; p: 0.006)
metabolite
gene
trait
up
down
same
NA
DEGs
up
down
same
DEPs
mQTL
partial correlation
coregulation
coexpression
coabundance
traitQTL/mWAS
B
Proportion of DEGs (down)
0.077 (0.22 +– 0.09; p: 0.935)
Proportion of DEGs (up)
0.5 (0.21 +– 0.1; p: 0.004)
Proportion of DEPs (down)
0.125 (0.22 +– 0.14; p: 0.749)
Proportion of DEPs (up)
0.542 (0.16 +– 0.12; p: 0.01)

**Figure S14. Molecular network of genes affected by repositioning candidates. (A)** Levetiracetam and **(B)** Candesartan. Both networks include genes from distinct genetic loci, as can be seen by using the "Node annotation – Show annotation for genes" function on the left panel of the AD Atlas. Furthermore, they are only connected by a few co-regulation links which often indicate common loci. The genes are connected at the messenger ribonucleic acid (mRNA) and protein level through co-expression (light green) and co-abundance (dark green) edges. Below each network the background distribution of the proportion of differentially expressed genes (DEGs) or differentially abundant proteins (DEPs) from 1000 random gene sets is shown (n=27 genes for Levetiracetam and n=26 genes for Candesartan). Networks were constructed using URL queries of the form: adatlas.org/?geneSymbol={gene symbols taken from EnrichR output}&gtex=Brain%20Cortex.

#### 4.2 Showcase 2: Statin target *ITGAL* links to neuroinflammation through TREM2 signaling

Statins are a class of lipid-lowering drugs that are used to reduce the risk of cardiovascular diseases, such as atherosclerosis and peripheral artery disease. Statins exert their primary therapeutic effect by inhibiting the rate-limiting enzyme in cholesterol biosynthesis, 3-hydroxy-3-methylglutaryl-coenzyme A reductase (HMGCR). In addition, statins have been associated with a wide range of secondary effects (*55*, *56*). Observational studies have reported a possible association between statin use and reduced risk of Alzheimer's disease (*57*–*59*), although the evidence has been inconsistent with reported differences between patient subpopulations as well as individual compounds in the statin drug class (*60*–*62*). To explore whether the observed differences may be attributed to distinct molecular targets of the individual compounds and investigate potential links to AD pathophysiology, we used the drug targets of statins as annotated in DrugBank (*63*) (**Supplementary Figure 15**). We built a context-specific network surrounding these targets (*HMGCR, ITGAL, HDAC2, DPP4, AHR, NR1I3)* and included their 1-step co-expression, co-regulation and protein co-abundance neighbors (**Supplementary Figure 16A**). Of the 321 genes contained in the resulting network, 20% are genetically linked to AD (endo-)phenotypes ($P_{emp}$ = 0.044; **Supplementary Figure 17**) and around 40% show differential expression at the transcript level in the temporal cortex of AD patients ($P_{emp}$ = 0.625; **Supplementary Figure 17**). The immediate neighborhood of each of the six input genes shows that all targets display some degree of dysregulation at the transcript or protein level in AD, either by direct evidence or via their neighbors, as shown in **Supplementary Figure 16B** and **Supplementary Figure 16C** for ITGAL and HMGCR, respectively.

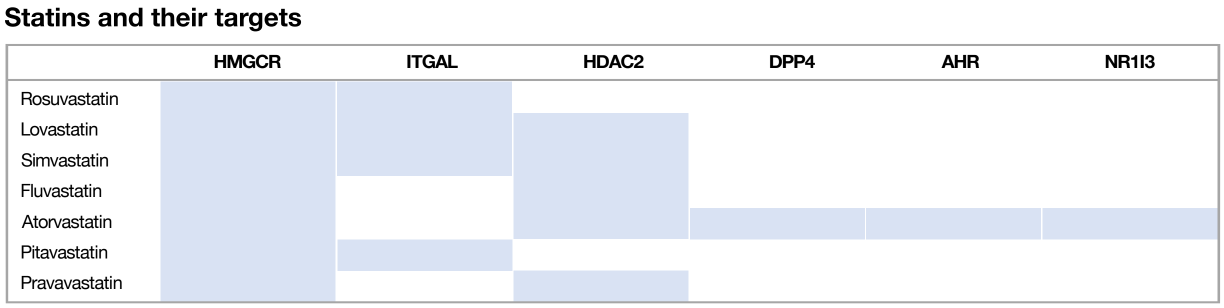

**Figure S15. Statins listed in DrugBank and their annotated targets**

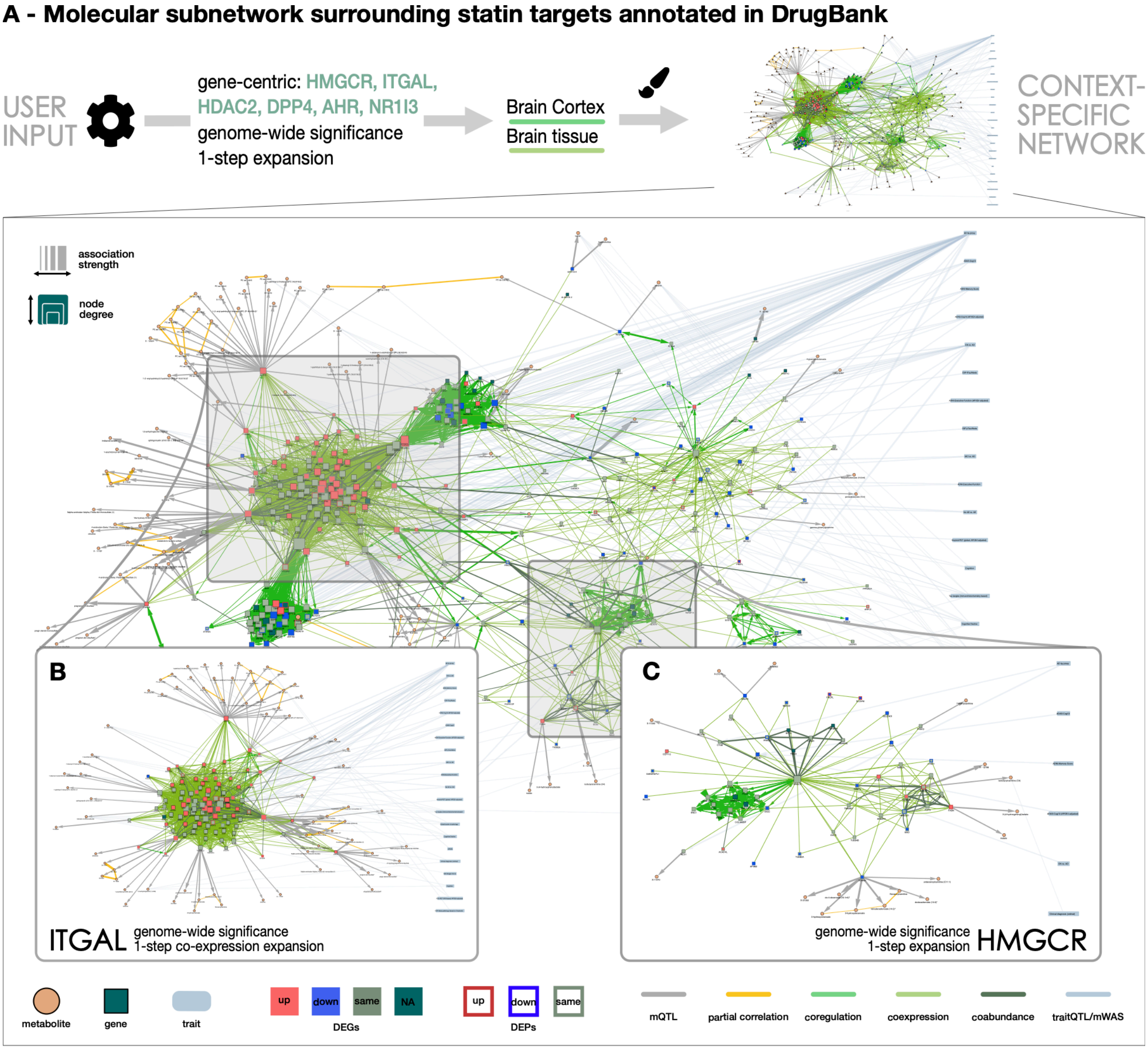

**D - Top 5 gene set enrichment results**

| *WikiPathway 2021 human (EnrichR)* | *overlap** | ***q-value**** *( ITGAL***)* | *OR***** | *genes* |
|---|---|---|---|---|
| **TYROBP causal network** in microglia WP3945 | 17/61 | **1.92e-14** (2.08e-23) | 24.95 | **RBM47**, IGSF6, **PLEK**, **ITGB2**, **TCIRG1**, **SLC1A5**, **TGFBR1**, **PYCARD**, APBB1IP, CD4, **SLC7A7**, ADAP2, BIN2, **ELF4**, CD37, GAL3ST4, TMEM106A |
| Cholesterol Biosynthesis Pathway WP197 | 7/15 | **2.46e-07** (-) | 54.82 | FDPS, SQLE, **HMGCS1**, SC5D, **MSMO1**, **MVD**, HMGCR |
| Cholesterol metabolism (includes both Bloch and Kandutsch-Russell pathways) WP4718 | 10/46 | **2.67e-07** (-) | 17.54 | FDPS, SQLE, **EBP**, **HMGCS1**, **FASN**, SC5D, **MSMO1**, **MVD**, HMGCR, SREBF2 |
| Microglia Pathogen Phagocytosis Pathway WP3937 | 9/40 | **9.26e-07** (1.86e-09) | 18.28 | HCK, **FCER1G**, **ARPC1B**, **ITGB2**, **RAC2**, **TREM2**, PTPN6, PIK3CG, VAV1 |
| Mevalonate pathway WP3963 | 5/7 | **1.40e-06** (-) | 155.67 | FDPS, **HMGCS1**, **MVD**, HMGCR, **ACAT2** |

*overlap network (nominator) with pathway gene set (denominator) **adjusted *p*-value using Benjamini-Hochberg
***enrichment for 1-step co-expression network surrounding ITGAL ****odds-ratio

**Figure S16. Network of statin targets links to TYROBP signaling. (A)** Multi-omics network surrounding the statin targets annotated in DrugBank; *HMGCR*, *ITGAL*, *HDAC2*, *DPP4*, *AHR* and *NR1I3*, as well as a co-expression network surrounding **(B)** only *ITGAL and a* co-expression, co-regulation and co-abundance network surrounding **(C)** only *HMGCR*. **(D)** Gene set enrichment analysis using EnrichR (*WikiPathway* gene set). Genes that overlap between pathway sets (column 'genes') and the network are color coded to indicate direction of expression change or protein abundance in disease (red: up-regulation, blue: down-regulation).

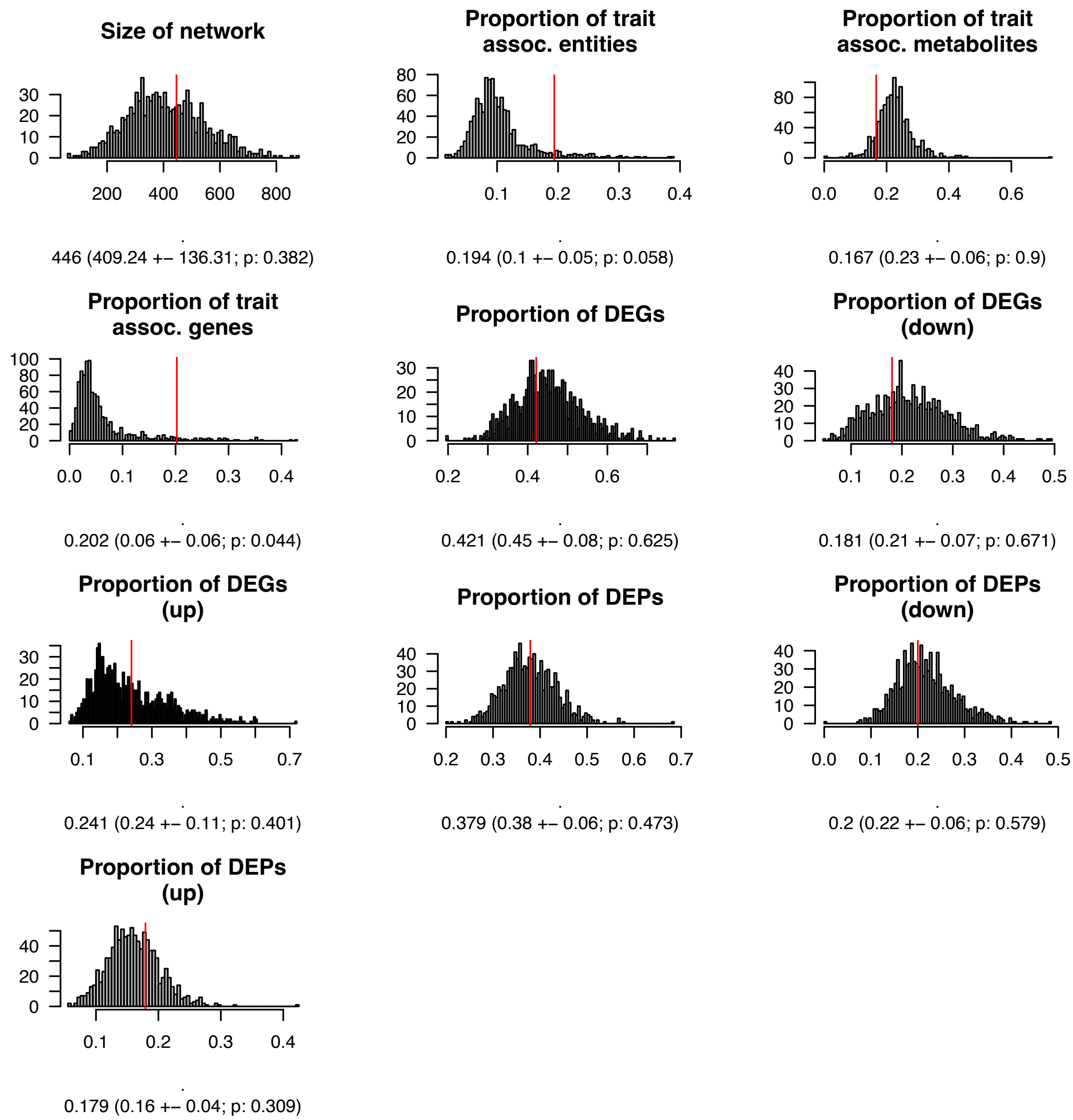

**Figure S17. Background distribution for statins example.** Network properties of 1000 random networks built by selecting genes (n=6) and expanding to their 1-step neighborhood, filtering co-regulation edges to the brain cortex. Red line indicates observed property for the gene-centric molecular network surrounding statin targets (seen in **Supplementary Figure 16**). Observed value with mean, standard deviation and empirical *p*-value are provided below each histogram.

To functionally characterize the pathways targeted by statins further, we used the gene set enrichment functionality of the AD Atlas. Here, we found a significant enrichment for the TYROBP causal network (EnrichR, *WikiPathways*, $P_{FDR}$ = 1.92e-14; **Supplementary Figure 16D**), an immune- and microglia-specific module that has been implicated in LOAD (*64*). This enrichment is driven by the co-expression network surrounding *ITGAL* (**Supplementary Figure 16B**), also known as *CD11a*, a subunit of the integrin leukocyte function associated antigen-1 (LFA-1), which is involved in a variety of immune-related functions (*65*). Interestingly, according to DrugBank, only a subset of statins (Rosuvastatin, Lovastatin, Simvastatin, Pitavastatin) target *ITGAL,* which may explain the inconsistent results reported in studies looking at those compounds vs. other statins. Furthermore, the co-expression neighborhood of *ITGAL* shows evidence of dysregulation in disease with 49 of 103 genes showing an up-regulated expression in AD ($P_{emp}$ = 0.126), 13 genes encoding for proteins that are up-regulated in disease ($P_{emp}$ = 0.044) and 16 genes are genetically associated with AD ($P_{emp}$ = 0.059; **Supplementary Figure 18**). TYROBP (Dap12) is an adaptor molecule involved in the transduction pathway of *TREM2* and *CD33*, both of which are known AD risk genes (*66*, *67*), as well as CR3 (*ITGAM* and *ITGB2*) (*68*). Both *TREM2* and *ITGB2* are up-regulated in disease and are contained in the network shown in **Supplementary Figure 16B**.

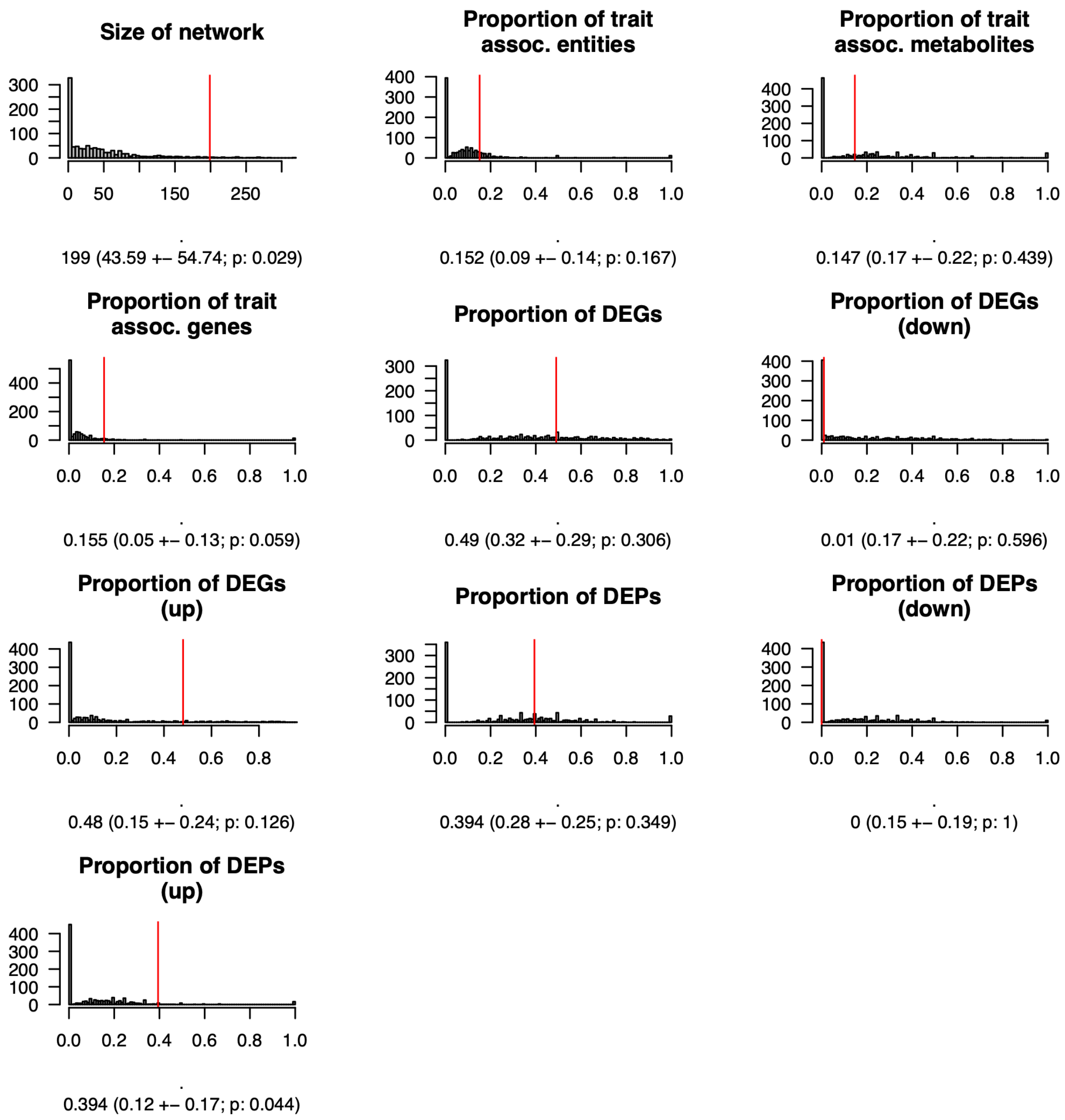

**Figure S18. Background distribution for statins example (gene *ITGAL*).** Network properties of 1000 random networks built by selecting one gene and expanding to their 1-step co-expression neighborhood. Red line indicates observed property for the gene-centric molecular network surrounding the gene *ITGAL* (seen in **Supplementary Figure 16B**). Observed value with mean, standard deviation and empirical *p*-value are provided below each histogram.

In summary, the identified functional link to TYROBP signaling suggests that the subset of statins targeting *ITGAL* may offer protective effects (*69*, *70*) by modulating neuroinflammatory pathways (*68*, *71*), providing an alternative hypothesis on the controversial effects of statins on AD risk. In light of discouraging randomized controlled trials of statins in AD dementia patients (*72*, *73*), these findings demonstrate how the AD Atlas can contribute to a more nuanced understanding of the diverse metabolic profiles associated with different statins, which may allow better selection of specific subpopulations to study in future clinical trials. Interestingly, a recent meta-analysis of electronic health records revealed a statistically significant protective treatment effect for simvastatin (*74*), one of the *ITGAL*-targeting statins, supporting this hypothesis.

##### 4.3 Showcase 3: Contextualization of links between the sphingomyelin pathway and AD pathology

In a previous study, we identified sphingomyelin species (SMs) of differing lengths to be implicated in early vs. late stages of AD (*37*). More precisely, we found SM C16:0 to be associated with CSF $A\beta_{1-42}$ pathology, while SMs with longer fatty acid chains (≥C20) were correlated with brain atrophy and cognitive decline. This study identified three SMs associated with AD, labeled as SM (OH) C14:1, SM C16:0, SM C20:2. We generated a metabolite-centric multi-omics network via the AD Atlas web interface to contextualize these findings and gain a better understanding of their potential functional role in AD. The resulting network can be seen in **Supplementary Figure 19A**.

To characterize the potential functional involvement of SM-associated genes contained in the network in AD, we performed an enrichment analysis using the *Reactome 2016* library via EnrichR. This identifies three genes (*CERS4, SPTLC3 and SGPP1)* involved in SM *de novo* biosynthesis (EnrichR, *Reactome*, $P_{FDR}$ = 2.53e-04; **Supplementary Figure 19B**). Interestingly, these genes have previously been identified in a study that involved multiple, time-intensive manual mapping steps (*75*). Here, the genes were categorized into two functional categories: global sphingomyelin synthesis (*SPTLC3*, *CERS4*) and synthesis and degradation of sphingosine-1-phosphate (*SGPP1*), highlighting a possible role for sphingosine-1-phosphate and its receptors in AD pathogenesis. AD mouse models indicate a potential benefit of Fingolimod, an FDA-approved S1P analog used for the treatment of multiple sclerosis (*76*–*78*). Furthermore, long-term Fingolimod treatment in multiple sclerosis patients has shown positive effects on cognition (*79*). Therefore, Baloni et al. applied a drug repositioning approach by treating APP/PS1 mice with Fingolimod, finding that prolonged S1P pathway modulation can rescue both the proposed cellular mechanism of hippocampus-related memory and cognitive deficits in these mice, further supporting this pathway as a high priority target for AD (*75*).

In conclusion, this analysis highlights the ability of molecular networks to contextualize hypotheses or findings from previous studies and to thereby point to novel, AD-related insights without the need for bioinformatics resources or time-intensive manual analyses. Here, we used three sphingomyelin species that have previously been associated with AD endophenotypes (*37*), to further investigate their involvement in mechanisms of disease. We find a link to the sphingosine-1-phosphate pathway, replicating results of a recent study (*78*).

**A - Molecular subnetwork surrounding SM (OH) C14:1, SM C16:0, SM C20:2**

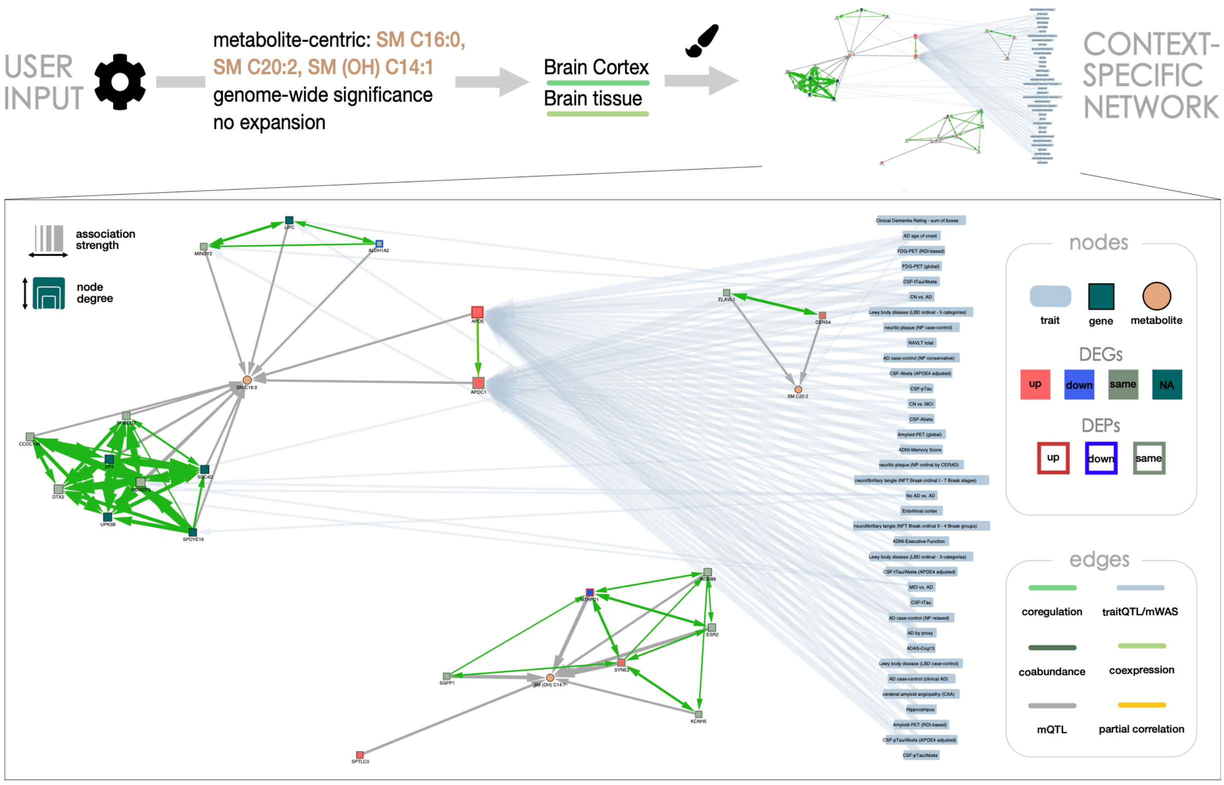

**B - Top 5 gene set enrichment results**

| *Reactome 2016 (EnrichR)* | *overlap** | *q-value*** | *OR**** | *genes* |
|---|---|---|---|---|
| **Sphingolipid de novo biosynthesis** Homo sapiens R-HSA-1660661 | 3/33 | 2.53e-04 | 104.99 | CERS4, SPTLC3, SGPP1 |
| **Sphingolipid metabolism** Homo sapiens R-HSA-428157 | 3/74 | 1.46e-03 | 44.27 | CERS4, SPTLC3, SGPP1 |
| Chylomicron-mediated lipid transport Homo sapiens R-HSA-174800 | 2/17 | 2.13e-03 | 133.09 | LIPC, APOE |
| Lipoprotein metabolism Homo sapiens R-HSA-174824 | 2/34 | 5.20e-03 | 62.33 | LIPC, APOE |
| Metabolism of lipids and lipoproteins Homo sapiens R-HSA-556833 | 5/659 | 5.20e-03 | 8.69 | CERS4, LIPC, SPTLC3, APOE, SGPP1 |

*overlap network (nominator) with pathway gene set (denominator) **adjusted *p*-value using Benjamini-Hochberg ***odds-ratio

**Figure S19. Contextualization of metabolomics-guided insights points to SM *de novo* biosynthesis. (A)** Multi-omics network surrounding three sphingomyelin (SM) species (SM (OH) C14:1, SM C16:0, SM C20:2) that are altered in biomarker defined stages of AD (*2*). **(B)** Gene set enrichment analysis using the *Reactome* gene set accessible via EnrichR identifies three genes (*CERS4, SPTLC3 and SGPP1)* involved in SM de novo biosynthesis. Genes that overlap between pathway sets (column genes) and network are color coded to indicate direction of expression change or protein abundance in disease (red: up-regulation, blue: down-regulation).

#### 4.4 Showcase 4: Networks surrounding marker genes for homeostatic microglia and disease-associated microglia suggest possible involvement of blood androgens

Genomic analyses in AD and animal models of the disease identified a specific activation program which drives the transition from homeostatic microglia to disease-associated microglia (*80*). However, the molecular mechanisms underlying this transition remain incompletely understood and the extent to which this process involves AD susceptibility genes has not been assessed in an integrated fashion. We used the AD Atlas to identify gene modules defining homeostatic vs. disease-associated microglia and to evaluate their respective links with large-scale genetics, proteomics and metabolomics data. Two canonical gene markers of homeostatic vs disease-associated microglia, namely *TMEM119* (*81*, *82*) for homeostatic microglia and *TREM2* (*83*) for disease-associated microglia, were entered as single query genes to construct brain co-expression-based molecular networks. An overview of the analysis steps can be seen in **Supplementary Figure 20A**.

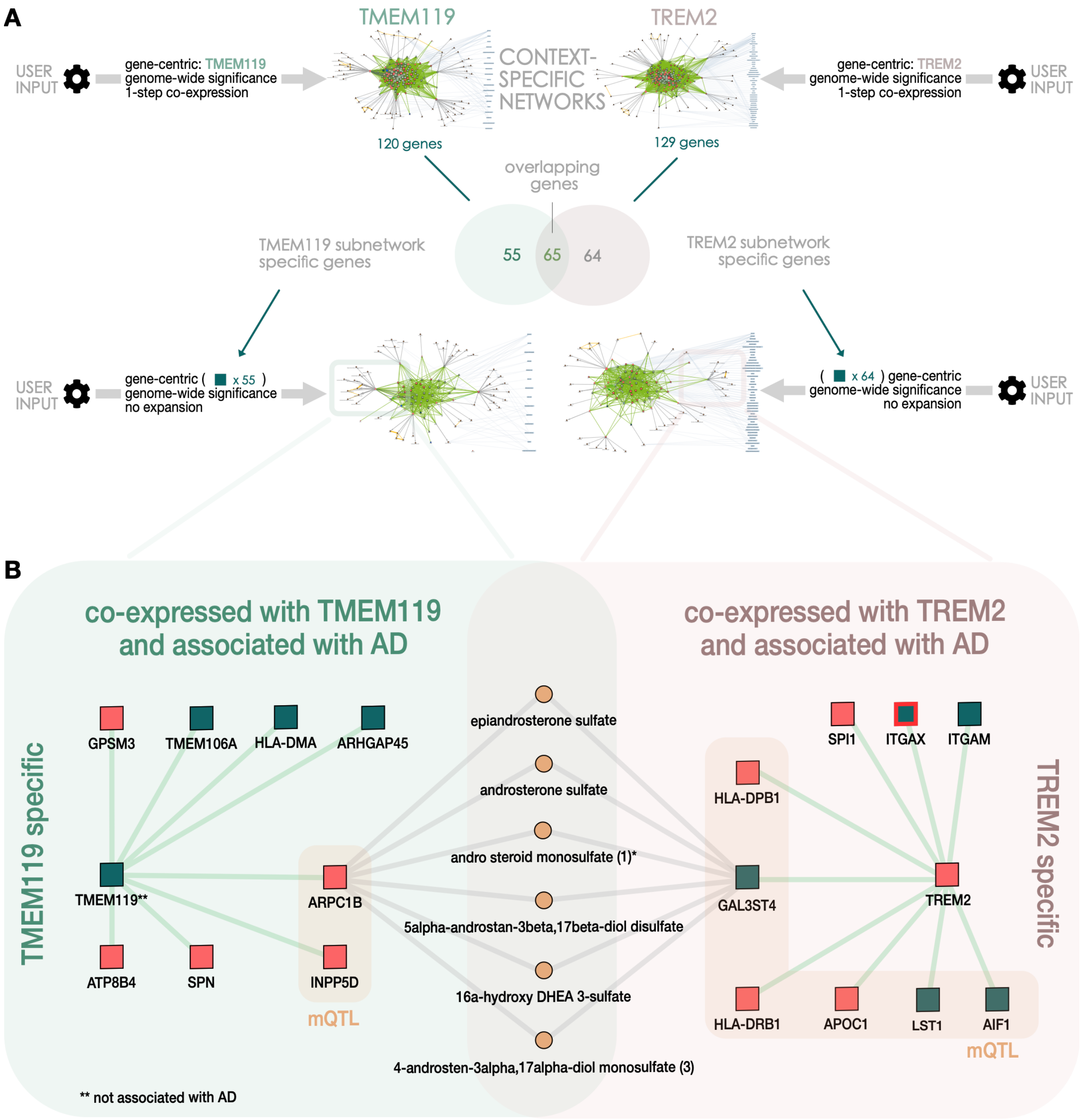

**Figure S20. Investigating the transition between homeostatic microglia and disease-associated microglia (DAM). (A)** Overview of the analysis. First, gene co-expression networks were built around marker genes for homeostatic (*TMEM119*) and DAM (*TREM2*). Then gene modules containing genes co-expressed exclusively with one of these genes were constructed and analyzed. **(B)** Schematic representation of the marker-specific networks. Genes that show co-expression in the brain with *TMEM119* but not *TREM2* and that are genetically associated with AD phenotypes are shown on the left (green background). Those that are additionally associated with metabolite levels are highlighted (orange). Genes that are significantly up-regulated at the mRNA (node filling) or protein (node border) level are highlighted in red. Genes that show co-expression in the brain with *TREM2* but not *TMEM119* and are genetically associated with AD phenotypes are shown on the right (red background). Those that are additionally associated with metabolite levels are highlighted (orange). Androgen steroids associated with both *ARPC1B* and *GAL3ST4* are depicted to highlight this overlap. It is important to note that only selected relationships are shown. For a detailed version of these networks please refer to our website (www.adatlas.org/?showcases).

Of the 55 genes co-expressed with *TMEM119* but not *TREM2*, eight are genetically associated with AD phenotypes (*ARHGAP45, ARPC1B, ATP8B4, GPSM3, HLA-DMA, INPP5D, SPN, TMEM106A*; **Supplementary Table 9**) and two of these (*ARPC1B, INPP5D*) are also associated with metabolite levels at genome-wide significance (mQTL associations), including multiple androgenic steroids (**Supplementary Table 10**). Focusing on this overlap of genetic associations with AD and levels of metabolites from the androgen pathway in the *ARPC1B* locus revealed interesting connections. Androgens are a class of steroid sex hormones that are responsible for the development of male sex characteristics (*84*) and also play important roles in female reproductive function (*85*). Females have a higher susceptibility to AD (*86*) and studies have linked age-related depletion of the androgen testosterone to an increased risk of AD in men (*87*, *88*). Interestingly, *in vitro* and *in vivo* models of neuroinflammation report that androgens may exert anti-inflammatory and neuroprotective effects by inhibiting microglial activation (*89*, *90*). Since *ARPC1B* has been linked to the branching and motility of microglial ramifications (*91*), this might suggest a potential molecular relationship between androgen levels, ageing and the ability of microglia to extend ramifications in the context of AD.

Of the 64 genes co-expressed with *TREM2* but not *TMEM119*, nine genes map to AD-associated loci (*AIF1, APOC1, GAL3ST4, HLA-DPB1, HLA-DRB1, ITGAM, ITGAX, LST1, SPI1*; **Supplementary Table 11**) and six of these (*AIF1, APOC1, GAL3ST4, HLA-DPB1, HLA-DRB1, LST1*) are also associated with the levels of at least one metabolite, again including multiple androgenic steroids (**Supplementary Table 12**). As observed in the *TMEM119* module, one gene, namely *GAL3ST4*, is associated with androgen metabolites. Although little is known about the role of *GAL3ST4* in microglia, the gene was reported to be part of a *TYROBP* brain-expressed gene module crucially involved in the development of LOAD (*64*). Interestingly, a metabolite-centric search for the direct network surrounding the androgen steroids associated with both *ARPC1B* and *GAL3ST4* (overlap of metabolites is seen in **Supplementary Figure 20B**) revealed an association of these metabolites with another AD-specific risk locus, *ZCWPW1/NYAP1/PILRA* (7q22.1) (*92*), which has been linked to myeloid enhancer activity, microglia function and neuroinflammation (*93*, *94*).

Lastly, to see if these networks can again point to drug compounds targeting the activation of microglia, we repeated the gene set enrichment analysis described previously using the 1-step co-expression network surrounding both *TMEM119* and *TREM2*. This identified Fasudil (EnrichR, *Drug Perturbations from GEO down*, $P_{\mathrm{FDR}}$ = 7.02e-16) among the most significant hits. Fasudil is an inhibitor of Rho-kinase (ROCK) and approved for the treatment of cerebral vasospasm in Japan. *Post mort*em data suggests that ROCK protein levels are elevated in AD brains (*95*) and preclinical data from *in vitro* and *in vivo* studies, including animal models of AD, indicate that Fasudil may be able to reduce the burden of tau protein (*96*) and promote an anti-inflammatory microglial phenotype (*97*, *98*). Fasudil has shown promising results in pre-clinical studies (*99*, *100*) and two ongoing clinical phase II trials are currently investigating the use of Fasudil in tauopathies (NCT04734379) and the effectiveness of an oral formulation of Fasudil in patients with dementia (NCT04793659). Interestingly, a previous *in vitro* study reported experimental evidence linking androgen levels through androgen receptor signaling to the levels of miRNA-135a, which targets ROCK, providing a potential mechanistic model integrating the different omics entities contained in the AD Atlas-derived network (*101*).

Overall, our analyses point to a potential involvement of blood androgens in the transition from homeostatic to disease-associated microglia during the course of AD. Using gene set enrichment, we were able to identify Fasudil, an inhibitor of ROCK, as a promising drug repositioning candidate. We  hypothesize that age-related decreases of androgen levels may result in an upregulation of ROCK via androgen-mediated pathways, leading to microglial activation and neuroinflammation (*102*, *103*). This exploratory analysis highlights how multi-omics networks can inform testable hypothesis, such as the potential link between androgen signaling, ROCK activity and microglial activation in AD, that can be investigated in follow-up experiments.

#### 5 DOCUMENTATION AND FEEDBACK

The Alzheimer's Disease Atlas is a new resource that was developed to enable the collection, consolidation and exploration of Alzheimer's disease related multi-omics data. It enables easy and efficient access to vast amounts of data generated across different studies as well as providing a continuously expanding toolbox of downstream analysis methods. Our web interface (*adatlas.org*) features multiple interactive showcases and provides users with additional documentation and help.

The resource is aimed at the Alzheimer's disease research community. Therefore, we are very thankful for feedback from our users regarding usability, missing features or data integration requests.

Please send bug reports, improvement ideas and feedback to **.

#### FIGURES AND TABLES

##### Box S1. Glossary of important terms and concepts.

***Step-wise multi-omics integration*** – Pair-wise association results are overlayed and integrated to interconnect entities within and between omics layers**,** enabling integration across different cohorts and studies.

***AD-related trait –*** traits that are tested for associations in genome- or metabolome-wide association studies (GWAS and MWAS), including cognitive measures as well as cerebrospinal fluid (CSF) and imaging biomarkers of Alzheimer's disease (AD). The term is used interchangeably with AD (endo-)phenotypes throughout this manuscript.

***Metabolic trait –*** measured concentrations of a metabolite which are tested for association with AD in metabolome-wide association studies (MWAS).

***SNP-to-gene mapping -*** For the assignment of SNPs to genes, we use the genomic location of a SNP (either directly within the gene body of a gene or within 2.5kb up- or downstream), gene-associated regulatory elements (promoters or enhancers from ENCODE and FANTOM5), and eQTL and pQTL associations.

***Context-specific molecular subnetwork –*** Multi-omics network built from pairwise association data surrounding entities of interest; associations (edges) are optionally filtered for tissue or sample type.

> ***Annotation –*** Addition of statistical results (nodes and edges) to an entity or set of entities to build a context-specific molecular network. For example, annotating metabolites through mQTL associations with metabolic genes.
>
> ***Significance threshold –*** Cutoff for statistical significance. Associations between two entities that are below this threshold are depicted as edges in the network.
>
> ***Edge filtering –*** To create context- and tissue-specific networks, users can filter associations (edges) by tissue or sample type. This is currently possible for co-expression, co-regulation, genetic associations and metabolic associations.

***Edge types –*** categorize statistically significant associations/correlations between two entities, represented as an edge in the multi-omics network. In the context of this manuscript, they are defined as following:

> ***Co-regulation –*** inferred from expression quantitative trait loci (eQTL studies) to inform about shared genetic and epigenetic regulation of gene or protein expression. Edges link two or more genes mostly within one locus, but can also be in trans.
>
> Interpretation examples of an edge (A)-[:COREGULATION]-(B) include:
>
> > *A SNP is an eQTL for both gene A and gene B;*
> > *A SNP is located in gene A and is an eQTL for gene B;*
> > *A SNP is located in a promoter/enhancer linked to gene A and is an eQTL for gene B;*
>
> ***Genetic association –*** inferred from GWAS and MWAS to inform about genetic risk for AD and metabolite quantitative trait loci (mQTL). Edges link genes with AD-related traits or metabolic traits.
>
> Interpretation examples of an edge (A)-[:GENETIC ASSOCIATION]-(B) include:
>
> > *A SNP is located in gene A and is a QTL for (metabolic) trait B;*
> > *A SNP is an eQTL for gene A and is a QTL for (metabolic) trait B;*
> > *A SNP is located in a promoter/enhancer linked to gene A and QTL for (metabolic) trait B;*
>
> ***Metabolic association –*** inferred from metabolome-wide association studies (MWAS). Edges link metabolites with AD-related traits and indicate a significant association.
>
> ***Co-expression –*** inferred from gene co-expression analysis. Edges indicates a statistically significant positive or negative correlation between the levels of two transcripts.
>
> ***Co-abundance –*** inferred from gaussian graphical models (GGMs) using proteomics data. Edges link two or more genes and indicate a significant positive or negative partial correlation between the abundance of proteins that are encoded by the genes***.***
>
> ***Partial correlation –*** inferred from GGMs using metabolomics data. Edge indicates a significant positive or negative partial correlation between the levels of two metabolites***.***

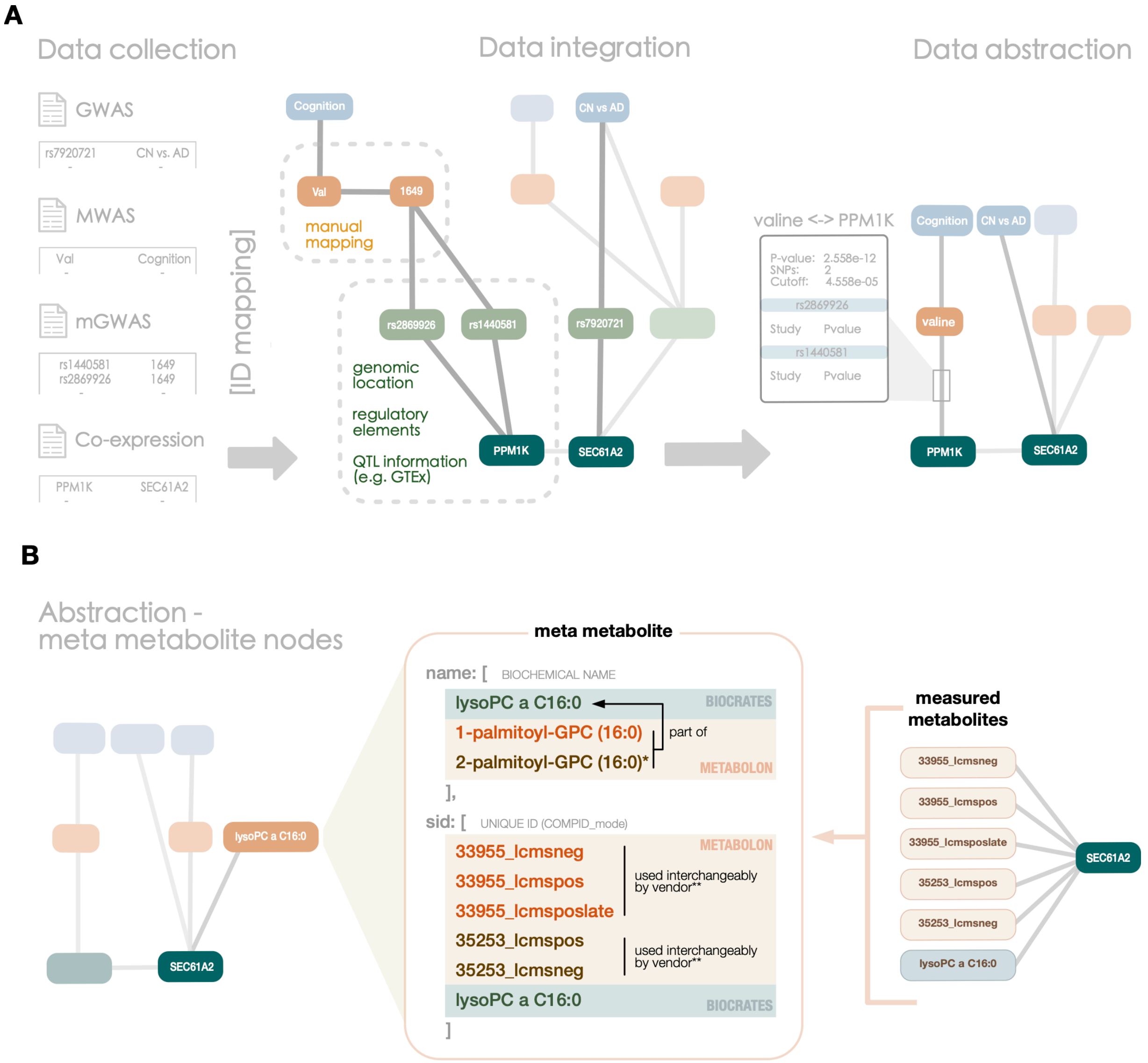

**Figure S1. Integration and abstraction pipeline.** A schematic example of the integration and abstraction pipeline. **(A)** First, statistical results are collected from different analyses and studies ("Data collection") and subsequently stored in comprehensive network structures that consist of different node types (trait, gene, SNP, metabolite) which are connected by relationships inferred from the collected datasets ("Data integration"). Lastly, the detailed data model is simplified and abstracted by mapping of entities across platforms (in case of metabolites) and mapping gene-linked node types (SNPs, transcripts, proteins) to their genes and by subsequent QTL-based integration to establish direct links between omics layers ("Data abstraction"). **(B)** Detailed example of a meta-metabolite. The meta-metabolite lysoPC a C16:0 is a consolidation of 6 different metabolite measurements (3 unique measurements, up to three MS modes) that have been measured on two different metabolomics platforms (Metabolon and Biocrates). As these two vendors provide different levels of resolution, we map higher resolved metabolites to the lowest given level through manual cross-platform mapping. Here, for example, 1-palmitoyl-GPC (16:0) and 2-palmitoyl-GPC (16:0)* are both constituents of lysoPC a C16:0, which is the sum of the two. Metabolites measured by Metabolon with the same compound ID but differing measurement modes are also summarized, as the vendor measures some compounds across several MS modes and reports the measurement from the platform/mode showing the best performance for the respective compound (determined on a per-study basis; thus, assumed as being interchangeable**).

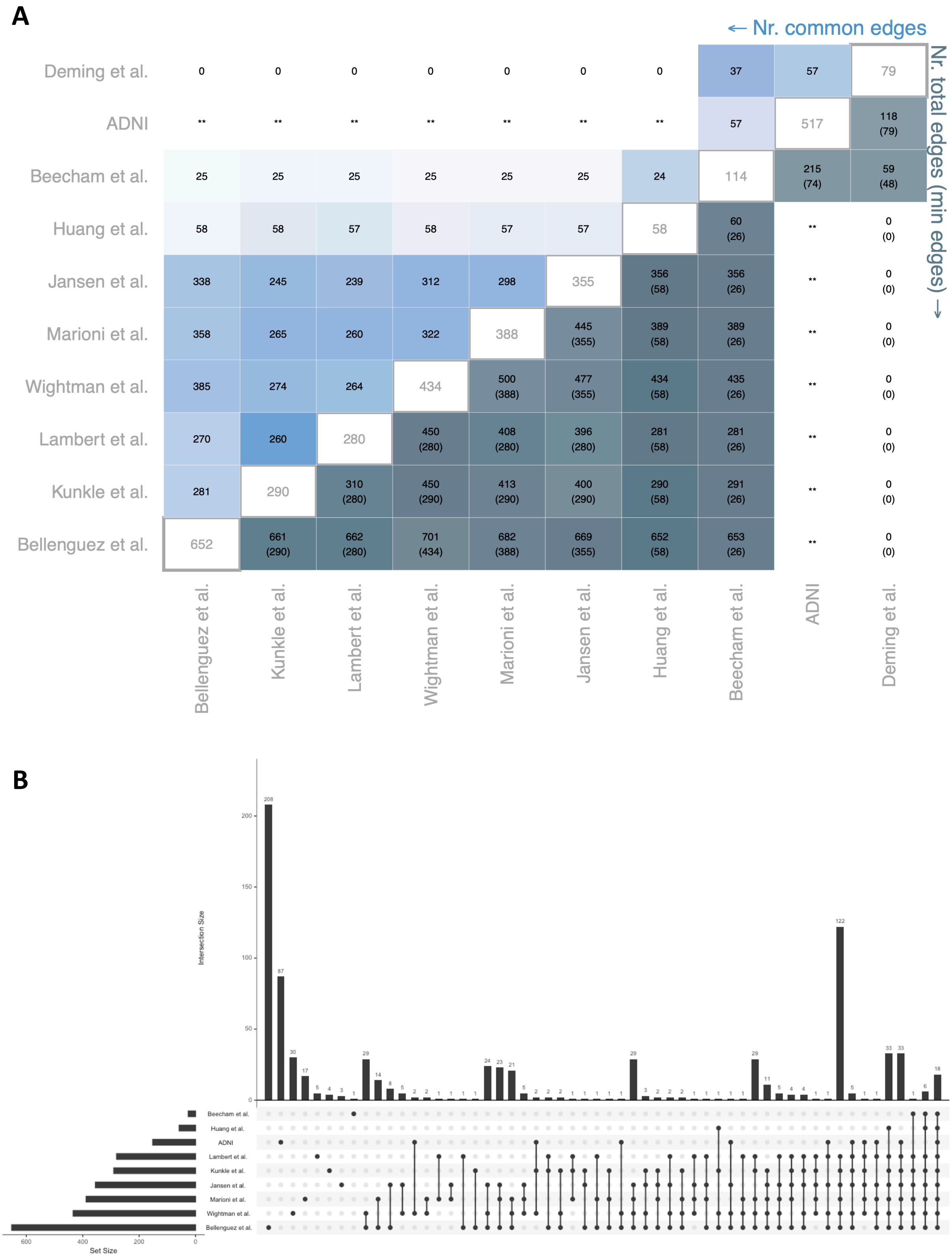

**Figure S2. Statistics used to calculate the Jaccard index and Overlap coefficient for Figure 2. (A)** Pairwise network statistics used to calculate the Jaccard index and Overlap coefficient. Diagonal: total number of edges in unfiltered network. Upper triangle: Number of common edges. Lower triangle: Number of total edges (union of both networks) regarded after filtering in the comparison and the number of edges in the smaller network. **(B)** Upset plot showing the meta-trait-gene overlap between studies.

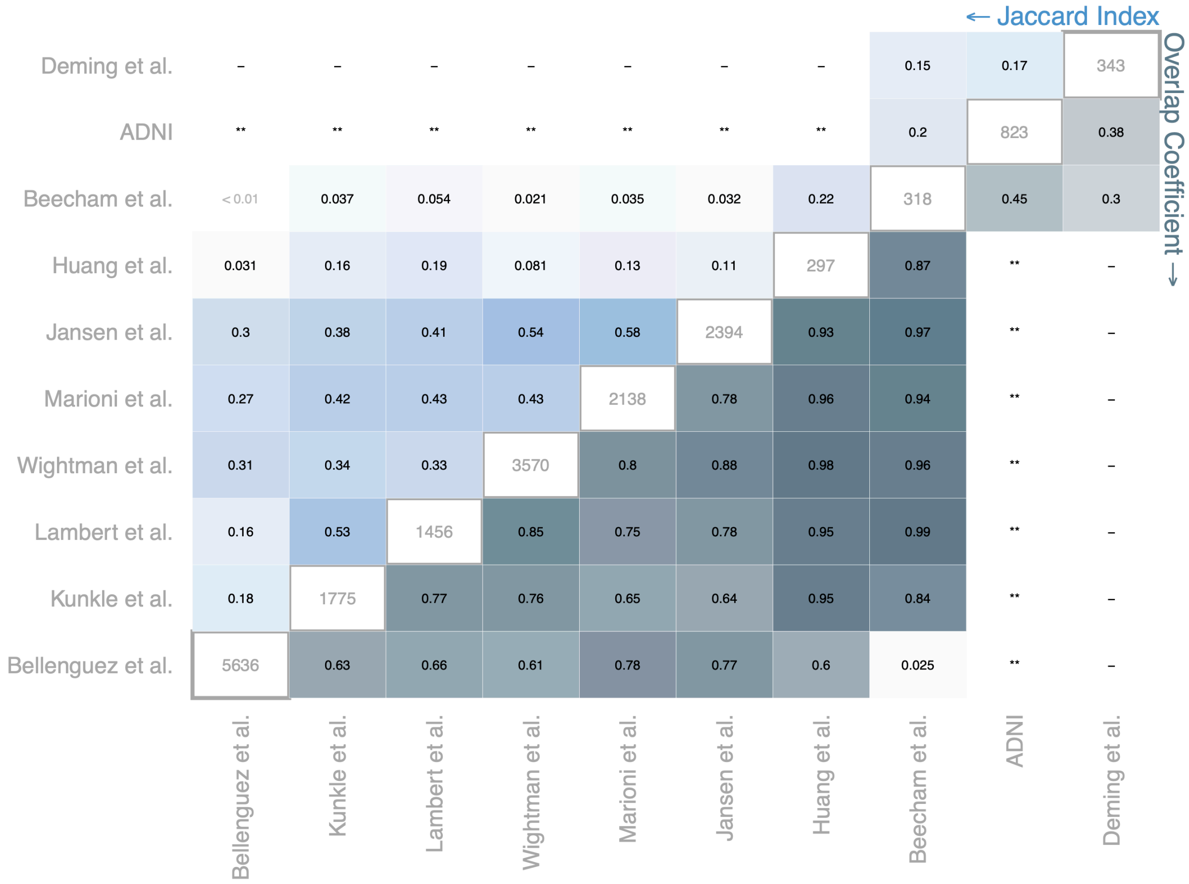

**Figure S3. Meta-trait-SNP (GWAS) network comparison.** Heatmap of genetic correlation between AD-relevant genome-wide association studies (GWAS). Pairwise comparisons were performed at the common meta-trait- and SNP-level. Diagonal: total number of edges in unfiltered network. Upper triangle: Jaccard index (JI). Lower triangle: Overlap coefficient (OC). Cells are colored by value; zero (white) to one (blue) and ordered in correspondence to **Figure 2A** for comparability. Statistics used to calculate the JI and OC are given in **Supplementary Figure 4.** Low overlap between Bellenguez et al. and Beecham et al. is most likely due to systematic difference in APOE reporting in Bellenguez et al. and lower statistical power in Beecham et al. **ADNI included in meta-analyses as part of IGAP

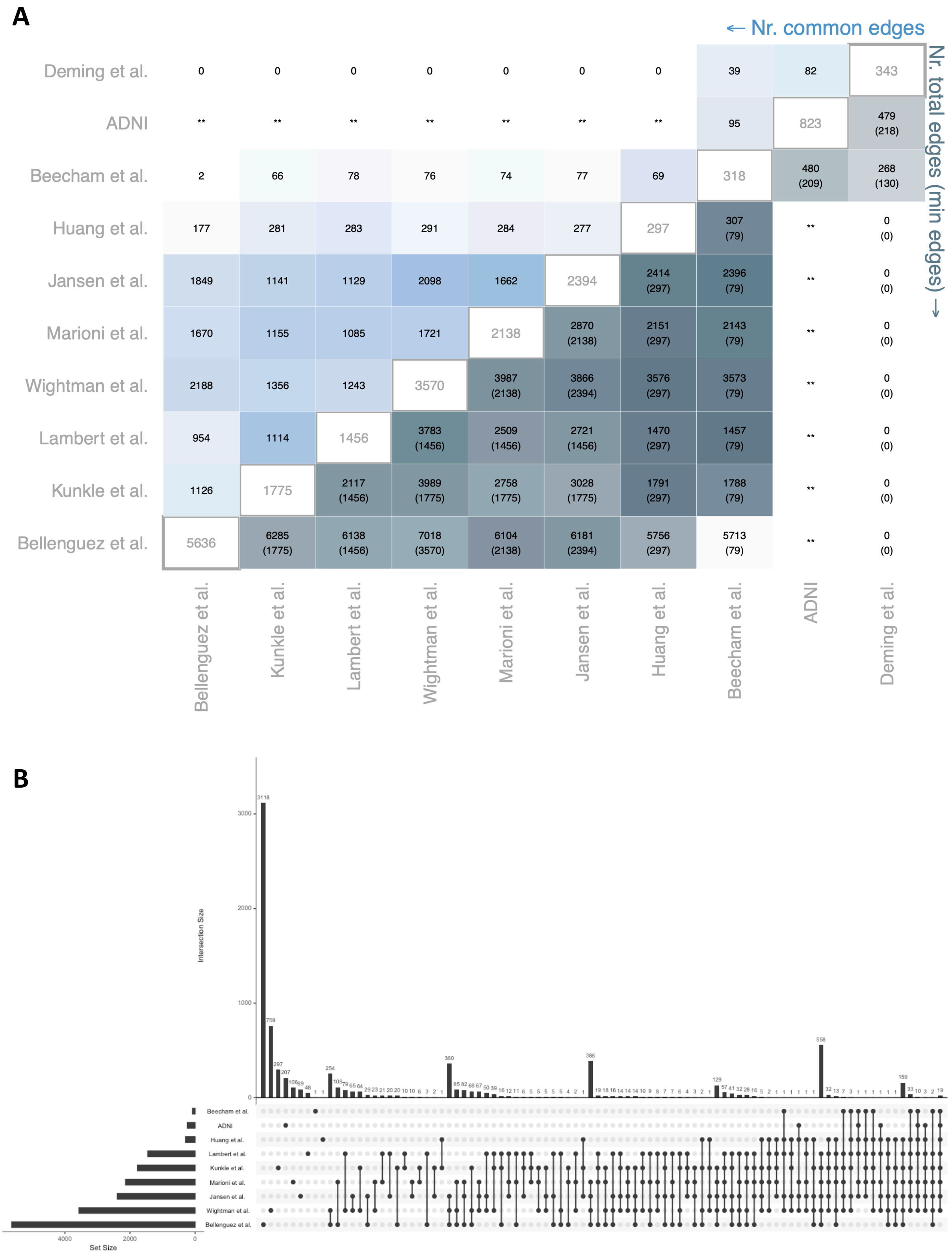

**Figure S4. Statistics used to calculate the Jaccard index and Overlap coefficient for Supplementary Figure 3. (A)** Pairwise network statistics used to calculate the Jaccard index and Overlap coefficient. Diagonal: total number of edges in unfiltered network. Upper triangle: Number of common edges. Lower triangle: Number of total edges (union of both networks) regarded after filtering in the comparison and the number of edges in the smaller network. **ADNI included in meta-analyses as part of IGAP **(B)** Upset plot showing the meta-trait-SNP overlap between studies.

← Nr. common edges

Nr. total edges (min edges) →

| | COREG (cortex, Sieberts) | COREG (blood) | COREG (CBE) | COREG (cortex, GTEx) | COREG (BA9) | COABUN (brain) | COABUN (plasma) | COEXPR (FP) | COEXPR (PHG) | COEXPR (STG) | COEXPR (IFG) | COEXPR (CBE) | COEXPR (TCX) | COEXPR (DLPFC) |
|---|---|---|---|---|---|---|---|---|---|---|---|---|---|---|
| COEXPR (DLPFC) | 565 | 436 | 336 | 307 | 248 | 902 | 12 | 5872 | 5069 | 5865 | 5167 | 4873 | 8624 | 71768 |
| COEXPR (TCX) | 567 | 374 | 311 | 287 | 217 | 659 | 8 | 4321 | 4221 | 4705 | 4150 | 5229 | 48297 | 98971 (41104) |
| COEXPR (CBE) | 657 | 449 | 405 | 347 | 289 | 467 | 8 | 2627 | 2429 | 2882 | 2252 | 47534 | 76590 (40898) | 93586 (37629) |
| COEXPR (IFG) | 266 | 207 | 149 | 137 | 103 | 521 | 9 | 5871 | 5670 | 6332 | 28864 | 48984 (24270) | 56332 (27089) | 72397 (25905) |
| COEXPR (STG) | 336 | 252 | 183 | 174 | 139 | 672 | 6 | 6881 | 6519 | 33511 | 50707 (26519) | 53329 (27722) | 60770 (31310) | 77815 (30057) |
| COEXPR (PHG) | 314 | 233 | 163 | 151 | 121 | 566 | 5 | 6229 | 31455 | 53347 (28999) | 48491 (26048) | 51082 (26361) | 58823 (29256) | 75419 (28085) |
| COEXPR (FP) | 340 | 255 | 180 | 160 | 128 | 629 | 8 | 35196 | 54225 (29347) | 56092 (31458) | 50633 (26569) | 56988 (29667) | 63381 (32448) | 82019 (31710) |
| COABUN (plasma) | 15 | 16 | 10 | 6 | 5 | 59 | 4203 | 1493 (367) | 1322 (318) | 1485 (420) | 1329 (340) | 1977 (422) | 1908 (453) | 1999 (594) |
| COABUN (brain) | 606 | 428 | 343 | 265 | 214 | 69323 | 1802 (834) | 51292 (13909) | 43320 (12405) | 44523 (12678) | 40618 (11026) | 64611 (15082) | 67200 (16420) | 78733 (27947) |
| COREG (BA9) | 42493 | 35035 | 35806 | 37909 | 53237 | 54715 (10232) | 2646 (406) | 35600 (14415) | 30769 (12103) | 31882 (12341) | 28443 (11545) | 54737 (26000) | 54723 (24983) | 67403 (23599) |
| COREG (cortex, GTEx) | 54820 | 44114 | 44251 | 70964 | 67183 (46702) | 64829 (13445) | 3162 (498) | 44340 (19405) | 38206 (16197) | 39590 (16575) | 35708 (15568) | 68237 (34064) | 67726 (32843) | 82720 (31069) |
| COREG (CBE) | 67935 | 54239 | 95397 | 93558 (60876) | 84589 (46843) | 75492 (17964) | 3680 (602) | 54204 (25888) | 46613 (21561) | 48465 (22082) | 43846 (20878) | 82676 (38312) | 83024 (39746) | 99032 (41032) |
| COREG (blood) | 87228 | 144410 | 136138 (79948) | 123104 (61401) | 112552 (47214) | 88268 (25882) | 4447 (959) | 68255 (31585) | 57980 (27911) | 60729 (29636) | 54917 (25580) | 104111 (41919) | 103550 (43238) | 121645 (57992) |
| COREG (cortex, Sieberts) | 233676 | 195923 (112569) | 175605 (78865) | 164144 (60708) | 151821 (46565) | 101810 (38267) | 5249 (1310) | 91352 (32668) | 77932 (29021) | 81058 (30958) | 74315 (26685) | 142689 (44284) | 141128 (44813) | 157454 (66620) |

**Figure S5. Gene-gene network comparison.** Pairwise network statistics used to calculate the Jaccard index and Overlap coefficient. Diagonal: total number of edges in unfiltered network. Upper triangle: Number of common edges. Lower triangle: Number of total edges (union of both networks) regarded after filtering in the comparison and the number of edges in the smaller network.

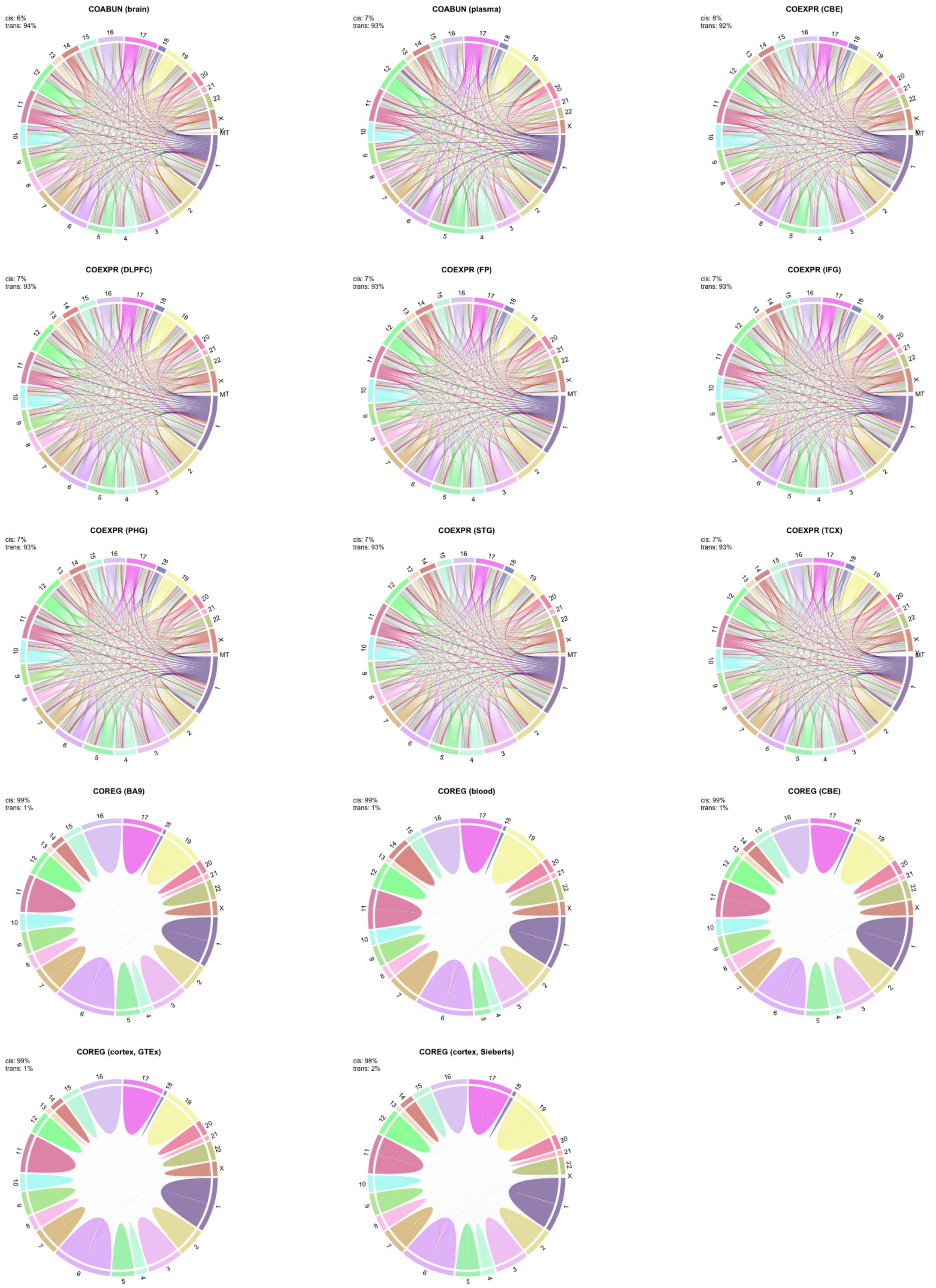

**Figure S6. Genetic architecture of gene-gene links.** Visualization of cis and trans links for selected edge types connecting genes with genes. *Cis* is defined as an edge connecting genes which are located on the *same* chromosome and *trans* is defined as an edge connecting genes located on *different* chromosomes.

| Omics-layer | n | Enrichment |
|---|---|---|
| SNP | 821 | Endolysosome (p:3.73e-06; OR:1.66)<br>Tau Homeostasis (p:1.02e-04; OR:4.94)<br>Lipid Metabolism (p:1.68e-04; OR:1.41)<br>Oxidative Stress (p:3.16e-03; OR:1.66)<br>Immune Response (p:3.17e-03; OR:1.29)<br>Autophagy (p:3.92e-03; OR:1.49)<br>APP Metabolism (p:6.07e-03; OR:1.88) |
| DEG (brain) | 3479 | Mitochondrial Metabolism (p:1.13e-09; OR:1.44)<br>Vasculature (p:5.22e-08; OR:1.47)<br>Proteostasis (p:2.46e-05; OR:1.19)<br>Tau Homeostasis (p:1.26e-02; OR:2.00)<br>Myelination (p:1.32e-02; OR:1.40)<br>Oxidative Stress (p:1.36e-02; OR:1.26)<br>Synapse (p:1.24e-02; OR:1.15)<br>Structural Stabilization (p:1.50e-02; OR:1.10) |
| DEG (DLPFC) | 1646 | Mitochondrial Metabolism (p:1.70e-03; OR:1.29) |
| DEP (DLPFC) | 3244 | Synapse (p:1.52e-106; OR:3.20)<br>Mitochondrial Metabolism (p:4.73e-60; OR:2.47)<br>Structural Stabilization (p:2.16e-39; OR:1.75)<br>Myelination (p:5.39e-10; OR:2.27)<br>Vasculature (p:1.35e-08; OR:1.48)<br>Endolysosome (p:3.14e-08; OR:1.38)<br>Tau Homeostasis (p:4.21e-08; OR:4.25)<br>Oxidative Stress (p:6.85e-08; OR:1.65)<br>Proteostasis (p:1.71e-06; OR:1.22)<br>RNA Spliceosome (p:2.92e-06; OR:1.63)<br>APP Metabolism (p:3.70e-05; OR:1.70)<br>Autophagy (p:5.60e-05; OR:1.35)<br>Lipid Metabolism (p:4.58e-04; OR:1.18)<br>Apoptosis (p:4.95e-04; OR:1.21) |

**Figure S7. Functional enrichment of gene sets implicated in AD by different omics layers.** SNP - set of genes that can be genetically linked to AD risk/diagnosis at genome-wide significance (n=821), DEG (brain) - set of genes differentially expressed in AD across brain regions, DEG (DLPFC) - set of genes differentially expressed in AD in the DLPFC, DEP - set of genes which encode proteins that are differentially abundant in AD in the DLPFC. Functional enrichment analysis was performed using AD-related biodomains. The significance of the enrichment was calculated using a one-sided *Fisher's exact test*. P: p-value; OR: odds-ratio.

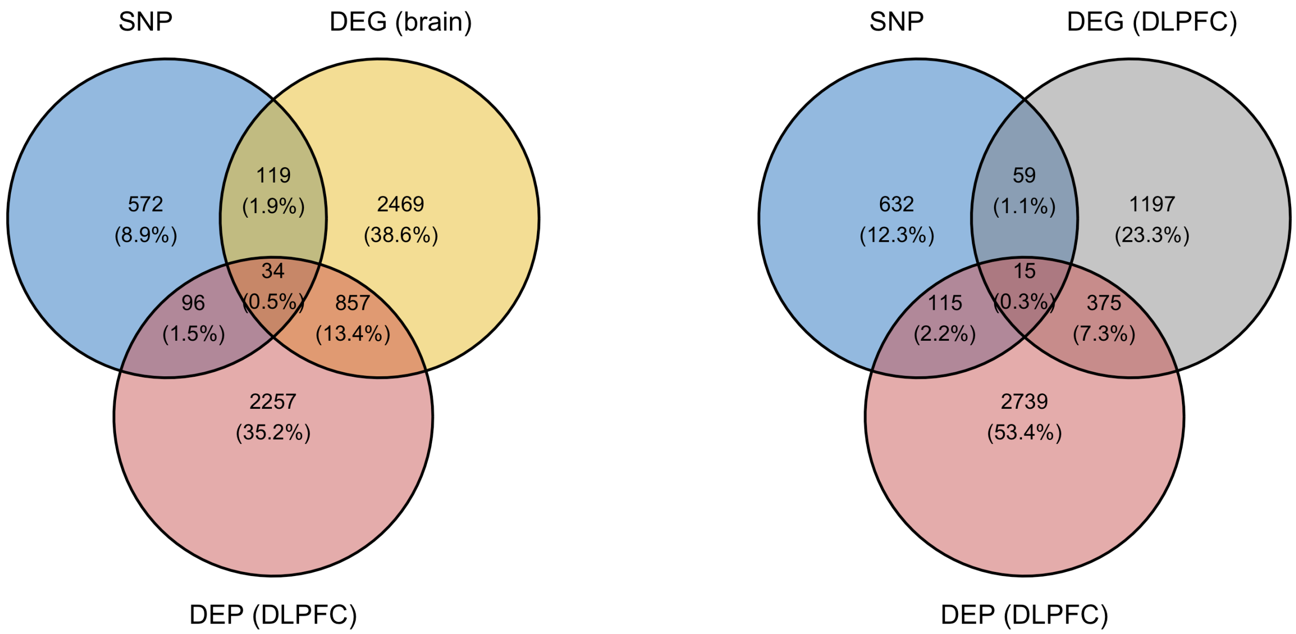

(I)

| Intersection | p-value | OR | 95% CI | Enrichment |
|---|---|---|---|---|
| SNP - DEG | 0.12 | 1.1 | 0.95 | Endolysosome (p:2.38e-04; OR:2.30)<br>Mitochondrial Metabolism (p:4.07e-03; OR:1.94)<br>Tau Homeostasis (p:1.05e-02; OR:7.06)<br>Synapse (p:8.23e-03; OR:1.82) |
| SNP - DEP | 0.55 | 0.99 | 0.84 | Synapse (p:8.12e-08; OR:3.21)<br>Tau Homeostasis (p:2.92e-07; OR:18.61)<br>Mitochondrial Metabolism (p:5.96e-06; OR:2.77)<br>Apoptosis (p:5.27e-06; OR:2.72)<br>Structural Stabilization (p:9.40e-05; OR:2.07)<br>Oxidative Stress (p:2.36e-04; OR:3.32)<br>Autophagy (p:6.31e-03; OR:2.16)<br>APP Metabolism (p:8.91e-03; OR:3.18)<br>Endolysosome (p:1.65e-02; OR:1.74) |
| DEG - DEP | 1.1e-59 | 2.1 | 2 | Mitochondrial Metabolism (p:3.66e-23; OR:2.50)<br>Synapse (p:1.36e-19; OR:2.30)<br>Structural Stabilization (p:3.65e-10; OR:1.60)<br>Vasculature (p:1.05e-05; OR:1.66)<br>Myelination (p:8.79e-05; OR:2.30)<br>Oxidative Stress (p:6.13e-04; OR:1.70)<br>Tau Homeostasis (p:1.03e-02; OR:2.90)<br>Lipid Metabolism (p:1.14e-02; OR:1.22)<br>Apoptosis (p:1.93e-02; OR:1.24) |
| all* | - | - | - | Mitochondrial Metabolism (p:1.95e-04; OR:4.46)<br>Synapse (p:1.36e-03; OR:3.64)<br>Tau Homeostasis (p:6.48e-03; OR:18.12)<br>Structural Stabilization (p:6.85e-03; OR:2.59) |

(II)

| Intersection | p-value | OR | 95% CI | Enrichment |
|---|---|---|---|---|
| SNP - DEG | 0.17 | 1.1 | 0.91 | – |
| SNP - DEP | 0.55 | 0.99 | 0.84 | Synapse (p:8.12e-08; OR:3.21)<br>Tau Homeostasis (p:2.92e-07; OR:18.61)<br>Mitochondrial Metabolism (p:5.96e-06; OR:2.77)<br>Apoptosis (p:5.27e-06; OR:2.72)<br>Structural Stabilization (p:9.40e-05; OR:2.07)<br>Oxidative Stress (p:2.36e-04; OR:3.32)<br>Autophagy (p:6.31e-03; OR:2.16)<br>APP Metabolism (p:8.91e-03; OR:3.18)<br>Endolysosome (p:1.65e-02; OR:1.74) |
| DEG - DEP | 8.8e-18 | 1.7 | 1.6 | Synapse (p:1.53e-08; OR:2.16)<br>Mitochondrial Metabolism (p:6.30e-07; OR:2.01)<br>Myelination (p:1.38e-03; OR:2.59)<br>Structural Stabilization (p:1.30e-03; OR:1.42)<br>Lipid Metabolism (p:8.89e-03; OR:1.35) |
| all* | - | - | - | – |

**Figure S8. Functional enrichment of the intersection of gene sets implicated in AD by different omics layers.** SNP - set of genes that can be genetically linked to AD risk/diagnosis at genome-wide significance (n=821), DEG (brain) - set of genes differentially expressed in AD across brain regions, DEG (DLPFC) - set of genes differentially expressed in AD in the DLPFC, DEP - set of genes which encode proteins that are differentially abundant in AD in the DLPFC. Functional enrichment analysis was performed using AD-related biodomains. The significance of the overlap and the enrichment was calculated using a one-sided *Fisher's exact test*. P: p-value; OR: odds-ratio; CI: confidence interval. *Intersection between all three omics, significance of overlap not tested.

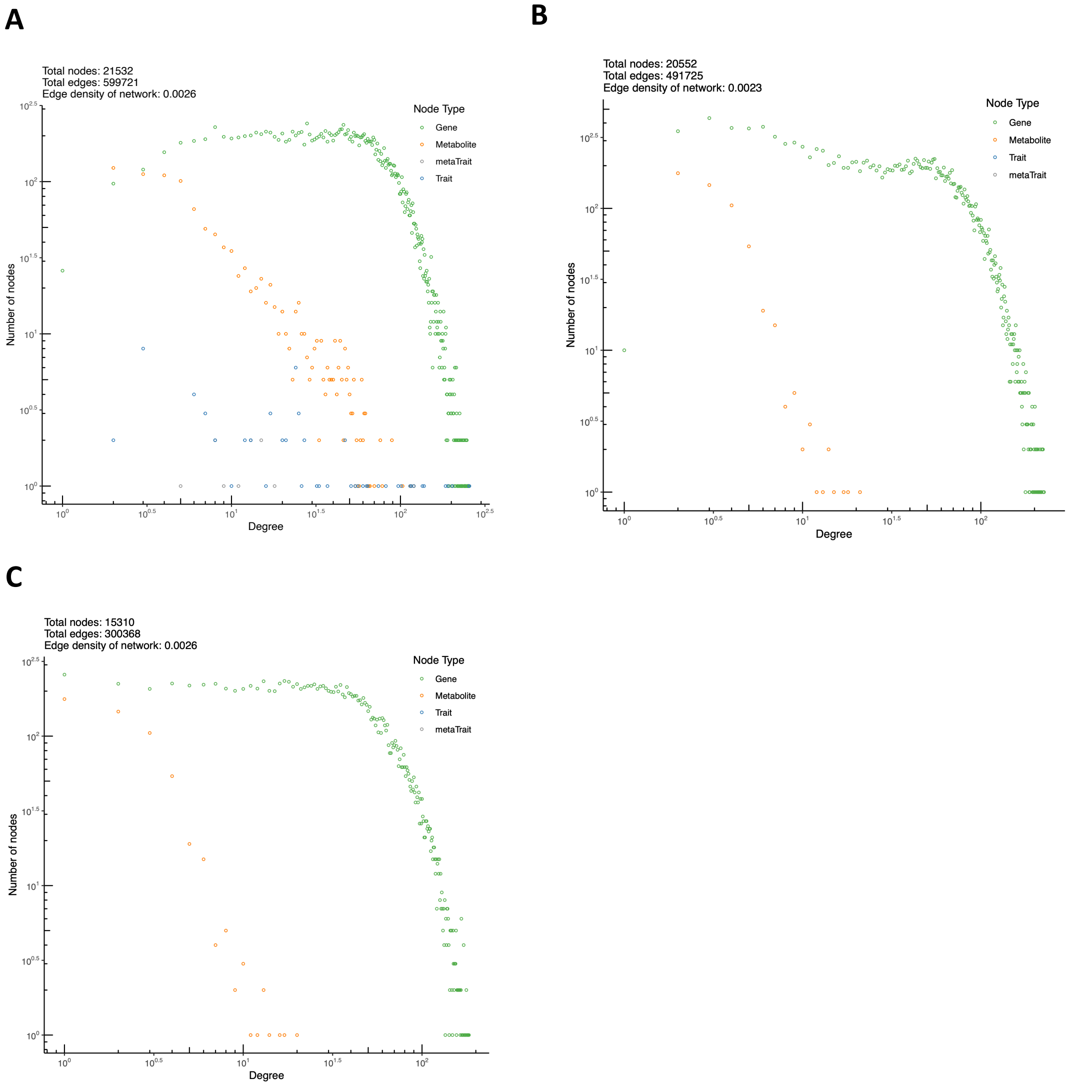

**Figure S9. Degree distribution of the AD Atlas network.** (**A**) Simple network representation (without self-loops and multiple edges between nodes) of the AD Atlas including all edge types, sample types and node types. (**B**) Simple network representation of the AD Atlas filtered to brain-specific edges and trait nodes. (**C**) Simple network representation of the AD Atlas filtered to brain-specific edges, and excluding co-regulation edges and trait nodes as used for the global assessment of biological content and visualization of the global network structure.

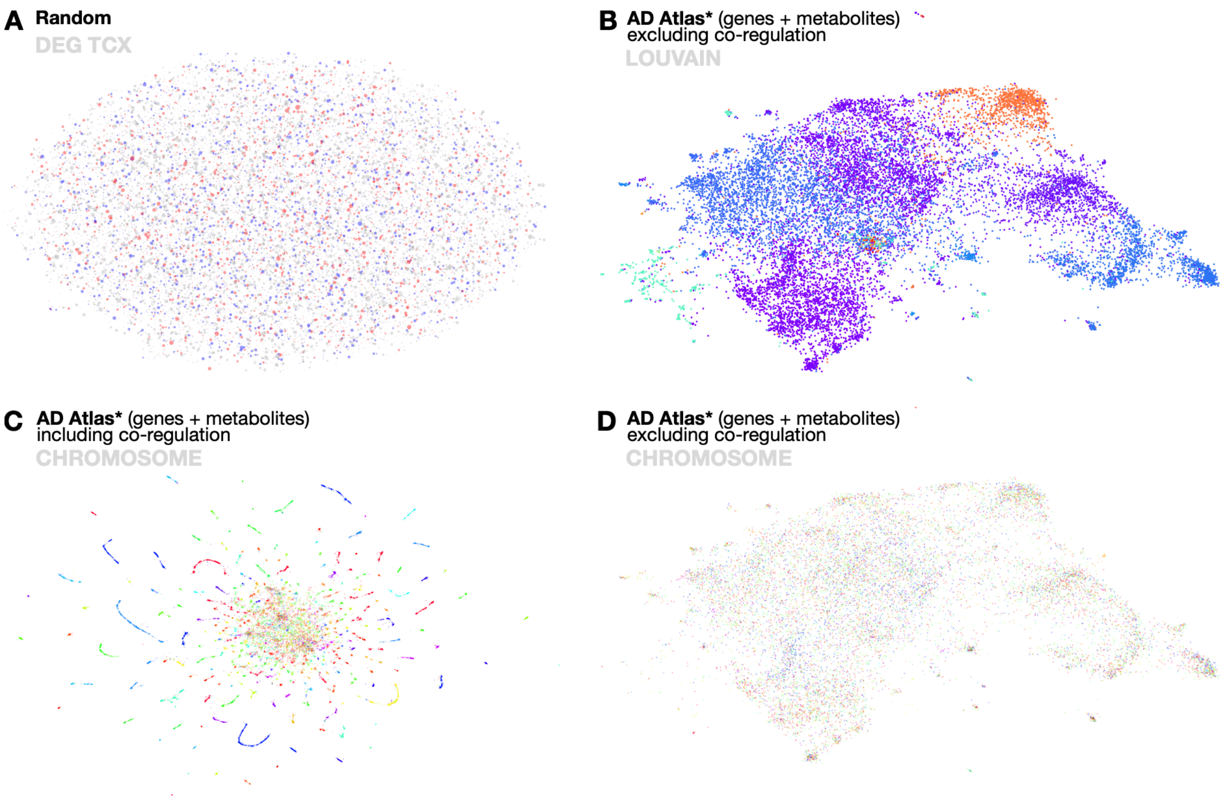

**Figure S10. Additional analysis of the global network structure of the AD Atlas.** The nodes of the AD Atlas network were embedded into a 130-dimensional vector space and projected to 2D for visualization, as described in **MATERIALS and METHODS**. **(A)** Randomized network as implemented by the GeneWalk (*104*) package, colored by differential expression in the temporal cortex. This shows, as expected, no structure or clustering. **(B)** Communities found by applying the network community detection algorithm Louvain (*105*) (as implemented by the Python package *NetworkX;* default settings) directly on the (simplified) network. Communities found in the network align with overall topology of embedding visualization. **(C)** Including co-regulation edges (filtered for the brain cortex and frontal cortex) in the network embedding (117-dimensional vector space) introduces clusters primarily driven by genetic architecture (genomic location of genes indicated by different colors for each chromosome). **(D)** Excluding co-regulation edges largely removes this effect with only few chromosomal clusters remaining. Gene nodes are again colored by chromosome.

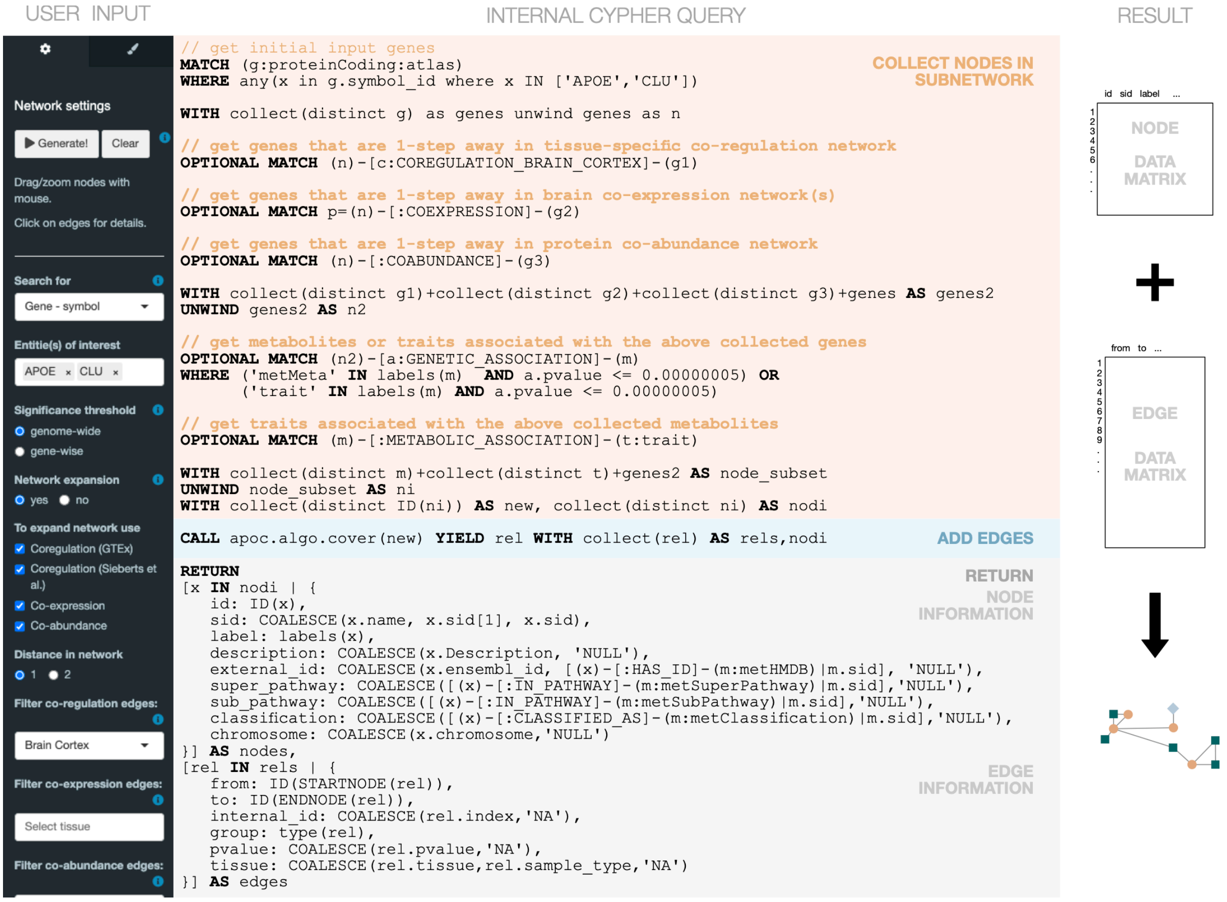

**Figure S11. Example cypher query.** Users can dynamically create context-specific networks by using the "Network settings" sidebar of the AD Atlas web interface. The user specified options are then translated internally into a cypher query that is used to communicate with the underlying Neo4j database. The returned data matrices are processed, which includes further edge filtering, and used by the AD Atlas frontend to visualize the interactive multi-omics network using VisNetwork.

**Table S1.** Data summary and statistics. Edge numbers differ from main manuscript table as undirected edges are modelled by two directed edges in Neo4j. For the edge type COREGULATION, we introduced tissue-specific edges named COREGULATION_*tissue* to enable more efficient data queries.

| | | n |
|---|---|---|
| Nodes | | |
| | metaTrait *mt* | 10 |
| | trait *t* | 67 |
| | gene (protein-coding) *g* | 20,363 |
| | DEG in min. 1 AD phenotype | 14,731 |
| | DEP in min. 1 AD phenotype | 7,867 |
| | metMeta *m* | 1,328 |
| Edges | | |
| | (*t*) - [ :PART_OF ] -> (m*t*) | 105 |
| | (*g*) - [ :GENETIC_ASSOCIATION ] –> (*t*) | 12,361 |
| | (*g*) - [ :GENETIC_ASSOCIATION ] – >(*m*) | 165,719 |
| | (*m*) - [ :METABOLIC_ASSOCIATION ] –> (*t*) | 1,018 |
| | (*g*) - [ :COEXPRESSION ] – (*g*) | 465,032 |
| | (g) - [ :COEXPRESSION {tissue:IFG} ] – > (g) | 30,670 |
| | (g) - [ :COEXPRESSION {tissue:PHG} ] – > (g) | 34,892 |
| | (g) - [ :COEXPRESSION {tissue:TCX} ] – > (g) | 64,230 |
| | (g) - [ :COEXPRESSION {tissue:CBE} ] – > (g) | 75,066 |
| | (g) - [ :COEXPRESSION {tissue:STG} ] – > (g) | 35,656 |
| | (g) - [ :COEXPRESSION {tissue:DLPFC} ] – > (g) | 106,156 |
| | (g) - [ :COEXPRESSION {tissue:FP} ] – > (g) | 40,294 |
| | (*g*) - [ :COREGULATION ] – > (*g)* | 493,117 |
| | (*g*) - [ :COREGULATION_{tissue} ] – > (*g)* | 4,345,276 |
| | (g) - [ :COREGULATION_BRAIN_CAUDATE_BASAL_GANGLIA ] – > (g) | 61,761 |
| | (g) - [ :COREGULATION_BRAIN_CORTEX ] – > (g) | 246,462 |
| | (g) - [ :COREGULATION_BRAIN_CEREBELLUM ] – > (g) | 95,397 |
| | (g) - [ :COREGULATION_BRAIN_CEREBELLAR_HEMISPHERE ] – > (g) | 78,049 |
| | (g) - [ :COREGULATION_BRAIN_ANTERIOR_CINGULATE_CORTEX_BA24 ] – > (g) | 37,213 |
| | (g) - [ :COREGULATION_BRAIN_NUCLEUS_ACCUMBENS_BASAL_GANGLIA ] – > (g) | 60,903 |
| | (g) - [ :COREGULATION_BRAIN_PUTAMEN_BASAL_GANGLIA ] – > (g) | 50,454 |
| | (g) - [ :COREGULATION_BRAIN_AMYGDALA ] – > (g) | 23,817 |
| | (g) - [ :COREGULATION_BRAIN_SPINAL_CORD_CERVICAL_C_1 ] – > (g) | 28,965 |
| | (g) - [ :COREGULATION_BRAIN_FRONTAL_CORTEX_BA9 ] – > (g) | 53,237 |
| | (g) - [ :COREGULATION_BRAIN_HYPOTHALAMUS ] – > (g) | 37,824 |
| | (g) - [ :COREGULATION_BRAIN_HIPPOCAMPUS ] – > (g) | 38,548 |
| | (g) - [ :COREGULATION_BRAIN_SUBSTANTIA_NIGRA ] – > (g) | 20,255 |
| | (*g*) - [ :COABUNDANCE ] – (*g*) | 146,592 |
| | (*m*) - [ :PARTIAL_CORRELATION ] – (*m*) | 2,326 |

**Table S2.** Data sources and references.

| Data | Edge Type | Significance threshold | Cohort | Reference |
|---|---|---|---|---|
| GGM | **COABUNDACE** | 0.05/all possible edges | KORA | Suhre et al. (*16*) |
| | | *q*-value < 0.05 and<br>*p*-value(cor) significant | BLSA<br>ACT<br>MSSB<br>Banner | Johnson et al. (*32*) |
| | | *q*-value < 0.05 and<br>*p*-value(cor) significant | ROSMAP<br>Banner | Johnson et al. (*33*) |
| | **COEXPRESSION** | see publication | ROSMAP<br>MSBB<br>Mayo | Wan et al. (*27*) |
| | **DEG** | $P_{FDR} \leq 0.05$ | ROSMAP<br>MSBB<br>Mayo | Wan et al. (*27*) |
| | **DEP** | $P_{Tukeys} \leq 0.05$ | BLSA<br>ACT<br>MSSB<br>Banner | Johnson et al. (*32*) |
| | | $P_{Holm} \leq 0.05$ | ROSMAP<br>Banner | Johnson et al. (*33*) |
| GGM | **PARTIAL_CORRELATION** | *p*-value ≤ 7.96e-7 and<br>abs(cor) ≤ 0.1603 | KORA | Krumsiek et al.<br>(*15*) |
| | | $P_{Bonf} \leq 0.05$ | ROSMAP | Batra et al. (*31*) |
| | | 0.05/all possible edges | ADNI | This study |
| | | 0.05/all possible edges | ADNI | This study |
| eQTL | **COREGULATION** | *q*-value ≤ 0.05 | GTEx | GTEx Consortium<br>(*2*) |
| | | $P_{FDR} \leq 0.05$ | AMP-AD<br>CMC | Sieberts et al. (*4*) |
| mQTL | **GENETIC_ASSOCIATION** | P ≤ 0.0001 [#] | KORA | Suhre et al. (*12*) |
| | | P ≤ 0.0001 [#] | TwinsUK | Suhre et al. (*12*) |

| | | | | |
|---|---|---|---|---|
| | | P ≤ 0.0001 # | KORA<br>TwinsUK | Shin et al. (*11*) |
| | | P ≤ 0.0001 # | SHIP-0<br>KORA | Raffler et al. (*10*) |
| | | P ≤ 0.0001 # | Meta-analysis** | Draisma et al. (*13*) |
| | | P ≤ 0.00001 # | TwinsUK | Long et al. (*14*) |
| | | P ≤ 0.05 # | ROSMAP | This study |
| | | P ≤ 0.05 # | ROSMAP<br>MAYO | This study |
| | | P ≤ 0.05 # | ADNI | This study |
| mWAS | **METABOLIC_ASSOCIATION** | *p*-value ≤ 0.05/15 | ADNI | MahmoudianDehkordi et al. (*34*)<br>Nho et al. (*35*) |
| | | *p*-value ≤ 0.05/55 | ADNI | Arnold et al. (*36*) |
| | | $P_{FDR}$ ≤ 0.05 | ROSMAP | Batra et al. (*31*) |
| traitQTL | **GENETIC_ASSOCIATION** | P ≤ 0.05 # | ADNI | This study |
| | | P ≤ 0.05 # | Meta-analysis** | Lambert et al. (*17*) |
| | | P ≤ 0.05 # | Meta-analysis* | Beecham et al. (*18*) |
| | | P ≤ 0.05 # | ADNI<br>Knight ADRC | Deming et al. (*19*) |
| | | P ≤ 0.05 # | Meta-analysis* | Deming et al. (*20*) |
| | | P ≤ 0.05 # | Meta-analysis** | Huang et al. (*21*) |
| | | P ≤ 0.05 # | IGAP<br>UK Biobank | Marioni et al. (*22*) |
| | | P ≤ 0.05 # | IGAP | Marioni et al. (*22*) |
| | | P ≤ 0.05 # | Meta-analysis** | Kunkle et al. (*23*) |

| | | | |
|---|---|---|---|
| | P ≤ 0.05 # | PGC-Alz<br>IGAP<br>ADSP<br>UK Biobank | Jansen et al. (*24*) |
| | P ≤ 0.05 # | Meta-analysis* | Wightman et al. (*25*) |
| | P ≤ 0.05 # | Meta-analysis* | Bellenguez et al. (*26*) |

**meta-analysis across more than four different cohorts; **meta-analysis across IGAP; #results up to reported threshold are integrated in resource and genome-wide or gene-wide significance is applied to generate context-specific networks*

**Table S3.** Collections of AD-related phenotypes summarized as "metaTraits". A trait can be connected to multiple meta traits. Further trait descriptions are given in **Supplementary Table 4**.

| Meta trait | Trait |
|---|---|
| **AD CSF biomarker** | CSF-Abeta<br>CSF-Abeta (APOE4 adjusted)<br>CSF-Abeta pathology (based on threshold)<br>CSF-CLU<br>CSF-pTau<br>CSF-pTau (APOE4 adjusted)<br>CSF-pTau/Abeta<br>CSF-pTau/Abeta (APOE4 adjusted)<br>CSF-tTau<br>CSF-tTau (APOE4 adjusted)<br>CSF-tTau/Abeta<br>CSF-tTau/Abeta (APOE4 adjusted) |
| **AD imaging biomarker** | Amyloid-PET (ROI-based)<br>Amyloid-PET (ROI-based; APOE4 adjusted)<br>Amyloid-PET (global)<br>Amyloid-PET (global; APOE4 adjusted)<br>Cortex<br>Entorhinal cortex<br>Entorhinal cortex (APOE4 adjusted)<br>FDG-PET (ROI-based)<br>FDG-PET (ROI-based; APOE4 adjusted)<br>FDG-PET (global)<br>FDG-PET (global; APOE4 adjusted)<br>Hippocampus<br>Hippocampus (APOE4 adjusted)<br>Ventricles<br>White matter hyperintensities<br>White matter hyperintensities (APOE4 adjusted) |
| **AD risk/diagnosis** | AD age of onset<br>AD case-control (NP conservative)<br>AD case-control (NP relaxed)<br>AD case-control (clinical AD)<br>AD_by_proxy<br>APOE4<br>Clinical diagnosis (ordinal)<br>CN vs. AD **<br>CN vs. MCI *<br>MCI vs. AD *<br>Neuropathology diagnosis<br>NIA-Reagan Score<br>No AD vs. AD * |
| **Amyloid pathology** | Amyloid beta (Immunohistochemistry-based)<br>Amyloid-PET (ROI-based)<br>Amyloid-PET (ROI-based; APOE4 adjusted)<br>Amyloid-PET (global)<br>Amyloid-PET (global; APOE4 adjusted)<br>CSF-Abeta<br>CSF-Abeta (APOE4 adjusted)<br>CSF-Abeta pathology (based on threshold)<br>neuritic plaque (NP case-control)<br>neuritic plaque (NP ordinal by CERAD) |
| **Brain glucose uptake** | FDG-PET (ROI-based)<br>FDG-PET (ROI-based; APOE4 adjusted)<br>FDG-PET (global)<br>FDG-PET (global; APOE4 adjusted) |
| **Cognition** | ADAS-Cog13<br>ADAS-Cog13 (APOE4 adjusted)<br>ADNI-Executive-Function<br>ADNI-Executive-Function (APOE4 adjusted)<br>ADNI-Memory Score |

| | |
|---|---|
| | ADNI-Memory Score (APOE4 adjusted)<br>Clinical Dementia Rating - sum of boxes<br>Cognition<br>Cognitive Decline<br>Mini-mental state exam<br>RAVLT total<br>RAVLT total (APOE4 adjusted) |
| **Neurodegeneration** | Cortex<br>Entorhinal cortex<br>Entorhinal cortex (APOE4 adjusted)<br>Hippocampus<br>Hippocampus (APOE4 adjusted)<br>Ventricles |
| **Neuropathology** | Amyloid beta (Immunohistochemistry-based)<br>cerebral amyloid angiopathy (CAA)<br>Lewy body disease (LBD case-control)<br>Lewy body disease (LBD ordinal - 3 categories)<br>Lewy body disease (LBD ordinal - 5 categories)<br>Global burden of pathology<br>hippocampal sclerosis (HS)<br>neuritic plaque (NP case-control)<br>neuritic plaque (NP ordinal by CERAD)<br>neurofibrillary tangle (NFT Braak ordinal I - 7 Braak stages)<br>neurofibrillary tangle (NFT Braak ordinal II - 4 Braak groups)<br>Tau tangles (immunohistochemistry-based)<br>vascular brain injury (VBI case-control)<br>vascular brain injury (VBI ordinal) |
| **Other brain pathologies** | Lewy body disease (LBD case-control)<br>Lewy body disease (LBD ordinal - 3 categories)<br>Lewy body disease (LBD ordinal - 5 categories)<br>White matter hyperintensities<br>White matter hyperintensities (APOE4 adjusted)<br>hippocampal sclerosis (HS)<br>vascular brain injury (VBI case-control)<br>vascular brain injury (VBI ordinal) |
| **Tau pathology** | CSF-pTau<br>CSF-pTau (APOE4 adjusted)<br>CSF-tTau<br>CSF-tTau (APOE4 adjusted)<br>neurofibrillary tangle (NFT Braak ordinal I - 7 Braak stages)<br>neurofibrillary tangle (NFT Braak ordinal II - 4 Braak groups)<br>Tau tangles (immunohistochemistry-based) |

*ADNI; **ADNI and IGAP studies

**Table S4.** Description of AD-related phenotypes with respective publications as currently included in the AD Atlas.

| Trait | Description | Publication | Type |
|---|---|---|---|
| **AD age of onset** | Age at onset of AD-defined survival (AAOS) in AD cases and nondemented elderly controls. Genome-wide survival analysis was performed through Cox proportional hazards regression where the time scale is defined as age in years ('age': AAO for cases and age at last assessment for controls). | Huang et al. | GWAS |
| **AD by proxy** | Proxy phenotype for AD case-control status inferred from UK Biobank self-report data on known parental LOAD status. Includes different types of analysis including meta-analysis of maternal and paternal AD status, parental LOAD status weighted by parental age as well as meta-analysis with both proxy and clinical AD case-controls. | Marioni et al. (meta-analysis)<br>Marioni et al. (parental)<br>Wightman et al.<br>Jansen et al.<br>Bellenguez et al. | GWAS<br>GWAS<br>GWAS<br>GWAS<br>GWAS |
| **AD case-control (clinical AD)** | Clinical AD case-control. Case: met DSM-IV criteria or had a clinical dementia rating greater than zero. Control: did not meet DSM-IV criteria for dementia, had no mild cognitive impairment and - when available - a clinical dementia rating of zero. | Beecham et al. | GWAS |
| **AD case-control (NP conservative)** | Clinico-pathologic AD dementia phenotype where cases had clinical dementia with core AD neuropathologic changes, and controls were not clinically demented and had none or minimal AD neuropathologic changes. Thorough documentation of neuropathologic assessment required, including documentation of the NIA/Reagan assessment or complete documentation of both the NFT Braak stage and the NP score. | Beecham et al. | GWAS |
| AD case-control (NP relaxed) | Clinico-pathologic AD dementia phenotype where cases had clinical dementia with core AD neuropathologic changes, and controls were not clinically demented and had none or minimal AD neuropathologic changes. No thorough documentation of neuropathologic assessment required. | Beecham et al. | GWAS |
| **ADAS-Cog13** | Alzheimer's disease assessment scale (ADAS) - 13-item cognitive subscale. | ADNI | GWAS |
| ADAS-Cog13 (APOE4 adjusted) | Alzheimer's disease assessment scale (ADAS) - 13-item cognitive subscale; Copies of APOE ε4 included as covariate. | ADNI<br>Arnold et al.<br>MahmoudianDehkordi et al. and Nho et al. | GWAS<br>MWAS<br>MWAS |

| | | | |
|---|---|---|---|
| **ADNI-Executive-Function** | ADNI composite score for executive function using items from different cognitive tests. | ADNI | GWAS |
| ADNI-Executive-Function (APOE4 adjusted) | ADNI composite score for executive function using items from different cognitive tests; Copies of APOE ε4 included as covariate. | ADNI | GWAS |
| **ADNI-Memory Score** | ADNI composite score for memory using items from different cognitive tests. | ADNI | GWAS |
| ADNI-Memory Score (APOE4 adjusted) | ADNI composite score for memory using items from different cognitive tests; Copies of APOE ε4 included as covariate. | ADNI | GWAS |
| **Amyloid beta (Immunohistochemistry-based)** | Immunohistochemistry-based overall Aβ load (square root). | Batra et al. | MWAS |
| Amyloid-PET (global; APOE4 adjusted) | Global cortical [18F] Florbetapir PET (co-registered, averaged, standardized image and voxel size, uniform resolution) SUVR (intensity-normalized using a whole cerebellum reference region); Copies of APOE ε4 included as covariate. | ADNI | GWAS |
| **Amyloid-PET (global)** | Global cortical [18F] Florbetapir PET (co-registered, averaged, standardized image and voxel size, uniform resolution) SUVR (intensity-normalized using a whole cerebellum reference region). | ADNI | GWAS |
| Amyloid-PET (ROI-based; APOE4 adjusted) | Global cortical [18F] Florbetapir PET (co-registered, averaged, standardized image and voxel size, uniform resolution) SUVR (intensity-normalized using a whole cerebellum reference region); SUVR value for region of interest was extracted using MarsBaR from global cortical values based on an independent comparison of ADNI-1 [11C] Pittsburgh Compound B SUVR scans (regions where AD > CN); Copies of APOE ε4 included as covariate. | ADNI | GWAS |
| Amyloid-PET (ROI-based) | Global cortical [18F] Florbetapir PET (co-registered, averaged, standardized image and voxel size, uniform resolution) SUVR (intensity-normalized using a whole cerebellum reference region); SUVR value for region of interest was extracted using MarsBaR from global cortical values based on an independent comparison of ADNI-1 [11C] Pittsburgh Compound B SUVR scans (regions where AD > CN). | ADNI | GWAS |
| **APOE4** | Metabolite associations with copies of APOE ε4 (additive model) | Arnold et al.<br>MahmoudianDehkordi et al. and Nho et al. | MWAS<br>MWAS |
| **cerebral amyloid angiopathy (CAA)** | Cerebral amyloid angiopathy (CAA) analyzed as co-morbid neuropathologic phenotype using presence vs. absence analyses. | Beecham et al. | GWAS |

| | | | |
|---|---|---|---|
| **Clinical Dementia Rating - sum of boxes** | Copies of APOE ε4 included as covariate. | Arnold et al.<br>MahmoudianDehkordi et al. and Nho et al. | MWAS<br>MWAS |
| **Clinical diagnosis (ordinal)** | Consensus cognitive diagnosis at time of death analyzed with ordinal ranking - three categories: AD, Mild cognitive impairment (MCI), No cognitive impairment (NCI). | Batra et al. | MWAS |
| **CN vs. AD** | Case-control AD study. Includes meta-analysis. | ADNI<br>Arnold et al.<br>MahmoudianDehkordi et al. and Nho et al.<br>Lambert et al.<br>Kunkle et al. | GWAS<br>MWAS<br>MWAS<br>GWAS<br>GWAS |
| **CN vs. MCI** | Case-control study of CN participants vs. participants with MCI (early and late). | ADNI<br>Arnold et al.<br>MahmoudianDehkordi et al. and Nho et al. | GWAS<br>MWAS<br>MWAS |
| **Cognition** | Global cognitive function determined at the last timepoint before death. | Batra et al. | MWAS |
| **Cognitive decline** | Rate of change in global cognition over time. | Batra et al. | MWAS |
| **Composite measure of brain atrophy (cross-region MRI analysis)** | Composite measure of atrophy in regions affected by AD (SPARE-AD). | Arnold et al.<br>MahmoudianDehkordi et al. and Nho et al. | MWAS<br>MWAS |
| **Cortex** | Global cortical grey matter volume based on MRI. Adjusted for Copies of APOE ε4. | Arnold et al.<br>MahmoudianDehkordi et al. and Nho et al. | MWAS<br>MWAS |
| **CSF-Abeta** | Amyloid beta (Aβ1-42) levels measured in cerebrospinal fluid (CSF). For meta-analysis the raw values were log10-transformed to approximate a normal distribution within each study and centralized by each study mean. | ADNI<br>Deming et al. | GWAS<br>GWAS |
| CSF-Abeta (APOE4 adjusted) | Amyloid beta (Aβ1-42) levels measured in cerebrospinal fluid (CSF) using the Roche Elecsys immunoassay. Copies of APOE ε4 included as covariate. | ADNI<br>Arnold et al.<br>MahmoudianDehkordi et al. and Nho et al. | GWAS<br>MWAS<br>MWAS |
| CSF-Abeta pathology (based on threshold) | Amyloid beta (Aβ1-42) positivity in cerebrospinal fluid (CSF) based on measures using the Roche Elecsys immunoassay and a threshold of 1073 pg/ml (< positive; >= negative); Copies of APOE ε4 included as covariate. | Arnold et al.<br>MahmoudianDehkordi et al. and Nho et al. | MWAS<br>MWAS |
| **CSF-CLU** | Clusterin (CLU) levels measured in cerebrospinal fluid (CSF). For meta-analysis the raw values were log10-transformed to approximate a normal distribution within each study and centralized by each study mean. | Deming et al. | GWAS |

| | | | |
|---|---|---|---|
| **CSF-pTau** | Phosphorylated tau (ptau181) levels measured in cerebrospinal fluid (CSF). For meta-analysis the raw values were log10-transformed to approximate a normal distribution within each study and centralized by each study mean. | ADNI<br>Deming et al. | GWAS<br>GWAS |
| CSF-pTau (APOE4 adjusted) | Phosphorylated tau (ptau181) levels measured in cerebrospinal fluid (CSF) using the Roche Elecsys immunoassay. Copies of APOE ε4 included as covariate. | ADNI<br>Arnold et al.<br>MahmoudianDehkordi et al. and Nho et al. | GWAS<br>MWAS<br>MWAS |
| **CSF-pTau/Abeta** | Ratio of phosphorylated tau levels and amyloid beta 1-42 levels measured in cerebrospinal fluid (CSF) using the Roche Elecsys immunoassay. | ADNI | GWAS |
| CSF-pTau/Abeta (APOE4 adjusted) | Ratio of phosphorylated tau levels and amyloid beta 1-42 levels measured in cerebrospinal fluid (CSF) using the Roche Elecsys immunoassay; Copies of APOE ε4 included as covariate. | ADNI<br>Arnold et al.<br>MahmoudianDehkordi et al. and Nho et al. | GWAS<br>MWAS<br>MWAS |
| **CSF-tTau** | Total tau levels measured in cerebrospinal fluid (CSF) using the Roche Elecsys immunoassay. For meta-analysis the raw values were log10-transformed to approximate a normal distribution within each study and centralized by each study mean. | ADNI<br>Deming et al. | GWAS<br>GWAS |
| CSF-tTau (APOE4 adjusted) | Total tau levels measured in cerebrospinal fluid (CSF) using the Roche Elecsys immunoassay; Copies of APOE ε4 included as covariate. | ADNI<br>Arnold et al.<br>MahmoudianDehkordi et al. and Nho et al. | GWAS<br>MWAS<br>MWAS |
| **CSF-tTau/Abeta** | Ratio of total tau levels and amyloid beta 1-42 levels measured in cerebrospinal fluid (CSF) using the Roche Elecsys immunoassay. | ADNI | GWAS |
| CSF-tTau/Abeta (APOE4 adjusted) | Ratio of total tau levels and amyloid beta 1-42 levels measured in cerebrospinal fluid (CSF) using the Roche Elecsys immunoassay; Copies of APOE ε4 included as covariate. | ADNI<br>Arnold et al.<br>MahmoudianDehkordi et al. and Nho et al. | GWAS<br>MWAS<br>MWAS |
| **Entorhinal cortex** | Entorhinal cortical thickness from MRI. | ADNI | GWAS |
| Entorhinal cortex (APOE4 adjusted) | Entorhinal cortical thickness from MRI; Copies of APOE ε4 included as covariate. | ADNI<br>Arnold et al.<br>MahmoudianDehkordi et al. and Nho et al. | GWAS<br>MWAS<br>MWAS |
| FDG-PET (global; APOE4 adjusted) | Global cortical [18F] FDG PET (co-registered, averaged, standardized image and voxel size, uniform resolution) SUVR (intensity-normalized using a pons reference region); Copies of APOE ε4 included as covariate. | ADNI | GWAS |
| **FDG-PET (global)** | Global cortical [18F] FDG PET (co-registered, averaged, standardized image and voxel size, | ADNI | GWAS |

| | | | |
|---|---|---|---|
| | uniform resolution) SUVR (intensity-normalized using a pons reference region). | | |
| FDG-PET (ROI-based; APOE4 adjusted) | Global cortical [18F] FDG PET (co-registered, averaged, standardized image and voxel size, uniform resolution) SUVR (intensity-normalized using a pons reference region); a mean SUVR value was extracted from global cortical values representing regions where AD patients show decreased glucose metabolism relative to cognitively normal older participants (CN) from the full ADNI-1 cohort; Copies of APOE ε4 included as covariate. | ADNI<br>Arnold et al.<br>MahmoudianDehkordi et al. and Nho et al. | GWAS<br>MWAS<br>MWAS |
| FDG-PET (ROI-based) | Global cortical [18F] FDG PET (co-registered, averaged, standardized image and voxel size, uniform resolution) SUVR (intensity-normalized using a pons reference region); a mean SUVR value was extracted from global cortical values representing regions where AD patients show decreased glucose metabolism relative to cognitively normal older participants (CN) from the full ADNI-1 cohort. | ADNI | GWAS |
| **Global burden of pathology** | Summary of pathology derived from counts of: neuritic plaques, diffuse plaques, and neurofibrillary tangles. | Batra et al. | MWAS |
| **hippocampal sclerosis (HS)** | Hippocampal sclerosis (HS) analyzed as co-morbid neuropathologic phenotype using presence vs. absence analyses. | Beecham et al. | GWAS |
| **Hippocampus** | Hippocampal grey matter volume from MRI. | ADNI | GWAS |
| Hippocampus (APOE4 adjusted) | Hippocampal grey matter volume from MRI; Copies of APOE ε4 included as covariate. | ADNI<br>Arnold et al.<br>MahmoudianDehkordi et al. and Nho et al. | GWAS<br>MWAS<br>MWAS |
| **Lewy body disease (LBD case-control)** | Lewy body disease (LBD) analyzed with a presence vs. absence - any LBD vs. no LBD - analysis. | Beecham et al. | GWAS |
| Lewy body disease (LBD ordinal - 3 categories) | Lewy body disease (LBD) analyzed with ordinal ranking - three categories: none, brainstem-predominant, and all other regions or not specified. | Beecham et al. | GWAS |
| Lewy body disease (LBD ordinal - 5 categories) | Lewy body disease (LBD) analyzed with ordinal ranking - five categories: none, brainstem-predominant, limbic, neocortical, and other regions or not specified. | Beecham et al. | GWAS |
| **MCI vs. AD** | case/control study of ADNI participants with MCI (early and late) vs. cases with clinical AD. | ADNI | GWAS |
| **Mini-mental state exam** | Adjusted for Copies of APOE ε4. | Arnold et al.<br>MahmoudianDehkordi et al. and Nho et al. | MWAS<br>MWAS |

| | | | |
|---|---|---|---|
| **neuritic plaque (NP case-control)** | Neuritic plaques (NPs) analyzed with a presence vs. absence - any NPs vs. no NPs - analysis. | Beecham et al. | GWAS |
| neuritic plaque (NP ordinal by CERAD) | Neuritic plaques (NPs) analyzed with ordinal ranking - four CERAD scores: none, sparse, moderate, frequent. | Beecham et al. | GWAS |
| **neurofibrillary tangle (NFT Braak ordinal I - 7 Braak stages)** | Neurofibrillary tangles (NFTs) analyzed by well-established ordinal ranking - seven Braak stages: none, I, II, III, IV, V, and VI. | Beecham et al. | GWAS |
| neurofibrillary tangle (NFT Braak ordinal II - 4 Braak groups) | Neurofibrillary tangles (NFTs) analyzed by well-established ordinal ranking - four Braak groups: none, transentorhinal, limbic, isocortical. | Beecham et al. | GWAS |
| **Neuropathology diagnosis** | Mayo clinic post mortem diagnosis of Alzheimer's disease derived from Braak and CERAD scores. AD case status was assigned where Braak stage was ≥4 and CERAD score was ≤2; control case status was assigned where Braak stage was ≤3 and CERAD score was ≥3. | Batra et al. | MWAS |
| **NIA-Reagan Score** | NIA Reagan diagnosis of Alzheimer's disease derived from Braak and CERAD scores; binarized (0: low likelihood of AD, 1: high likelihood of AD). | Batra et al. | MWAS |
| No AD vs. AD | Case/control setting combining all non-demented individuals (CN, SMC, EMCI, LMCI) into the control group vs. cases with clinical AD. | ADNI | GWAS |
| **Progression MCI -> AD** | Ever-progression (coded as 0/1) observed in ADNI participants during follow-up (up to ten years) with baseline diagnosis of late MCI. | Arnold et al.<br>MahmoudianDehkordi et al. and Nho et al. | MWAS<br>MWAS |
| **RAVLT total** | Rey Auditory-Verbal Learning Test (RAVLT). | ADNI | GWAS |
| RAVLT total (APOE4 adjusted) | Rey Auditory-Verbal Learning Test (RAVLT); Copies of APOE ε4 included as covariate. | ADNI | GWAS |
| **Tau tangles (immunohistochemistry-based)** | Immunohistochemistry-based overall paired helical filament (PHF)-tau tangles load. | Batra et al. | MWAS |
| **vascular brain injury (VBI case-control)** | Vascular brain injury (VBI) analyzed with a presence vs. absence - any VBI vs. no VBI - analysis. | Beecham et al. | GWAS |
| vascular brain injury (VBI ordinal) | Vascular brain injury (VBI) analyzed with ordinal ranking - three categories: none, any microinfarcts, any lacunar or territorial infarcts. | Beecham et al. | GWAS |
| **Ventricles** | Ventricular grey matter volume from MRI. | Arnold et al.<br>MahmoudianDehkordi et al. and Nho et al. | MWAS<br>MWAS |

| | | | |
|---|---|---|---|
| **White matter hyperintensities** | Global cortical white matter hyperintensities from MRI. | ADNI | GWAS |
| White matter hyperintensities (APOE4 adjusted) | Global cortical white matter hyperintensities from MRI; Copies of APOE ε4 included as covariate. | ADNI | GWAS |

**Table S5.** Statistics for clusters obtained by applying hierarchical clustering on the 130D embedding vectors and using cut h = 30.

| Cluster* | Color HEX | $n$ | $n_{metabolites}$ | $n_{genes}$ | $n_{genesAnno}$** |
|---|---|---|---|---|---|
| **1** | #CDC08C | 318 | 301 | 17 | 9 |
| **2** | #A2A475 | 368 | 0 | 368 | 329 |
| **3** | #F1BB7B | 356 | 0 | 356 | 304 |
| **4** | #5B1A18 | 425 | 0 | 425 | 370 |
| **5** | #FDDDA0 | 93 | 12 | 81 | 77 |
| **6** | #D67236 | 726 | 0 | 726 | 563 |
| **7** | #C6CDF7 | 1047 | 0 | 1047 | 852 |
| **8** | #FD6467 | 15 | 15 | 0 | 0 |
| **9** | #02401B | 476 | 2 | 474 | 351 |
| **10** | #E1AF00 | 614 | 1 | 613 | 446 |
| **11** | #E6A0C4 | 818 | 0 | 818 | 611 |
| **12** | #7294D4 | 146 | 0 | 146 | 111 |
| **13** | #3B9AB2 | 696 | 16 | 680 | 501 |
| **14** | #81A88D | 748 | 0 | 748 | 593 |
| **15** | #EBCC2A | 532 | 2 | 530 | 367 |
| **16** | #F4B5BD | 2678 | 3 | 2675 | 2113 |
| **17** | #FAD77B | 1928 | 2 | 1926 | 1464 |
| **18** | #D8A499 | 397 | 0 | 397 | 355 |
| **19** | #85D4E3 | 113 | 0 | 113 | 109 |
| **20** | #D8B70A | 33 | 9 | 24 | 21 |
| **21** | #9A8822 | 573 | 180 | 393 | 252 |
| **22** | #78B7C5 | 261 | 0 | 261 | 211 |
| **23** | #F8AFA8 | 1003 | 3 | 1000 | 678 |
| **24** | #F5CDB4 | 548 | 1 | 547 | 385 |
| **25** | #F21A00 | 437 | 9 | 428 | 269 |

* corresponding to clusters indicated in **Figure 5** of the main manuscript **genes annotated with one or more biodomain annotations

**Table S6.** Clusters with significant enrichment for biodomains. Clusters were obtained by applying hierarchical clustering on the 130D node vectors and using cut h = 30. Multiple testing correction was performed using Bonferroni, resulting in a significance threshold of *p*-value ≤ 0.002(0.05/25 clusters). Significance for enrichment of differentially expressed genes (temporal cortex in AD) and average path length (to AD phenotypes) is also given.

| Cluster* | Term | *p*-value | OR | 95% CI** | $P_{adj}$ |
|---|---|---|---|---|---|
| 1 | path_length_less | 4.96e-17 | 1.72 | -0.43 | 1.24e-15 |
| 2 | Autophagy | 1.46e-13 | 3.31 | 2.57 | 3.65e-12 |
| 2 | Endolysosome | 6.10e-10 | 2.32 | 1.86 | 1.53e-08 |
| 2 | Immune_Response | 2.60e-81 | 9.20 | 7.51 | 6.51e-80 |
| 2 | Lipid_Metabolism | 6.27e-08 | 1.91 | 1.56 | 1.57e-06 |
| 2 | Vasculature | 2.52e-04 | 1.78 | 1.36 | 6.30e-03 |
| 2 | path_length_less | 3.77e-42 | 1.92 | -0.29 | 9.44e-41 |
| 2 | DEG_TCX_up | 2.37e-30 | 3.60 | 3.00 | 5.92e-29 |
| 3 | Apoptosis | 2.01e-12 | 2.62 | 2.10 | 5.01e-11 |
| 3 | Immune_Response | 1.23e-41 | 5.10 | 4.18 | 3.08e-40 |
| 3 | Lipid_Metabolism | 2.70e-05 | 1.69 | 1.36 | 6.75e-04 |
| 3 | Oxidative_Stress | 3.25e-04 | 2.18 | 1.51 | 8.12e-03 |
| 3 | Vasculature | 2.24e-08 | 2.40 | 1.86 | 5.60e-07 |
| 3 | path_length_less | 1.49e-11 | 2.08 | -0.13 | 3.73e-10 |
| 3 | DEG_TCX_up | 2.05e-77 | 7.98 | 6.57 | 5.12e-76 |
| 4 | Immune_Response | 9.40e-04 | 1.48 | 1.20 | 2.35e-02 |
| 4 | Lipid_Metabolism | 3.19e-06 | 1.70 | 1.40 | 7.97e-05 |
| 4 | Structural_Stabilization | 8.18e-19 | 2.56 | 2.14 | 2.05e-17 |
| 4 | Vasculature | 4.04e-44 | 5.72 | 4.71 | 1.01e-42 |
| 4 | path_length_less | 3.29e-06 | 2.14 | -0.07 | 8.22e-05 |
| 4 | DEG_TCX_up | 9.08e-108 | 10.27 | 8.49 | 2.27e-106 |
| 5 | Autophagy | 1.91e-05 | 3.80 | 2.28 | 4.76e-04 |
| 5 | Endolysosome | 2.48e-08 | 4.12 | 2.71 | 6.20e-07 |
| 5 | Immune_Response | 1.01e-24 | 11.81 | 7.52 | 2.53e-23 |
| 5 | Lipid_Metabolism | 8.67e-06 | 2.85 | 1.90 | 2.17e-04 |
| 5 | path_length_less | 4.19e-08 | 1.95 | -0.21 | 1.05e-06 |
| 6 | Lipid_Metabolism | 7.41e-07 | 1.60 | 1.36 | 1.85e-05 |
| 6 | Myelination | 3.41e-18 | 5.01 | 3.79 | 8.52e-17 |
| 6 | Structural_Stabilization | 8.43e-06 | 1.47 | 1.27 | 2.11e-04 |
| 6 | path_length_less | 4.94e-05 | 2.18 | -0.04 | 1.24e-03 |
| 6 | DEG_TCX_unchanged | 6.86e-30 | 2.61 | 2.24 | 1.72e-28 |
| 7 | Lipid_Metabolism | 1.81e-19 | 2.01 | 1.77 | 4.52e-18 |
| 7 | Myelination | 6.64e-09 | 2.81 | 2.11 | 1.66e-07 |
| 7 | Structural_Stabilization | 1.44e-08 | 1.50 | 1.33 | 3.61e-07 |
| 7 | Vasculature | 3.21e-17 | 2.32 | 1.98 | 8.02e-16 |
| 7 | path_length_less | 4.07e-11 | 2.15 | -0.07 | 1.02e-09 |
| 7 | DEG_TCX_up | 1.67e-240 | 9.45 | 8.41 | 4.17e-239 |
| 8 | path_length_more | 3.44e-06 | 3.07 | 0.61 | 8.61e-05 |
| 9 | Epigenetic | 6.38e-04 | 1.54 | 1.24 | 1.60e-02 |
| 9 | DEG_TCX_up | 1.16e-63 | 5.11 | 4.35 | 2.90e-62 |
| 10 | Synapse | 5.54e-20 | 2.74 | 2.29 | 1.39e-18 |
| 10 | path_length_less | 4.74e-04 | 2.18 | -0.04 | 1.19e-02 |
| 10 | DEG_TCX_unchanged | 2.63e-13 | 1.89 | 1.63 | 6.58e-12 |
| 11 | Structural_Stabilization | 2.85e-10 | 1.69 | 1.47 | 7.12e-09 |
| 11 | Synapse | 2.19e-48 | 3.84 | 3.32 | 5.47e-47 |

| | | | | | |
|---|---|---|---|---|---|
| **11** | DEG_TCX_down | 1.71e-84 | 4.48 | 3.95 | 4.27e-83 |
| **12** | Synapse | 1.20e-03 | 2.02 | 1.38 | 3.00e-02 |
| **12** | path_length_more | 7.39e-04 | 2.40 | 0.07 | 1.85e-02 |
| **13** | Structural_Stabilization | 7.80e-06 | 1.50 | 1.28 | 1.95e-04 |
| **13** | Synapse | 2.51e-31 | 3.29 | 2.79 | 6.28e-30 |
| **13** | Vasculature | 1.67e-04 | 1.64 | 1.31 | 4.18e-03 |
| **13** | path_length_more | 5.58e-04 | 2.32 | 0.04 | 1.39e-02 |
| **13** | DEG_TCX_down | 2.31e-29 | 2.62 | 2.28 | 5.77e-28 |
| **14** | Epigenetic | 4.32e-07 | 1.66 | 1.40 | 1.08e-05 |
| **14** | RNA_Spliceosome | 9.16e-36 | 5.93 | 4.79 | 2.29e-34 |
| **14** | DEG_TCX_unchanged | 2.26e-04 | 1.32 | 1.16 | 5.65e-03 |
| **15** | Epigenetic | 7.22e-06 | 1.74 | 1.41 | 1.81e-04 |
| **15** | path_length_more | 4.37e-20 | 2.51 | 0.21 | 1.09e-18 |
| **15** | DEG_TCX_unchanged | 2.16e-14 | 2.07 | 1.75 | 5.40e-13 |
| **16** | Mitochondrial_Metabolism | 4.81e-11 | 1.52 | 1.36 | 1.20e-09 |
| **16** | Proteostasis | 1.37e-04 | 1.19 | 1.10 | 3.42e-03 |
| **16** | DEG_TCX_unchanged | 3.62e-05 | 1.19 | 1.11 | 9.06e-04 |
| **17** | Autophagy | 4.31e-06 | 1.57 | 1.33 | 1.08e-04 |
| **17** | Mitochondrial_Metabolism | 3.60e-08 | 1.49 | 1.32 | 9.00e-07 |
| **17** | Proteostasis | 1.06e-06 | 1.31 | 1.19 | 2.65e-05 |
| **17** | path_length_more | 4.99e-05 | 2.30 | 0.03 | 1.25e-03 |
| **17** | DEG_TCX_down | 5.01e-45 | 2.18 | 1.99 | 1.25e-43 |
| **18** | Mitochondrial_Metabolism | 5.77e-84 | 9.10 | 7.54 | 1.44e-82 |
| **18** | Proteostasis | 1.85e-04 | 1.48 | 1.23 | 4.63e-03 |
| **18** | DEG_TCX_unchanged | 5.97e-06 | 1.61 | 1.34 | 1.49e-04 |
| **19** | APP_Metabolism | 3.88e-12 | 10.53 | 6.44 | 9.70e-11 |
| **19** | Proteostasis | 6.02e-17 | 6.06 | 3.96 | 1.50e-15 |
| **19** | Structural_Stabilization | 2.03e-16 | 5.23 | 3.60 | 5.06e-15 |
| **19** | Synapse | 3.90e-04 | 2.17 | 1.49 | 9.76e-03 |
| **19** | path_length_less | 1.70e-06 | 2.09 | -0.10 | 4.25e-05 |
| **19** | DEG_TCX_unchanged | 1.10e-10 | 5.04 | 3.04 | 2.75e-09 |
| **20** | Structural_Stabilization | 2.00e-05 | 7.96 | 3.00 | 5.01e-04 |
| **21** | path_length_more | 4.70e-04 | 2.37 | 0.06 | 1.18e-02 |
| **21** | DEG_TCX_unchanged | 1.17e-06 | 1.74 | 1.42 | 2.94e-05 |
| **22** | Epigenetic | 2.38e-06 | 2.10 | 1.61 | 5.95e-05 |
| **22** | Proteostasis | 2.86e-10 | 2.42 | 1.89 | 7.16e-09 |
| **22** | DEG_TCX_unchanged | 1.65e-06 | 1.87 | 1.48 | 4.13e-05 |
| **23** | RNA_Spliceosome | 1.43e-04 | 1.90 | 1.43 | 3.57e-03 |
| **23** | path_length_more | 7.74e-24 | 2.45 | 0.17 | 1.93e-22 |
| **23** | DEG_TCX_down | 5.12e-37 | 2.48 | 2.21 | 1.28e-35 |
| **24** | Epigenetic | 1.76e-07 | 1.87 | 1.53 | 4.41e-06 |
| **24** | RNA_Spliceosome | 2.38e-06 | 2.61 | 1.88 | 5.95e-05 |
| **24** | DEG_TCX_unchanged | 3.84e-11 | 1.85 | 1.57 | 9.61e-10 |
| **25** | path_length_more | 2.21e-16 | 2.53 | 0.23 | 5.52e-15 |
| **25** | DEG_TCX_unchanged | 7.28e-14 | 2.24 | 1.85 | 1.82e-12 |

[*] corresponding to clusters indicated in **Figure 5** of the main manuscript **lower bound of one-sided test (alternative = 'greater') reported except for 'path_length_less', where the upper bound is given (alternative = 'less')

**Table S7.** Number of measured metabolites that are consolidated in each meta-metabolite listed with the number of their respective occurrence. For example, n=148 meta-metabolites consolidate 3 measured metabolites. An example is given in **Supplementary Figure 1B**.

| Number of measured metabolites/compounds consolidated to one "meta-Metabolite" | n |
|---|---|
| 1 = no consolidation | 806 |
| 2 | 318 |
| 3 | 148 |
| 4 | 37 |
| 5 | 12 |
| 6 | 6 |
| 7 | 1 |

**Table S8.** Details on the metabolomics platform providers for the 522 meta-metabolites that consolidate 2 or more measured metabolites. For example, cross-platform mapping was performed for n=60 metabolites.

| Metabolomics platform provider | n |
|---|---|
| Metabolon | 462 |
| Biocrates | 0 |
| both | 60 |
| **Total meta-metabolites that consolidate >1 measured metabolite:** | 522 |

**Table S9.** Genes co-expressed with *TMEM119* but not *TREM2* that show genetic associations with AD-related phenotypes. The lowest p-value is reported for each locus.

| Gene symbol | AD-related phenotype | *p*-value |
|---|---|---|
| ARHGAP45 | AD by proxy | 4.09e-30 |
| ARHGAP45 | CN vs. AD | 3.06e-16 |
| ARPC1B | AD by proxy | 4.83e-11 |
| ATP8B4 | AD by proxy | 5.12e-10 |
| GPSM3 | AD by proxy | 8.76e-11 |
| HLA-DMA | AD by proxy | 2.09e-16 |
| HLA-DMA | ADAS-Cog13 | 1.04e-09 |

| | | |
|---|---|---|
| HLA-DMA | ADAS-Cog13 (APOE4 adjusted) | 3.15e-09 |
| HLA-DMA | ADNI-Executive-Function | 4.81e-08 |
| HLA-DMA | ADNI-Memory Score | 3.09e-08 |
| HLA-DMA | CN vs. AD | 5.10e-12 |
| HLA-DMA | MCI vs. AD | 3.16e-10 |
| HLA-DMA | No AD vs. AD | 2.78e-10 |
| INPP5D | AD by proxy | 1.04e-17 |
| INPP5D | CN vs. AD | 3.42e-09 |
| SPN | AD by proxy | 5.39e-10 |
| TMEM106A | ADAS-Cog13 | 2.95e-10 |
| TMEM106A | ADAS-Cog13 (APOE4 adjusted) | 4.92e-09 |
| TMEM106A | ADNI-Memory Score | 3.52e-09 |
| TMEM106A | CN vs. AD | 2.56e-08 |

** metabolonic lactone sulfate (partially characterized metabolite)

**Table S10.** Genes co-expressed with *TMEM119* but not *TREM2* that show genetic associations with AD phenotypes and metabolite traits (metabolite quantitative trait loci (mQTL)). The lowest p-value is reported for each locus.

| **Gene symbol** | **Metabolic trait** | ***p*-value** |
|---|---|---|
| ARPC1B | 16a-hydroxy DHEA 3-sulfate | 1.46e-28 |
| ARPC1B | 4-androsten-3alpha,17alpha-diol monosulfate (3) | 1.85e-27 |
| ARPC1B | 4-androsten-3beta,17beta-diol disulfate (1) | 6.27e-09 |
| ARPC1B | 4-androsten-3beta,17beta-diol monosulfate (1) | 1.52e-11 |
| ARPC1B | 5alpha-androstan-3alpha,17beta-diol monosulfate (1) | 2.97e-09 |
| ARPC1B | 5alpha-androstan-3beta,17beta-diol disulfate | 9.27e-33 |
| ARPC1B | andro steroid monosulfate (1)* | 1.40e-21 |
| ARPC1B | androsterone sulfate | 8.82e-113 |
| ARPC1B | dehydroisoandrosterone sulfate (DHEA-S) | 6.70e-14 |
| ARPC1B | epiandrosterone sulfate | 2.80e-75 |
| ARPC1B | X-12063** | 1.67e-109 |
| INPP5D | 1-arachidonoyl-GPA (20:4) | 1.68e-09 |
| INPP5D | bilirubin | 3.21e-11 |
| INPP5D | biliverdin | 6.33e-17 |
| INPP5D | X-11441 | 5.48e-17 |
| INPP5D | X-11442 | 1.56e-16 |
| INPP5D | X-11530 | 6.42e-13 |

** metabolonic lactone sulfate (partially characterized metabolite)

**Table S11.** Genes co-expressed with *TREM2* but not *TMEM119* that show genetic associations with AD-related phenotypes. The lowest p-value is reported for each locus.

| Gene symbol | AD-related phenotype | *p*-value |
|---|---|---|
| AIF1 | AD by proxy | 1.19e-16 |
| AIF1 | CN vs. AD | 7.16e-09 |
| APOC1 | AD age of onset | 4.32e-131 |
| APOC1 | AD by proxy | 0.00e+00 |
| APOC1 | AD case-control (clinical AD) | 2.64e-63 |
| APOC1 | AD case-control (NP conservative) | 3.48e-38 |
| APOC1 | AD case-control (NP relaxed) | 2.02e-62 |
| APOC1 | ADAS-Cog13 | 1.04e-29 |
| APOC1 | ADNI-Executive-Function | 2.80e-17 |
| APOC1 | ADNI-Memory Score | 4.29e-38 |
| APOC1 | Amyloid-PET (global) | 1.14e-44 |
| APOC1 | Amyloid-PET (ROI-based) | 1.16e-42 |
| APOC1 | cerebral amyloid angiopathy (CAA) | 2.92e-21 |
| APOC1 | CN vs. AD | 0.00e+00 |
| APOC1 | CN vs. MCI | 4.30e-15 |
| APOC1 | CSF-Abeta | 4.78e-94 |
| APOC1 | CSF-Abeta (APOE4 adjusted) | 4.67e-16 |
| APOC1 | CSF-pTau | 5.67e-37 |
| APOC1 | CSF-pTau/Abeta | 8.84e-95 |
| APOC1 | CSF-pTau/Abeta (APOE4 adjusted) | 3.17e-16 |
| APOC1 | CSF-tTau | 4.61e-30 |
| APOC1 | CSF-tTau/Abeta | 7.97e-97 |
| APOC1 | CSF-tTau/Abeta (APOE4 adjusted) | 1.76e-17 |
| APOC1 | Entorhinal cortex | 1.14e-10 |
| APOC1 | FDG-PET (global) | 7.15e-10 |
| APOC1 | FDG-PET (ROI-based) | 9.45e-16 |
| APOC1 | Hippocampus | 1.80e-27 |
| APOC1 | Lewy body disease (LBD case-control) | 2.83e-11 |
| APOC1 | Lewy body disease (LBD ordinal - 3 categories) | 4.87e-12 |
| APOC1 | Lewy body disease (LBD ordinal - 5 categories) | 1.10e-12 |
| APOC1 | MCI vs. AD | 8.24e-09 |

| | | |
|---|---|---|
| APOC1 | neuritic plaque (NP case-control) | 1.78e-27 |
| APOC1 | neuritic plaque (NP ordinal by CERAD) | 1.37e-46 |
| APOC1 | neurofibrillary tangle (NFT Braak ordinal I - 7 Braak stages) | 4.73e-47 |
| APOC1 | neurofibrillary tangle (NFT Braak ordinal II - 4 Braak groups) | 4.83e-44 |
| APOC1 | No AD vs. AD | 3.66e-18 |
| APOC1 | RAVLT total | 1.98e-26 |
| GAL3ST4 | AD by proxy | 9.92e-19 |
| GAL3ST4 | CN vs. AD | 5.58e-10 |
| HLA-DPB1 | AD by proxy | 4.34e-16 |
| HLA-DPB1 | ADAS-Cog13 | 2.68e-11 |
| HLA-DPB1 | ADAS-Cog13 (APOE4 adjusted) | 6.17e-11 |
| HLA-DPB1 | ADNI-Executive-Function | 4.19e-09 |
| HLA-DPB1 | ADNI-Executive-Function (APOE4 adjusted) | 1.30e-08 |
| HLA-DPB1 | ADNI-Memory Score | 3.18e-10 |
| HLA-DPB1 | ADNI-Memory Score (APOE4 adjusted) | 8.76e-10 |
| HLA-DPB1 | CN vs. AD | 2.94e-12 |
| HLA-DPB1 | MCI vs. AD | 2.25e-12 |
| HLA-DPB1 | No AD vs. AD | 1.62e-12 |
| HLA-DRB1 | AD by proxy | 1.05e-17 |
| HLA-DRB1 | ADAS-Cog13 | 6.16e-09 |
| HLA-DRB1 | ADNI-Memory Score | 1.00e-09 |
| HLA-DRB1 | ADNI-Memory Score (APOE4 adjusted) | 8.24e-09 |
| HLA-DRB1 | CN vs. AD | 2.94e-12 |
| HLA-DRB1 | Entorhinal cortex | 6.18e-09 |
| HLA-DRB1 | MCI vs. AD | 6.42e-11 |
| HLA-DRB1 | No AD vs. AD | 3.00e-11 |
| HLA-DRB1 | RAVLT total | 3.52e-08 |
| ITGAM | AD by proxy | 8.50e-10 |
| ITGAX | AD by proxy | 7.71e-10 |
| LST1 | AD by proxy | 1.19e-16 |
| LST1 | CN vs. AD | 1.38e-09 |
| SPI1 | AD by proxy | 7.84e-12 |
| SPI1 | CN vs. AD | 5.46e-13 |

**Table S12.** Genes co-expressed with *TREM2* but not *TMEM119* that show genetic associations with AD phenotypes and metabolite traits (mQTLs). The lowest p-value is reported for each locus.

| **Gene symbol** | **Metabolic trait** | ***p*-value** |
|---|---|---|
| AIF1 | X-12712 | 2.12e-12 |
| APOC1 | cholesterol | 4.10e-10 |
| APOC1 | SM (OH) C22:1 | 1.36e-12 |
| APOC1 | SM (OH) C22:2 | 2.23e-08 |
| APOC1 | SM (OH) C24:1 | 7.91e-11 |
| APOC1 | SM C16:0 | 1.97e-09 |
| APOC1 | SM C18:0 | 1.16e-08 |
| APOC1 | SM C24:0 | 9.31e-10 |
| APOC1 | X-11820 | 1.22e-16 |
| GAL3ST4 | 16a-hydroxy DHEA 3-sulfate | 9.09e-12 |
| GAL3ST4 | 4-androsten-3alpha,17alpha-diol monosulfate (3) | 1.65e-13 |
| GAL3ST4 | 5alpha-androstan-3beta,17beta-diol disulfate | 1.76e-14 |
| GAL3ST4 | andro steroid monosulfate (1)* | 4.91e-10 |
| GAL3ST4 | androsterone sulfate | 3.07e-31 |
| GAL3ST4 | epiandrosterone sulfate | 9.11e-24 |
| GAL3ST4 | X-12063** | 1.67e-109 |
| HLA-DPB1 | N6-methyllysine | 3.87e-08 |
| HLA-DRB1 | X-11470 | 6.56e-13 |
| LST1 | X-11470 | 3.41e-09 |

** metabolonic lactone sulfate (partially characterized metabolite)

#### REFERENCES

1. M. A. Wörheide, J. Krumsiek, G. Kastenmüller, M. Arnold, Multi-omics integration in biomedical research - A metabolomics-centric review. *Anal. Chim. Acta*. **1141**, 144–162 (2021).

2. GTEx Consortium, The GTEx Consortium atlas of genetic regulatory effects across human tissues. *Science*. **369**, 1318–1330 (2020).

3. M. Arnold, J. Raffler, A. Pfeufer, K. Suhre, G. Kastenmüller, SNiPA: an interactive, genetic variant-centered annotation browser. *Bioinformatics*. **31**, 1334–1336 (2015).

4. S. K. Sieberts, T. M. Perumal, M. M. Carrasquillo, M. Allen, J. S. Reddy, G. E. Hoffman, K. K. Dang, J. Calley, P. J. Ebert, J. Eddy, X. Wang, A. K. Greenwood, S. Mostafavi, CommonMind Consortium (CMC), The AMP-AD Consortium, L. Omberg, M. A. Peters, B. A. Logsdon, P. L. De Jager, N. Ertekin-Taner, L. M. Mangravite, Large eQTL meta-analysis reveals differing patterns between cerebral cortical and cerebellar brain regions. *Sci Data*. **7**, 340 (2020).

5. F. Cunningham, P. Achuthan, W. Akanni, J. Allen, M. R. Amode, I. M. Armean, R. Bennett, J. Bhai, K. Billis, S. Boddu, C. Cummins, C. Davidson, K. J. Dodiya, A. Gall, C. G. Girón, L. Gil, T. Grego, L. Haggerty, E. Haskell, T. Hourlier, O. G. Izuogu, S. H. Janacek, T. Juettemann, M. Kay, M. R. Laird, I. Lavidas, Z. Liu, J. E. Loveland, J. C. Marugán, T. Maurel, A. C. McMahon, B. Moore, J. Morales, J. M. Mudge, M. Nuhn, D. Ogeh, A. Parker, A. Parton, M. Patricio, A. I. Abdul Salam, B. M. Schmitt, H. Schuilenburg, D. Sheppard, H. Sparrow, E. Stapleton, M. Szuba, K. Taylor, G. Threadgold, A. Thormann, A. Vullo, B. Walts, A. Winterbottom, A. Zadissa, M. Chakiachvili, A. Frankish, S. E. Hunt, M. Kostadima, N. Langridge, F. J. Martin, M. Muffato, E. Perry, M. Ruffier, D. M. Staines, S. J. Trevanion, B. L. Aken, A. D. Yates, D. R. Zerbino, P. Flicek, Ensembl 2019. *Nucleic Acids Res.* **47**, D745–D751 (2019).

6. S. Durinck, P. T. Spellman, E. Birney, W. Huber, Mapping identifiers for the integration of genomic datasets with the R/Bioconductor package biomaRt. *Nat. Protoc.* **4**, 1184–1191 (2009).

7. B. Braschi, P. Denny, K. Gray, T. Jones, R. Seal, S. Tweedie, B. Yates, E. Bruford, Genenames.org: the HGNC and VGNC resources in 2019. *Nucleic Acids Res.* **47**, D786–D792 (2019).

8. R. E. Thurman, E. Rynes, R. Humbert, J. Vierstra, M. T. Maurano, E. Haugen, N. C. Sheffield, A. B. Stergachis, H. Wang, B. Vernot, K. Garg, S. John, R. Sandstrom, D. Bates, L. Boatman, T. K. Canfield, M. Diegel, D. Dunn, A. K. Ebersol, T. Frum, E. Giste, A. K. Johnson, E. M. Johnson, T. Kutyavin, B. Lajoie, B.-K. Lee, K. Lee, D. London, D. Lotakis, S. Neph, F. Neri, E. D. Nguyen, H. Qu, A. P. Reynolds, V. Roach, A. Safi, M. E. Sanchez, A. Sanyal, A. Shafer, J. M. Simon, L. Song, S. Vong, M. Weaver, Y. Yan, Z. Zhang, Z. Zhang, B. Lenhard, M. Tewari, M. O. Dorschner, R. S. Hansen, P. A. Navas, G. Stamatoyannopoulos, V. R. Iyer, J. D. Lieb, S. R. Sunyaev, J. M. Akey, P. J. Sabo, R. Kaul, T. S. Furey, J. Dekker, G. E. Crawford, J. A. Stamatoyannopoulos, The accessible chromatin landscape of the human genome. *Nature*. **489**, 75–82 (2012).

9. FANTOM Consortium and the RIKEN PMI and CLST (DGT), A. R. R. Forrest, H. Kawaji, M. Rehli, J. K. Baillie, M. J. L. de Hoon, V. Haberle, T. Lassmann, I. V. Kulakovskiy, M. Lizio, M. Itoh, R. Andersson, C. J. Mungall, T. F. Meehan, S. Schmeier, N. Bertin, M. Jørgensen, E. Dimont, E. Arner, C. Schmidl, U. Schaefer, Y. A. Medvedeva, C. Plessy, M. Vitezic, J. Severin, C. A. Semple, Y. Ishizu, R. S. Young, M. Francescatto, I. Alam, D. Albanese, G. M. Altschuler, T. Arakawa, J. A. C. Archer, P. Arner, M. Babina, S. Rennie, P. J. Balwierz, A. G. Beckhouse, S. Pradhan-Bhatt, J. A. Blake, A. Blumenthal, B. Bodega, A. Bonetti, J. Briggs, F. Brombacher, A. M. Burroughs, A. Califano, C. V. Cannistraci, D. Carbajo, Y. Chen, M. Chierici, Y. Ciani, H. C. Clevers, E. Dalla, C. A. Davis, M. Detmar, A. D. Diehl, T. Dohi, F. Drabløs, A. S. B. Edge, M. Edinger, K. Ekwall, M. Endoh, H. Enomoto, M. Fagiolini, L. Fairbairn, H. Fang, M. C. Farach-Carson, G. J. Faulkner, A. V. Favorov, M. E. Fisher, M. C. Frith, R. Fujita, S. Fukuda, C. Furlanello, M. Furino, J.-I. Furusawa, T. B. Geijtenbeek, A. P. Gibson, T. Gingeras, D. Goldowitz, J. Gough, S. Guhl, R. Guler, S. Gustincich, T. J. Ha, M. Hamaguchi, M. Hara, M. Harbers, J. Harshbarger, A. Hasegawa, Y. Hasegawa, T. Hashimoto, M. Herlyn, K. J. Hitchens, S. J. Ho Sui, O. M. Hofmann, I. Hoof, F. Hori, L. Huminiecki, K. Iida, T. Ikawa, B. R. Jankovic, H. Jia, A. Joshi, G. Jurman, B. Kaczkowski, C. Kai, K. Kaida, A. Kaiho, K. Kajiyama, M. Kanamori-Katayama, A. S. Kasianov, T. Kasukawa, S. Katayama, S. Kato, S. Kawaguchi, H. Kawamoto, Y. I. Kawamura, T. Kawashima, J. S. Kempfle, T. J. Kenna, J. Kere, L. M. Khachigian, T. Kitamura, S. P. Klinken, A. J. Knox, M. Kojima, S. Kojima, N. Kondo, H. Koseki, S. Koyasu, S. Krampitz, A. Kubosaki, A. T. Kwon, J. F. J. Laros, W. Lee, A. Lennartsson, K. Li, B. Lilje, L. Lipovich, A. Mackay-Sim, R.-I. Manabe, J. C. Mar, B. Marchand, A. Mathelier, N. Mejhert, A.

Meynert, Y. Mizuno, D. A. de Lima Morais, H. Morikawa, M. Morimoto, K. Moro, E. Motakis, H. Motohashi, C. L. Mummery, M. Murata, S. Nagao-Sato, Y. Nakachi, F. Nakahara, T. Nakamura, Y. Nakamura, K. Nakazato, E. van Nimwegen, N. Ninomiya, H. Nishiyori, S. Noma, S. Noma, T. Noazaki, S. Ogishima, N. Ohkura, H. Ohimiya, H. Ohno, M. Ohshima, M. Okada-Hatakeyama, Y. Okazaki, V. Orlando, D. A. Ovchinnikov, A. Pain, R. Passier, M. Patrikakis, H. Persson, S. Piazza, J. G. D. Prendergast, O. J. L. Rackham, J. A. Ramilowski, M. Rashid, T. Ravasi, P. Rizzu, M. Roncador, S. Roy, M. B. Rye, E. Saijyo, A. Sajantila, A. Saka, S. Sakaguchi, M. Sakai, H. Sato, S. Savvi, A. Saxena, C. Schneider, E. A. Schultes, G. G. Schulze-Tanzil, A. Schwegmann, T. Sengstag, G. Sheng, H. Shimoji, Y. Shimoni, J. W. Shin, C. Simon, D. Sugiyama, T. Sugiyama, M. Suzuki, N. Suzuki, R. K. Swoboda, P. A. C. ’t Hoen, M. Tagami, N. Takahashi, J. Takai, H. Tanaka, H. Tatsukawa, Z. Tatum, M. Thompson, H. Toyodo, T. Toyoda, E. Valen, M. van de Wetering, L. M. van den Berg, R. Verado, D. Vijayan, I. E. Vorontsov, W. W. Wasserman, S. Watanabe, C. A. Wells, L. N. Winteringham, E. Wolvetang, E. J. Wood, Y. Yamaguchi, M. Yamamoto, M. Yoneda, Y. Yonekura, S. Yoshida, S. E. Zabierowski, P. G. Zhang, X. Zhao, S. Zucchelli, K. M. Summers, H. Suzuki, C. O. Daub, J. Kawai, P. Heutink, W. Hide, T. C. Freeman, B. Lenhard, V. B. Bajic, M. S. Taylor, V. J. Makeev, A. Sandelin, D. A. Hume, P. Carninci, Y. Hayashizaki, A promoter-level mammalian expression atlas. *Nature*. **507**, 462–470 (2014).

10. J. Raffler, N. Friedrich, M. Arnold, T. Kacprowski, R. Rueedi, E. Altmaier, S. Bergmann, K. Budde, C. Gieger, G. Homuth, M. Pietzner, W. Römisch-Margl, K. Strauch, H. Völzke, M. Waldenberger, H. Wallaschofski, M. Nauck, U. Völker, G. Kastenmüller, K. Suhre, Genome-Wide Association Study with Targeted and Non-targeted NMR Metabolomics Identifies 15 Novel Loci of Urinary Human Metabolic Individuality. *PLoS Genet.* **11**, e1005487 (2015).

11. S.-Y. Shin, E. B. Fauman, A.-K. Petersen, J. Krumsiek, R. Santos, J. Huang, M. Arnold, I. Erte, V. Forgetta, T.-P. Yang, K. Walter, C. Menni, L. Chen, L. Vasquez, A. M. Valdes, C. L. Hyde, V. Wang, D. Ziemek, P. Roberts, L. Xi, E. Grundberg, Multiple Tissue Human Expression Resource (MuTHER) Consortium, M. Waldenberger, J. B. Richards, R. P. Mohney, M. V. Milburn, S. L. John, J. Trimmer, F. J. Theis, J. P. Overington, K. Suhre, M. J. Brosnan, C. Gieger, G. Kastenmüller, T. D. Spector, N. Soranzo, An atlas of genetic influences on human blood metabolites. *Nat. Genet.* **46**, 543–550 (2014).

12. K. Suhre, S.-Y. Shin, A.-K. Petersen, R. P. Mohney, D. Meredith, B. Wägele, E. Altmaier, CARDIoGRAM, P. Deloukas, J. Erdmann, E. Grundberg, C. J. Hammond, M. H. de Angelis, G. Kastenmüller, A. Köttgen, F. Kronenberg, M. Mangino, C. Meisinger, T. Meitinger, H.-W. Mewes, M. V. Milburn, C. Prehn, J. Raffler, J. S. Ried, W. Römisch-Margl, N. J. Samani, K. S. Small, H.-E. Wichmann, G. Zhai, T. Illig, T. D. Spector, J. Adamski, N. Soranzo, C. Gieger, Human metabolic individuality in biomedical and pharmaceutical research. *Nature*. **477**, 54–60 (2011).

13. H. H. M. Draisma, R. Pool, M. Kobl, R. Jansen, A.-K. Petersen, A. A. M. Vaarhorst, I. Yet, T. Haller, A. Demirkan, T. Esko, G. Zhu, S. Böhringer, M. Beekman, J. B. van Klinken, W. Römisch-Margl, C. Prehn, J. Adamski, A. J. M. de Craen, E. M. van Leeuwen, N. Amin, H. Dharuri, H.-J. Westra, L. Franke, E. J. C. de Geus, J. J. Hottenga, G. Willemsen, A. K. Henders, G. W. Montgomery, D. R. Nyholt, J. B. Whitfield, B. W. Penninx, T. D. Spector, A. Metspalu, P. E. Slagboom, K. W. van Dijk, P. A. C. ’t Hoen, K. Strauch, N. G. Martin, G.-J. B. van Ommen, T. Illig, J. T. Bell, M. Mangino, K. Suhre, M. I. McCarthy, C. Gieger, A. Isaacs, C. M. van Duijn, D. I. Boomsma, Genome-wide association study identifies novel genetic variants contributing to variation in blood metabolite levels. *Nat. Commun.* **6**, 7208 (2015).

14. T. Long, M. Hicks, H.-C. Yu, W. H. Biggs, E. F. Kirkness, C. Menni, J. Zierer, K. S. Small, M. Mangino, H. Messier, S. Brewerton, Y. Turpaz, B. A. Perkins, A. M. Evans, L. A. D. Miller, L. Guo, C. T. Caskey, N. J. Schork, C. Garner, T. D. Spector, J. C. Venter, A. Telenti, Whole-genome sequencing identifies common-to-rare variants associated with human blood metabolites. *Nat. Genet.* **49**, 568–578 (2017).

15. J. Krumsiek, K. Suhre, A. M. Evans, M. W. Mitchell, R. P. Mohney, M. V. Milburn, B. Wägele, W. Römisch-Margl, T. Illig, J. Adamski, C. Gieger, F. J. Theis, G. Kastenmüller, Mining the unknown: a systems approach to metabolite identification combining genetic and metabolic information. *PLoS Genet.* **8**, e1003005 (2012).

16. K. Suhre, M. Arnold, A. M. Bhagwat, R. J. Cotton, R. Engelke, J. Raffler, H. Sarwath, G. Thareja, A. Wahl, R. K. DeLisle, L. Gold, M. Pezer, G. Lauc, M. A. El-Din Selim, D. O. Mook-Kanamori, E. K. Al-Dous, Y. A. Mohamoud, J. Malek, K. Strauch, H. Grallert, A. Peters, G. Kastenmüller, C. Gieger, J. Graumann, Connecting genetic risk to disease end points through the human blood plasma proteome. *Nat. Commun.* **8**, 14357 (2017).

17. J. C. Lambert, C. A. Ibrahim-Verbaas, D. Harold, A. C. Naj, R. Sims, C. Bellenguez, A. L. DeStafano, J. C. Bis, G. W. Beecham, B. Grenier-Boley, G. Russo, T. A. Thorton-Wells, N. Jones, A. V. Smith, V. Chouraki, C. Thomas, M. A. Ikram, D. Zelenika, B. N. Vardarajan, Y. Kamatani, C. F. Lin, A. Gerrish, H. Schmidt, B. Kunkle, M. L. Dunstan, A. Ruiz, M. T. Bihoreau, S. H. Choi, C. Reitz, F. Pasquier, C. Cruchaga, D. Craig, N. Amin, C. Berr, O. L. Lopez, P. L. De Jager, V. Deramecourt, J. A. Johnston, D. Evans, S. Lovestone, L. Letenneur, F. J. Morón, D. C. Rubinsztein, G. Eiriksdottir, K. Sleegers, A. M. Goate, N. Fiévet, M. W. Huentelman, M. Gill, K. Brown, M. I. Kamboh, L. Keller, P. Barberger-Gateau, B. McGuiness, E. B. Larson, R. Green, A. J. Myers, C. Dufouil, S. Todd, D. Wallon, S. Love, E. Rogaeva, J. Gallacher, P. St George-Hyslop, J. Clarimon, A. Lleo, A. Bayer, D. W. Tsuang, L. Yu, M. Tsolaki, P. Bossù, G. Spalletta, P. Proitsi, J. Collinge, S. Sorbi, F. Sanchez-Garcia, N. C. Fox, J. Hardy, M. C. Deniz Naranjo, P. Bosco, R. Clarke, C. Brayne, D. Galimberti, M. Mancuso, F. Matthews, European Alzheimer's Disease Initiative (EADI), Genetic and Environmental Risk in Alzheimer's Disease, Alzheimer's Disease Genetic Consortium, Cohorts for Heart and Aging Research in Genomic Epidemiology, S. Moebus, P. Mecocci, M. Del Zompo, W. Maier, H. Hampel, A. Pilotto, M. Bullido, F. Panza, P. Caffarra, B. Nacmias, J. R. Gilbert, M. Mayhaus, L. Lannefelt, H. Hakonarson, S. Pichler, M. M. Carrasquillo, M. Ingelsson, D. Beekly, V. Alvarez, F. Zou, O. Valladares, S. G. Younkin, E. Coto, K. L. Hamilton-Nelson, W. Gu, C. Razquin, P. Pastor, I. Mateo, M. J. Owen, K. M. Faber, P. V. Jonsson, O. Combarros, M. C. O'Donovan, L. B. Cantwell, H. Soininen, D. Blacker, S. Mead, T. H. Mosley Jr, D. A. Bennett, T. B. Harris, L. Fratiglioni, C. Holmes, R. F. de Bruijn, P. Passmore, T. J. Montine, K. Bettens, J. I. Rotter, A. Brice, K. Morgan, T. M. Foroud, W. A. Kukull, D. Hannequin, J. F. Powell, M. A. Nalls, K. Ritchie, K. L. Lunetta, J. S. Kauwe, E. Boerwinkle, M. Riemenschneider, M. Boada, M. Hiltuenen, E. R. Martin, R. Schmidt, D. Rujescu, L. S. Wang, J. F. Dartigues, R. Mayeux, C. Tzourio, A. Hofman, M. M. Nöthen, C. Graff, B. M. Psaty, L. Jones, J. L. Haines, P. A. Holmans, M. Lathrop, M. A. Pericak-Vance, L. J. Launer, L. A. Farrer, C. M. van Duijn, C. Van Broeckhoven, V. Moskvina, S. Seshadri, J. Williams, G. D. Schellenberg, P. Amouyel, Meta-analysis of 74,046 individuals identifies 11 new susceptibility loci for Alzheimer's disease. *Nat. Genet.* **45**, 1452–1458 (2013).

18. G. W. Beecham, K. Hamilton, A. C. Naj, E. R. Martin, M. Huentelman, A. J. Myers, J. J. Corneveaux, J. Hardy, J.-P. Vonsattel, S. G. Younkin, D. A. Bennett, P. L. De Jager, E. B. Larson, P. K. Crane, M. I. Kamboh, J. K. Kofler, D. C. Mash, L. Duque, J. R. Gilbert, H. Gwirtsman, J. D. Buxbaum, P. Kramer, D. W. Dickson, L. A. Farrer, M. P. Frosch, B. Ghetti, J. L. Haines, B. T. Hyman, W. A. Kukull, R. P. Mayeux, M. A. Pericak-Vance, J. A. Schneider, J. Q. Trojanowski, E. M. Reiman, Alzheimer's Disease Genetics Consortium (ADGC), G. D. Schellenberg, T. J. Montine, Genome-wide association meta-analysis of neuropathologic features of Alzheimer's disease and related dementias. *PLoS Genet.* **10**, e1004606 (2014).

19. Y. Deming, J. Xia, Y. Cai, J. Lord, P. Holmans, S. Bertelsen, D. Holtzman, J. C. Morris, K. Bales, E. H. Pickering, J. Kauwe, A. Goate, C. Cruchaga, Alzheimer's Disease Neuroimaging Initiative (ADNI), *Neurobiol. Aging*, in press.

20. Y. Deming, Z. Li, M. Kapoor, O. Harari, J. L. Del-Aguila, K. Black, D. Carrell, Y. Cai, M. V. Fernandez, J. Budde, S. Ma, B. Saef, B. Howells, K.-L. Huang, S. Bertelsen, A. M. Fagan, D. M. Holtzman, J. C. Morris, S. Kim, A. J. Saykin, P. L. De Jager, M. Albert, A. Moghekar, R. O'Brien, M. Riemenschneider, R. C. Petersen, K. Blennow, H. Zetterberg, L. Minthon, V. M. Van Deerlin, V. M.-Y. Lee, L. M. Shaw, J. Q. Trojanowski, G. Schellenberg, J. L. Haines, R. Mayeux, M. A. Pericak-Vance, L. A. Farrer, E. R. Peskind, G. Li, A. F. Di Narzo, Alzheimer's Disease Neuroimaging Initiative (ADNI), Alzheimer Disease Genetic Consortium (ADGC), J. S. K. Kauwe, A. M. Goate, C. Cruchaga, Genome-wide association study identifies four novel loci associated with Alzheimer's endophenotypes and disease modifiers. *Acta Neuropathol.* **133**, 839–856 (2017).

21. K.-L. Huang, E. Marcora, A. A. Pimenova, A. F. Di Narzo, M. Kapoor, S. C. Jin, O. Harari, S. Bertelsen, B. P. Fairfax, J. Czajkowski, V. Chouraki, B. Grenier-Boley, C. Bellenguez, Y. Deming, A. McKenzie, T. Raj, A. E. Renton, J. Budde, A. Smith, A. Fitzpatrick, J. C. Bis, A. DeStefano, H. H. H. Adams, M. A. Ikram, S. van der Lee, J. L. Del-Aguila, M. V. Fernandez, L. Ibañez, International Genomics of Alzheimer's Project, Alzheimer's Disease Neuroimaging Initiative, R. Sims, V. Escott-Price, R. Mayeux, J. L. Haines, L. A. Farrer, M. A. Pericak-Vance, J. C. Lambert, C. van Duijn, L. Launer, S. Seshadri, J. Williams, P. Amouyel, G. D. Schellenberg, B. Zhang, I. Borecki, J. S. K. Kauwe, C. Cruchaga, K. Hao, A. M. Goate, A common haplotype lowers PU.1 expression in myeloid cells and delays onset of Alzheimer's disease. *Nat. Neurosci.* **20**, 1052–1061 (2017).

22. R. E. Marioni, S. E. Harris, Q. Zhang, A. F. McRae, S. P. Hagenaars, W. D. Hill, G. Davies, C. W. Ritchie, C. R. Gale, J. M. Starr, A. M. Goate, D. J. Porteous, J. Yang, K. L. Evans, I. J. Deary, N. R. Wray, P. M. Visscher, GWAS on family history of Alzheimer's disease. *Transl. Psychiatry*. **8**, 99 (2018).

23. B. W. Kunkle, B. Grenier-Boley, R. Sims, J. C. Bis, V. Damotte, A. C. Naj, A. Boland, M. Vronskaya, S. J. van der Lee, A. Amlie-Wolf, C. Bellenguez, A. Frizatti, V. Chouraki, E. R. Martin, K. Sleegers, N. Badarinarayan, J. Jakobsdottir, K. L. Hamilton-Nelson, S. Moreno-Grau, R. Olaso, R. Raybould, Y. Chen, A. B. Kuzma, M. Hiltunen, T. Morgan, S. Ahmad, B. N. Vardarajan, J. Epelbaum, P. Hoffmann, M. Boada, G. W. Beecham, J.-G. Garnier, D. Harold, A. L. Fitzpatrick, O. Valladares, M.-L. Moutet, A. Gerrish, A. V. Smith, L. Qu, D. Bacq, N. Denning, X. Jian, Y. Zhao, M. Del Zompo, N. C. Fox, S.-H. Choi, I. Mateo, J. T. Hughes, H. H. Adams, J. Malamon, F. Sanchez-Garcia, Y. Patel, J. A. Brody, B. A. Dombroski, M. C. D. Naranjo, M. Daniilidou, G. Eiriksdottir, S. Mukherjee, D. Wallon, J. Uphill, T. Aspelund, L. B. Cantwell, F. Garzia, D. Galimberti, E. Hofer, M. Butkiewicz, B. Fin, E. Scarpini, C. Sarnowski, W. S. Bush, S. Meslage, J. Kornhuber, C. C. White, Y. Song, R. C. Barber, S. Engelborghs, S. Sordon, D. Voijnovic, P. M. Adams, R. Vandenberghe, M. Mayhaus, L. A. Cupples, M. S. Albert, P. P. De Deyn, W. Gu, J. J. Himali, D. Beekly, A. Squassina, A. M. Hartmann, A. Orellana, D. Blacker, E. Rodriguez-Rodriguez, S. Lovestone, M. E. Garcia, R. S. Doody, C. Munoz-Fernadez, R. Sussams, H. Lin, T. J. Fairchild, Y. A. Benito, C. Holmes, H. Karamujić-Čomić, M. P. Frosch, H. Thonberg, W. Maier, G. Roshchupkin, B. Ghetti, V. Giedraitis, A. Kawalia, S. Li, R. M. Huebinger, L. Kilander, S. Moebus, I. Hernández, M. I. Kamboh, R. Brundin, J. Turton, Q. Yang, M. J. Katz, L. Concari, J. Lord, A. S. Beiser, C. D. Keene, S. Helisalmi, I. Kloszewska, W. A. Kukull, A. M. Koivisto, A. Lynch, L. Tarraga, E. B. Larson, A. Haapasalo, B. Lawlor, T. H. Mosley, R. B. Lipton, V. Solfrizzi, M. Gill, W. T. Longstreth Jr, T. J. Montine, V. Frisardi, M. Diez-Fairen, F. Rivadeneira, R. C. Petersen, V. Deramecourt, I. Alvarez, F. Salani, A. Ciaramella, E. Boerwinkle, E. M. Reiman, N. Fievet, J. I. Rotter, J. S. Reisch, O. Hanon, C. Cupidi, A. G. Andre Uitterlinden, D. R. Royall, C. Dufouil, R. G. Maletta, I. de Rojas, M. Sano, A. Brice, R. Cecchetti, P. S. George-Hyslop, K. Ritchie, M. Tsolaki, D. W. Tsuang, B. Dubois, D. Craig, C.-K. Wu, H. Soininen, D. Avramidou, R. L. Albin, L. Fratiglioni, A. Germanou, L. G. Apostolova, L. Keller, M. Koutroumani, S. E. Arnold, F. Panza, O. Gkatzima, S. Asthana, D. Hannequin, P. Whitehead, C. S. Atwood, P. Caffarra, H. Hampel, I. Quintela, Á. Carracedo, L. Lannfelt, D. C. Rubinsztein, L. L. Barnes, F. Pasquier, L. Frölich, S. Barral, B. McGuinness, T. G. Beach, J. A. Johnston, J. T. Becker, P. Passmore, E. H. Bigio, J. M. Schott, T. D. Bird, J. D. Warren, B. F. Boeve, M. K. Lupton, J. D. Bowen, P. Proitsi, A. Boxer, J. F. Powell, J. R. Burke, J. S. K. Kauwe, J. M. Burns, M. Mancuso, J. D. Buxbaum, U. Bonuccelli, N. J. Cairns, A. McQuillin, C. Cao, G. Livingston, C. S. Carlson, N. J. Bass, C. M. Carlsson, J. Hardy, R. M. Carney, J. Bras, M. M. Carrasquillo, R. Guerreiro, M. Allen, H. C. Chui, E. Fisher, C. Masullo, E. A. Crocco, C. DeCarli, G. Bisceglio, M. Dick, L. Ma, R. Duara, N. R. Graff-Radford, D. A. Evans, A. Hodges, K. M. Faber, M. Scherer, K. B. Fallon, M. Riemenschneider, D. W. Fardo, R. Heun, M. R. Farlow, H. Kölsch, S. Ferris, M. Leber, T. M. Foroud, I. Heuser, D. R. Galasko, I. Giegling, M. Gearing, M. Hüll, D. H. Geschwind, J. R. Gilbert, J. Morris, R. C. Green, K. Mayo, J. H. Growdon, T. Feulner, R. L. Hamilton, L. E. Harrell, D. Drichel, L. S. Honig, T. D. Cushion, M. J. Huentelman, P. Hollingworth, C. M. Hulette, B. T. Hyman, R. Marshall, G. P. Jarvik, A. Meggy, E. Abner, G. E. Menzies, L.-W. Jin, G. Leonenko, L. M. Real, G. R. Jun, C. T. Baldwin, D. Grozeva, A. Karydas, G. Russo, J. A. Kaye, R. Kim, F. Jessen, N. W. Kowall, B. Vellas, J. H. Kramer, E. Vardy, F. M. LaFerla, K.-H. Jöckel, J. J. Lah, M. Dichgans, J. B. Leverenz, D. Mann, A. I. Levey, S. Pickering-Brown, A. P. Lieberman, N. Klopp, K. L. Lunetta, H.-E. Wichmann, C. G. Lyketsos, K. Morgan, D. C. Marson, K. Brown, F. Martiniuk, C. Medway, D. C. Mash, M. M. Nöthen, E. Masliah, N. M. Hooper, W. C. McCormick, A. Daniele, S. M. McCurry, A. Bayer, A. N. McDavid, J. Gallacher, A. C. McKee, H. van den Bussche, M. Mesulam, C. Brayne, B. L. Miller, S. Riedel-Heller, C. A. Miller, J. W. Miller, A. Al-Chalabi, J. C. Morris, C. E. Shaw, A. J. Myers, J. Wiltfang, S. O'Bryant, J. M. Olichney, V. Alvarez, J. E. Parisi, A. B. Singleton, H. L. Paulson, J. Collinge, W. R. Perry, S. Mead, E. Peskind, D. H. Cribbs, M. Rossor, A. Pierce, N. S. Ryan, W. W. Poon, B. Nacmias, H. Potter, S. Sorbi, J. F. Quinn, E. Sacchinelli, A. Raj, G. Spalletta, M. Raskind, C. Caltagirone, P. Bossù, M. D. Orfei, B. Reisberg, R. Clarke, C. Reitz, A. D. Smith, J. M. Ringman, D. Warden, E. D. Roberson, G. Wilcock, E. Rogaeva, A. C. Bruni, H. J. Rosen, M. Gallo, R. N. Rosenberg, Y. Ben-Shlomo, M. A. Sager, P. Mecocci, A. J. Saykin, P. Pastor, M. L. Cuccaro, J. M. Vance, J. A. Schneider, L. S. Schneider, S. Slifer, W. W. Seeley, A. G. Smith, J. A. Sonnen, S. Spina, R. A. Stern, R. H. Swerdlow, M. Tang, R. E. Tanzi, J. Q. Trojanowski, J. C. Troncoso, V. M. Van Deerlin, L. J. Van Eldik, H. V. Vinters, J. P. Vonsattel, S. Weintraub, K. A. Welsh-Bohmer, K. C. Wilhelmsen, J. Williamson, T. S. Wingo, R. L. Woltjer, C. B. Wright, C.-E. Yu, L. Yu, Y. Saba, A. Pilotto, M. J. Bullido, O. Peters, P. K. Crane, D. Bennett, P. Bosco, E. Coto, V. Boccardi, P. L. De Jager, A. Lleo, N. Warner, O. L. Lopez, M. Ingelsson, P. Deloukas, C. Cruchaga, C. Graff, R. Gwilliam, M. Fornage, A. M. Goate, P. Sanchez-Juan, P. G. Kehoe, N. Amin, N. Ertekin-Taner, C. Berr, S. Debette, S. Love, L. J. Launer, S. G. Younkin, J.-F. Dartigues, C. Corcoran, M. A. Ikram, D. W. Dickson, G. Nicolas, D. Campion, J. Tschanz, H. Schmidt, H. Hakonarson, J. Clarimon, R. Munger, R. Schmidt, L. A. Farrer, C. Van Broeckhoven, M. C O'Donovan, A. L. DeStefano, L. Jones, J. L. Haines, J.-F. Deleuze, M. J. Owen, V. Gudnason, R. Mayeux, V. Escott-Price, B. M. Psaty, A. Ramirez, L.-S. Wang, A. Ruiz, C. M. van Duijn, P. A. Holmans, S. Seshadri, J. Williams, P. Amouyel, G. D. Schellenberg, J.-C. Lambert, M. A. Pericak-Vance, Alzheimer Disease Genetics Consortium (ADGC), European Alzheimer's Disease Initiative (EADI), Cohorts for Heart and Aging Research in Genomic Epidemiology Consortium (CHARGE),, Genetic and Environmental Risk in AD/Defining Genetic, Polygenic and Environmental Risk for Alzheimer's Disease Consortium (GERAD/PERADES), Genetic meta-

analysis of diagnosed Alzheimer's disease identifies new risk loci and implicates Aβ, tau, immunity and lipid processing. *Nat. Genet.* **51**, 414–430 (2019).

24. I. E. Jansen, J. E. Savage, K. Watanabe, J. Bryois, D. M. Williams, S. Steinberg, J. Sealock, I. K. Karlsson, S. Hägg, L. Athanasiu, N. Voyle, P. Proitsi, A. Witoelar, S. Stringer, D. Aarsland, I. S. Almdahl, F. Andersen, S. Bergh, F. Bettella, S. Bjornsson, A. Brækhus, G. Bråthen, C. de Leeuw, R. S. Desikan, S. Djurovic, L. Dumitrescu, T. Fladby, T. J. Hohman, P. V. Jonsson, S. J. Kiddle, A. Rongve, I. Saltvedt, S. B. Sando, G. Selbæk, M. Shoai, N. G. Skene, J. Snaedal, E. Stordal, I. D. Ulstein, Y. Wang, L. R. White, J. Hardy, J. Hjerling-Leffler, P. F. Sullivan, W. M. van der Flier, R. Dobson, L. K. Davis, H. Stefansson, K. Stefansson, N. L. Pedersen, S. Ripke, O. A. Andreassen, D. Posthuma, Genome-wide meta-analysis identifies new loci and functional pathways influencing Alzheimer's disease risk. *Nat. Genet.* **51**, 404–413 (2019).

25. D. P. Wightman, I. E. Jansen, J. E. Savage, A. A. Shadrin, S. Bahrami, D. Holland, A. Rongve, S. Børte, B. S. Winsvold, O. K. Drange, A. E. Martinsen, A. H. Skogholt, C. Willer, G. Bråthen, I. Bosnes, J. B. Nielsen, L. G. Fritsche, L. F. Thomas, L. M. Pedersen, M. E. Gabrielsen, M. B. Johnsen, T. W. Meisingset, W. Zhou, P. Proitsi, A. Hodges, R. Dobson, L. Velayudhan, K. Heilbron, A. Auton, 23andMe Research Team, J. M. Sealock, L. K. Davis, N. L. Pedersen, C. A. Reynolds, I. K. Karlsson, S. Magnusson, H. Stefansson, S. Thordardottir, P. V. Jonsson, J. Snaedal, A. Zettergren, I. Skoog, S. Kern, M. Waern, H. Zetterberg, K. Blennow, E. Stordal, K. Hveem, J.-A. Zwart, L. Athanasiu, P. Selnes, I. Saltvedt, S. B. Sando, I. Ulstein, S. Djurovic, T. Fladby, D. Aarsland, G. Selbæk, S. Ripke, K. Stefansson, O. A. Andreassen, D. Posthuma, A genome-wide association study with 1,126,563 individuals identifies new risk loci for Alzheimer's disease. *Nat. Genet.* **53**, 1276–1282 (2021).

26. C. Bellenguez, F. Küçükali, I. E. Jansen, L. Kleineidam, S. Moreno-Grau, N. Amin, A. C. Naj, R. Campos-Martin, B. Grenier-Boley, V. Andrade, P. A. Holmans, A. Boland, V. Damotte, S. J. van der Lee, M. R. Costa, T. Kuulasmaa, Q. Yang, I. de Rojas, J. C. Bis, A. Yaqub, I. Prokic, J. Chapuis, S. Ahmad, V. Giedraitis, D. Aarsland, P. Garcia-Gonzalez, C. Abdelnour, E. Alarcón-Martín, D. Alcolea, M. Alegret, I. Alvarez, V. Álvarez, N. J. Armstrong, A. Tsolaki, C. Antúnez, I. Appollonio, M. Arcaro, S. Archetti, A. A. Pastor, B. Arosio, L. Athanasiu, H. Bailly, N. Banaj, M. Baquero, S. Barral, A. Beiser, A. B. Pastor, J. E. Below, P. Benchek, L. Benussi, C. Berr, C. Besse, V. Bessi, G. Binetti, A. Bizarro, R. Blesa, M. Boada, E. Boerwinkle, B. Borroni, S. Boschi, P. Bossù, G. Bråthen, J. Bressler, C. Bresner, H. Brodaty, K. J. Brookes, L. I. Brusco, D. Buiza-Rueda, K. Bûrger, V. Burholt, W. S. Bush, M. Calero, L. B. Cantwell, G. Chene, J. Chung, M. L. Cuccaro, Á. Carracedo, R. Cecchetti, L. Cervera-Carles, C. Charbonnier, H.-H. Chen, C. Chillotti, S. Ciccone, J. A. H. R. Claassen, C. Clark, E. Conti, A. Corma-Gómez, E. Costantini, C. Custodero, D. Daian, M. C. Dalmasso, A. Daniele, E. Dardiotis, J.-F. Dartigues, P. P. de Deyn, K. de Paiva Lopes, L. D. de Witte, S. Debette, J. Deckert, T. Del Ser, N. Denning, A. DeStefano, M. Dichgans, J. Diehl-Schmid, M. Diez-Fairen, P. D. Rossi, S. Djurovic, E. Duron, E. Düzel, C. Dufouil, G. Eiriksdottir, S. Engelborghs, V. Escott-Price, A. Espinosa, M. Ewers, K. M. Faber, T. Fabrizio, S. F. Nielsen, D. W. Fardo, L. Farotti, C. Fenoglio, M. Fernández-Fuertes, R. Ferrari, C. B. Ferreira, E. Ferri, B. Fin, P. Fischer, T. Fladby, K. Fließbach, B. Fongang, M. Fornage, J. Fortea, T. M. Foroud, S. Fostinelli, N. C. Fox, E. Franco-Macías, M. J. Bullido, A. Frank-García, L. Froelich, B. Fulton-Howard, D. Galimberti, J. M. García-Alberca, P. García-González, S. Garcia-Madrona, G. Garcia-Ribas, R. Ghidoni, I. Giegling, G. Giorgio, A. M. Goate, O. Goldhardt, D. Gomez-Fonseca, A. González-Pérez, C. Graff, G. Grande, E. Green, T. Grimmer, E. Grünblatt, M. Grunin, V. Gudnason, T. Guetta-Baranes, A. Haapasalo, G. Hadjigeorgiou, J. L. Haines, K. L. Hamilton-Nelson, H. Hampel, O. Hanon, J. Hardy, A. M. Hartmann, L. Hausner, J. Harwood, S. Heilmann-Heimbach, S. Helisalmi, M. T. Heneka, I. Hernández, M. J. Herrmann, P. Hoffmann, C. Holmes, H. Holstege, R. H. Vilas, M. Hulsman, J. Humphrey, G. J. Biessels, X. Jian, C. Johansson, G. R. Jun, Y. Kastumata, J. Kauwe, P. G. Kehoe, L. Kilander, A. K. Ståhlbom, M. Kivipelto, A. Koivisto, J. Kornhuber, M. H. Kosmidis, W. A. Kukull, P. P. Kuksa, B. W. Kunkle, A. B. Kuzma, C. Lage, E. J. Laukka, L. Launer, A. Lauria, C.-Y. Lee, J. Lehtisalo, O. Lerch, A. Lleó, W. Longstreth Jr, O. Lopez, A. L. de Munain, S. Love, M. Löwemark, L. Luckcuck, K. L. Lunetta, Y. Ma, J. Macías, C. A. MacLeod, W. Maier, F. Mangialasche, M. Spallazzi, M. Marquié, R. Marshall, E. R. Martin, A. M. Montes, C. M. Rodríguez, C. Masullo, R. Mayeux, S. Mead, P. Mecocci, M. Medina, A. Meggy, S. Mehrabian, S. Mendoza, M. Menéndez-González, P. Mir, S. Moebus, M. Mol, L. Molina-Porcel, L. Montrreal, L. Morelli, F. Moreno, K. Morgan, T. Mosley, M. M. Nöthen, C. Muchnik, S. Mukherjee, B. Nacmias, T. Ngandu, G. Nicolas, B. G. Nordestgaard, R. Olaso, A. Orellana, M. Orsini, G. Ortega, A. Padovani, C. Paolo, G. Papenberg, L. Parnetti, F. Pasquier, P. Pastor, G. Peloso, A. Pérez-Cordón, J. Pérez-Tur, P. Pericard, O. Peters, Y. A. L. Pijnenburg, J. A. Pineda, G. Piñol-Ripoll, C. Pisanu, T. Polak, J. Popp, D. Posthuma, J. Priller, R. Puerta, O. Quenez, I. Quintela, J. Q. Thomassen, A. Rábano, I. Rainero, F. Rajabli, I. Ramakers, L. M. Real, M. J. T. Reinders, C. Reitz, D. Reyes-Dumeyer, P. Ridge, S. Riedel-Heller, P. Riederer, N. Roberto, E. Rodriguez-Rodriguez, A. Rongve, I. R. Allende, M. Rosende-Roca, J. L. Royo, E. Rubino, D. Rujescu, M. E. Sáez, P. Sakka, I. Saltvedt, Á. Sanabria, M. B. Sánchez-Arjona, F. Sanchez-Garcia, P. S. Juan,

R. Sánchez-Valle, S. B. Sando, C. Sarnowski, C. L. Satizabal, M. Scamosci, N. Scarmeas, E. Scarpini, P. Scheltens, N. Scherbaum, M. Scherer, M. Schmid, A. Schneider, J. M. Schott, G. Selbæk, D. Seripa, M. Serrano, J. Sha, A. A. Shadrin, O. Skrobot, S. Slifer, G. J. L. Snijders, H. Soininen, V. Solfrizzi, A. Solomon, Y. Song, S. Sorbi, O. Sotolongo-Grau, G. Spalletta, A. Spottke, A. Squassina, E. Stordal, J. P. Tartan, L. Tárraga, N. Tesí, A. Thalamuthu, T. Thomas, G. Tosto, L. Traykov, L. Tremolizzo, A. Tybjærg-Hansen, A. Uitterlinden, A. Ullgren, I. Ulstein, S. Valero, O. Valladares, C. Van Broeckhoven, J. Vance, B. N. Vardarajan, A. van der Lugt, J. Van Dongen, J. van Rooij, J. van Swieten, R. Vandenberghe, F. Verhey, J.-S. Vidal, J. Vogelgsang, M. Vyhnalek, M. Wagner, D. Wallon, L.-S. Wang, R. Wang, L. Weinhold, J. Wiltfang, G. Windle, B. Woods, M. Yannakoulia, H. Zare, Y. Zhao, X. Zhang, C. Zhu, M. Zulaica, EADB, GR@ACE, DEGESCO, EADI, GERAD, Demgene, FinnGen, ADGC, CHARGE, L. A. Farrer, B. M. Psaty, M. Ghanbari, T. Raj, P. Sachdev, K. Mather, F. Jessen, M. A. Ikram, A. de Mendonça, J. Hort, M. Tsolaki, M. A. Pericak-Vance, P. Amouyel, J. Williams, R. Frikke-Schmidt, J. Clarimon, J.-F. Deleuze, G. Rossi, S. Seshadri, O. A. Andreassen, M. Ingelsson, M. Hiltunen, K. Sleegers, G. D. Schellenberg, C. M. van Duijn, R. Sims, W. M. van der Flier, A. Ruiz, A. Ramirez, J.-C. Lambert, New insights into the genetic etiology of Alzheimer's disease and related dementias. *Nat. Genet.* **54**, 412–436 (2022).

27. Y.-W. Wan, R. Al-Ouran, C. G. Mangleburg, T. M. Perumal, T. V. Lee, K. Allison, V. Swarup, C. C. Funk, C. Gaiteri, M. Allen, M. Wang, S. M. Neuner, C. C. Kaczorowski, V. M. Philip, G. R. Howell, H. Martini-Stoica, H. Zheng, H. Mei, X. Zhong, J. W. Kim, V. L. Dawson, T. M. Dawson, P.-C. Pao, L.-H. Tsai, J.-V. Haure-Mirande, M. E. Ehrlich, P. Chakrabarty, Y. Levites, X. Wang, E. B. Dammer, G. Srivastava, S. Mukherjee, S. K. Sieberts, L. Omberg, K. D. Dang, J. A. Eddy, P. Snyder, Y. Chae, S. Amberkar, W. Wei, W. Hide, C. Preuss, A. Ergun, P. J. Ebert, D. C. Airey, S. Mostafavi, L. Yu, H.-U. Klein, Accelerating Medicines Partnership-Alzheimer's Disease Consortium, G. W. Carter, D. A. Collier, T. E. Golde, A. I. Levey, D. A. Bennett, K. Estrada, T. M. Townsend, B. Zhang, E. Schadt, P. L. De Jager, N. D. Price, N. Ertekin-Taner, Z. Liu, J. M. Shulman, L. M. Mangravite, B. A. Logsdon, Meta-Analysis of the Alzheimer's Disease Human Brain Transcriptome and Functional Dissection in Mouse Models. *Cell Rep.* **32**, 107908 (2020).

28. P. L. De Jager, Y. Ma, C. McCabe, J. Xu, B. N. Vardarajan, D. Felsky, H.-U. Klein, C. C. White, M. A. Peters, B. Lodgson, P. Nejad, A. Tang, L. M. Mangravite, L. Yu, C. Gaiteri, S. Mostafavi, J. A. Schneider, D. A. Bennett, A multi-omic atlas of the human frontal cortex for aging and Alzheimer's disease research. *Sci Data*. **5**, 180142 (2018).

29. M. Allen, M. M. Carrasquillo, C. Funk, B. D. Heavner, F. Zou, C. S. Younkin, J. D. Burgess, H.-S. Chai, J. Crook, J. A. Eddy, H. Li, B. Logsdon, M. A. Peters, K. K. Dang, X. Wang, D. Serie, C. Wang, T. Nguyen, S. Lincoln, K. Malphrus, G. Bisceglio, M. Li, T. E. Golde, L. M. Mangravite, Y. Asmann, N. D. Price, R. C. Petersen, N. R. Graff-Radford, D. W. Dickson, S. G. Younkin, N. Ertekin-Taner, Human whole genome genotype and transcriptome data for Alzheimer's and other neurodegenerative diseases. *Sci Data*. **3**, 160089 (2016).

30. M. Wang, N. D. Beckmann, P. Roussos, E. Wang, X. Zhou, Q. Wang, C. Ming, R. Neff, W. Ma, J. F. Fullard, M. E. Hauberg, J. Bendl, M. A. Peters, B. Logsdon, P. Wang, M. Mahajan, L. M. Mangravite, E. B. Dammer, D. M. Duong, J. J. Lah, N. T. Seyfried, A. I. Levey, J. D. Buxbaum, M. Ehrlich, S. Gandy, P. Katsel, V. Haroutunian, E. Schadt, B. Zhang, The Mount Sinai cohort of large-scale genomic, transcriptomic and proteomic data in Alzheimer's disease. *Sci Data*. **5**, 180185 (2018).

31. R. Batra, M. Arnold, M. A. Wörheide, M. Allen, X. Wang, C. Blach, A. I. Levey, N. T. Seyfried, N. Ertekin-Taner, D. A. Bennett, G. Kastenmüller, R. F. Kaddurah-Daouk, J. Krumsiek, Alzheimer's Disease Metabolomics Consortium (ADMC), The landscape of metabolic brain alterations in Alzheimer's disease. *Alzheimers. Dement.*, 1–19 (2022).

32. E. C. B. Johnson, E. B. Dammer, D. M. Duong, L. Ping, M. Zhou, L. Yin, L. A. Higginbotham, A. Guajardo, B. White, J. C. Troncoso, M. Thambisetty, T. J. Montine, E. B. Lee, J. Q. Trojanowski, T. G. Beach, E. M. Reiman, V. Haroutunian, M. Wang, E. Schadt, B. Zhang, D. W. Dickson, N. Ertekin-Taner, T. E. Golde, V. A. Petyuk, P. L. De Jager, D. A. Bennett, T. S. Wingo, S. Rangaraju, I. Hajjar, J. M. Shulman, J. J. Lah, A. I. Levey, N. T. Seyfried, Large-scale proteomic analysis of Alzheimer's disease brain and cerebrospinal fluid reveals early changes in energy metabolism associated with microglia and astrocyte activation. *Nat. Med.* **26**, 769–780 (2020).

33. E. C. B. Johnson, E. K. Carter, E. B. Dammer, D. M. Duong, E. S. Gerasimov, Y. Liu, J. Liu, R. Betarbet, L. Ping, L. Yin, G. E. Serrano, T. G. Beach, J. Peng, P. L. De Jager, V. Haroutunian, B. Zhang, C. Gaiteri, D. A. Bennett, M. Gearing, T. S. Wingo, A. P. Wingo, J. J. Lah, A. I. Levey, N. T. Seyfried, Large-scale deep multi-layer

analysis of Alzheimer's disease brain reveals strong proteomic disease-related changes not observed at the RNA level. *Nat. Neurosci.* **25**, 213–225 (2022).

34. S. MahmoudianDehkordi, M. Arnold, K. Nho, S. Ahmad, W. Jia, G. Xie, G. Louie, A. Kueider-Paisley, M. A. Moseley, J. W. Thompson, L. St John Williams, J. D. Tenenbaum, C. Blach, R. Baillie, X. Han, S. Bhattacharyya, J. B. Toledo, S. Schafferer, S. Klein, T. Koal, S. L. Risacher, M. A. Kling, A. Motsinger-Reif, D. M. Rotroff, J. Jack, T. Hankemeier, D. A. Bennett, P. L. De Jager, J. Q. Trojanowski, L. M. Shaw, M. W. Weiner, P. M. Doraiswamy, C. M. van Duijn, A. J. Saykin, G. Kastenmüller, R. Kaddurah-Daouk, Alzheimer's Disease Neuroimaging Initiative and the Alzheimer Disease Metabolomics Consortium, Altered bile acid profile associates with cognitive impairment in Alzheimer's disease-An emerging role for gut microbiome. *Alzheimers. Dement.* **15**, 76–92 (2019).

35. K. Nho, A. Kueider-Paisley, S. MahmoudianDehkordi, M. Arnold, S. L. Risacher, G. Louie, C. Blach, R. Baillie, X. Han, G. Kastenmüller, W. Jia, G. Xie, S. Ahmad, T. Hankemeier, C. M. van Duijn, J. Q. Trojanowski, L. M. Shaw, M. W. Weiner, P. M. Doraiswamy, A. J. Saykin, R. Kaddurah-Daouk, Alzheimer's Disease Neuroimaging Initiative and the Alzheimer Disease Metabolomics Consortium, Altered bile acid profile in mild cognitive impairment and Alzheimer's disease: Relationship to neuroimaging and CSF biomarkers. *Alzheimers. Dement.* **15**, 232–244 (2019).

36. M. Arnold, K. Nho, A. Kueider-Paisley, T. Massaro, K. Huynh, B. Brauner, S. MahmoudianDehkordi, G. Louie, M. A. Moseley, J. W. Thompson, L. S. John-Williams, J. D. Tenenbaum, C. Blach, R. Chang, R. D. Brinton, R. Baillie, X. Han, J. Q. Trojanowski, L. M. Shaw, R. Martins, M. W. Weiner, E. Trushina, J. B. Toledo, P. J. Meikle, D. A. Bennett, J. Krumsiek, P. M. Doraiswamy, A. J. Saykin, R. Kaddurah-Daouk, G. Kastenmüller, Sex and APOE ε4 genotype modify the Alzheimer's disease serum metabolome. *Nat. Commun.* **11**, 1148 (2020).

37. J. B. Toledo, M. Arnold, G. Kastenmüller, R. Chang, R. A. Baillie, X. Han, M. Thambisetty, J. D. Tenenbaum, K. Suhre, J. W. Thompson, L. S. John-Williams, S. MahmoudianDehkordi, D. M. Rotroff, J. R. Jack, A. Motsinger-Reif, S. L. Risacher, C. Blach, J. E. Lucas, T. Massaro, G. Louie, H. Zhu, G. Dallmann, K. Klavins, T. Koal, S. Kim, K. Nho, L. Shen, R. Casanova, S. Varma, C. Legido-Quigley, M. A. Moseley, K. Zhu, M. Y. R. Henrion, S. J. van der Lee, A. C. Harms, A. Demirkan, T. Hankemeier, C. M. van Duijn, J. Q. Trojanowski, L. M. Shaw, A. J. Saykin, M. W. Weiner, P. M. Doraiswamy, R. Kaddurah-Daouk, Alzheimer's Disease Neuroimaging Initiative and the Alzheimer Disease Metabolomics Consortium, Metabolic network failures in Alzheimer's disease: A biochemical road map. *Alzheimers. Dement.* **13**, 965–984 (2017).

38. M. W. Weiner, D. P. Veitch, P. S. Aisen, L. A. Beckett, N. J. Cairns, R. C. Green, D. Harvey, C. R. Jack, W. Jagust, J. C. Morris, R. C. Petersen, J. Salazar, A. J. Saykin, L. M. Shaw, A. W. Toga, J. Q. Trojanowski, The Alzheimer's Disease Neuroimaging Initiative 3: Continued innovation for clinical trial improvement. *Alzheimer's and Dementia*. **13** (2017), pp. 561–571.

39. C. Ballard, D. Aarsland, J. Cummings, J. O'Brien, R. Mills, J. L. Molinuevo, T. Fladby, G. Williams, P. Doherty, A. Corbett, J. Sultana, Drug repositioning and repurposing for Alzheimer disease. *Nat. Rev. Neurol.* **16**, 661–673 (2020).

40. W. J. Strittmatter, K. H. Weisgraber, D. Y. Huang, L. M. Dong, G. S. Salvesen, M. Pericak-Vance, D. Schmechel, A. M. Saunders, D. Goldgaber, A. D. Roses, Binding of human apolipoprotein E to synthetic amyloid beta peptide: isoform-specific effects and implications for late-onset Alzheimer disease. *Proc. Natl. Acad. Sci. U. S. A.* **90**, 8098–8102 (1993).

41. J.-C. Lambert, S. Heath, G. Even, D. Campion, K. Sleegers, M. Hiltunen, O. Combarros, D. Zelenika, M. J. Bullido, B. Tavernier, L. Letenneur, K. Bettens, C. Berr, F. Pasquier, N. Fiévet, P. Barberger-Gateau, S. Engelborghs, P. De Deyn, I. Mateo, A. Franck, S. Helisalmi, E. Porcellini, O. Hanon, European Alzheimer's Disease Initiative Investigators, M. M. de Pancorbo, C. Lendon, C. Dufouil, C. Jaillard, T. Leveillard, V. Alvarez, P. Bosco, M. Mancuso, F. Panza, B. Nacmias, P. Bossù, P. Piccardi, G. Annoni, D. Seripa, D. Galimberti, D. Hannequin, F. Licastro, H. Soininen, K. Ritchie, H. Blanché, J.-F. Dartigues, C. Tzourio, I. Gut, C. Van Broeckhoven, A. Alpérovitch, M. Lathrop, P. Amouyel, Genome-wide association study identifies variants at CLU and CR1 associated with Alzheimer's disease. *Nat. Genet.* **41**, 1094–1099 (2009).

42. D. Harold, R. Abraham, P. Hollingworth, R. Sims, A. Gerrish, M. L. Hamshere, J. S. Pahwa, V. Moskvina, K. Dowzell, A. Williams, N. Jones, C. Thomas, A. Stretton, A. R. Morgan, S. Lovestone, J. Powell, P. Proitsi, M. K. Lupton, C. Brayne, D. C. Rubinsztein, M. Gill, B. Lawlor, A. Lynch, K. Morgan, K. S. Brown, P. A. Passmore, D. Craig, B. McGuinness, S. Todd, C. Holmes, D. Mann, A. D. Smith, S. Love, P. G. Kehoe, J. Hardy, S. Mead, N. Fox, M. Rossor, J. Collinge, W. Maier, F. Jessen, B. Schürmann, R. Heun, H. van den Bussche, I. Heuser, J.

Kornhuber, J. Wiltfang, M. Dichgans, L. Frölich, H. Hampel, M. Hüll, D. Rujescu, A. M. Goate, J. S. K. Kauwe, C. Cruchaga, P. Nowotny, J. C. Morris, K. Mayo, K. Sleegers, K. Bettens, S. Engelborghs, P. P. De Deyn, C. Van Broeckhoven, G. Livingston, N. J. Bass, H. Gurling, A. McQuillin, R. Gwilliam, P. Deloukas, A. Al-Chalabi, C. E. Shaw, M. Tsolaki, A. B. Singleton, R. Guerreiro, T. W. Mühleisen, M. M. Nöthen, S. Moebus, K.-H. Jöckel, N. Klopp, H.-E. Wichmann, M. M. Carrasquillo, V. S. Pankratz, S. G. Younkin, P. A. Holmans, M. O'Donovan, M. J. Owen, J. Williams, Genome-wide association study identifies variants at CLU and PICALM associated with Alzheimer's disease. *Nat. Genet.* **41**, 1088–1093 (2009).

43. Y.-W. Wan, R. Al-Ouran, C. G. Mangleburg, T. M. Perumal, T. V. Lee, K. Allison, V. Swarup, C. C. Funk, C. Gaiteri, M. Allen, M. Wang, S. M. Neuner, C. C. Kaczorowski, V. M. Philip, G. R. Howell, H. Martini-Stoica, H. Zheng, H. Mei, X. Zhong, J. W. Kim, V. L. Dawson, T. M. Dawson, P.-C. Pao, L.-H. Tsai, J.-V. Haure-Mirande, M. E. Ehrlich, P. Chakrabarty, Y. Levites, X. Wang, E. B. Dammer, G. Srivastava, S. Mukherjee, S. K. Sieberts, L. Omberg, K. D. Dang, J. A. Eddy, P. Snyder, Y. Chae, S. Amberkar, W. Wei, W. Hide, C. Preuss, A. Ergun, P. J. Ebert, D. C. Airey, S. Mostafavi, L. Yu, H.-U. Klein, Accelerating Medicines Partnership-Alzheimer's Disease Consortium, G. W. Carter, D. A. Collier, T. E. Golde, A. I. Levey, D. A. Bennett, K. Estrada, T. M. Townsend, B. Zhang, E. Schadt, P. L. De Jager, N. D. Price, N. Ertekin-Taner, Z. Liu, J. M. Shulman, L. M. Mangravite, B. A. Logsdon, Meta-Analysis of the Alzheimer's Disease Human Brain Transcriptome and Functional Dissection in Mouse Models. *Cell Rep.* **32**, 107908 (2020).

44. M. V. Kuleshov, M. R. Jones, A. D. Rouillard, N. F. Fernandez, Q. Duan, Z. Wang, S. Koplev, S. L. Jenkins, K. M. Jagodnik, A. Lachmann, M. G. McDermott, C. D. Monteiro, G. W. Gundersen, A. Ma'ayan, Enrichr: a comprehensive gene set enrichment analysis web server 2016 update. *Nucleic Acids Res.* **44**, W90-7 (2016).

45. J. Bauzon, G. Lee, J. Cummings, Repurposed agents in the Alzheimer's disease drug development pipeline. *Alzheimers. Res. Ther.* **12**, 98 (2020).

46. J. Cummings, Y. Zhou, G. Lee, K. Zhong, J. Fonseca, F. Cheng, Alzheimer's disease drug development pipeline: 2023. *Alzheimers Dement. (N. Y.)* **9** (2023).

47. I. Hajjar, M. Okafor, D. McDaniel, M. Obideen, E. Dee, M. Shokouhi, A. A. Quyyumi, A. Levey, F. Goldstein, Effects of Candesartan vs Lisinopril on Neurocognitive Function in Older Adults With Executive Mild Cognitive Impairment: A Randomized Clinical Trial. *JAMA Netw Open* **3**, e2012252 (2020).

48. I. Hajjar, M. Hart, Y.-L. Chen, W. Mack, W. Milberg, H. Chui, L. Lipsitz, Effect of antihypertensive therapy on cognitive function in early executive cognitive impairment: a double-blind randomized clinical trial. *Arch. Intern. Med.* **172**, 442–444 (2012).

49. J. Cummings, G. Lee, A. Ritter, M. Sabbagh, K. Zhong, Alzheimer's disease drug development pipeline: 2020. *Alzheimers. Dement.* **6**, e12050 (2020).

50. S. Lehrer, P. H. Rheinstein, Alzheimer's disease and intranasal fluticasone propionate in the FDA MedWatch adverse events database. *J. Alzheimers Dis. Rep.* **2**, 111–115 (2018).

51. J. Fang, P. Zhang, Q. Wang, C.-W. Chiang, Y. Zhou, Y. Hou, J. Xu, R. Chen, B. Zhang, S. J. Lewis, J. B. Leverenz, A. A. Pieper, B. Li, L. Li, J. Cummings, F. Cheng, Artificial intelligence framework identifies candidate targets for drug repurposing in Alzheimer's disease. *Alzheimers. Res. Ther.* **14**, 7 (2022).

52. J. Xu, P. Zhang, Y. Huang, Y. Zhou, Y. Hou, L. M. Bekris, J. Lathia, C.-W. Chiang, L. Li, A. A. Pieper, J. B. Leverenz, J. Cummings, F. Cheng, Multimodal single-cell/nucleus RNA sequencing data analysis uncovers molecular networks between disease-associated microglia and astrocytes with implications for drug repurposing in Alzheimer's disease. *Genome Res.* **31**, 1900–1912 (2021).

53. A. S. Tang, T. Oskotsky, S. Havaldar, W. G. Mantyh, M. Bicak, C. W. Solsberg, S. Woldemariam, B. Zeng, Z. Hu, B. Oskotsky, D. Dubal, I. E. Allen, B. S. Glicksberg, M. Sirota, Deep phenotyping of Alzheimer's disease leveraging electronic medical records identifies sex-specific clinical associations. *Nat. Commun.* **13**, 675 (2022).

54. C. Zang, H. Zhang, J. Xu, H. Zhang, S. Fouladvand, S. Havaldar, F. Cheng, K. Chen, Y. Chen, B. S. Glicksberg, J. Chen, J. Bian, F. Wang, High-throughput target trial emulation for Alzheimer's disease drug repurposing with real-world data. *Nat. Commun.* **14**, 8180 (2023).

55. A. O. Sodero, F. J. Barrantes, Pleiotropic effects of statins on brain cells. *Biochim. Biophys. Acta Biomembr.* **1862**, 183340 (2020).

56. J. K. Liao, U. Laufs, Pleiotropic effects of statins. *Annu. Rev. Pharmacol. Toxicol.* **45**, 89–118 (2005).

57. M. D. M. Haag, A. Hofman, P. J. Koudstaal, B. H. C. Stricker, M. M. B. Breteler, Statins are associated with a reduced risk of Alzheimer disease regardless of lipophilicity. The Rotterdam Study. *J. Neurol. Neurosurg. Psychiatry* **80**, 13–17 (2009).

58. G. Torrandell-Haro, G. L. Branigan, F. Vitali, N. Geifman, J. M. Zissimopoulos, R. D. Brinton, Statin therapy and risk of Alzheimer's and age-related neurodegenerative diseases. *Alzheimers. Dement.* **6**, e12108 (2020).

59. C.-S. Chu, P.-T. Tseng, B. Stubbs, T.-Y. Chen, C.-H. Tang, D.-J. Li, W.-C. Yang, Y.-W. Chen, C.-K. Wu, N. Veronese, A. F. Carvalho, B. S. Fernandes, N. Herrmann, P.-Y. Lin, Use of statins and the risk of dementia and mild cognitive impairment: A systematic review and meta-analysis. *Sci. Rep.* **8**, 5804 (2018).

60. J. M. Zissimopoulos, D. Barthold, R. D. Brinton, G. Joyce, Sex and Race Differences in the Association Between Statin Use and the Incidence of Alzheimer Disease. *JAMA Neurol.* **74**, 225–232 (2017).

61. N. Geifman, R. D. Brinton, R. E. Kennedy, L. S. Schneider, A. J. Butte, Evidence for benefit of statins to modify cognitive decline and risk in Alzheimer's disease. *Alzheimers. Res. Ther.* **9**, 10 (2017).

62. B. G. Schultz, D. K. Patten, D. J. Berlau, The role of statins in both cognitive impairment and protection against dementia: a tale of two mechanisms. *Transl. Neurodegener.* **7**, 5 (2018).

63. D. S. Wishart, Y. D. Feunang, A. C. Guo, E. J. Lo, A. Marcu, J. R. Grant, T. Sajed, D. Johnson, C. Li, Z. Sayeeda, N. Assempour, I. Iynkkaran, Y. Liu, A. Maciejewski, N. Gale, A. Wilson, L. Chin, R. Cummings, D. Le, A. Pon, C. Knox, M. Wilson, DrugBank 5.0: a major update to the DrugBank database for 2018. *Nucleic Acids Res.* **46**, D1074–D1082 (2018).

64. B. Zhang, C. Gaiteri, L.-G. Bodea, Z. Wang, J. McElwee, A. A. Podtelezhnikov, C. Zhang, T. Xie, L. Tran, R. Dobrin, E. Fluder, B. Clurman, S. Melquist, M. Narayanan, C. Suver, H. Shah, M. Mahajan, T. Gillis, J. Mysore, M. E. MacDonald, J. R. Lamb, D. A. Bennett, C. Molony, D. J. Stone, V. Gudnason, A. J. Myers, E. E. Schadt, H. Neumann, J. Zhu, V. Emilsson, Integrated systems approach identifies genetic nodes and networks in late-onset Alzheimer's disease. *Cell* **153**, 707–720 (2013).

65. B. L. Walling, M. Kim, LFA-1 in T Cell Migration and Differentiation. *Front. Immunol.* **9**, 952 (2018).

66. G. W. Beecham, K. Hamilton, A. C. Naj, E. R. Martin, M. Huentelman, A. J. Myers, J. J. Corneveaux, J. Hardy, J.-P. Vonsattel, S. G. Younkin, D. A. Bennett, P. L. De Jager, E. B. Larson, P. K. Crane, M. I. Kamboh, J. K. Kofler, D. C. Mash, L. Duque, J. R. Gilbert, H. Gwirtsman, J. D. Buxbaum, P. Kramer, D. W. Dickson, L. A. Farrer, M. P. Frosch, B. Ghetti, J. L. Haines, B. T. Hyman, W. A. Kukull, R. P. Mayeux, M. A. Pericak-Vance, J. A. Schneider, J. Q. Trojanowski, E. M. Reiman, Alzheimer's Disease Genetics Consortium (ADGC), G. D. Schellenberg, T. J. Montine, Genome-wide association meta-analysis of neuropathologic features of Alzheimer's disease and related dementias. *PLoS Genet.* **10**, e1004606 (2014).

67. B. W. Kunkle, B. Grenier-Boley, R. Sims, J. C. Bis, V. Damotte, A. C. Naj, A. Boland, M. Vronskaya, S. J. van der Lee, A. Amlie-Wolf, C. Bellenguez, A. Frizatti, V. Chouraki, E. R. Martin, K. Sleegers, N. Badarinarayan, J. Jakobsdottir, K. L. Hamilton-Nelson, S. Moreno-Grau, R. Olaso, R. Raybould, Y. Chen, A. B. Kuzma, M. Hiltunen, T. Morgan, S. Ahmad, B. N. Vardarajan, J. Epelbaum, P. Hoffmann, M. Boada, G. W. Beecham, J. G. Garnier, D. Harold, A. L. Fitzpatrick, O. Valladares, M. L. Moutet, A. Gerrish, A. V. Smith, L. Qu, D. Bacq, N. Denning, X. Jian, Y. Zhao, M. Del Zompo, N. C. Fox, S. H. Choi, I. Mateo, J. T. Hughes, H. H. Adams, J. Malamon, F. Sanchez-Garcia, Y. Patel, J. A. Brody, B. A. Dombroski, M. C. D. Naranjo, M. Daniilidou, G. Eiriksdottir, S. Mukherjee, D. Wallon, J. Uphill, T. Aspelund, L. B. Cantwell, F. Garzia, D. Galimberti, E. Hofer, M. Butkiewicz, B. Fin, E. Scarpini, C. Sarnowski, W. S. Bush, S. Meslage, J. Kornhuber, C. C. White, Y. Song, R. C. Barber, S. Engelborghs, S. Sordon, D. Voijnovic, P. M. Adams, R. Vandenberghe, M. Mayhaus, L. A. Cupples, M. S. Albert, P. P. De Deyn, W. Gu, J. J. Himali, D. Beekly, A. Squassina, A. M. Hartmann, A. Orellana, D. Blacker, E. Rodriguez-Rodriguez, S. Lovestone, M. E. Garcia, R. S. Doody, C. Munoz-Fernadez, R. Sussams, H. Lin, T. J. Fairchild, Y. A. Benito, C. Holmes, H. Karamujić-Čomić, M. P. Frosch, H. Thonberg, W. Maier, G. Roschupkin, B. Ghetti, V. Giedraitis, A. Kawalia, S. Li, R. M. Huebinger, L. Kilander, S. Moebus, I. Hernández, M. I. Kamboh, R. M. Brundin, J. Turton, Q. Yang, M. J. Katz, L. Concari, J. Lord, A. S. Beiser, C. D. Keene, S. Helisalmi, I. Kloszewska, W. A. Kukull, A. M. Koivisto, A. Lynch, L. Tarraga, E. B. Larson, A. Haapasalo, B. Lawlor, T. H. Mosley, R. B. Lipton, V. Solfrizzi, M. Gill, W. T. Longstreth, T. J. Montine, V. Frisardi, M. Diez-Fairen, F. Rivadeneira, R. C. Petersen, V. Deramecourt, I. Alvarez, F. Salani, A. Ciaramella, E. Boerwinkle, E. M. Reiman, N. Fievet, J. I. Rotter, J. S. Reisch, O. Hanon, C. Cupidi, A. G. Andre Uitterlinden, D. R. Royall, C. Dufouil, R. G. Maletta, I. de Rojas, M. Sano, A. Brice, R. Cecchetti, P. S. George-Hyslop, K. Ritchie, M. Tsolaki, D. W. Tsuang, B. Dubois, D. Craig, C. K. Wu, H. Soininen, D. Avramidou, R. L. Albin, L. Fratiglioni, A. Germanou, L. G. Apostolova, L. Keller, M. Koutroumani, S. E. Arnold, F. Panza, O. Gkatzima, S. Asthana, D. Hannequin, P. Whitehead, C. S. Atwood, P. Caffarra, H. Hampel, I. Quintela, Á. Carracedo, L. Lannfelt, D. C. Rubinsztein, L. L. Barnes, F. Pasquier, L. Frölich, S. Barral, B. McGuinness, T. G. Beach, J. A. Johnston, J. T. Becker, P. Passmore, E. H. Bigio, J. M. Schott, T. D. Bird, J. D. Warren, B. F. Boeve, M. K. Lupton, J. D. Bowen, P. Proitsi, A. Boxer, J. F. Powell, J. R. Burke, J. S. K. Kauwe, J. M. Burns, M. Mancuso, J. D. Buxbaum, U. Bonuccelli, N. J. Cairns, A. McQuillin, C. Cao, G. Livingston, C. S. Carlson, N. J. Bass, C. M. Carlsson, J. Hardy, R. M. Carney, J. Bras, M. M. Carrasquillo, R. Guerreiro, M. Allen, H. C. Chui, E.

Fisher, C. Masullo, E. A. Crocco, C. DeCarli, G. Bisceglio, M. Dick, L. Ma, R. Duara, N. R. Graff-Radford, D. A. Evans, A. Hodges, K. M. Faber, M. Scherer, K. B. Fallon, M. Riemenschneider, D. W. Fardo, R. Heun, M. R. Farlow, H. Kölsch, S. Ferris, M. Leber, T. M. Foroud, I. Heuser, D. R. Galasko, I. Giegling, M. Gearing, M. Hüll, D. H. Geschwind, J. R. Gilbert, J. Morris, R. C. Green, K. Mayo, J. H. Growdon, T. Feulner, R. L. Hamilton, L. E. Harrell, D. Drichel, L. S. Honig, T. D. Cushion, M. J. Huentelman, P. Hollingworth, C. M. Hulette, B. T. Hyman, R. Marshall, G. P. Jarvik, A. Meggy, E. Abner, G. E. Menzies, L. W. Jin, G. Leonenko, L. M. Real, G. R. Jun, C. T. Baldwin, D. Grozeva, A. Karydas, G. Russo, J. A. Kaye, R. Kim, F. Jessen, N. W. Kowall, B. Vellas, J. H. Kramer, E. Vardy, F. M. LaFerla, K. H. Jöckel, J. J. Lah, M. Dichgans, J. B. Leverenz, D. Mann, A. I. Levey, S. Pickering-Brown, A. P. Lieberman, N. Klopp, K. L. Lunetta, H. E. Wichmann, C. G. Lyketsos, K. Morgan, D. C. Marson, K. Brown, F. Martiniuk, C. Medway, D. C. Mash, M. M. Nöthen, E. Masliah, N. M. Hooper, W. C. McCormick, A. Daniele, S. M. McCurry, A. Bayer, A. N. McDavid, J. Gallacher, A. C. McKee, H. van den Bussche, M. Mesulam, C. Brayne, B. L. Miller, S. Riedel-Heller, C. A. Miller, J. W. Miller, A. Al-Chalabi, J. C. Morris, C. E. Shaw, A. J. Myers, J. Wiltfang, S. O'Bryant, J. M. Olichney, V. Alvarez, J. E. Parisi, A. B. Singleton, H. L. Paulson, J. Collinge, W. R. Perry, S. Mead, E. Peskind, D. H. Cribbs, M. Rossor, A. Pierce, N. S. Ryan, W. W. Poon, B. Nacmias, H. Potter, S. Sorbi, J. F. Quinn, E. Sacchinelli, A. Raj, G. Spalletta, M. Raskind, C. Caltagirone, P. Bossù, M. D. Orfei, B. Reisberg, R. Clarke, C. Reitz, A. D. Smith, J. M. Ringman, D. Warden, E. D. Roberson, G. Wilcock, E. Rogaeva, A. C. Bruni, H. J. Rosen, M. Gallo, R. N. Rosenberg, Y. Ben-Shlomo, M. A. Sager, P. Mecocci, A. J. Saykin, P. Pastor, M. L. Cuccaro, J. M. Vance, J. A. Schneider, L. S. Schneider, S. Slifer, W. W. Seeley, A. G. Smith, J. A. Sonnen, S. Spina, R. A. Stern, R. H. Swerdlow, M. Tang, R. E. Tanzi, J. Q. Trojanowski, J. C. Troncoso, V. M. Van Deerlin, L. J. Van Eldik, H. V. Vinters, J. P. Vonsattel, S. Weintraub, K. A. Welsh-Bohmer, K. C. Wilhelmsen, J. Williamson, T. S. Wingo, R. L. Woltjer, C. B. Wright, C. E. Yu, L. Yu, Y. Saba, A. Pilotto, M. J. Bullido, O. Peters, P. K. Crane, D. Bennett, P. Bosco, E. Coto, V. Boccardi, P. L. De Jager, A. Lleo, N. Warner, O. L. Lopez, M. Ingelsson, P. Deloukas, C. Cruchaga, C. Graff, R. Gwilliam, M. Fornage, A. M. Goate, P. Sanchez-Juan, P. G. Kehoe, N. Amin, N. Ertekin-Taner, C. Berr, S. Debette, S. Love, L. J. Launer, S. G. Younkin, J. F. Dartigues, C. Corcoran, M. A. Ikram, D. W. Dickson, G. Nicolas, D. Campion, J. A. Tschanz, H. Schmidt, H. Hakonarson, J. Clarimon, R. Munger, R. Schmidt, L. A. Farrer, C. Van Broeckhoven, M. C. O'Donovan, A. L. DeStefano, L. Jones, J. L. Haines, J. F. Deleuze, M. J. Owen, V. Gudnason, R. Mayeux, V. Escott-Price, B. M. Psaty, A. Ramirez, L. S. Wang, A. Ruiz, C. M. van Duijn, P. A. Holmans, S. Seshadri, J. Williams, P. Amouyel, G. D. Schellenberg, J. C. Lambert, M. A. Pericak-Vance, Genetic meta-analysis of diagnosed Alzheimer's disease identifies new risk loci and implicates Aβ, tau, immunity and lipid processing. *Nat. Genet.* **51**, 414–430 (2019).

68. J.-V. Haure-Mirande, M. Audrain, T. Fanutza, S. H. Kim, W. L. Klein, C. Glabe, B. Readhead, J. T. Dudley, R. D. Blitzer, M. Wang, B. Zhang, E. E. Schadt, S. Gandy, M. E. Ehrlich, Deficiency of TYROBP, an adapter protein for TREM2 and CR3 receptors, is neuroprotective in a mouse model of early Alzheimer's pathology. *Acta Neuropathol.* **134**, 769–788 (2017).

69. G. Weitz-Schmidt, K. Welzenbach, V. Brinkmann, T. Kamata, J. Kallen, C. Bruns, S. Cottens, Y. Takada, U. Hommel, Statins selectively inhibit leukocyte function antigen-1 by binding to a novel regulatory integrin site. *Nat. Med.* **7**, 687–692 (2001).

70. E. Zenaro, E. Pietronigro, V. Della Bianca, G. Piacentino, L. Marongiu, S. Budui, E. Turano, B. Rossi, S. Angiari, S. Dusi, A. Montresor, T. Carlucci, S. Nanì, G. Tosadori, L. Calciano, D. Catalucci, G. Berton, B. Bonetti, G. Constantin, Neutrophils promote Alzheimer's disease-like pathology and cognitive decline via LFA-1 integrin. *Nat. Med.* **21**, 880–886 (2015).

71. J.-C. Jiang, C. Hu, A. M. McIntosh, S. Shah, Investigating the potential anti-depressive mechanisms of statins: a transcriptomic and Mendelian randomization analysis. *Transl. Psychiatry* **13**, 110 (2023).

72. M. Sano, K. L. Bell, D. Galasko, J. E. Galvin, R. G. Thomas, C. H. van Dyck, P. S. Aisen, A randomized, double-blind, placebo-controlled trial of simvastatin to treat Alzheimer disease. *Neurology* **77**, 556–563 (2011).

73. H. H. Feldman, R. S. Doody, M. Kivipelto, D. L. Sparks, D. D. Waters, R. W. Jones, E. Schwam, R. Schindler, J. Hey-Hadavi, D. A. DeMicco, A. Breazna, On behalf of the LEADe Investigators, Randomized controlled trial of atorvastatin in mild to moderate Alzheimer disease: LEADe. *Neurology* **74**, 956–964 (2010).

74. C. Yan, M. E. Grabowska, A. L. Dickson, B. Li, Z. Wen, D. M. Roden, C. Michael Stein, P. J. Embí, J. F. Peterson, Q. Feng, B. A. Malin, W.-Q. Wei, Leveraging generative AI to prioritize drug repurposing candidates for Alzheimer's disease with real-world clinical validation. *NPJ Digit. Med.* **7**, 46 (2024).

75. P. Baloni, M. Arnold, L. Buitrago, K. Nho, H. Moreno, K. Huynh, B. Brauner, G. Louie, A. Kueider-Paisley, K. Suhre, A. J. Saykin, K. Ekroos, P. J. Meikle, L. Hood, N. D. Price, Alzheimer's Disease Metabolomics Consortium, P. M. Doraiswamy, C. C. Funk, A. I. Hernández, G. Kastenmüller, R. Baillie, X. Han, R. Kaddurah-Daouk, Multi-Omic analyses characterize the ceramide/sphingomyelin pathway as a therapeutic target in Alzheimer's disease. *Commun Biol* **5**, 1074 (2022).

76. S. Vasiliou, Oral fingolimod for the treatment of relapsing-remitting multiple sclerosis. *Drugs Today* **46**, 315–325 (2010).

77. I. Carreras, N. Aytan, J.-K. Choi, C. M. Tognoni, N. W. Kowall, B. G. Jenkins, A. Dedeoglu, Dual dose-dependent effects of fingolimod in a mouse model of Alzheimer's disease. *Sci. Rep.* **9**, 10972 (2019).

78. H. Jeśko, P. L. Wencel, W. J. Lukiw, R. P. Strosznajder, Modulatory Effects of Fingolimod (FTY720) on the Expression of Sphingolipid Metabolism-Related Genes in an Animal Model of Alzheimer's Disease. *Mol. Neurobiol.* **56**, 174–185 (2019).

79. S. Ozakbas, B. Piri Cinar, P. Yigit, C. Baba, O. Sagici, Multiple Sclerosis Research Group, Five-year real-world data on fingolimod treatment's effects on cognitive function. *Mult. Scler. Relat. Disord.* **54**, 103089 (2021).

80. H. Keren-Shaul, A. Spinrad, A. Weiner, O. Matcovitch-Natan, R. Dvir-Szternfeld, T. K. Ulland, E. David, K. Baruch, D. Lara-Astaiso, B. Toth, S. Itzkovitz, M. Colonna, M. Schwartz, I. Amit, A Unique Microglia Type Associated with Restricting Development of Alzheimer's Disease. *Cell* **169**, 1276-1290.e17 (2017).

81. M. L. Bennett, F. C. Bennett, S. A. Liddelow, B. Ajami, J. L. Zamanian, N. B. Fernhoff, S. B. Mulinyawe, C. J. Bohlen, A. Adil, A. Tucker, I. L. Weissman, E. F. Chang, G. Li, G. A. Grant, M. G. Hayden Gephart, B. A. Barres, New tools for studying microglia in the mouse and human CNS. *Proc. Natl. Acad. Sci. U. S. A.* **113**, E1738-46 (2016).

82. D. Boche, M. N. Gordon, Diversity of transcriptomic microglial phenotypes in aging and Alzheimer's disease. *Alzheimers. Dement.* **18**, 360–376 (2022).

83. T. K. Ulland, M. Colonna, TREM2 - a key player in microglial biology and Alzheimer disease. *Nat. Rev. Neurol.* **14**, 667–675 (2018).

84. G. R. Dohle, M. Smit, R. F. A. Weber, Androgens and male fertility. *World J. Urol.* **21**, 341–345 (2003).

85. S. R. Davis, S. Wahlin-Jacobsen, Testosterone in women--the clinical significance. *Lancet Diabetes Endocrinol* **3**, 980–992 (2015).

86. R. A. Nebel, N. T. Aggarwal, L. L. Barnes, A. Gallagher, J. M. Goldstein, K. Kantarci, M. P. Mallampalli, E. C. Mormino, L. Scott, W. H. Yu, P. M. Maki, M. M. Mielke, Understanding the impact of sex and gender in Alzheimer's disease: A call to action. *Alzheimers. Dement.* **14**, 1171–1183 (2018).

87. S. J. Fuller, R. S. Tan, R. N. Martins, Androgens in the etiology of Alzheimer's disease in aging men and possible therapeutic interventions. *J. Alzheimers. Dis.* **12**, 129–142 (2007).

88. E. R. Rosario, L. Chang, E. H. Head, F. Z. Stanczyk, C. J. Pike, Brain levels of sex steroid hormones in men and women during normal aging and in Alzheimer's disease. *Neurobiol. Aging* **32**, 604–613 (2011).

89. L. Yang, R. Zhou, Y. Tong, P. Chen, Y. Shen, S. Miao, X. Liu, Neuroprotection by dihydrotestosterone in LPS-induced neuroinflammation. *Neurobiol. Dis.* **140**, 104814 (2020).

90. K. Maekawa, K. Yamanaka, Role of sex hormones in neuroinflammation in Alzheimer's disease. *Clin. Exp. Neuroimmunol.* **14**, 100–109 (2023).

91. D. K. Franco-Bocanegra, C. McAuley, J. A. R. Nicoll, D. Boche, Molecular Mechanisms of Microglial Motility: Changes in Ageing and Alzheimer's Disease. *Cells* **8**, 639 (2019).

92. J. C. Lambert, C. A. Ibrahim-Verbaas, D. Harold, A. C. Naj, R. Sims, C. Bellenguez, G. Jun, A. L. DeStefano, J. C. Bis, G. W. Beecham, B. Grenier-Boley, G. Russo, T. A. Thornton-Wells, N. Jones, A. V. Smith, V. Chouraki, C. Thomas, M. A. Ikram, D. Zelenika, B. N. Vardarajan, Y. Kamatani, C. F. Lin, A. Gerrish, H. Schmidt, B. Kunkle, N. Fiévet, P. Amouyel, F. Pasquier, V. Deramecourt, R. F. A. G. De Bruijn, N. Amin, A. Hofman, C. M. Van Duijn, M. L. Dunstan, P. Hollingworth, M. J. Owen, M. C. O'Donovan, L. Jones, P. A. Holmans, V. Moskvina, J. Williams, C. Baldwin, L. A. Farrer, S. H. Choi, K. L. Lunetta, A. L. Fitzpatrick, T. B. Harris, B. M. Psaty, J. R. Gilbert, K. L. Hamilton-Nelson, E. R. Martin, M. A. Pericak-Vance, J. L. Haines, V. Gudnason, P. V. Jonsson, G. Eiriksdottir, M. T. Bihoreau, M. Lathrop, O. Valladares, L. B. Cantwell, L. S. Wang, G. D. Schellenberg, A. Ruiz, M. Boada, C. Reitz, R. Mayeux, A. Ramirez, W. Maier, O. Hanon, W. A. Kukull, J. D. Buxbaum, D. Campion, D. Wallon, D. Hannequin, P. K. Crane, E. B. Larson, T. Becker, C. Cruchaga, A. M. Goate, D. Craig, J. A. Johnston, B. Mc-Guinness, S. Todd, P. Passmore, C. Berr, K. Ritchie, O. L. Lopez, P. L. De Jager, D. Evans, S. Lovestone, P. Proitsi, J. F. Powell, L. Letenneur, P. Barberger-Gateau, C. Dufouil, J. F. Dartigues, F. J. Morón, D. C. Rubinsztein, P. St. George-Hyslop, K. Sleegers, K. Bettens, C. Van Broeckhoven, M. J. Huentelman, M. Gill, K. Brown, K. Morgan, M. I. Kamboh, L. Keller, L. Fratiglioni, R. Green, A. J. Myers, S. Love, E. Rogaeva, J. Gallacher, A. Bayer, J. Clarimon, A. Lleo, D. W. Tsuang, L. Yu, D. A. Bennett, M. Tsolaki, P. Bossù, G. Spalletta, J. Collinge, S. Mead, S. Sorbi, B. Nacmias, F. Sanchez-Garcia, M. C. Deniz Naranjo, N. C. Fox, J. Hardy, P. Bosco, R. Clarke, C. Brayne, D. Galimberti, M. Mancuso, F. Matthews, S.

Moebus, P. Mecocci, M. Del Zompo, H. Hampel, A. Pilotto, M. Bullido, F. Panza, P. Caffarra, M. Mayhaus, S. Pichler, W. Gu, M. Riemenschneider, L. Lannfelt, M. Ingelsson, H. Hakonarson, M. M. Carrasquillo, F. Zou, S. G. Younkin, D. Beekly, V. Alvarez, E. Coto, C. Razquin, P. Pastor, I. Mateo, O. Combarros, K. M. Faber, T. M. Foroud, H. Soininen, M. Hiltunen, D. Blacker, T. H. Mosley, C. Graff, C. Holmes, T. J. Montine, J. I. Rotter, A. Brice, M. A. Nalls, J. S. K. Kauwe, E. Boerwinkle, R. Schmidt, D. Rujescu, C. Tzourio, M. M. Nöthen, L. J. Launer, S. Seshadri, Meta-analysis of 74,046 individuals identifies 11 new susceptibility loci for Alzheimer's disease. *Nat. Genet.* **45**, 1452–1458 (2013).

93. C. M. Karch, L. A. Ezerskiy, S. Bertelsen, Alzheimer's Disease Genetics Consortium (ADGC), A. M. Goate, Alzheimer's Disease Risk Polymorphisms Regulate Gene Expression in the ZCWPW1 and the CELF1 Loci. *PLoS One* **11**, e0148717 (2016).

94. G. Novikova, M. Kapoor, J. Tcw, E. M. Abud, A. G. Efthymiou, S. X. Chen, H. Cheng, J. F. Fullard, J. Bendl, Y. Liu, P. Roussos, J. L. Björkegren, Y. Liu, W. W. Poon, K. Hao, E. Marcora, A. M. Goate, Integration of Alzheimer's disease genetics and myeloid genomics identifies disease risk regulatory elements and genes. *Nat. Commun.* **12**, 1610 (2021).

95. B. W. Henderson, E. G. Gentry, T. Rush, J. C. Troncoso, M. Thambisetty, T. J. Montine, J. H. Herskowitz, Rho-associated protein kinase 1 (ROCK1) is increased in Alzheimer's disease and ROCK1 depletion reduces amyloid-β levels in brain. *J. Neurochem.* **138**, 525–531 (2016).

96. E. G. Gentry, B. W. Henderson, A. E. Arrant, M. Gearing, Y. Feng, N. C. Riddle, J. H. Herskowitz, Rho Kinase Inhibition as a Therapeutic for Progressive Supranuclear Palsy and Corticobasal Degeneration. *J. Neurosci.* **36**, 1316–1323 (2016).

97. M.-F. Guo, H.-Y. Zhang, Y.-H. Li, Q.-F. Gu, W.-Y. Wei, Y.-Y. Wang, X.-J. Zhang, X.-Q. Liu, L.-J. Song, Z. Chai, J.-Z. Yu, C.-G. Ma, Fasudil inhibits the activation of microglia and astrocytes of transgenic Alzheimer's disease mice via the downregulation of TLR4/Myd88/NF-κB pathway. *J. Neuroimmunol.* **346**, 577284 (2020).

98. J. Chen, Z. Sun, M. Jin, Y. Tu, S. Wang, X. Yang, Q. Chen, X. Zhang, Y. Han, R. Pi, Inhibition of AGEs/RAGE/Rho/ROCK pathway suppresses non-specific neuroinflammation by regulating BV2 microglial M1/M2 polarization through the NF-κB pathway. *J. Neuroimmunol.* **305**, 108–114 (2017).

99. R. Killick, C. Elliott, E. Ribe, M. Broadstock, C. Ballard, D. Aarsland, G. Williams, Neurodegenerative disease associated pathways in the brains of triple transgenic Alzheimer's model mice are reversed following two weeks of peripheral administration of fasudil. *Int. J. Mol. Sci.* **24** (2023).

100. R. Collu, Z. Yin, E. Giunti, S. Daley, M. Chen, P. Morin, R. Killick, S. T. C. Wong, W. Xia, Effect of the ROCK inhibitor fasudil on the brain proteomic profile in the tau transgenic mouse model of Alzheimer's disease. *Front. Aging Neurosci.* **16**, 1323563 (2024).

101. A. Kroiss, S. Vincent, M. Decaussin-Petrucci, E. Meugnier, J. Viallet, A. Ruffion, F. Chalmel, J. Samarut, N. Allioli, Androgen-regulated microRNA-135a decreases prostate cancer cell migration and invasion through downregulating ROCK1 and ROCK2. *Oncogene* **34**, 2846–2855 (2015).

102. A.-E. Roser, L. Tönges, P. Lingor, Modulation of Microglial Activity by Rho-Kinase (ROCK) Inhibition as Therapeutic Strategy in Parkinson's Disease and Amyotrophic Lateral Sclerosis. *Front. Aging Neurosci.* **9**, 94 (2017).

103. A. M. Horstman, E. L. Dillon, R. J. Urban, M. Sheffield-Moore, The role of androgens and estrogens on healthy aging and longevity. *J. Gerontol. A Biol. Sci. Med. Sci.* **67**, 1140–1152 (2012).

104. R. Ietswaart, B. M. Gyori, J. A. Bachman, P. K. Sorger, L. S. Churchman, GeneWalk identifies relevant gene functions for a biological context using network representation learning. *Genome Biol.* **22**, 55 (2021).

105. V. D. Blondel, J.-L. Guillaume, R. Lambiotte, E. Lefebvre, Fast unfolding of communities in large networks. *J. Stat. Mech: Theory Exp.* **P10008**, 1–12 (2008).
